# The health and wellbeing impact of arts engagement: A longitudinal outcome-wide approach in 22 countries

**DOI:** 10.64898/2026.11.18.26368421

**Authors:** Hei Wan Mak, Keyu Li, Christos A. Makridis, Ying Chen, Dorota Weziak-Bialowolska, Eric S. Kim, Daisy Fancourt

## Abstract

Arts and cultural engagement is increasingly recognised as a potential resource for health and wellbeing, yet evidence remains dominated by Western, high-income countries. Using data from the Global Flourishing Study across 22 countries (N=118,045), we examined associations between arts engagement and 19 health and wellbeing outcomes across five domains, applying an outcome-wide analytical framework and random-effects meta-analysis. Arts engagement was consistently associated with greater positive wellbeing and character strengths, higher evaluative wellbeing, and better social and general health, including lower loneliness. By contrast, associations with future-oriented wellbeing and perceived capability were less robust, and no pooled associations emerged for psychological distress. Country-level characteristics explained little cross-national variation. These findings suggest that arts engagement may promote flourishing across diverse cultural contexts, with benefits extending beyond the prevention of psychological distress, highlighting its potential role as a global population health resource.

## Introduction

Arts engagement is a universal human behaviour, ubiquitous across societies globally. Whilst artistic practices vary widely across cultures, arts engagement is often grouped into broad categories, including attending, performing, participating in, consuming or learning about dance/movement, literary arts, media, music, theatre/performance, and visual arts, craft and design.^1^ Beyond its associations with enjoyment, appreciation, creativity, innovation, and emotional expression, arts engagement has deep evolutionary roots in supporting social bonding, group cohesion, and cooperation, as well as individual affect, emotion regulation, and stress reduction with potential implications for health, longevity, and survival.^2^ Over the past few decades, a growing body of literature has demonstrated the association between arts engagement and health. For instance, epidemiological studies have found longitudinal associations between arts engagement and reduced incidence of depression,^3^ slower cognitive decline and longer time to onset of dementia,^4^ reduced frailty and disability,^5,6^ decelerated physiological ageing,^7^ and lower all-cause mortality,^8^ with associations independent of socioeconomic, behavioural, and health-related confounders. Randomised controlled trials have focused on the ‘treatment’ effect of arts interventions on diagnosed depression, anxiety, stress, social competences, and quality of life in both clinical and non-clinical samples, with pooled effect sizes showing some comparability to those of established psychological and pharmacological treatments (with this comparability echoed in the first recent head-to-head trials).^9–12^ Complexity science theories suggest that arts engagement achieves these diverse health benefits because it comprises multiple salutogenic ingredients,^13^ such as sensory stimulation, cognitive engagement, and social and physical activity, which may activate diverse and interacting mechanisms.^14^ This theoretical positioning is supported by increasing evidence elucidating the neurocognitive, bio-physiological, and socio-behavioural processes through which arts engagement may influence health. These processes (demonstrated through laboratory experiments and qualitative research) include the induction of positive emotions, emotion regulation, processing of unconscious material, fulfilment of psychological needs, meaning-making, identity formation, activation of neural reward networks, downregulation of neuroendocrine stress response systems and inflammatory immune pathways, increases in oxytocin, enhanced immune function, structural and functional brain changes consistent with neuroplasticity, engagement in health-promoting behaviours, and potential slowing of epigenetic and physiological ageing processes.^15,14,16–20^

Despite this momentum, important gaps remain. First, the evidence base is geographically and demographically narrow. Most arts-and-health research has been conducted in high-income, Global North settings – a limitation explicitly acknowledged within the field – leaving the majority of the world’s population untested.^19,21^ Whether these potential benefits extend to populations with different cultural traditions, social values and norms, and cultural resources remains an open question. Exploring and comparing cross-country differences in arts and health is timely. In Western contexts, arts engagement is increasingly recognised as an additional pillar of health and a health-promoting behaviour.^22^ However, this framing cannot simply be extended uncritically to other cultural contexts due to differences in the way that arts engagement is defined, valued and emerges cross-culturally.^1^ Nevertheless, understanding whether the arts may support health in a similar way in other geographical regions could open up important conversations about the role of arts engagement in population health globally. Such conversations would be particularly relevant because arts-and-health research and policy often carry an implicit expectation that the arts can become more accessible, inclusive, and relevant to wider populations. Existing evidence suggests that there are social gradients in arts participation within and across geographically and culturally diverse countries.^23,24^ Cross-country evidence can help establish, therefore, whether inequalities in art engagement may also have implications for health across cultural contexts.^23^

A second major gap relates to limited availability of cross-country evidence based on standardised measures. Epidemiological literature on arts engagement and health, despite being relatively young, has begun to extend earlier single-country evidence to cross-country comparisons, largely through harmonising data from individual population-based cohort studies.^25,26^ A key strength of these cohorts is that they are often designed to reflect local cultural norms, values, and traditions, meaning that measures of arts engagement may be culturally specific and locally meaningful. For instance, the Japanese ageing cohort, the Japan Gerontological Evaluation Study (JAGES), includes questions on activities such as flower arrangement and Haiku poetry, which do not typically appear in Western ageing cohorts. However, harmonisation also brings challenges: measures may not be fully comparable across countries, and subtle differences in wording, response options, and cultural interpretation can complicate interpretation. In addition, many of the most comprehensive cohorts are located in Western, more developed, or Global North countries, further skewing the evidence base towards Western contexts. To mitigate these limitations, and to cross-validate findings from previous comparative cohort studies, there is a need for complementary evidence using cross-national datasets in which the same standardised survey instruments are jointly selected and administered across a broad range of countries.^25^ This approach can reduce bias introduced through post-hoc harmonisation, generate evidence from non-Western, less economically developed, or Global South countries where cohort data may be limited or unavailable, and provide a clearer basis for comparing associations between arts engagement and health across cultural contexts.

Finally, from an ecological perspective, there remains a critical gap in understanding how the effect of arts engagement on individual health may be shaped by broader environmental and societal factors. Social-ecological models emphasise that individual behaviours and health are not formed in isolation, but through interaction with wider social, cultural, economic, and policy environments.^27,28^ This perspective is also reflected in the World Health Organization’s social determinants of health framework, which highlights how individual lifestyles and health behaviours are shaped by broader conditions such as socioeconomic circumstances, inequalities, and cultural norms.^29,30^ Applying this perspective to arts engagement and health is important as it can help understand whether such associations are observed across countries regardless of country-level characteristics, or whether these associations are moderated by them. This approach can help move the field beyond looking at individual-level associations towards conditions under which arts engagement may be most health-promoting.

To address these gaps and adopt a causal framing approach,^31^ this study examined the longitudinal effects of arts engagement on a range of mental and physical health outcomes using data from the Global Flourishing Study (GFS). The GFS includes over 200,000 participations across 22 geographically and culturally diverse countries, spanning both the Global North and South. This provides an opportunity to reduce the current Western bias in the evidence base, to assess the consistency and robustness of the effects of arts on health across countries using standardised survey measures, and to explore whether these effects vary according to country-level characteristics. Specifically, we investigated: (a) whether and how arts engagement affected a wide range of subsequent health outcomes; (b) whether these effects were consistent across countries; and (c) whether they were moderated by a range of country-level factors.

## Methods

### The Global Flourishing Study (GFS)

GFS is a longitudinal study of over 200,000 participants in 22 geographically and culturally diverse countries and one Special Administrative Region (Hong Kong SAR, China), with samples designed to be nationally representative. These include Argentina, Australia, Brazil, China, Egypt, Germany, Hong Kong SAR, India, Indonesia, Israel, Japan, Kenya, Mexico, Nigeria, the Philippines, Poland, South Africa, Spain, Sweden, Tanzania, Türkiye, United Kingdom (UK), and United States (US). Some countries used both or either probability-based and non-probability based sampling approaches for recruitment, whereas the US, Sweden and Hong Kong (S.A.R. of China) only used existing web panels with set quotas for age, gender, region and education to ensure adequate representation of the population.^32^ GFS collects rich data to understand various aspects of flourishing and its determinants, and contains numerous socio-demographic, economic, political, religious, personality, lifestyle, community, health and wellbeing variables. Data were also collected on retrospective assessments of childhood experiences. The data are publicly available upon preregistration through the Center for Open sciences (COS; Johnson et al., 2024, https://osf.io/3jtz8/).

This present study analysed data from Wave 1 (2022-23), mid-year retention data (2023-24), and Wave 3 (2025). The mid-year survey was not administered in a few countries and territories for various reasons such as cost implications and the utility of retention activity. These countries included China, Hong Kong (S.A.R. of China), Japan, Israel, Sweden and the US. In these countries or territories, the substantive mid-year survey questions (including the arts engagement question) were included in the second annual survey (2024) so that all respondents had an opportunity to answer them. Our analyses included participants who provided responses to these questions. Of all Wave 1 respondents (N=207,919), N=131,487 answered the mid-year survey items and N=131,183 completed the arts measure. Of these, N=118,045 further provided complete data across all demographic, socio-economic, family and social relationships, and health and health behaviour measures considered in our present study. Ethical approval was granted by the institutional review boards at Baylor University (IRB Reference #: 1841317) and Gallup (IRB Reference #: 2021-11-02), and all participants provided informed consent. Analytical plans were pre-registered at https://osf.io/3w8bj/overview. See **Figure 1** for the overall methodological framework.

**Figure 1.**
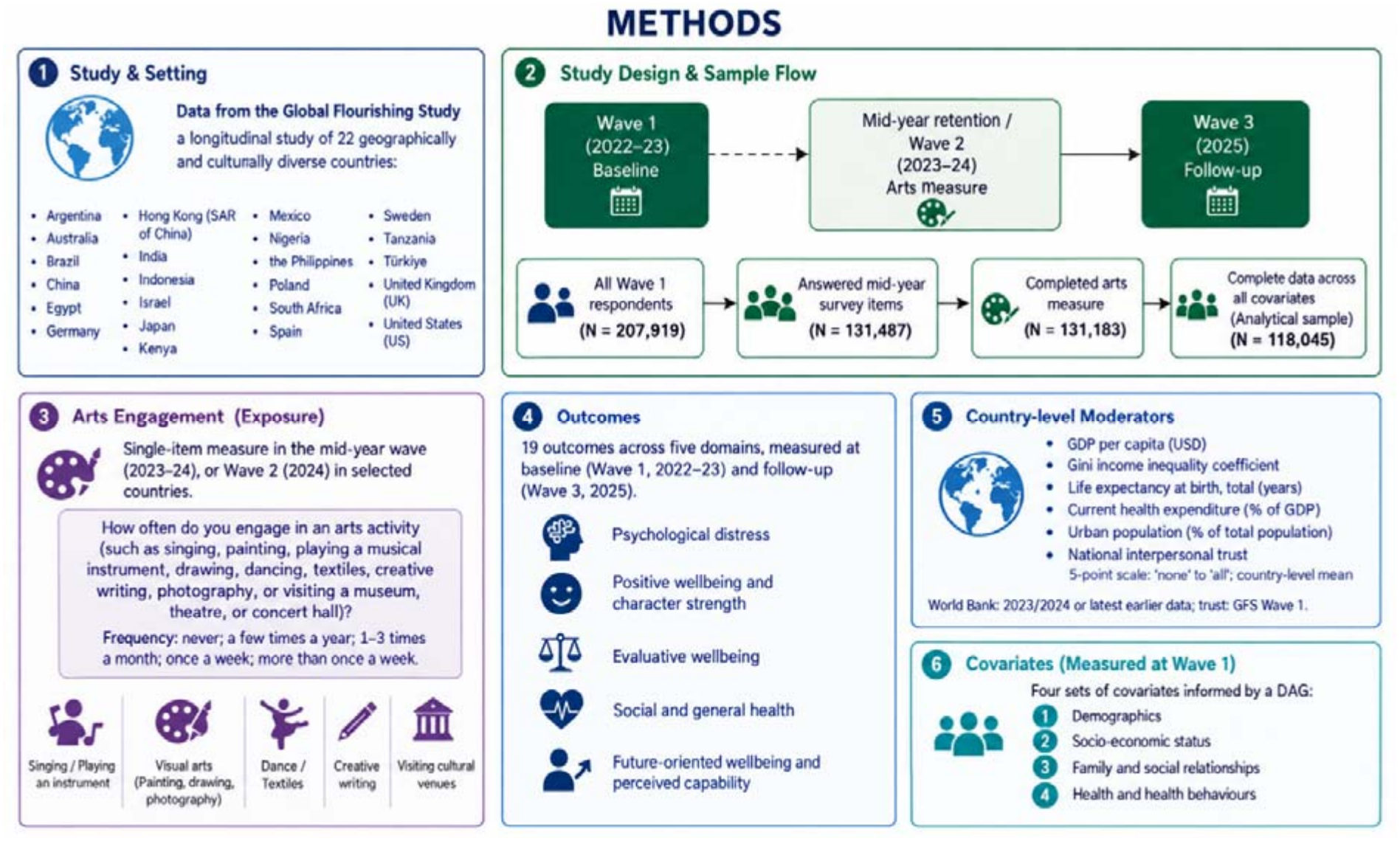
Overall methodological framework. Figure was generated using ChatGPT and reviewed and approved by authors.

### Measures

#### Arts engagement

Arts engagement was measured via a single-item question in the mid-year wave, 2023-24, (or in Wave 2, 2024, for some countries as mentioned earlier) asking *How often do you engage in an arts activity (such as singing, painting, playing a musical instrument, drawing, dancing, textiles, creative writing, photography, or visiting a museum, theatre, or concert hall)?* Response options included never, a few times a year, one to three times a month, once a week, and more than once a week. These categories were retained in the analyses.

While the single-item measure focuses specifically on the frequency of arts engagement, one of several dimensions of arts participation acknowledged by UNESCO,^33^ previous studies using GFS data suggest that a single-item measure may be well-suited to cross-national surveys.^24^ First, measuring arts participation across culturally and geographically diverse is inherently challenging because understanding of what considers as ‘arts’, including whether engagement is considered ‘active’ or ‘receptive’, vary across contexts. A broad measure can capture practices embedded in everyday life without imposing categories that may not translate across social groups, cultures, or regions. Second, the item did not require arts engagement to be the participants’ primary activity. Responses may therefore encompass engagement occurring through religious participation, physical activity, community traditions or festivals, including activities that participants engage in secondarily and might not otherwise identify as arts engagement.

#### Health and wellbeing outcomes

We took an outcome-wide approach to examine multiple outcome indicators across five domains of health and wellbeing.^34^ All these indicators were measured at baseline (Wave 1, 2022-23) and three years later (2025).

##### Psychological distress

We used five measures, all of which assessed symptoms experienced in the last two weeks: (1) feeling nervous, anxious or on edge, (2) not being able to stop or control worrying, (3) feeling down, depressed or hopeless, and (4) little interest of pleasure in doing things. Each item was measured on a 4-point scale ranging from ‘not at all’ to ‘nearly every day’. We also considered (5) the extent to which participants felt they were suffering,^35,36^ either physically or mentally (also on a 4-point scale ranging from ‘not at all’ to ‘a lot’). As a sensitivity analysis, we additional explored how arts engagement might be associated with a combination of anxiety and depressive symptoms, measured using a composite score derived from the group items of the PHQ-4 measures.^37^

##### Positive wellbeing and character strength

We used five measures: (6) happiness, (7) understanding purpose in life, (8) feeling that things done in life were worthwhile, (9) gratitude (measured by feeling grateful), and (10) feeling at peace with one’s thoughts and feelings. Happiness, understanding purpose in life, feeling that things done in life were worthwhile, and gratefulness were measured on 11-point scales (with higher scores indicate greater levels), while feeling at peace with one’s thoughts and feelings was measured on a 4-point scale ranging from ‘never’ to ‘always.

##### Evaluative wellbeing

We used three measures: (11) life satisfaction, (12) life evaluation today, (13) expected life evaluation five years from now. Life satisfaction was measured on an 11-point scale ranging from ‘not at all satisfied with your life’ to ‘completely satisfied with your life’. Life evaluation today and life evaluation five years from now was measured on 11-point scales ranging from the ‘worst possible’ to the ‘best possible’.

##### Social and general health

We used three measures: (14) loneliness, (15) overall mental health, and (16) overall physical health. Loneliness was measured on an 11-point scale ranging from ‘never’ to ‘always’. Overall mental health and physical health were each measured on 11-point scales ranging from ‘poor’ to ‘excellent’.

##### Future-oriented wellbeing and perceived capability

We used three measures: (17) remaining hopeful about the future, (18) expecting more good things to happen in life, and (19) feeling very capable in most things done in life. Remaining hopeful about the future and expecting more good things to happen in life were each measured on 11-point scales ranging from ‘strongly disagree’ to ‘strongly agree’. Feeling capable was measured on a 4-point scale ranging from ‘never’ to ‘always’.

#### Country-level moderators

We explored a set of country-level factors that might moderate the individual-level association between arts engagement and the outcomes. These factors were selected to capture national economic conditions, income inequality, urbanisation, population health, healthcare expenditure, and interpersonal trust. They included the logged GDP per capita (USD),^38^ Gini income inequality coefficient (data were not available for Hong Kong SAR),^39^ life expectancy at birth, total (years),^40^ current health expenditure (% of GDP; data were not available for Hong Kong SAR),^41^ urban population (% of total population),^42^ and national level of interpersonal trust (the proportion of people in their country trusting on another; a 5-point scale ranging from ‘none’ to ‘all’). Data on the GDP per capita, Gini income inequality coefficient, life expectancy at birth, health expenditure (% of GDP), and urban population were extracted from the World Bank Group data for 2023 or 2024. Where data were unavailable for either year, the most recent available from earlier years were used. National level of interpersonal trust was extracted from the Wave 1 of the GFS and calculated as the country-level mean.

#### Covariates

We controlled for four sets of covariates identified through a Directed Acyclic Graph (DAG) and measured in Wave 1. A simplified DAG illustrating the assumed causal structure underlying the outcome-wide analyse is reported **in Supplementary Figure S1. Set 1: Demographics**. This included age (a continuous variable ranging from 18-99), gender (female vs male; participants who selected either ‘other’ or ‘prefer not to say’ were dropped), country of birth (native-born vs foreign-born), marital status (married/domestic partner vs single vs separated/divorced/widowed), number of children (a continuous variable ranging from 0-6+), living area (urban vs rural/town), and religious attendance (at least weekly vs less than once a week or never). **Set 2: Socio-economic status**. This included education (up to 8 years of basic education, 9-15 years of education, completed four years of education beyond high school and/or received a 4-year college degree), employment (employed/self-employed, unemployed, not in labour force e.g. retired, student, homemaker), and feelings about household income (finding it difficult/very difficult on present income vs getting by/living comfortably). **Set 3: Family and social relationships**. This included relationship with parents while growing up, derived by combining responses on relationships with the mother and father and using the highest reported level of relationship equality. Responses categories were very bad, somewhat bad, somewhat good, very good. The set also considered physical or sexual abuse experiences while growing up (yes vs no; data were unavailable for Israel), and whether people counted on others to help if they were in trouble (an 11-point scale ranging from ‘never’ to ‘always’). Finally, **Set 4: Health and health behaviours**. This included physical activity (≥5 days a week vs <5 days a week), number of alcoholic drinks in the past seven days (a continuous variable ranging from 0-10+), smoking habit (smoked vs did not smoke), and health problems that limiting activities (yes vs no).^43^

#### Analysis

Using an outcome-wide approach, we tested the effect of arts engagement frequency on health and wellbeing outcomes a year later in regression models. The type of regression was determined by the outcome: ordered logistic regression for ordinal outcomes and linear regression for continuous. Models were built sequentially: Model 1: unadjusted; Model 2: adjusted for demographics, socio-economic status and baseline outcome measured at Wave 1, Model 3: adjusted for demographics, socio-economic status, family and social relationships, and baseline outcome measured at Wave 1, and Model 4: fully adjusted for baseline outcome measured at Wave 1, demographics, socio-economic status, family and social relationship, and health and health behaviours. Israel was excluded from the main analyses due to data unavailable in one of the confounders (which was abuse experience while growing up). We nonetheless provide findings from Israel in **Supplementary Figure S2**, based on fully-adjusted analyses conducted but without adjusting for this confounder.

Regression models were run separately for each country to obtain country-specific effect estimates (unstandardized coefficients/ odds ratios and 95%CIs). These estimates were then pooled using a random-effects meta-analysis to derive an overall effect size. Heterogeneity was examined by computing I^2^, H^2^ and τ^2^ statistics. I^2^ is the percentage of variability in the effect size that is caused by between-country heterogeneity, rather than by sampling error, with values of 25%, 50%, and 75% indicating low, moderate, and high, respectively.^44^ The H^2^ statistics describes the ratio of the observed variation and the expected variance due to sampling error, with larger values indicating greater heterogeneity.^45^ The τ^2^ statistics estimates the between-country variance in the true effect sizes on the scale used for meta-analysis, with larger values indicating greater heterogeneity.^47,48^ We further conducted exploratory meta-regression to examine whether country-level characteristics moderated variation in effect sizes across countries. Each country-level moderator was entered in the analysis separately. It is important to note that these analyses were based on 22 country-level units.

Several sensitivity analyses were performed. First, to assess whether the findings were sensitive to adjustment for baseline health, we ran the analysis without including baseline health as a covariate. Second, to assess whether ceiling effects influenced the findings, we repeated the analyses after excluding respondents with the highest levels of health or wellbeing at baseline, as these individuals had limited potential for subsequent improvement. For ordinal variables, we excluded those who reported the ‘healthiest’ response option; for continuous variables, we excluded those in the healthiest 20% of the baseline distribution. Third, to examine whether any engagement vs frequency of engagement is more important, we ran two parallel analyses: (a) treating arts engagement as a binary indicator (engaged vs not engaged), and (b) restricting the sample to those who engaged and examining frequency of engagement.

Across all meta-analyses, 11 of 3,784 country-specific estimates (0.29%) were unavailable because the models failed to converge. In Indonesia, estimates for little interest or pleasure in doing things failed to converge in both the fully adjusted main analysis and the sensitivity analysis using a binary measure of arts engagement. Estimates for suffering failed to converge in two sensitivity analyses: one that omitted baseline outcome adjustment and one that restricted the sample to participants below the healthiest baseline category. The model examining uncontrollable worrying also failed to converge in the latter analysis. In Nigeria, estimates for feeling at peace failed to converge in the nested model adjusted for sociodemographic confounders, social and family relationship confounders, and the baseline outcome, in the fully adjusted main model, and in all four sensitivity analyses examining this outcome.

Annual sampling weights provided by GFS were applied for regression models to adjust for unequal probabilities of selection and differential response rates, ensuring estimates are more representative of the population.^49^ Missing data were handled using listwise deletion. Analytical plans were pre-registered at https://osf.io/kz7b9/overview. All code for the analysis can be found at https://osf.io/mwgxz/files/osfstorage. Analyses were carried out in Stata v19.5.

## Results

### Sample descriptives

The average age of the respondents across countries ranged from 34.8 to 54.3 years, with participants in Nigeria being the youngest on average and those in Japan the oldest (**Supplementary Table S1**). Gender distributions were generally balanced. In most countries and territories, more than 90% of participants were native born, with lower proportions observed in Australia, the UK, and Spain. The proportion of participants who were married ranged from 31.5% in South Africa to 78.6% in India, despite the two countries having similar mean age. Urban residence also varied considerably, from 11.6% in Kenya to 92.8% in Hong Kong SAR. The proportion of participants with a degree ranged from 0.9% in Tanzania to 40.2% in the US, while employment ranged from 32.6% in South Africa to 73.8% in Hong Kong SAR. Arts engagement also differed markedly across countries, with the lowest prevalence observed in Egypt at 12.9% and the highest in Nigeria at 86.6%. Notably, arts engagement also varied even across neighbouring countries. For example, within East Asia, 41.6% of participants in Japan reported engaging in the arts, compared with 62.4% in Hong Kong SAR and 80.0% in China. However, these proportions should be interpreted with caution, as they may partly reflect cross-country differences in how respondents understood and interpreted the survey question on arts engagement.

### Outcome-wide analyses

Our meta-analyses showed that the effects of arts engagement on positive wellbeing and character strength outcomes, as well as evaluative wellbeing, persisted after full adjustment, although they generally attenuated slightly after adjustment for baseline outcome, with little further attenuation with additional adjustments (**Figure 2**). Effects on lower loneliness and better overall mental health became more apparent after adjustment. Effects on future-oriented wellbeing and perceived capability associations were weaker and less clearly supported after adjustment for socio-demographic confounders and baseline outcome.

**Figure 2.**
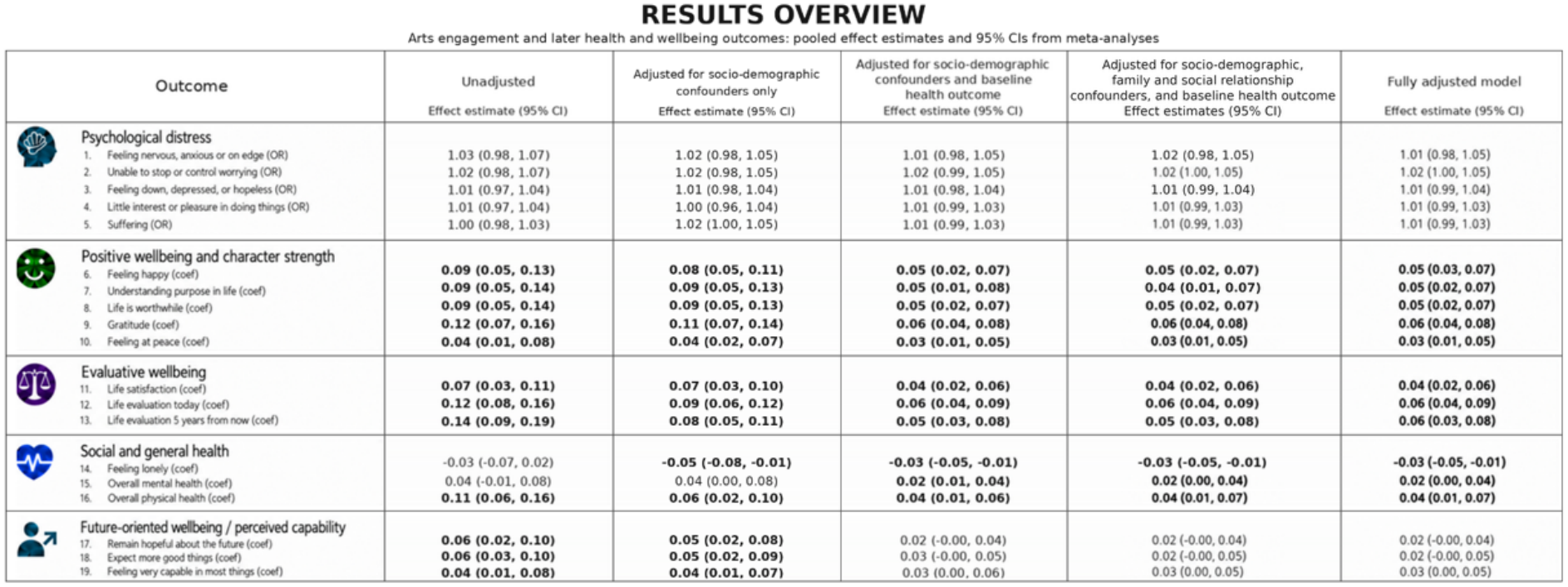
Pooled unstandardised regression coefficients and 95%CIs from meta-analysis of associations between arts engagement and later health and wellbeing outcomes. Bold values denote p <0.05. Column 1 as unadjusted, column 2 adjusted for confounder sets 1 and 2, column 3 additionally adjusted for baseline health, column 4 additionally set 3, and column 5 additionally adjusted for set 4. The figure was created from tables of results using ChatGPT (OpenAI) and reviewed for accuracy and approved by the authors.

In our fully adjusted model, arts engagement was positively associated with positive wellbeing and character strength, evaluative wellbeing, as well as social and general health, reported in the subsequent wave. Specifically, engagement was related to happiness (coef=0.05, 95%CI=0.03, 0.07), understanding purpose in life (coef=0.05, 95%CI=0.02, 0.07), life satisfaction (coef=0.04, 95%CI=0.02, 0.08), life evaluation today (coef=0.06, 95%CI=0.04, 0.09), life evaluation 5 years from now (coef=0.06, 95%CI=0.03, 0.08), feeling grateful (coef=0.06, 95%CI=0.04, 0.08), feeling life is worthwhile (coef=0.05, 95%CI=0.02, 0.07), feeling at peace (coef=0.03, 95%CI=0.01, 0.05), and overall physical health (coef=0.04, 95%CI=0.01, 0.07). Engagement was also negatively associated with loneliness (coef=-0.03, 95%CI=-0.05, -0.01). Associations between arts engagement and individual psychological distress symptoms, the composite PHQ-4 measure, future-oriented wellbeing, and perceived capability were less clear.

Full meta-analysis forest plots are presented in **Supplementary Figure S3** for unadjusted models, **Supplementary Figure S4** for models adjusted only for sociodemographic confounders, **Supplementary Figure S5** for models adjusted for sociodemographic confounders and baseline outcome, **Supplementary Figure S6** for models adjusted for sociodemographic confounders, family and social relationships confounders, and baseline outcome, and **Supplementary Figure S7** for fully adjusted models, and **Supplementary Figure 8** for composite PHQ-4 measure.

#### Variation across countries

While some patterns appeared relatively consistent across countries, others varied across context settings. For instance, most countries showed no clear association between arts engagement and feeling nervous, yet arts engagement was associated with higher odds of feeling nervous in Poland (OR=1.24, 95%CI=1.12, 1.38) and the United States (OR=1.10, 95%CI=1.06, 1.13), but lower odds in Egypt (OR=0.79, 95%CI=0.69, 0.90) (**Figure 3a**). Similar cross-national variation amongst these countries was observed for feeling down, depressed, or hopeless (**Figure 3b**), and for being unable to stop or control worrying (**Figure 3c**). Notably, variation was also observed between neighbouring regions. For instance, arts engagement was associated with understanding one’s purpose in life in China (coef=0.07, 95%CI=0.01, 0.13) and Japan (coef=0.11, 95%CI=0.07, 0.14), but no clear association was observed in Hong Kong SAR (**Figure 3d**). (NB differences should be interpreted cautiously, particularly where odds ratios were small or confidence intervals were wide.)

**Figure 3.**
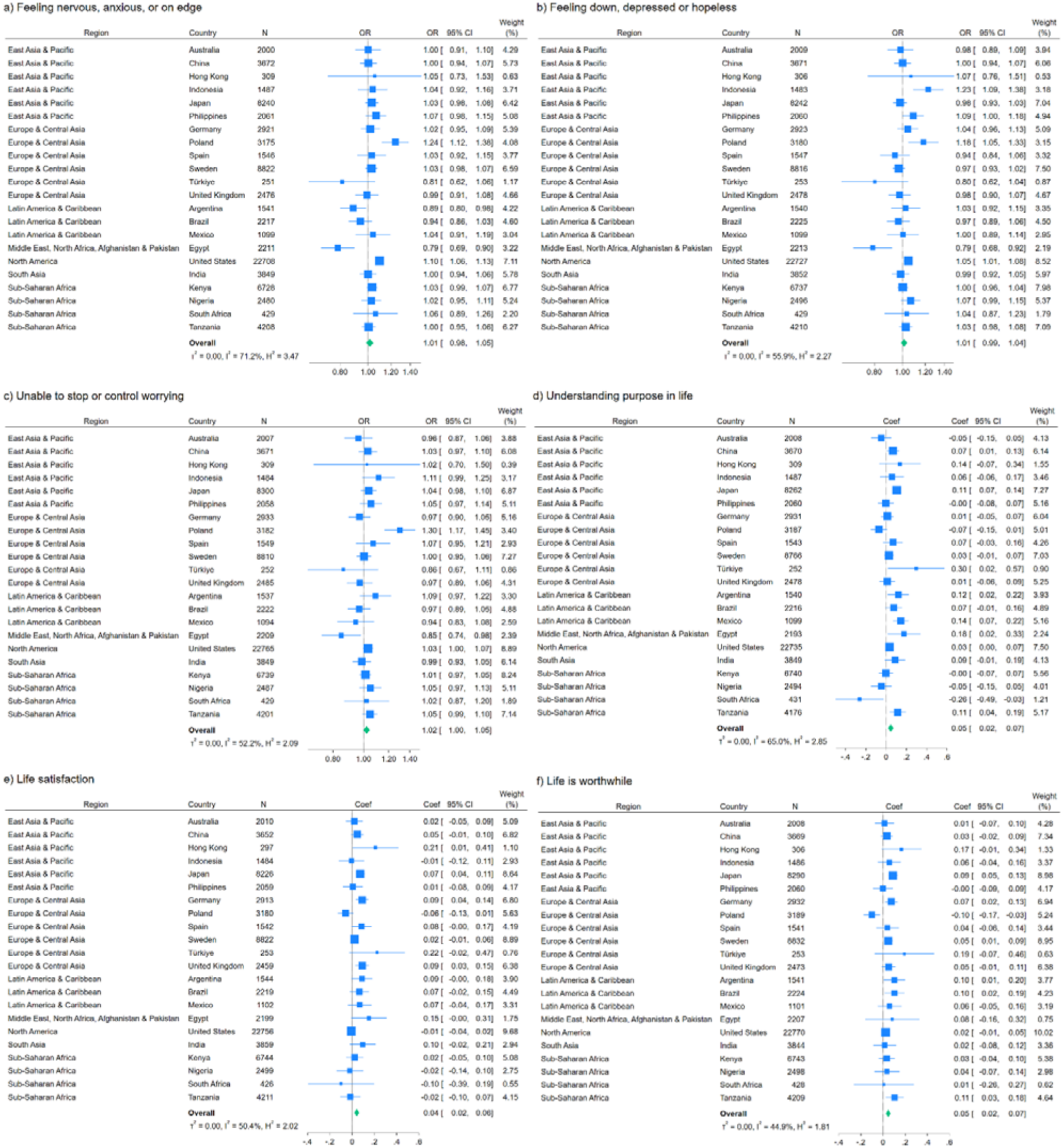
Selected meta-analysis forest plots showing country-specific and pooled associations between arts engagement and subsequent health and wellbeing outcomes from fully adjusted models. Forest plots for all outcomes are presented in the Supplementary Figures S7.

#### Robustness checks

When baseline outcome was excluded from the models, associations with positive wellbeing and character strength, evaluative wellbeing, and social and general health remained, with additional associations observed for future-oriented wellbeing and perceived capability (**Supplementary Figure S9**). When restricting the sample to participants below the healthiest baseline category (i.e. removing the ceiling effect), results were broadly consistent with the main analyses (**Supplementary Figure S10**). When arts engagement was binarised as engaged vs not engaged, associations with positive wellbeing and character strength, evaluative wellbeing, and social and general health were broadly consistent with the main analyses, with additional associations observed for future-oriented wellbeing (**Supplementary Figure S11**). When analyses were restricted to participants who engaged in the arts and focused on frequency of engagement, positive associations between arts engagement and several health and wellbeing outcomes remained evident (**Supplementary Figure S12**). See **Figure 4** for the results overview.

**Figure 4.**
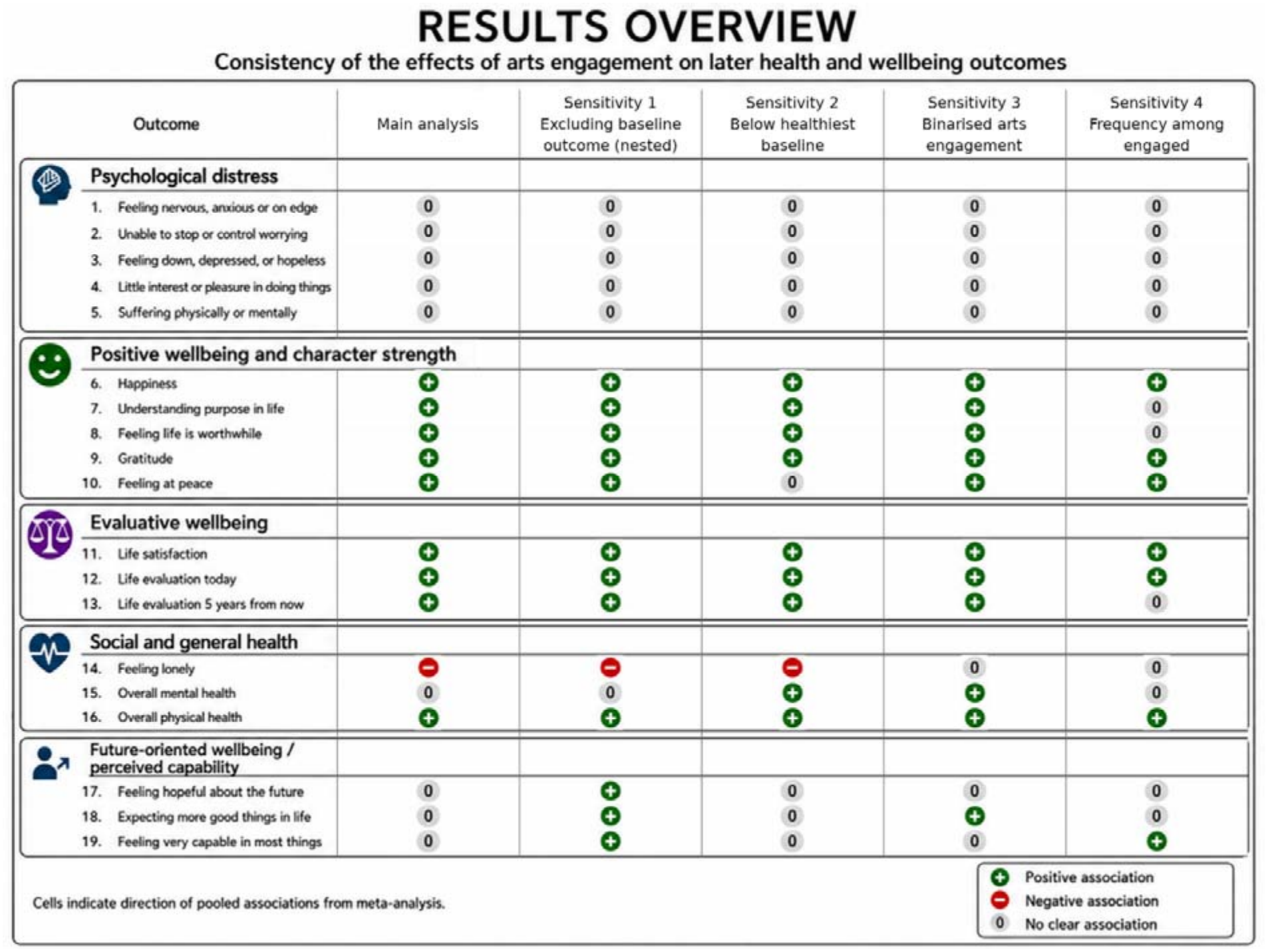
Overview of pooled effect estimates from the main meta-analysis model and sensitivity analyses. The figure was created using ChatGPT (OpenAI) and reviewed for accuracy and approved by the authors.

#### Country-level moderators

To examine whether country-level characteristics moderated between-country variation in effect sizes, we conducted meta-regression analyses. Overall, we found little evidence of moderation by country-level factors. The only evidence of moderation was observed for the association between arts engagement and gratitude: this positive association tended to be stronger in countries with higher national levels of trust (coef=0.07, 95%CI=0.01, 0.13) (**Figure 5**). See **Supplementary Figure S13** for the full meta-regression results.

**Figure 5.**
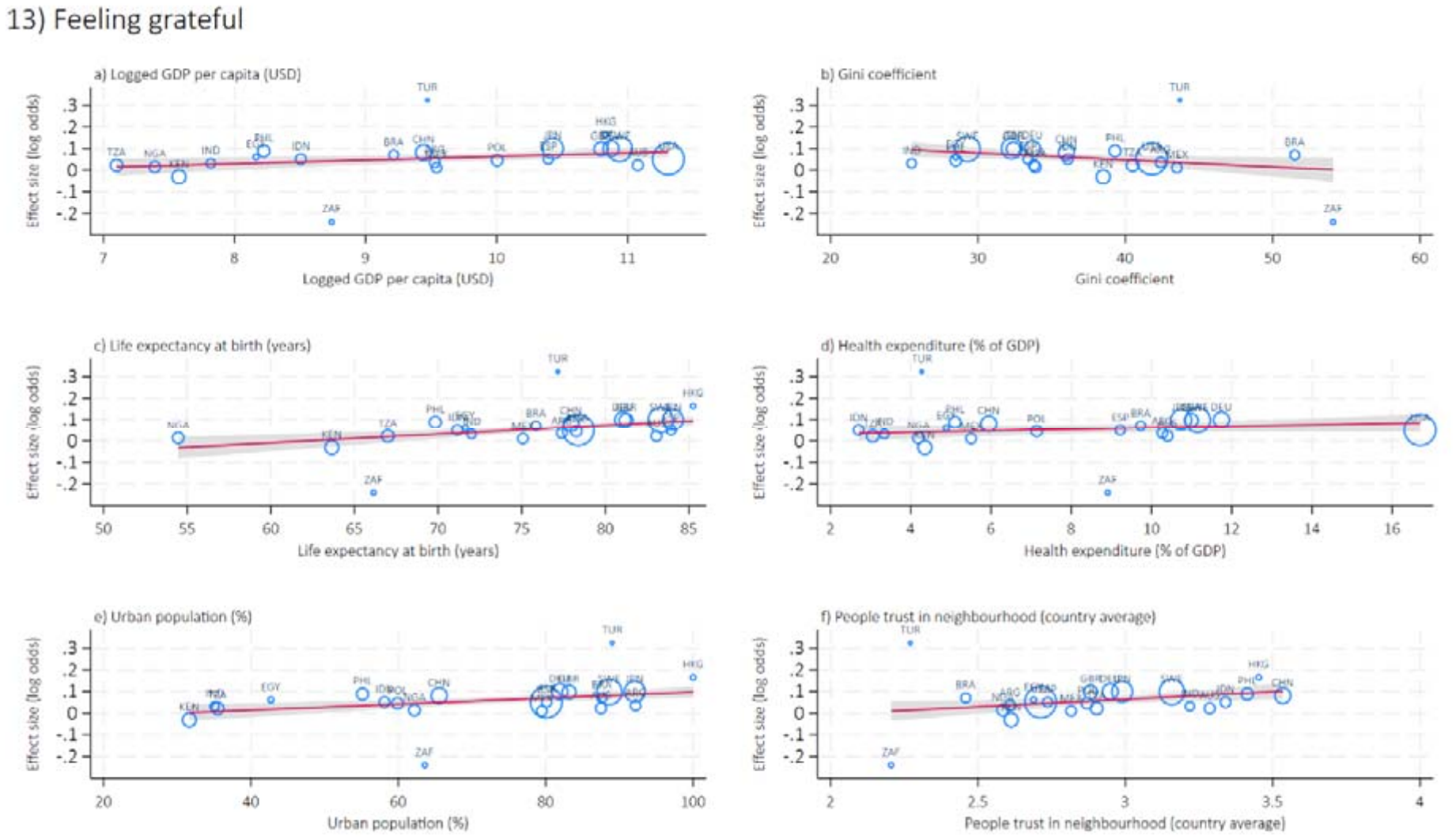
Bubble plot for meta-regression analyses examining the moderating effect of country-level factors on the association between arts engagement and gratitude (measured by feeling grateful). Bubble plots for all meta-regression outcomes are presented in the Supplementary Figure S13.

## Discussion

Across 22 countries and territories spanning the Global South, East Asia, Western Europe, the Middle East, and North America, our findings suggest that the positive effects of arts engagement on health and wellbeing are broadly universal. The effects were most consistently observed for **positive wellbeing and character strength**, including understanding purpose in life, feeling happy, grateful, and at peace; **evaluative wellbeing**, including life satisfaction, current and anticipated life evaluation, and feeling that life is worthwhile; and **social and general health**, including lower levels of loneliness and higher levels of overall physical health. By contrast, the effects of the arts on future-oriented wellbeing and perceived capability were less robust, and no meta-analytic effects across countries were observed for psychological distress (both individual symptoms and as a composite measure). The pattern of findings suggests that arts engagement may be most cross-culturally relevant to the promotion of positive mental health and wellbeing, and potential effects on the reduction of psychological distress may be more culturally specific. This distinction is consistent with conceptualisations of positive health and positive mental health in particular in which wellbeing extends beyond the absence of disease symptoms.^50,51^ Arts engagement may therefore represent a health-promoting resource that contributes to flourishing beyond the prevention or alleviation of psychological ill-health. When examining whether country-level characteristics moderated the effects of arts on the outcomes across countries, we found little evidence. The only exception was the modest moderating effect for feeling grateful: the effect of arts engagement was stronger in countries with higher national levels of trust. To our knowledge, this is the first study to compare the effects of arts engagement on health and wellbeing directly across a diverse range of countries using meta-analysis approach and to test for potential cross-country moderation. In doing so, the study addresses three important gaps in the arts-and-health literature. First, it demonstrates that the health and wellbeing effects of arts engagement are not confined to Western, high-income settings, which have historically dominated the evidence base. Second, it provides a direct test of cross-national consistency, rather than drawing conclusions about universality by synthesising findings from separate single-country studies. Third, it provides an initial basis for understanding whether, and to what extent, country-level characteristics may moderate the effects of arts on health and wellbeing.

Previous studies have suggested that arts engagement may help to prevent the onset of illnesses, support symptom management, and promote population health.^52,53^ The present study extends this literature by showing that the most consistent and strongest cross-national evidence appears to be across domains of positive wellbeing and character strength, evaluative wellbeing, and social and general health,^54^ even in countries with a lower arts engagement rate such as Egypt and India. These domains are important outcomes in their own right, supporting the view that arts engagement may function as a health-promoting behaviour.^19,22^ The consistently positive associations between arts engagement and wellbeing across countries are compatible with Keye’s dual-continuum model, which conceptualises positive mental health and mental illness as related but distinct dimensions.^55^ Improvements in wellbeing may therefore not necessarily correspond to reductions in psychological distress. However, methodological factors may also contribute to this pattern. Participants reporting no or very few symptoms at baseline may have limited scope for further reductions in distress, resulting in a floor effect. We attempted to explore this possibility, but the models did not converge, leaving this explanation unconfirmed. At the same time, these domains may also operate as potential mechanisms through which arts engagement influences downstream health outcomes and more distal clinical outcomes such as psychiatric morbidity, physical illness, and mortality.^14^ Treating these as potential mechanisms may help identify the active ingredients of arts engagement^13^ and the extent to which these may be context specific. For example, in more collectivist settings, the ‘social’ active ingredients of an activity (e.g. social interaction, shared participation and experiences, cooperation) may operate more strongly, and therefore activating the social pathway, such as trust, belonging, and reduced loneliness, which may in turn contribute to better health outcomes. This possibility is illustrated by Hong Kong SAR, a relatively collectivist cultural context in which arts engagement was strongly associated with lower levels of loneliness. However, the potential of arts engagement on health and wellbeing may extend beyond these domains. Notably, when analyses were restricted to participants who already engaged in the arts, some associations were attenuated, whereas others became more apparent. For instance, amongst participants who engaged in the arts, more frequent engagement was associated with higher levels of feeling very capable in most things in life in the later wave. This finding is consistent with the theory of emotion regulation through the arts, whereby people may use artistic activities for self-development and build confidence in their ability to approach challenges.^56^ In contrast, for some outcomes (e.g. understanding purpose in life and feeling that life is worthwhile), initiating arts engagement may be sufficient to generate potential health benefits, with limited additional gains from engaging more frequently thereafter.

In this study, we explicitly framed our research question causally,^31^ with the aim of estimating the effects of arts engagement on subsequent health and wellbeing. Several approaches were used to strengthen the plausibility of a causal interpretation, which all pointed to very similar results. First, we analysed data on arts engagement from an earlier wave to establish temporal ordering. Second, models adjusted for a broad range of theoretically and empirically established covariates to reduce confounding. Third, adjustment for baseline outcomes helped account for pre-existing differences in health and wellbeing and reduce concerns about reverse causation. Fourth, consistency of the effects observed across countries helped provider stronger evidence of causality by showing that the effects of arts engagement on health and wellbeing were seen across populations in which the underlying confounding structures such as the social, cultural, economic, and healthcare environments are very different.^57^ Finally, sensitivity analysis restricted to participants who were below the healthiest level of each outcome at baseline suggested that the estimated effects of arts engagement may contribute to later improvements amongst individuals with scope to benefit. Together, these findings suggest the plausibility of a health-enhancing effect of arts engagement, and align with other multi-national studies that have triangulated across different statistical approaches to strengthen causal inference.^58^ Yet, despite these strengthens, further triangulation using various and complementary analytical approaches is still needed to strengthen confidence in the causal interpretation, including counter-factual causal inference approaches.

Whilst some effects of arts engagement were broadly consistent across countries and aligned with patterns reported in previous studies, others varied across countries in both magnitude and direction. Poland and the United States were particularly notable, with estimated effects in one or both countries showing in the opposite direction to those reported in the existing literature. Although the reasons for these findings remain uncertain, we had undertaken an ecological approach to systematically examine the potential role of country-level moderators, capturing national economic conditions, income inequality, urbanisation, population health, healthcare expenditure, and interpersonal trust. However, evidence of moderation was limited. These findings can be interpreted from both methodological and conceptual perspectives. Methodologically, this may partly reflect the relatively small number of countries and territories included in the analysis, which constrained statistical power to detect any interactions. Contextually, the findings point to three possible interpretations. First, the associations may be broadly consistent across countries, as indicated in most of the outcomes identified in the meta-analysis, and hence country-level factors only had minimal moderating effects. Second, other relevant country-level factors (e.g. national cultural policies and infrastructure supporting cultural participation) may remain unidentified and require further investigation. One important factor not considered in the present analysis, and one that is difficult to quantify, is the cultural meanings attached to arts engagement and to mental health and wellbeing. Although standardised questions in a cross-national survey enable direct comparison, they also assume that the same items carry equivalent meanings across cultural and language settings. As a result, they may not fully capture culturally specific forms of arts participation or dimensions of health and wellbeing, while apparently identical survey items may remain polysemous across societies.^59^ This may help explain why associations between arts and mental health and wellbeing were identified in single-country studies using nationally developed cohort data,^25^ yet were weaker or absent when they were assessed using standardised cross-national questions. Third, participation rates, motivation for, and the social patterning of arts engagement varied across countries.^23,24,26^ In some countries, especially high-income countries,^24^ participation may be more concentrated amongst those with higher levels of education and income. Elsewhere, arts engagement may be more socially widespread and embedded in community life (e.g. 87% of people living in Nigeria engaged in the arts and 86% in Australia).^23,24,26^ This embedding could lead to health benefits arising as an indirect outcome of participation, whereas in countries where arts engagement is more explicitly understood and promoted as a health-promoting behaviour, alongside physical activity, sleep and diet, and undertaken with an intention of maintaining or improving health and wellbeing,^19,22^ there could potentially be increased engagement amongst people experiencing poor health or psychological distress, which could increase the risks of reverse causality. Overall, this suggests that, in cross-national research, comparing estimates derived from standardised measures is only part of the evidence jigsaw required, and that measurement equivalence, cultural context and baseline population differences need to be considered through triangulation of findings with insight from other methodologies including qualitative research.

This study had several strengths. It drew on large-scale, quota-representative samples from 22 countries and regions, collectively covering more than half of the world’s population. Survey questions were standardised and administered in local languages, facilitating direct comparison across countries. The analysis also used meta-analysis to assess the consistency of associations across countries and meta-regression to examine the extent to which country-level factors might moderate the associations. However, several limitations should also be acknowledged. First, although the study framed its research questions causally and applied several causal inference approaches, future research should seek to strength confidence in the causal interpretation by replicating these findings in longitudinal cohort data and applying complementary causal inference methods to further establish temporality and assess the robustness of the estimated effects found in this study. Second, the study lacked granular measures of the nature and pattern of arts exposure. We were unable to determine whether arts engagement was the primary activity or occurred alongside other activities, such as religious or community participation; to distinguish between different forms of engagement, such as active participation, receptive engagement, group-based activities; or to assess its consistency over time. Consequently, we could not examine whether associations with mental health and wellbeing varied according to the type, context, intensity or length of arts engagement. Third, further work is needed to strengthen the measurement of cultural engagement measurement and to examine the broader contextual influences and mechanisms that may shape its associations with health and wellbeing. With respective to measurement, countries may differ in what is recognised as ‘arts engagement’, in the prevalence and social patterning of such engagement, and in how these definitions and practices evolve over time. Future research should therefore develop measures that are sufficiently flexible to capture the fluid and context-specific nature of cultural engagement, while remaining harmonizable enough to support meaningful cross-country comparison. In relation to contextual influences, while our study examined several country-level indicators, future research should identify a broader range of ecological determinants, including access to the arts, cultural infrastructure, opportunities for engagement, and structural barriers to engagement. With respective to mechanisms, there is growing interest in how arts engagement may influence health outcomes, including emerging research on epigenetic and physiological ageing ^7,60^ and a review identifying more than 50 interconnected mechanisms.^20^ Further research should examine the mechanisms underlying the associations observed in the study, whether these pathways operate consistently across cultural contexts, and whether some of the outcomes identified here may themselves function as intermediate mechanisms linking arts engagement to more distal clinical outcomes.

In conclusion, this study provides important evidence that there are effects of arts engagement on positive wellbeing and character strength, evaluative wellbeing, and social and general health, but not psychological distress, with these effects remaining broadly consistent across diverse countries and cultural settings. The presence of these effects across societies and populations with varying country-level characteristics is encouraging. However, the variation observed between countries also highlights the complexity of determining whether the health impacts of arts engagement are universal and raise questions about how arts engagement is conceptualised across settings. Arts engagement may function both as an everyday cultural, social or leisure practice with indirect effects on health and as an intentional health-promoting behaviour. Addressing this question will require culturally grounded cross-national research that combines harmonised measures with in-depth qualitative research, capturing local meanings, motivations, and practices while retaining the capacity for systematic comparison across countries. Such research is encouraged to understand how the meaning and contexts of arts engagement shape its relationship with health and wellbeing across societies.

## Data Availability

The data are publicly available upon preregistration through the Center for Open sciences (COS; Johnson et al., 2024, https://osf.io/3jtz8/).

## Authors contributions

HWM and DF designed the study. HWM conducted the analyses, and drafted the paper with support from KL and DF. All authors contributed to the writing, made critical revisions, and approved the final paper.

## Funding & Acknowledgements

This paper was also supported by Wellcome Trust Discovery Award (GRACE-Epi: 326117/Z/25/Z). It received supplementary support from the EpiArts Lab, a National Endowment for the Arts Research Lab at the University of Florida, which is supported in part by an award from the National Endowment for the Arts (1862896–38-C-20). The opinions expressed are those of the authors and do not represent the views of the National Endowment for the Arts Office of Research & Analysis or the National Endowment for the Arts. The National Endowment for the Arts does not guarantee the accuracy or completeness of the information included in this material and is not responsible for any consequences of its use. The EpiArts Lab is also supported by the University of Florida, Americans for the Arts, the Pabst Steinmetz Foundation, and Bloomberg Philanthropies. In addition, HWM was supported by the Jameel Arts & Health Lab Global Population Health Fellowship. DWB was supported by the National Science Centre, Poland [2024/55/B/HS6/00041]. The Global Flourishing Study was supported by funding from the John Templeton Foundation (grant no. 61665, B.R.J., T.J.V.), Templeton Religion Trust (no. 1308, B.R.J., T.J.V.), Templeton World Charity Foundation (no. 0605, B.R.J., T.J.V.), Well-Being for Planet Earth Foundation, Fetzer Institute (no. 4354, B.R.J., T.J.V.), Well Being Trust, Paul L. Foster Family Foundation and the David and Carol Myers Foundation. The opinions expressed in this publication are those of the authors and do not necessarily reflect the views of these organizations.

