## Supplementary for "The health and wellbeing impact of arts engagement: A longitudinal outcome-wide approach in 22 countries"

Supplementary Figure S5: Meta-analysis forest plots for the effects of arts engagement on later health and wellbeing outcomes (nested adjusted only for socio-demographic confounders and baseline health outcome) 12

Supplementary Table S1: Baseline characteristics of analytical sample from weighted data.

| Country | N | Age, mean (SD) | Arts engagement, % | Female, % | Native-born, % | Married, % | Urban residence, % | University degree, % | Employed, % |
|---|---|---|---|---|---|---|---|---|---|
| Argentina | 2,696 | 47.5 (17.1) | 58.5 | 52.9 | 95.6 | 46.3 | 65.6 | 10.0 | 61.3 |
| Australia | 2,457 | 49.6 (18.1) | 86.2 | 49.2 | 79.1 | 58.1 | 72.0 | 31.0 | 59.2 |
| Brazil | 3,864 | 46.3 (0.0) | 67.4 | 51.2 | 98.7 | 53.1 | 64.2 | 22.7 | 50.8 |
| China | 4,536 | 44.2 (15.4) | 80.0 | 50.0 | 99.8 | 72.0 | 58.4 | 9.4 | 63.9 |
| Egypt | 3,363 | 36.2 (0.0) | 12.9 | 46.5 | 99.7 | 67.7 | 33.7 | 11.4 | 47.4 |
| Germany | 4,079 | 50.9 (16.8) | 74.9 | 48.7 | 93.3 | 59.8 | 47.2 | 35.5 | 60.5 |
| Hong Kong SAR | 681 | 50.0 (14.3) | 62.4 | 56.6 | 90.7 | 62.4 | 92.8 | 31.7 | 73.8 |
| India | 8,079 | 37.4 (0.0) | 40.0 | 46.5 | 99.0 | 78.6 | 23.2 | 4.9 | 49.1 |
| Indonesia | 2,644 | 38.9 (13.9) | 42.8 | 49.1 | 99.5 | 69.3 | 27.7 | 4.5 | 51.6 |
| Japan | 13,341 | 54.3 (16.1) | 41.6 | 51.4 | 99.4 | 62.3 | 44.5 | 22.4 | 61.9 |
| Kenya | 9,062 | 35.7 (15.1) | 77.4 | 51.2 | 99.0 | 58.3 | 11.6 | 6.1 | 42.6 |
| Mexico | 2,056 | 45.4 (16.5) | 62.0 | 52.2 | 98.2 | 57.4 | 49.5 | 25.5 | 52.5 |
| Nigeria | 4,889 | 34.8 (13.9) | 86.6 | 50.2 | 99.3 | 58.9 | 40.8 | 1.6 | 65.4 |
| Philippines | 3,426 | 40.1 (0.0) | 62.8 | 51.6 | 99.9 | 68.5 | 40.4 | 6.0 | 51.3 |
| Poland | 4,225 | 48.1 (17.1) | 55.1 | 52.9 | 99.5 | 66.0 | 37.1 | 28.7 | 58.7 |
| South Africa | 1,601 | 39.3 (15.6) | 63.3 | 53.9 | 93.8 | 31.5 | 32.7 | 4.8 | 32.6 |
| Spain | 2,183 | 51.1 (14.5) | 79.7 | 49.5 | 89.8 | 63.8 | 54.2 | 23.0 | 54.0 |
| Sweden | 11,347 | 51.1 (17.8) | 81.7 | 51.5 | 93.3 | 64.6 | 45.6 | 25.4 | 60.9 |
| Tanzania | 6,557 | 36.3 (15.6) | 47.6 | 51.7 | 99.8 | 63.4 | 25.1 | 0.9 | 55.6 |
| Türkiye | 653 | 44.2 (0.0) | 64.1 | 50.4 | 94.5 | 67.7 | 72.9 | 19.5 | 43.1 |
| United Kingdom | 3,273 | 52.3 (16.5) | 71.8 | 52.1 | 88.2 | 58.3 | 44.0 | 36.0 | 60.8 |
| United States | 31,378 | 52.5 (16.3) | 80.8 | 52.2 | 92.6 | 63.2 | 58.8 | 40.2 | 59.8 |

Note: Sample sizes are unweighted. All characteristics were measured at Wave 1, and descriptive estimates were weighted using the Wave 3 sampling weight applied in the regression analyses.

Supplementary Figure S1: A simplified DAG illustrating the assumed causal structure underlying the outcome-wide analysis

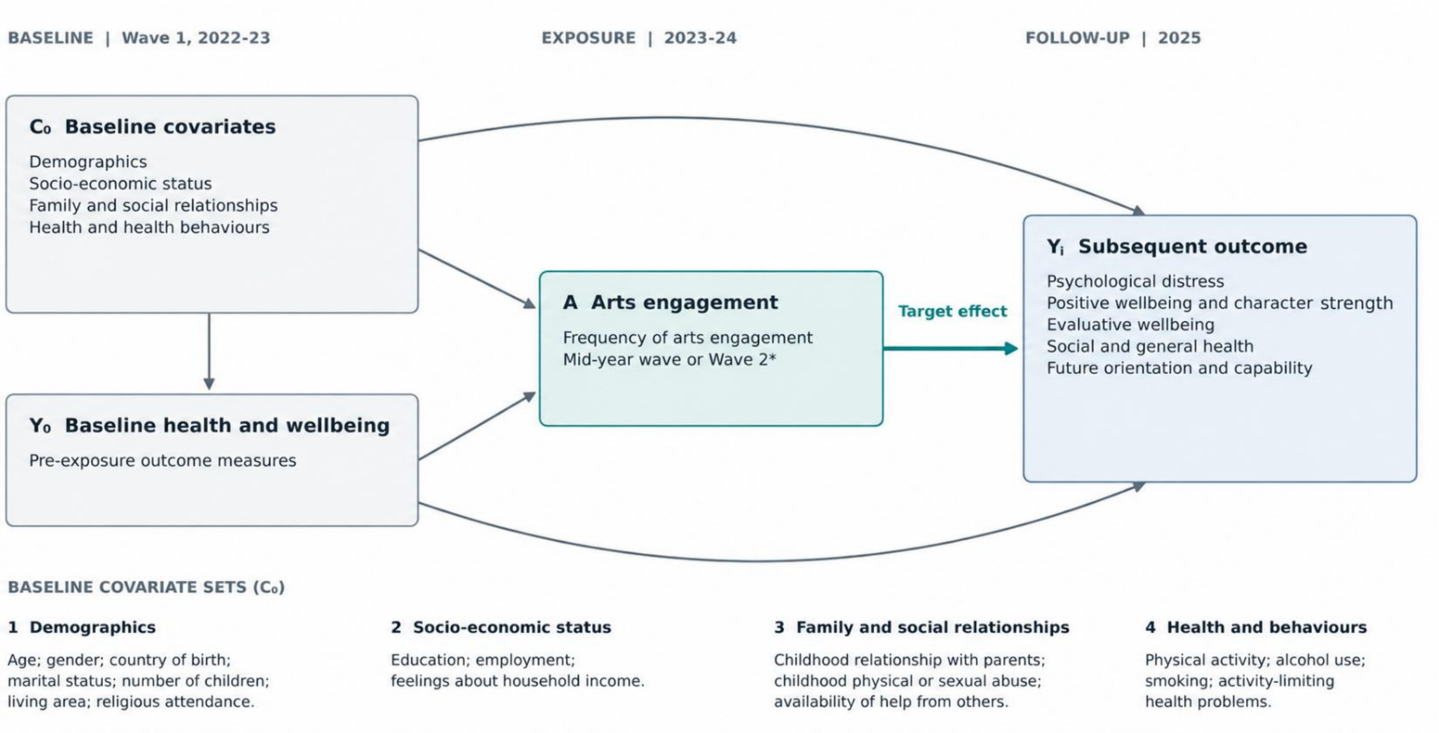

Note: Arrows represent hypothesised causal directions. Each $Y_j$ was analysed separately (19 outcomes across five domains). *Exposure measured in Wave 2 (2024) in some countries. Childhood variables refer to earlier life but were reported at baseline. The figure was created using ChatGPT (OpenAI) and reviewed for accuracy and approved by the authors.

Supplementary Figure S2: Findings from Israel based on the fully adjusted model, excluding childhood abuse experiences as a covariate due to data unavailability.

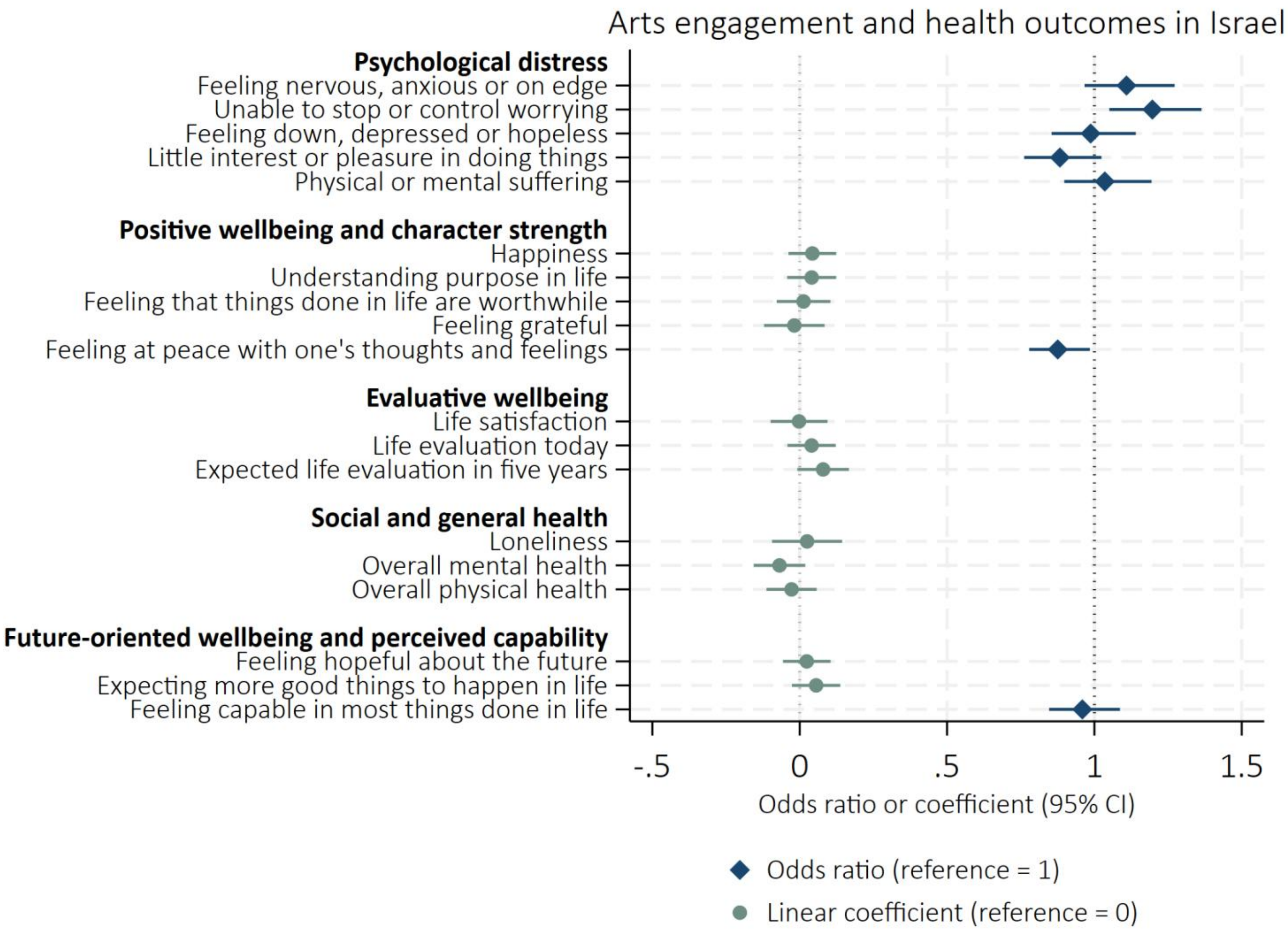

#### Supplementary Figure S3: Meta-analysis forest plots for the effects of arts engagement on later health and wellbeing outcomes (nested unadjusted models)

a) Feeling nervous, anxious, or on edge

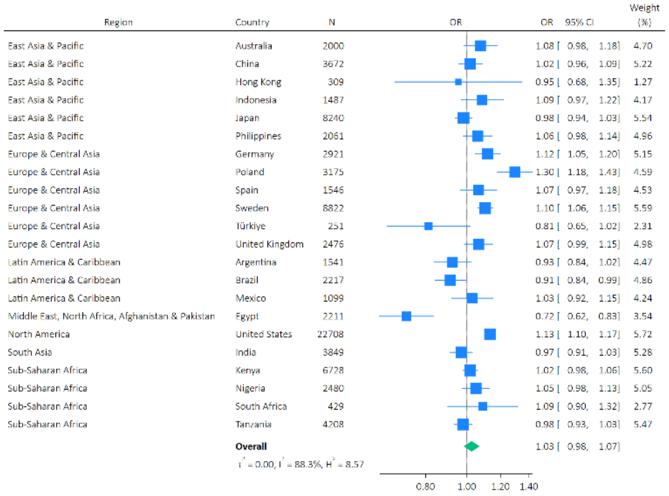

b) Feeling down, depressed or hopeless

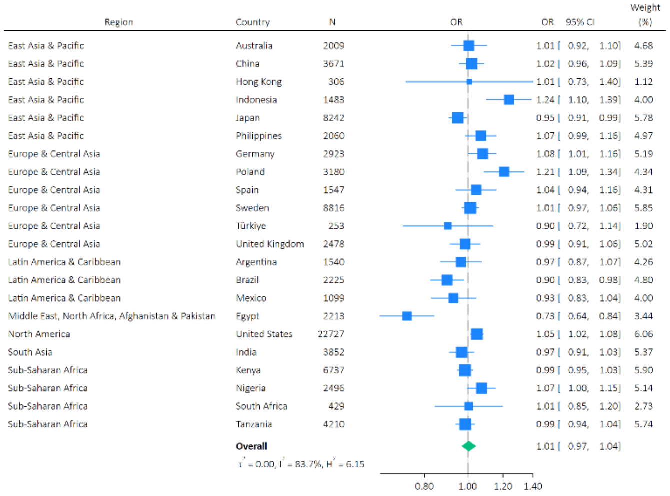

c) Unable to stop or control worrying

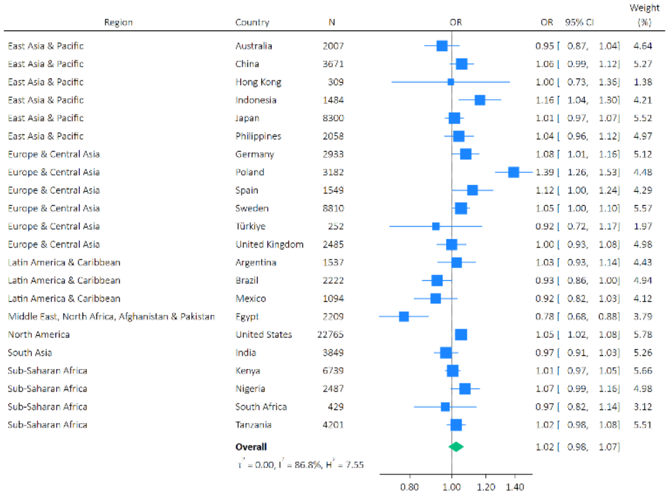

d) Little interest or pleasure in doing things

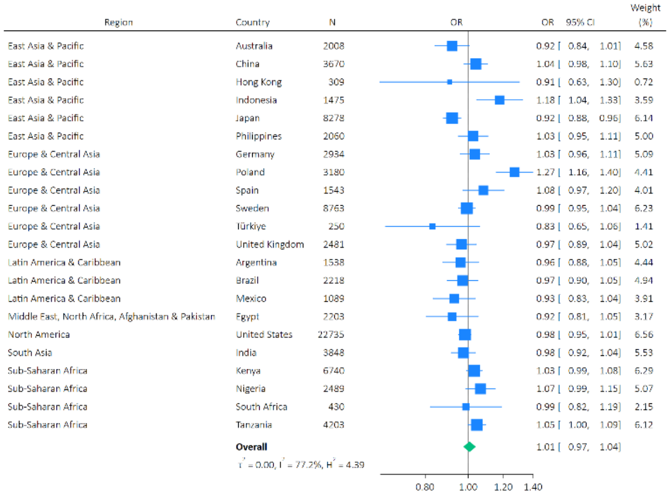

e) Suffering

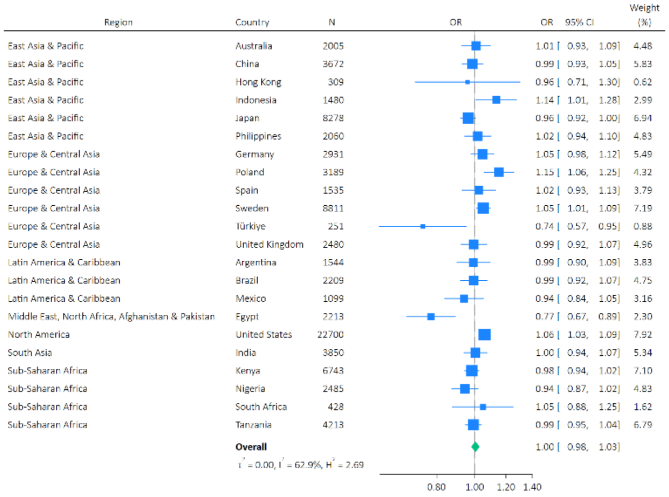

f) Feeling happy

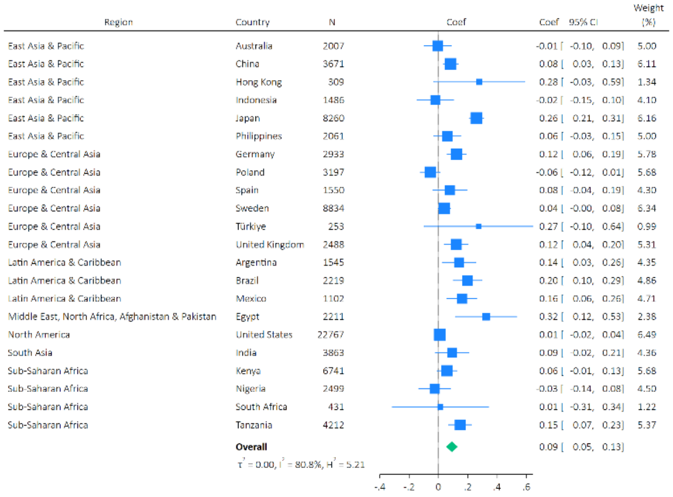

g) Understanding purpose in life

| Region | Country | N | Coef | 95% CI | Weight (%) |
|---|---|---|---|---|---|
| East Asia & Pacific | Australia | 2008 | 0.04 | [-0.10, 0.17] | 4.07 |
| East Asia & Pacific | China | 3670 | 0.11 | [0.05, 0.17] | 5.78 |
| East Asia & Pacific | Hong Kong | 309 | 0.31 | [-0.02, 0.64] | 1.47 |
| East Asia & Pacific | Indonesia | 1487 | 0.04 | [-0.09, 0.17] | 4.20 |
| East Asia & Pacific | Japan | 8262 | 0.30 | [0.25, 0.35] | 5.88 |
| East Asia & Pacific | Philippines | 2060 | 0.01 | [-0.07, 0.09] | 5.32 |
| Europe & Central Asia | Germany | 2931 | 0.06 | [-0.02, 0.14] | 5.35 |
| Europe & Central Asia | Poland | 3187 | -0.11 | [-0.20, -0.02] | 5.08 |
| Europe & Central Asia | Spain | 1543 | 0.10 | [-0.01, 0.22] | 4.44 |
| Europe & Central Asia | Sweden | 8766 | 0.05 | [-0.00, 0.11] | 5.85 |
| Europe & Central Asia | Türkiye | 252 | 0.32 | [-0.10, 0.74] | 0.98 |
| Europe & Central Asia | United Kingdom | 2478 | 0.10 | [-0.01, 0.21] | 4.67 |
| Latin America & Caribbean | Argentina | 1540 | 0.09 | [-0.03, 0.21] | 4.33 |
| Latin America & Caribbean | Brazil | 2216 | 0.20 | [0.09, 0.30] | 4.84 |
| Latin America & Caribbean | Mexico | 1099 | 0.16 | [0.06, 0.27] | 4.80 |
| Middle East, North Africa, Afghanistan & Pakistan | Egypt | 2193 | 0.20 | [0.05, 0.34] | 3.82 |
| North America | United States | 22735 | 0.07 | [0.03, 0.12] | 6.02 |
| South Asia | India | 3849 | 0.13 | [0.03, 0.23] | 4.85 |
| Sub-Saharan Africa | Kenya | 6740 | 0.05 | [-0.02, 0.12] | 5.53 |
| Sub-Saharan Africa | Nigeria | 2494 | -0.03 | [-0.13, 0.07] | 4.83 |
| Sub-Saharan Africa | South Africa | 431 | -0.22 | [-0.44, 0.00] | 2.53 |
| Sub-Saharan Africa | Tanzania | 4176 | 0.19 | [0.11, 0.27] | 5.37 |
| | **Overall** | | 0.09 | [0.05, 0.14] | |

$\tau^2 = 0.00$, $I^2 = 81.5\%$, $H^2 = 5.40$

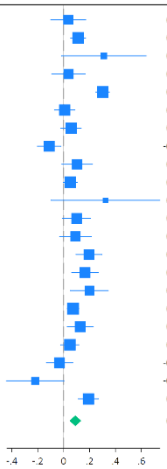

h) Life satisfaction

| Region | Country | N | Coef | 95% CI | Weight (%) |
|---|---|---|---|---|---|
| East Asia & Pacific | Australia | 2010 | 0.02 | [-0.08, 0.13] | 4.75 |
| East Asia & Pacific | China | 3652 | 0.09 | [0.03, 0.14] | 6.07 |
| East Asia & Pacific | Hong Kong | 297 | 0.39 | [0.03, 0.76] | 1.10 |
| East Asia & Pacific | Indonesia | 1484 | -0.03 | [-0.16, 0.09] | 4.24 |
| East Asia & Pacific | Japan | 8226 | 0.27 | [0.21, 0.32] | 6.07 |
| East Asia & Pacific | Philippines | 2059 | 0.01 | [-0.08, 0.10] | 5.09 |
| Europe & Central Asia | Germany | 2913 | 0.11 | [0.04, 0.18] | 5.77 |
| Europe & Central Asia | Poland | 3180 | -0.09 | [-0.17, -0.02] | 5.55 |
| Europe & Central Asia | Spain | 1542 | 0.10 | [-0.02, 0.22] | 4.34 |
| Europe & Central Asia | Sweden | 8822 | 0.02 | [-0.02, 0.06] | 6.30 |
| Europe & Central Asia | Türkiye | 253 | 0.01 | [-0.39, 0.40] | 0.95 |
| Europe & Central Asia | United Kingdom | 2459 | 0.16 | [0.07, 0.24] | 5.27 |
| Latin America & Caribbean | Argentina | 1544 | 0.12 | [0.00, 0.25] | 4.26 |
| Latin America & Caribbean | Brazil | 2219 | 0.14 | [0.04, 0.25] | 4.71 |
| Latin America & Caribbean | Mexico | 1102 | 0.10 | [-0.02, 0.21] | 4.48 |
| Middle East, North Africa, Afghanistan & Pakistan | Egypt | 2199 | 0.18 | [0.03, 0.33] | 3.63 |
| North America | United States | 22756 | -0.00 | [-0.04, 0.03] | 6.45 |
| South Asia | India | 3859 | 0.13 | [0.02, 0.24] | 4.57 |
| Sub-Saharan Africa | Kenya | 6744 | 0.03 | [-0.04, 0.11] | 5.54 |
| Sub-Saharan Africa | Nigeria | 2499 | -0.05 | [-0.17, 0.07] | 4.34 |
| Sub-Saharan Africa | South Africa | 426 | -0.09 | [-0.40, 0.23] | 1.40 |
| Sub-Saharan Africa | Tanzania | 4211 | -0.01 | [-0.10, 0.08] | 5.11 |
| | **Overall** | | 0.07 | [0.03, 0.11] | |

$\tau^2 = 0.00$, $I^2 = 79.8\%$, $H^2 = 4.94$

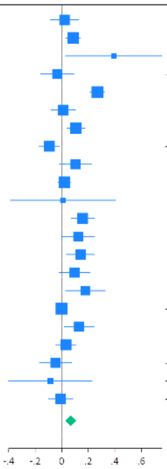

i) Life evaluation: Today

| Region | Country | N | Coef | 95% CI | Weight (%) |
|---|---|---|---|---|---|
| East Asia & Pacific | Australia | 2011 | -0.00 | [-0.10, 0.09] | 4.96 |
| East Asia & Pacific | China | 3671 | 0.06 | [0.01, 0.12] | 5.82 |
| East Asia & Pacific | Hong Kong | 309 | 0.33 | [0.06, 0.60] | 1.77 |
| East Asia & Pacific | Indonesia | 1485 | 0.04 | [-0.10, 0.18] | 3.75 |
| East Asia & Pacific | Japan | 8293 | 0.27 | [0.22, 0.32] | 5.99 |
| East Asia & Pacific | Philippines | 2056 | 0.13 | [0.04, 0.22] | 5.03 |
| Europe & Central Asia | Germany | 2928 | 0.20 | [0.13, 0.26] | 5.71 |
| Europe & Central Asia | Poland | 3195 | 0.04 | [-0.03, 0.11] | 5.51 |
| Europe & Central Asia | Spain | 1550 | 0.09 | [-0.02, 0.20] | 4.49 |
| Europe & Central Asia | Sweden | 8831 | 0.03 | [-0.00, 0.07] | 6.18 |
| Europe & Central Asia | Türkiye | 252 | 0.31 | [-0.04, 0.66] | 1.17 |
| Europe & Central Asia | United Kingdom | 2485 | 0.18 | [0.09, 0.26] | 5.16 |
| Latin America & Caribbean | Argentina | 1547 | 0.16 | [0.05, 0.28] | 4.37 |
| Latin America & Caribbean | Brazil | 2226 | 0.15 | [0.05, 0.24] | 4.85 |
| Latin America & Caribbean | Mexico | 1102 | 0.15 | [0.04, 0.25] | 4.64 |
| Middle East, North Africa, Afghanistan & Pakistan | Egypt | 2201 | 0.43 | [0.26, 0.59] | 3.16 |
| North America | United States | 22777 | 0.03 | [-0.00, 0.06] | 6.28 |
| South Asia | India | 3842 | 0.13 | [0.02, 0.24] | 4.45 |
| Sub-Saharan Africa | Kenya | 6744 | 0.07 | [-0.00, 0.14] | 5.53 |
| Sub-Saharan Africa | Nigeria | 2503 | -0.03 | [-0.15, 0.08] | 4.37 |
| Sub-Saharan Africa | South Africa | 429 | -0.09 | [-0.39, 0.21] | 1.50 |
| Sub-Saharan Africa | Tanzania | 4215 | 0.17 | [0.09, 0.25] | 5.29 |
| | **Overall** | | 0.12 | [0.08, 0.16] | |

$\tau^2 = 0.00$, $I^2 = 83.3\%$, $H^2 = 5.99$

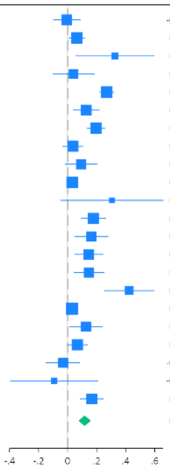

j) Life evaluation: Five years from now

| Region | Country | N | Coef | 95% CI | Weight (%) |
|---|---|---|---|---|---|
| East Asia & Pacific | Australia | 2010 | 0.15 | [0.05, 0.25] | 4.79 |
| East Asia & Pacific | China | 3653 | 0.06 | [0.01, 0.11] | 5.63 |
| East Asia & Pacific | Hong Kong | 300 | 0.51 | [0.23, 0.80] | 1.94 |
| East Asia & Pacific | Indonesia | 1467 | 0.12 | [0.02, 0.22] | 4.77 |
| East Asia & Pacific | Japan | 8297 | 0.29 | [0.24, 0.34] | 5.64 |
| East Asia & Pacific | Philippines | 2057 | 0.04 | [-0.04, 0.12] | 5.12 |
| Europe & Central Asia | Germany | 2912 | 0.29 | [0.22, 0.36] | 5.37 |
| Europe & Central Asia | Poland | 3153 | 0.06 | [-0.03, 0.15] | 5.04 |
| Europe & Central Asia | Spain | 1540 | 0.13 | [-0.01, 0.26] | 4.16 |
| Europe & Central Asia | Sweden | 8806 | 0.07 | [0.03, 0.11] | 5.79 |
| Europe & Central Asia | Türkiye | 236 | 0.44 | [-0.03, 0.92] | 0.89 |
| Europe & Central Asia | United Kingdom | 2460 | 0.25 | [0.16, 0.34] | 5.03 |
| Latin America & Caribbean | Argentina | 1510 | 0.16 | [0.04, 0.28] | 4.40 |
| Latin America & Caribbean | Brazil | 2216 | 0.10 | [0.02, 0.19] | 5.08 |
| Latin America & Caribbean | Mexico | 1078 | 0.23 | [0.12, 0.34] | 4.61 |
| Middle East, North Africa, Afghanistan & Pakistan | Egypt | 1632 | 0.42 | [0.22, 0.62] | 3.02 |
| North America | United States | 22765 | 0.02 | [-0.02, 0.05] | 5.83 |
| South Asia | India | 3576 | 0.13 | [0.03, 0.22] | 4.87 |
| Sub-Saharan Africa | Kenya | 6699 | 0.09 | [0.02, 0.15] | 5.42 |
| Sub-Saharan Africa | Nigeria | 2470 | -0.05 | [-0.14, 0.04] | 5.02 |
| Sub-Saharan Africa | South Africa | 423 | -0.08 | [-0.33, 0.17] | 2.32 |
| Sub-Saharan Africa | Tanzania | 3979 | 0.19 | [0.11, 0.26] | 5.25 |
| | **Overall** | | 0.14 | [0.09, 0.19] | |

$\tau^2 = 0.00$, $I^2 = 87.5\%$, $H^2 = 8.02$

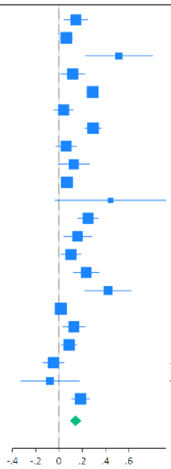

k) Feeling lonely

| Region | Country | N | Coef | 95% CI | Weight (%) |
|---|---|---|---|---|---|
| East Asia & Pacific | Australia | 2010 | 0.07 | [-0.07, 0.20] | 4.64 |
| East Asia & Pacific | China | 3672 | -0.06 | [-0.13, 0.01] | 6.67 |
| East Asia & Pacific | Hong Kong | 308 | -0.28 | [-0.68, 0.13] | 1.03 |
| East Asia & Pacific | Indonesia | 1487 | 0.06 | [-0.13, 0.26] | 3.14 |
| East Asia & Pacific | Japan | 8309 | -0.19 | [-0.25, -0.13] | 6.92 |
| East Asia & Pacific | Philippines | 2063 | -0.04 | [-0.16, 0.08] | 4.95 |
| Europe & Central Asia | Germany | 2938 | -0.01 | [-0.10, 0.08] | 6.00 |
| Europe & Central Asia | Poland | 3190 | 0.14 | [0.05, 0.23] | 5.93 |
| Europe & Central Asia | Spain | 1553 | 0.02 | [-0.15, 0.20] | 3.57 |
| Europe & Central Asia | Sweden | 8848 | 0.01 | [-0.04, 0.07] | 7.16 |
| Europe & Central Asia | Türkiye | 250 | -0.00 | [-0.45, 0.45] | 0.85 |
| Europe & Central Asia | United Kingdom | 2476 | -0.09 | [-0.21, 0.02] | 5.31 |
| Latin America & Caribbean | Argentina | 1549 | -0.18 | [-0.34, -0.01] | 3.70 |
| Latin America & Caribbean | Brazil | 2221 | -0.12 | [-0.26, 0.03] | 4.29 |
| Latin America & Caribbean | Mexico | 1099 | -0.20 | [-0.38, -0.02] | 3.42 |
| Middle East, North Africa, Afghanistan & Pakistan | Egypt | 2210 | 0.03 | [-0.28, 0.34] | 1.62 |
| North America | United States | 22786 | -0.02 | [-0.07, 0.02] | 7.35 |
| South Asia | India | 3863 | 0.12 | [-0.01, 0.25] | 4.73 |
| Sub-Saharan Africa | Kenya | 6739 | -0.05 | [-0.13, 0.03] | 6.33 |
| Sub-Saharan Africa | Nigeria | 2496 | 0.08 | [-0.06, 0.21] | 4.64 |
| Sub-Saharan Africa | South Africa | 431 | 0.29 | [-0.02, 0.60] | 1.61 |
| Sub-Saharan Africa | Tanzania | 4213 | -0.08 | [-0.17, 0.00] | 6.13 |
| | **Overall** | | -0.03 | [-0.07, 0.02] | |

$\tau^2 = 0.00$, $I^2 = 70.8\%$, $H^2 = 3.42$

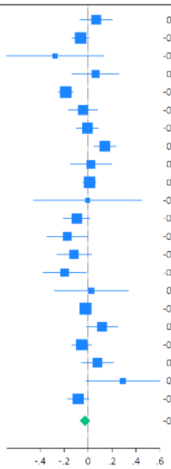

l) Life is worthwhile

| Region | Country | N | Coef | 95% CI | Weight (%) |
|---|---|---|---|---|---|
| East Asia & Pacific | Australia | 2008 | 0.04 | [-0.07, 0.16] | 4.47 |
| East Asia & Pacific | China | 3669 | 0.06 | [0.01, 0.11] | 5.90 |
| East Asia & Pacific | Hong Kong | 306 | 0.36 | [0.07, 0.66] | 1.69 |
| East Asia & Pacific | Indonesia | 1486 | 0.03 | [-0.08, 0.14] | 4.60 |
| East Asia & Pacific | Japan | 8290 | 0.32 | [0.26, 0.37] | 5.88 |
| East Asia & Pacific | Philippines | 2060 | -0.01 | [-0.10, 0.08] | 5.01 |
| Europe & Central Asia | Germany | 2932 | 0.12 | [0.05, 0.18] | 5.59 |
| Europe & Central Asia | Poland | 3189 | -0.14 | [-0.22, -0.07] | 5.39 |
| Europe & Central Asia | Spain | 1541 | 0.05 | [-0.08, 0.18] | 4.18 |
| Europe & Central Asia | Sweden | 8832 | 0.08 | [0.03, 0.13] | 5.97 |
| Europe & Central Asia | Türkiye | 253 | 0.22 | [-0.17, 0.61] | 1.09 |
| Europe & Central Asia | United Kingdom | 2473 | 0.16 | [0.08, 0.25] | 5.19 |
| Latin America & Caribbean | Argentina | 1541 | 0.14 | [0.03, 0.25] | 4.56 |
| Latin America & Caribbean | Brazil | 2224 | 0.18 | [0.08, 0.28] | 4.93 |
| Latin America & Caribbean | Mexico | 1101 | 0.11 | [0.00, 0.21] | 4.71 |
| Middle East, North Africa, Afghanistan & Pakistan | Egypt | 2207 | 0.10 | [-0.12, 0.31] | 2.58 |
| North America | United States | 22770 | 0.06 | [0.03, 0.10] | 6.09 |
| South Asia | India | 3844 | 0.03 | [-0.07, 0.13] | 4.81 |
| Sub-Saharan Africa | Kenya | 6743 | 0.08 | [0.01, 0.15] | 5.50 |
| Sub-Saharan Africa | Nigeria | 2498 | 0.03 | [-0.08, 0.14] | 4.62 |
| Sub-Saharan Africa | South Africa | 428 | -0.01 | [-0.27, 0.26] | 1.96 |
| Sub-Saharan Africa | Tanzania | 4209 | 0.20 | [0.12, 0.28] | 5.30 |
| | **Overall** | | 0.09 | [0.05, 0.14] | |

$\tau^2 = 0.00$, $I^2 = 83.1\%$, $H^2 = 5.92$

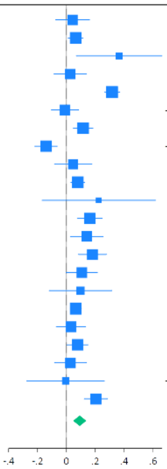

m) Feeling grateful

| Region | Country | N | Coef | 95% CI | Weight (%) |
|---|---|---|---|---|---|
| East Asia & Pacific | Australia | 1930 | 0.12 | [-0.00, 0.24] | 4.34 |
| East Asia & Pacific | China | 3670 | 0.12 | [0.07, 0.18] | 5.55 |
| East Asia & Pacific | Hong Kong | 308 | 0.52 | [0.21, 0.82] | 1.73 |
| East Asia & Pacific | Indonesia | 1482 | 0.06 | [-0.03, 0.15] | 4.95 |
| East Asia & Pacific | Japan | 8240 | 0.33 | [0.28, 0.39] | 5.61 |
| East Asia & Pacific | Philippines | 2061 | 0.07 | [-0.01, 0.15] | 5.18 |
| Europe & Central Asia | Germany | 2921 | 0.25 | [0.18, 0.32] | 5.31 |
| Europe & Central Asia | Poland | 3189 | 0.04 | [-0.05, 0.13] | 4.98 |
| Europe & Central Asia | Spain | 1547 | 0.15 | [0.03, 0.27] | 4.31 |
| Europe & Central Asia | Sweden | 8836 | 0.24 | [0.19, 0.28] | 5.71 |
| Europe & Central Asia | Türkiye | 246 | 0.10 | [-0.33, 0.53] | 1.02 |
| Europe & Central Asia | United Kingdom | 2445 | 0.24 | [0.15, 0.33] | 4.90 |
| Latin America & Caribbean | Argentina | 1543 | 0.03 | [-0.08, 0.14] | 4.56 |
| Latin America & Caribbean | Brazil | 2215 | 0.14 | [0.04, 0.24] | 4.73 |
| Latin America & Caribbean | Mexico | 1101 | 0.04 | [-0.05, 0.14] | 4.91 |
| Middle East, North Africa, Afghanistan & Pakistan | Egypt | 2181 | 0.06 | [-0.11, 0.22] | 3.51 |
| North America | United States | 21475 | 0.16 | [0.12, 0.20] | 5.81 |
| South Asia | India | 3837 | 0.06 | [-0.04, 0.16] | 4.72 |
| Sub-Saharan Africa | Kenya | 6744 | -0.00 | [-0.07, 0.06] | 5.40 |
| Sub-Saharan Africa | Nigeria | 2490 | 0.01 | [-0.08, 0.09] | 5.10 |
| Sub-Saharan Africa | South Africa | 428 | -0.21 | [-0.45, 0.02] | 2.38 |
| Sub-Saharan Africa | Tanzania | 4197 | 0.05 | [-0.02, 0.12] | 5.28 |
| | **Overall** | | 0.12 | [0.07, 0.16] | |

$\tau^2 = 0.00, I^2 = 85.7\%, H^2 = 7.00$

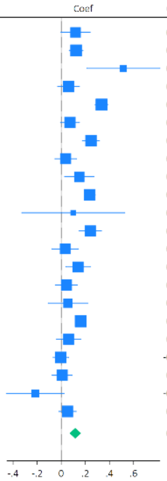

n) Feeling at peace

| Region | Country | N | Coef | 95% CI | Weight (%) |
|---|---|---|---|---|---|
| East Asia & Pacific | Australia | 2007 | -0.09 | [-0.19, 0.00] | 4.48 |
| East Asia & Pacific | China | 3669 | 0.04 | [-0.03, 0.10] | 5.69 |
| East Asia & Pacific | Hong Kong | 309 | 0.25 | [-0.11, 0.60] | 0.69 |
| East Asia & Pacific | Indonesia | 1485 | -0.06 | [-0.17, 0.05] | 4.00 |
| East Asia & Pacific | Japan | 8263 | 0.21 | [0.16, 0.26] | 6.32 |
| East Asia & Pacific | Philippines | 2061 | 0.02 | [-0.05, 0.10] | 5.20 |
| Europe & Central Asia | Germany | 2923 | 0.01 | [-0.07, 0.08] | 5.17 |
| Europe & Central Asia | Poland | 3166 | 0.16 | [0.07, 0.25] | 4.64 |
| Europe & Central Asia | Spain | 1541 | 0.07 | [-0.03, 0.17] | 4.13 |
| Europe & Central Asia | Sweden | 8824 | 0.03 | [-0.01, 0.08] | 6.34 |
| Europe & Central Asia | Türkiye | 253 | -0.01 | [-0.26, 0.24] | 1.30 |
| Europe & Central Asia | United Kingdom | 2479 | 0.06 | [-0.02, 0.14] | 4.94 |
| Latin America & Caribbean | Argentina | 1538 | -0.01 | [-0.12, 0.10] | 3.92 |
| Latin America & Caribbean | Brazil | 2219 | 0.07 | [-0.02, 0.16] | 4.72 |
| Latin America & Caribbean | Mexico | 1096 | 0.10 | [-0.01, 0.21] | 3.72 |
| Middle East, North Africa, Afghanistan & Pakistan | Egypt | 2210 | 0.15 | [-0.02, 0.31] | 2.35 |
| North America | United States | 22702 | 0.01 | [-0.02, 0.05] | 6.84 |
| South Asia | India | 3845 | 0.01 | [-0.06, 0.08] | 5.47 |
| Sub-Saharan Africa | Kenya | 6743 | 0.00 | [-0.04, 0.04] | 6.61 |
| Sub-Saharan Africa | Nigeria | 2493 | 0.01 | [-0.07, 0.09] | 4.99 |
| Sub-Saharan Africa | South Africa | 430 | 0.03 | [-0.15, 0.21] | 2.12 |
| Sub-Saharan Africa | Tanzania | 4215 | 0.06 | [0.01, 0.11] | 6.35 |
| | **Overall** | | 0.04 | [0.01, 0.08] | |

$\tau^2 = 0.00, I^2 = 72.4\%, H^2 = 3.63$

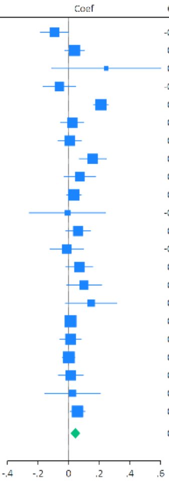

o) Remain hopeful about the future

| Region | Country | N | Coef | 95% CI | Weight (%) |
|---|---|---|---|---|---|
| East Asia & Pacific | Australia | 1934 | 0.02 | [-0.09, 0.13] | 4.25 |
| East Asia & Pacific | China | 3673 | 0.07 | [0.02, 0.13] | 5.78 |
| East Asia & Pacific | Hong Kong | 309 | 0.37 | [0.05, 0.70] | 1.17 |
| East Asia & Pacific | Indonesia | 1484 | -0.06 | [-0.18, 0.05] | 4.14 |
| East Asia & Pacific | Japan | 8266 | 0.30 | [0.25, 0.35] | 5.85 |
| East Asia & Pacific | Philippines | 2062 | -0.04 | [-0.10, 0.02] | 5.53 |
| Europe & Central Asia | Germany | 2928 | 0.06 | [-0.02, 0.13] | 5.33 |
| Europe & Central Asia | Poland | 3195 | 0.03 | [-0.04, 0.11] | 5.19 |
| Europe & Central Asia | Spain | 1546 | 0.03 | [-0.06, 0.13] | 4.76 |
| Europe & Central Asia | Sweden | 8837 | -0.00 | [-0.05, 0.05] | 5.92 |
| Europe & Central Asia | Türkiye | 251 | 0.01 | [-0.39, 0.40] | 0.85 |
| Europe & Central Asia | United Kingdom | 2446 | 0.10 | [0.01, 0.19] | 4.82 |
| Latin America & Caribbean | Argentina | 1542 | 0.03 | [-0.07, 0.12] | 4.71 |
| Latin America & Caribbean | Brazil | 2219 | 0.08 | [-0.02, 0.17] | 4.74 |
| Latin America & Caribbean | Mexico | 1101 | 0.07 | [-0.01, 0.14] | 5.19 |
| Middle East, North Africa, Afghanistan & Pakistan | Egypt | 2198 | 0.12 | [-0.05, 0.30] | 2.77 |
| North America | United States | 21460 | -0.02 | [-0.05, 0.02] | 6.13 |
| South Asia | India | 3854 | 0.15 | [0.05, 0.24] | 4.78 |
| Sub-Saharan Africa | Kenya | 6740 | 0.05 | [-0.01, 0.11] | 5.55 |
| Sub-Saharan Africa | Nigeria | 2498 | 0.07 | [-0.03, 0.16] | 4.66 |
| Sub-Saharan Africa | South Africa | 429 | -0.18 | [-0.37, 0.01] | 2.58 |
| Sub-Saharan Africa | Tanzania | 4212 | 0.07 | [0.00, 0.15] | 5.30 |
| | **Overall** | | 0.06 | [0.02, 0.10] | |

$\tau^2 = 0.00, I^2 = 80.5\%, H^2 = 5.12$

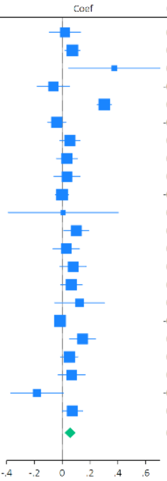

p) Feeling very capable in most things

| Region | Country | N | Coef | 95% CI | Weight (%) |
|---|---|---|---|---|---|
| East Asia & Pacific | Australia | 1994 | -0.11 | [-0.22, 0.00] | 4.18 |
| East Asia & Pacific | China | 3667 | 0.05 | [-0.01, 0.12] | 5.51 |
| East Asia & Pacific | Hong Kong | 309 | 0.07 | [-0.31, 0.44] | 0.87 |
| East Asia & Pacific | Indonesia | 1481 | 0.01 | [-0.10, 0.11] | 4.29 |
| East Asia & Pacific | Japan | 8227 | 0.27 | [0.23, 0.32] | 6.03 |
| East Asia & Pacific | Philippines | 2063 | 0.07 | [-0.00, 0.15] | 5.18 |
| Europe & Central Asia | Germany | 2924 | -0.08 | [-0.17, 0.01] | 4.84 |
| Europe & Central Asia | Poland | 3151 | 0.02 | [-0.07, 0.10] | 4.94 |
| Europe & Central Asia | Spain | 1530 | -0.00 | [-0.12, 0.12] | 3.98 |
| Europe & Central Asia | Sweden | 8791 | 0.01 | [-0.03, 0.06] | 5.98 |
| Europe & Central Asia | Türkiye | 252 | -0.08 | [-0.34, 0.18] | 1.55 |
| Europe & Central Asia | United Kingdom | 2466 | 0.05 | [-0.04, 0.14] | 4.80 |
| Latin America & Caribbean | Argentina | 1543 | -0.07 | [-0.18, 0.04] | 4.26 |
| Latin America & Caribbean | Brazil | 2208 | 0.09 | [-0.00, 0.17] | 4.86 |
| Latin America & Caribbean | Mexico | 1094 | 0.06 | [-0.06, 0.17] | 4.11 |
| Middle East, North Africa, Afghanistan & Pakistan | Egypt | 2209 | 0.07 | [-0.09, 0.23] | 3.00 |
| North America | United States | 22625 | 0.00 | [-0.03, 0.04] | 6.23 |
| South Asia | India | 3858 | 0.05 | [-0.02, 0.11] | 5.51 |
| Sub-Saharan Africa | Kenya | 6735 | 0.05 | [0.01, 0.09] | 6.15 |
| Sub-Saharan Africa | Nigeria | 2492 | 0.07 | [-0.00, 0.15] | 5.12 |
| Sub-Saharan Africa | South Africa | 430 | 0.07 | [-0.11, 0.25] | 2.61 |
| Sub-Saharan Africa | Tanzania | 4212 | 0.12 | [0.08, 0.17] | 6.04 |
| | **Overall** | | 0.04 | [0.01, 0.08] | |

$\tau^2 = 0.00, I^2 = 80.7\%, H^2 = 5.18$

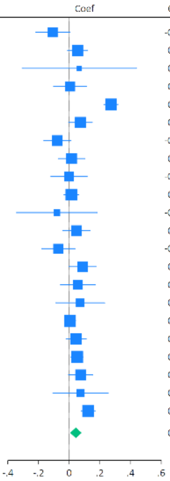

q) Expect more good things to happen to you

| Region | Country | N | Coef | 95% CI | Weight (%) |
|---|---|---|---|---|---|
| East Asia & Pacific | Australia | 1988 | 0.09 | [-0.03, 0.20] | 3.83 |
| East Asia & Pacific | China | 3673 | 0.06 | [0.00, 0.11] | 5.95 |
| East Asia & Pacific | Hong Kong | 308 | 0.35 | [-0.00, 0.71] | 0.83 |
| East Asia & Pacific | Indonesia | 1487 | -0.04 | [-0.14, 0.05] | 4.56 |
| East Asia & Pacific | Japan | 8308 | 0.24 | [0.19, 0.29] | 6.06 |
| East Asia & Pacific | Philippines | 2061 | 0.00 | [-0.07, 0.08] | 5.20 |
| Europe & Central Asia | Germany | 2929 | 0.09 | [0.02, 0.16] | 5.33 |
| Europe & Central Asia | Poland | 3191 | -0.05 | [-0.13, 0.03] | 5.07 |
| Europe & Central Asia | Spain | 1548 | 0.03 | [-0.07, 0.13] | 4.36 |
| Europe & Central Asia | Sweden | 8827 | 0.08 | [0.03, 0.13] | 6.13 |
| Europe & Central Asia | Türkiye | 253 | 0.44 | [0.06, 0.83] | 0.73 |
| Europe & Central Asia | United Kingdom | 2470 | 0.14 | [0.04, 0.23] | 4.58 |
| Latin America & Caribbean | Argentina | 1545 | 0.07 | [-0.04, 0.18] | 4.17 |
| Latin America & Caribbean | Brazil | 2221 | 0.06 | [-0.00, 0.13] | 5.56 |
| Latin America & Caribbean | Mexico | 1103 | 0.04 | [-0.04, 0.12] | 5.01 |
| Middle East, North Africa, Afghanistan & Pakistan | Egypt | 2161 | 0.20 | [0.04, 0.35] | 2.85 |
| North America | United States | 22673 | 0.02 | [-0.02, 0.06] | 6.36 |
| South Asia | India | 3854 | 0.12 | [0.02, 0.22] | 4.47 |
| Sub-Saharan Africa | Kenya | 6747 | 0.00 | [-0.06, 0.07] | 5.73 |
| Sub-Saharan Africa | Nigeria | 2494 | -0.07 | [-0.15, 0.01] | 5.04 |
| Sub-Saharan Africa | South Africa | 427 | 0.07 | [-0.08, 0.23] | 2.83 |
| Sub-Saharan Africa | Tanzania | 4211 | 0.03 | [-0.05, 0.10] | 5.33 |
| | **Overall** | | 0.06 | [0.03, 0.10] | |

$\tau^2 = 0.00, I^2 = 75.2\%, H^2 = 4.04$

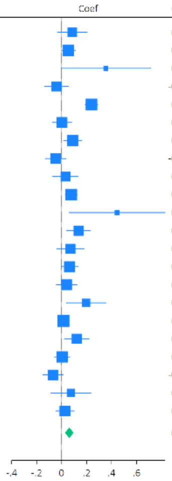

r) Overall mental health

| Region | Country | N | Coef | 95% CI | Weight (%) |
|---|---|---|---|---|---|
| East Asia & Pacific | Australia | 2009 | -0.04 | [-0.15, 0.07] | 4.50 |
| East Asia & Pacific | China | 3670 | 0.06 | [0.01, 0.12] | 5.58 |
| East Asia & Pacific | Hong Kong | 307 | 0.22 | [-0.13, 0.58] | 1.39 |
| East Asia & Pacific | Indonesia | 1486 | -0.13 | [-0.26, 0.00] | 4.14 |
| East Asia & Pacific | Japan | 8320 | 0.22 | [0.16, 0.27] | 5.58 |
| East Asia & Pacific | Philippines | 2062 | -0.02 | [-0.09, 0.06] | 5.17 |
| Europe & Central Asia | Germany | 2931 | 0.03 | [-0.05, 0.11] | 5.15 |
| Europe & Central Asia | Poland | 3196 | -0.21 | [-0.30, -0.12] | 5.01 |
| Europe & Central Asia | Spain | 1546 | 0.03 | [-0.07, 0.14] | 4.59 |
| Europe & Central Asia | Sweden | 8855 | -0.02 | [-0.06, 0.03] | 5.67 |
| Europe & Central Asia | Türkiye | 252 | 0.38 | [-0.00, 0.77] | 1.21 |
| Europe & Central Asia | United Kingdom | 2465 | 0.05 | [-0.04, 0.15] | 4.84 |
| Latin America & Caribbean | Argentina | 1546 | -0.02 | [-0.13, 0.09] | 4.53 |
| Latin America & Caribbean | Brazil | 2225 | 0.15 | [0.05, 0.26] | 4.66 |
| Latin America & Caribbean | Mexico | 1100 | 0.02 | [-0.08, 0.11] | 4.89 |
| Middle East, North Africa, Afghanistan & Pakistan | Egypt | 2199 | 0.21 | [0.10, 0.33] | 4.39 |
| North America | United States | 22777 | -0.03 | [-0.07, 0.01] | 5.77 |
| South Asia | India | 3857 | 0.17 | [0.07, 0.28] | 4.67 |
| Sub-Saharan Africa | Kenya | 6738 | 0.04 | [-0.02, 0.10] | 5.48 |
| Sub-Saharan Africa | Nigeria | 2500 | -0.02 | [-0.12, 0.09] | 4.69 |
| Sub-Saharan Africa | South Africa | 429 | -0.16 | [-0.39, 0.07] | 2.56 |
| Sub-Saharan Africa | Tanzania | 4201 | 0.12 | [0.07, 0.18] | 5.53 |
| | **Overall** | | 0.04 | [-0.01, 0.08] | |

$\tau^2 = 0.00, I^2 = 86.5\%, H^2 = 7.42$

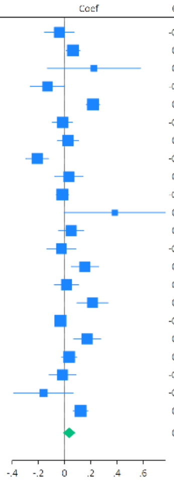

s) Overall physical health

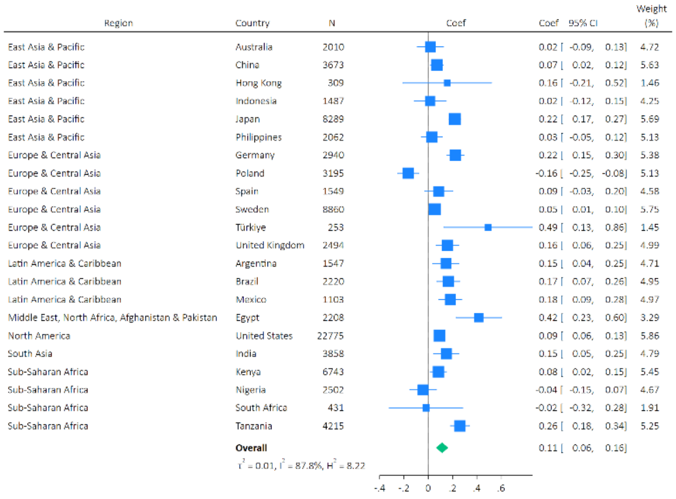

#### Supplementary Figure S4: Meta-analysis forest plots for the effects of arts engagement on later health and wellbeing outcomes (nested adjusted only for socio-demographic confounders)

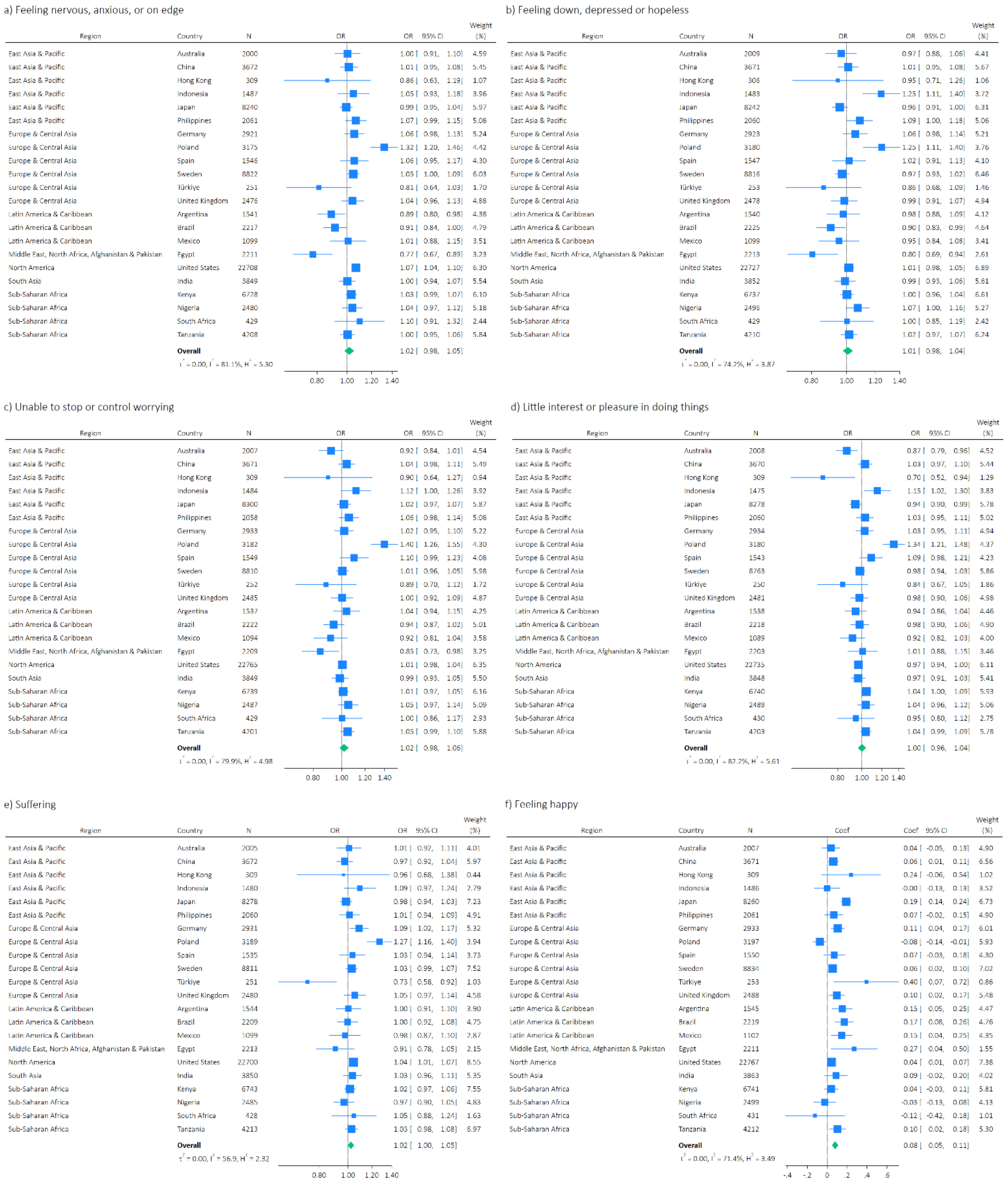

g) Understanding purpose in life

| Region | Country | N | Coef | 95% CI | Weight (%) |
|---|---|---|---|---|---|
| East Asia & Pacific | Australia | 2008 | 0.10 | [ -0.02, 0.23] | 4.05 |
| East Asia & Pacific | China | 3670 | 0.09 | [ 0.03, 0.15] | 5.78 |
| East Asia & Pacific | Hong Kong | 309 | 0.33 | [ 0.07, 0.59] | 1.80 |
| East Asia & Pacific | Indonesia | 1487 | 0.06 | [ -0.06, 0.18] | 4.19 |
| East Asia & Pacific | Japan | 8262 | 0.24 | [ 0.19, 0.29] | 5.94 |
| East Asia & Pacific | Philippines | 2060 | 0.01 | [ -0.06, 0.09] | 5.32 |
| Europe & Central Asia | Germany | 2931 | 0.07 | [ -0.01, 0.15] | 5.27 |
| Europe & Central Asia | Poland | 3187 | -0.10 | [ -0.18, -0.02] | 5.26 |
| Europe & Central Asia | Spain | 1543 | 0.12 | [ 0.02, 0.23] | 4.48 |
| Europe & Central Asia | Sweden | 8766 | 0.06 | [ 0.01, 0.12] | 5.90 |
| Europe & Central Asia | Türkiye | 252 | 0.35 | [ 0.04, 0.66] | 1.38 |
| Europe & Central Asia | United Kingdom | 2478 | 0.04 | [ -0.06, 0.14] | 4.72 |
| Latin America & Caribbean | Argentina | 1540 | 0.17 | [ 0.06, 0.29] | 4.33 |
| Latin America & Caribbean | Brazil | 2216 | 0.17 | [ 0.07, 0.27] | 4.70 |
| Latin America & Caribbean | Mexico | 1099 | 0.17 | [ 0.08, 0.26] | 4.94 |
| Middle East, North Africa, Afghanistan & Pakistan | Egypt | 2193 | 0.20 | [ 0.05, 0.36] | 3.38 |
| North America | United States | 22735 | 0.11 | [ 0.07, 0.15] | 6.18 |
| South Asia | India | 3849 | 0.10 | [ -0.00, 0.19] | 4.74 |
| Sub-Saharan Africa | Kenya | 6740 | -0.00 | [ -0.07, 0.07] | 5.52 |
| Sub-Saharan Africa | Nigeria | 2494 | -0.05 | [ -0.15, 0.05] | 4.66 |
| Sub-Saharan Africa | South Africa | 431 | -0.25 | [ -0.48, -0.01] | 2.11 |
| Sub-Saharan Africa | Tanzania | 4176 | 0.13 | [ 0.05, 0.20] | 5.32 |
| | **Overall** | | 0.09 | [ 0.05, 0.13] | |

$\tau^2 = 0.00$, $I^2 = 79.1\%$, $H^2 = 4.79$

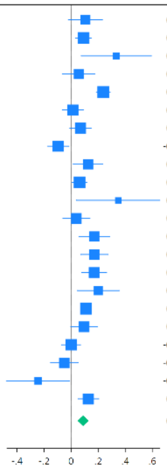

h) Life satisfaction

| Region | Country | N | Coef | 95% CI | Weight (%) |
|---|---|---|---|---|---|
| East Asia & Pacific | Australia | 2010 | 0.07 | [ -0.03, 0.17] | 4.74 |
| East Asia & Pacific | China | 3652 | 0.06 | [ 0.00, 0.11] | 6.46 |
| East Asia & Pacific | Hong Kong | 297 | 0.31 | [ -0.02, 0.63] | 0.96 |
| East Asia & Pacific | Indonesia | 1484 | -0.01 | [ -0.13, 0.12] | 3.79 |
| East Asia & Pacific | Japan | 8226 | 0.19 | [ 0.14, 0.24] | 6.61 |
| East Asia & Pacific | Philippines | 2059 | 0.02 | [ -0.07, 0.11] | 5.02 |
| Europe & Central Asia | Germany | 2913 | 0.09 | [ 0.03, 0.16] | 5.96 |
| Europe & Central Asia | Poland | 3180 | -0.11 | [ -0.18, -0.04] | 5.78 |
| Europe & Central Asia | Spain | 1542 | 0.09 | [ -0.02, 0.20] | 4.37 |
| Europe & Central Asia | Sweden | 8822 | 0.05 | [ 0.00, 0.09] | 6.88 |
| Europe & Central Asia | Türkiye | 253 | 0.26 | [ -0.05, 0.57] | 1.05 |
| Europe & Central Asia | United Kingdom | 2459 | 0.11 | [ 0.03, 0.19] | 5.44 |
| Latin America & Caribbean | Argentina | 1544 | 0.17 | [ 0.06, 0.27] | 4.45 |
| Latin America & Caribbean | Brazil | 2219 | 0.12 | [ 0.02, 0.22] | 4.49 |
| Latin America & Caribbean | Mexico | 1102 | 0.11 | [ -0.01, 0.22] | 4.08 |
| Middle East, North Africa, Afghanistan & Pakistan | Egypt | 2199 | 0.17 | [ 0.01, 0.32] | 2.99 |
| North America | United States | 22756 | 0.03 | [ -0.00, 0.06] | 7.24 |
| South Asia | India | 3859 | 0.12 | [ 0.01, 0.23] | 4.19 |
| Sub-Saharan Africa | Kenya | 6744 | 0.02 | [ -0.05, 0.10] | 5.58 |
| Sub-Saharan Africa | Nigeria | 2499 | -0.02 | [ -0.14, 0.11] | 3.93 |
| Sub-Saharan Africa | South Africa | 426 | -0.13 | [ -0.43, 0.17] | 1.10 |
| Sub-Saharan Africa | Tanzania | 4211 | -0.02 | [ -0.11, 0.08] | 4.90 |
| | **Overall** | | 0.07 | [ 0.03, 0.10] | |

$\tau^2 = 0.00$, $I^2 = 71.7\%$, $H^2 = 3.53$

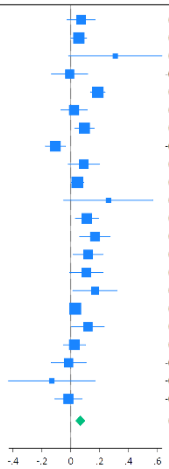

i) Life evaluation: Today

| Region | Country | N | Coef | 95% CI | Weight (%) |
|---|---|---|---|---|---|
| East Asia & Pacific | Australia | 2011 | 0.02 | [ -0.07, 0.11] | 4.81 |
| East Asia & Pacific | China | 3671 | 0.03 | [ -0.02, 0.09] | 6.64 |
| East Asia & Pacific | Hong Kong | 309 | 0.16 | [ -0.06, 0.39] | 1.31 |
| East Asia & Pacific | Indonesia | 1485 | 0.07 | [ -0.07, 0.21] | 2.81 |
| East Asia & Pacific | Japan | 8293 | 0.20 | [ 0.15, 0.24] | 7.32 |
| East Asia & Pacific | Philippines | 2056 | 0.13 | [ 0.04, 0.22] | 4.79 |
| Europe & Central Asia | Germany | 2928 | 0.12 | [ 0.07, 0.18] | 6.56 |
| Europe & Central Asia | Poland | 3195 | 0.05 | [ -0.02, 0.12] | 5.94 |
| Europe & Central Asia | Spain | 1550 | 0.07 | [ -0.03, 0.17] | 4.32 |
| Europe & Central Asia | Sweden | 8831 | 0.05 | [ 0.01, 0.08] | 7.89 |
| Europe & Central Asia | Türkiye | 252 | 0.26 | [ -0.06, 0.58] | 0.71 |
| Europe & Central Asia | United Kingdom | 2485 | 0.13 | [ 0.05, 0.20] | 5.46 |
| Latin America & Caribbean | Argentina | 1547 | 0.14 | [ 0.03, 0.25] | 3.69 |
| Latin America & Caribbean | Brazil | 2226 | 0.12 | [ 0.03, 0.21] | 4.66 |
| Latin America & Caribbean | Mexico | 1102 | 0.06 | [ -0.04, 0.17] | 3.89 |
| Middle East, North Africa, Afghanistan & Pakistan | Egypt | 2201 | 0.29 | [ 0.09, 0.48] | 1.64 |
| North America | United States | 22777 | 0.04 | [ 0.02, 0.07] | 8.41 |
| South Asia | India | 3842 | 0.11 | [ 0.00, 0.23] | 3.74 |
| Sub-Saharan Africa | Kenya | 6744 | 0.04 | [ -0.03, 0.11] | 5.88 |
| Sub-Saharan Africa | Nigeria | 2503 | -0.04 | [ -0.16, 0.07] | 3.53 |
| Sub-Saharan Africa | South Africa | 429 | -0.16 | [ -0.45, 0.13] | 0.85 |
| Sub-Saharan Africa | Tanzania | 4215 | 0.14 | [ 0.06, 0.22] | 5.17 |
| | **Overall** | | 0.09 | [ 0.06, 0.12] | |

$\tau^2 = 0.00$, $I^2 = 63.1\%$, $H^2 = 2.71$

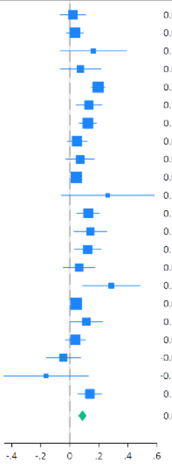

j) Life evaluation: Five years from now

| Region | Country | N | Coef | 95% CI | Weight (%) |
|---|---|---|---|---|---|
| East Asia & Pacific | Australia | 2010 | 0.09 | [ -0.01, 0.18] | 4.48 |
| East Asia & Pacific | China | 3653 | 0.04 | [ -0.01, 0.09] | 6.29 |
| East Asia & Pacific | Hong Kong | 300 | 0.32 | [ 0.06, 0.58] | 1.29 |
| East Asia & Pacific | Indonesia | 1467 | 0.10 | [ -0.00, 0.20] | 4.28 |
| East Asia & Pacific | Japan | 8297 | 0.22 | [ 0.17, 0.27] | 6.39 |
| East Asia & Pacific | Philippines | 2057 | 0.04 | [ -0.04, 0.12] | 5.10 |
| Europe & Central Asia | Germany | 2912 | 0.15 | [ 0.08, 0.21] | 5.78 |
| Europe & Central Asia | Poland | 3153 | 0.06 | [ -0.02, 0.15] | 5.00 |
| Europe & Central Asia | Spain | 1540 | 0.09 | [ -0.02, 0.20] | 4.03 |
| Europe & Central Asia | Sweden | 8806 | 0.05 | [ 0.01, 0.09] | 6.78 |
| Europe & Central Asia | Türkiye | 236 | 0.13 | [ -0.20, 0.47] | 0.85 |
| Europe & Central Asia | United Kingdom | 2460 | 0.14 | [ 0.06, 0.22] | 5.13 |
| Latin America & Caribbean | Argentina | 1510 | 0.04 | [ -0.06, 0.15] | 4.16 |
| Latin America & Caribbean | Brazil | 2216 | 0.09 | [ 0.01, 0.16] | 5.28 |
| Latin America & Caribbean | Mexico | 1078 | 0.08 | [ -0.03, 0.18] | 4.23 |
| Middle East, North Africa, Afghanistan & Pakistan | Egypt | 1632 | 0.29 | [ 0.09, 0.48] | 2.08 |
| North America | United States | 22765 | -0.01 | [ -0.04, 0.03] | 6.96 |
| South Asia | India | 3576 | 0.10 | [ 0.00, 0.20] | 4.49 |
| Sub-Saharan Africa | Kenya | 6699 | 0.04 | [ -0.02, 0.11] | 5.80 |
| Sub-Saharan Africa | Nigeria | 2470 | -0.06 | [ -0.15, 0.03] | 4.75 |
| Sub-Saharan Africa | South Africa | 423 | -0.19 | [ -0.42, 0.05] | 1.52 |
| Sub-Saharan Africa | Tanzania | 3979 | 0.13 | [ 0.05, 0.20] | 5.31 |
| | **Overall** | | 0.08 | [ 0.05, 0.11] | |

$\tau^2 = 0.00$, $I^2 = 72.8\%$, $H^2 = 3.68$

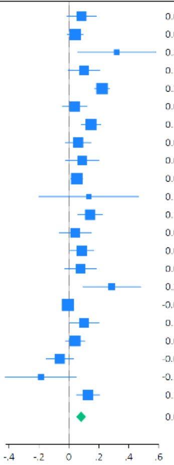

k) Feeling lonely

| Region | Country | N | Coef | 95% CI | Weight (%) |
|---|---|---|---|---|---|
| East Asia & Pacific | Australia | 2010 | -0.03 | [ -0.15, 0.10] | 4.36 |
| East Asia & Pacific | China | 3672 | -0.04 | [ -0.11, 0.03] | 7.26 |
| East Asia & Pacific | Hong Kong | 308 | -0.31 | [ -0.68, 0.06] | 0.83 |
| East Asia & Pacific | Indonesia | 1487 | 0.00 | [ -0.19, 0.19] | 2.55 |
| East Asia & Pacific | Japan | 8309 | -0.14 | [ -0.20, -0.07] | 7.81 |
| East Asia & Pacific | Philippines | 2063 | -0.05 | [ -0.17, 0.07] | 4.66 |
| Europe & Central Asia | Germany | 2938 | -0.06 | [ -0.15, 0.03] | 6.17 |
| Europe & Central Asia | Poland | 3190 | 0.12 | [ 0.02, 0.21] | 5.93 |
| Europe & Central Asia | Spain | 1553 | -0.00 | [ -0.16, 0.16] | 3.30 |
| Europe & Central Asia | Sweden | 8848 | -0.05 | [ -0.11, -0.00] | 8.39 |
| Europe & Central Asia | Türkiye | 250 | -0.15 | [ -0.57, 0.26] | 0.67 |
| Europe & Central Asia | United Kingdom | 2476 | -0.10 | [ -0.21, 0.00] | 5.26 |
| Latin America & Caribbean | Argentina | 1549 | -0.20 | [ -0.35, -0.04] | 3.47 |
| Latin America & Caribbean | Brazil | 2221 | -0.09 | [ -0.24, 0.05] | 3.81 |
| Latin America & Caribbean | Mexico | 1099 | -0.18 | [ -0.36, 0.00] | 2.79 |
| Middle East, North Africa, Afghanistan & Pakistan | Egypt | 2210 | -0.10 | [ -0.41, 0.22] | 1.10 |
| North America | United States | 22786 | -0.08 | [ -0.12, -0.03] | 9.02 |
| South Asia | India | 3863 | 0.11 | [ -0.02, 0.24] | 4.30 |
| Sub-Saharan Africa | Kenya | 6739 | -0.04 | [ -0.12, 0.04] | 6.71 |
| Sub-Saharan Africa | Nigeria | 2496 | 0.08 | [ -0.05, 0.22] | 4.11 |
| Sub-Saharan Africa | South Africa | 431 | 0.19 | [ -0.09, 0.47] | 1.33 |
| Sub-Saharan Africa | Tanzania | 4213 | -0.06 | [ -0.15, 0.03] | 6.17 |
| | **Overall** | | -0.05 | [ -0.08, -0.01] | |

$\tau^2 = 0.00$, $I^2 = 55.1\%$, $H^2 = 2.23$

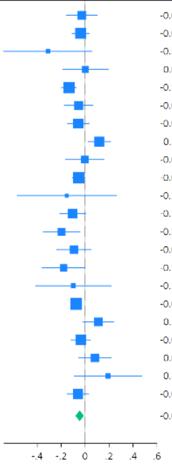

l) Life is worthwhile

| Region | Country | N | Coef | 95% CI | Weight (%) |
|---|---|---|---|---|---|
| East Asia & Pacific | Australia | 2008 | 0.08 | [ -0.03, 0.20] | 4.33 |
| East Asia & Pacific | China | 3669 | 0.04 | [ -0.01, 0.10] | 6.19 |
| East Asia & Pacific | Hong Kong | 306 | 0.37 | [ 0.11, 0.64] | 1.62 |
| East Asia & Pacific | Indonesia | 1486 | 0.06 | [ -0.04, 0.17] | 4.64 |
| East Asia & Pacific | Japan | 8290 | 0.24 | [ 0.19, 0.30] | 6.18 |
| East Asia & Pacific | Philippines | 2060 | 0.00 | [ -0.08, 0.09] | 5.05 |
| Europe & Central Asia | Germany | 2932 | 0.11 | [ 0.05, 0.18] | 5.71 |
| Europe & Central Asia | Poland | 3189 | -0.13 | [ -0.21, -0.06] | 5.46 |
| Europe & Central Asia | Spain | 1541 | 0.03 | [ -0.08, 0.15] | 4.18 |
| Europe & Central Asia | Sweden | 8832 | 0.09 | [ 0.04, 0.13] | 6.34 |
| Europe & Central Asia | Türkiye | 253 | 0.30 | [ -0.02, 0.61] | 1.18 |
| Europe & Central Asia | United Kingdom | 2473 | 0.12 | [ 0.04, 0.20] | 5.32 |
| Latin America & Caribbean | Argentina | 1541 | 0.17 | [ 0.07, 0.28] | 4.58 |
| Latin America & Caribbean | Brazil | 2224 | 0.16 | [ 0.07, 0.25] | 4.95 |
| Latin America & Caribbean | Mexico | 1101 | 0.11 | [ -0.00, 0.23] | 4.24 |
| Middle East, North Africa, Afghanistan & Pakistan | Egypt | 2207 | 0.12 | [ -0.12, 0.36] | 1.79 |
| North America | United States | 22770 | 0.08 | [ 0.05, 0.12] | 6.60 |
| South Asia | India | 3844 | 0.03 | [ -0.07, 0.13] | 4.64 |
| Sub-Saharan Africa | Kenya | 6743 | 0.04 | [ -0.03, 0.12] | 5.60 |
| Sub-Saharan Africa | Nigeria | 2498 | 0.04 | [ -0.07, 0.15] | 4.39 |
| Sub-Saharan Africa | South Africa | 428 | 0.06 | [ -0.18, 0.31] | 1.77 |
| Sub-Saharan Africa | Tanzania | 4209 | 0.13 | [ 0.05, 0.22] | 5.25 |
| | **Overall** | | 0.09 | [ 0.05, 0.13] | |

$\tau^2 = 0.00$, $I^2 = 77.3\%$, $H^2 = 4.41$

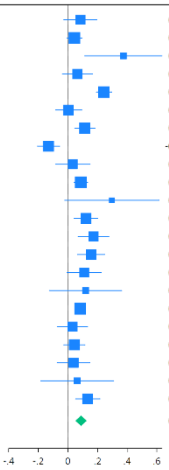

m) Feeling grateful

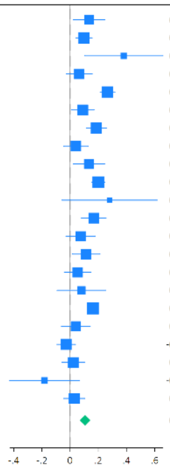

| Region | Country | N | Coef | 95% CI | Weight (%) |
|---|---|---|---|---|---|
| East Asia & Pacific | Australia | 1980 | 0.13 | [ 0.02, 0.25] | 4.20 |
| East Asia & Pacific | China | 3670 | 0.10 | [ 0.04, 0.16] | 5.92 |
| East Asia & Pacific | Hong Kong | 308 | 0.38 | [ 0.10, 0.66] | 1.41 |
| East Asia & Pacific | Indonesia | 1482 | 0.06 | [ -0.03, 0.16] | 4.83 |
| East Asia & Pacific | Japan | 8240 | 0.27 | [ 0.21, 0.32] | 6.04 |
| East Asia & Pacific | Philippines | 2061 | 0.09 | [ 0.01, 0.17] | 5.21 |
| Europe & Central Asia | Germany | 2921 | 0.19 | [ 0.11, 0.26] | 5.48 |
| Europe & Central Asia | Poland | 3189 | 0.04 | [ -0.05, 0.13] | 4.98 |
| Europe & Central Asia | Spain | 1547 | 0.13 | [ 0.02, 0.25] | 4.23 |
| Europe & Central Asia | Sweden | 8836 | 0.20 | [ 0.15, 0.25] | 6.24 |
| Europe & Central Asia | Türkiye | 246 | 0.28 | [ -0.06, 0.62] | 1.02 |
| Europe & Central Asia | United Kingdom | 2445 | 0.17 | [ 0.08, 0.26] | 4.98 |
| Latin America & Caribbean | Argentina | 1543 | 0.07 | [ -0.03, 0.18] | 4.46 |
| Latin America & Caribbean | Brazil | 2215 | 0.11 | [ 0.02, 0.21] | 4.64 |
| Latin America & Caribbean | Mexico | 1101 | 0.05 | [ -0.04, 0.15] | 4.74 |
| Middle East, North Africa, Afghanistan & Pakistan | Egypt | 2181 | 0.08 | [ -0.09, 0.25] | 2.74 |
| North America | United States | 21475 | 0.16 | [ 0.13, 0.20] | 6.47 |
| South Asia | India | 3837 | 0.04 | [ -0.06, 0.14] | 4.53 |
| Sub-Saharan Africa | Kenya | 6744 | -0.03 | [ -0.09, 0.04] | 5.63 |
| Sub-Saharan Africa | Nigeria | 2490 | 0.02 | [ -0.06, 0.11] | 5.14 |
| Sub-Saharan Africa | South Africa | 428 | -0.18 | [ -0.43, 0.07] | 1.69 |
| Sub-Saharan Africa | Tanzania | 4197 | 0.03 | [ -0.04, 0.10] | 5.40 |
| | **Overall** | | 0.11 | [ 0.07, 0.14] | |

$\tau^2 = 0.00$, $I^2 = 76.9\%$, $H^2 = 4.33$

n) Feeling at peace

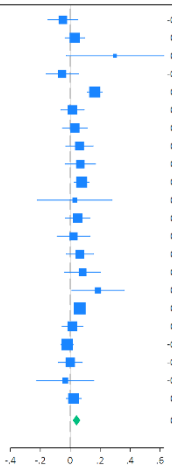

| Region | Country | N | Coef | 95% CI | Weight (%) |
|---|---|---|---|---|---|
| East Asia & Pacific | Australia | 2007 | -0.05 | [ -0.15, 0.05] | 3.89 |
| East Asia & Pacific | China | 3669 | 0.03 | [ -0.03, 0.10] | 5.95 |
| East Asia & Pacific | Hong Kong | 309 | 0.30 | [ -0.03, 0.63] | 0.56 |
| East Asia & Pacific | Indonesia | 1485 | -0.05 | [ -0.16, 0.05] | 3.52 |
| East Asia & Pacific | Japan | 8263 | 0.16 | [ 0.11, 0.21] | 7.08 |
| East Asia & Pacific | Philippines | 2061 | 0.02 | [ -0.06, 0.09] | 5.11 |
| Europe & Central Asia | Germany | 2923 | 0.03 | [ -0.05, 0.11] | 4.81 |
| Europe & Central Asia | Poland | 3166 | 0.06 | [ -0.03, 0.15] | 4.29 |
| Europe & Central Asia | Spain | 1541 | 0.07 | [ -0.03, 0.17] | 3.82 |
| Europe & Central Asia | Sweden | 8824 | 0.08 | [ 0.03, 0.13] | 7.13 |
| Europe & Central Asia | Türkiye | 253 | 0.03 | [ -0.22, 0.28] | 0.93 |
| Europe & Central Asia | United Kingdom | 2479 | 0.05 | [ -0.03, 0.13] | 4.86 |
| Latin America & Caribbean | Argentina | 1538 | 0.02 | [ -0.09, 0.13] | 3.45 |
| Latin America & Caribbean | Brazil | 2219 | 0.06 | [ -0.03, 0.16] | 4.33 |
| Latin America & Caribbean | Mexico | 1096 | 0.08 | [ -0.04, 0.20] | 3.10 |
| Middle East, North Africa, Afghanistan & Pakistan | Egypt | 2210 | 0.19 | [ 0.01, 0.36] | 1.72 |
| North America | United States | 22702 | 0.06 | [ 0.03, 0.10] | 8.37 |
| South Asia | India | 3845 | 0.01 | [ -0.05, 0.08] | 5.72 |
| Sub-Saharan Africa | Kenya | 6743 | -0.02 | [ -0.06, 0.02] | 7.80 |
| Sub-Saharan Africa | Nigeria | 2493 | -0.00 | [ -0.08, 0.08] | 4.99 |
| Sub-Saharan Africa | South Africa | 430 | -0.03 | [ -0.22, 0.16] | 1.49 |
| Sub-Saharan Africa | Tanzania | 4215 | 0.02 | [ -0.03, 0.07] | 7.05 |
| | **Overall** | | 0.04 | [ 0.02, 0.07] | |

$\tau^2 = 0.00$, $I^2 = 55.1\%$, $H^2 = 2.23$

o) Remain hopeful about the future

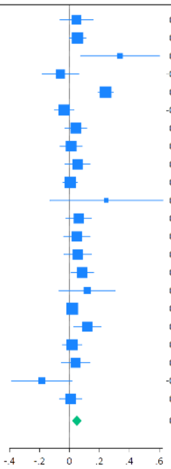

| Region | Country | N | Coef | 95% CI | Weight (%) |
|---|---|---|---|---|---|
| East Asia & Pacific | Australia | 1934 | 0.05 | [ -0.06, 0.16] | 3.97 |
| East Asia & Pacific | China | 3673 | 0.05 | [ -0.00, 0.11] | 6.07 |
| East Asia & Pacific | Hong Kong | 309 | 0.34 | [ 0.07, 0.60] | 1.25 |
| East Asia & Pacific | Indonesia | 1484 | -0.06 | [ -0.18, 0.06] | 3.61 |
| East Asia & Pacific | Japan | 8266 | 0.24 | [ 0.19, 0.29] | 6.21 |
| East Asia & Pacific | Philippines | 2062 | -0.04 | [ -0.10, 0.03] | 5.71 |
| Europe & Central Asia | Germany | 2928 | 0.04 | [ -0.03, 0.11] | 5.39 |
| Europe & Central Asia | Poland | 3195 | 0.01 | [ -0.06, 0.08] | 5.34 |
| Europe & Central Asia | Spain | 1546 | 0.05 | [ -0.03, 0.14] | 4.94 |
| Europe & Central Asia | Sweden | 8837 | 0.00 | [ -0.04, 0.05] | 6.30 |
| Europe & Central Asia | Türkiye | 251 | 0.25 | [ -0.13, 0.62] | 0.67 |
| Europe & Central Asia | United Kingdom | 2446 | 0.06 | [ -0.02, 0.15] | 4.88 |
| Latin America & Caribbean | Argentina | 1542 | 0.05 | [ -0.04, 0.14] | 4.77 |
| Latin America & Caribbean | Brazil | 2219 | 0.06 | [ -0.04, 0.15] | 4.66 |
| Latin America & Caribbean | Mexico | 1101 | 0.09 | [ 0.01, 0.16] | 5.26 |
| Middle East, North Africa, Afghanistan & Pakistan | Egypt | 2198 | 0.12 | [ -0.07, 0.30] | 2.13 |
| North America | United States | 21460 | 0.02 | [ -0.02, 0.05] | 6.74 |
| South Asia | India | 3854 | 0.12 | [ 0.03, 0.21] | 4.64 |
| Sub-Saharan Africa | Kenya | 6740 | 0.02 | [ -0.05, 0.08] | 5.72 |
| Sub-Saharan Africa | Nigeria | 2498 | 0.04 | [ -0.05, 0.14] | 4.51 |
| Sub-Saharan Africa | South Africa | 429 | -0.18 | [ -0.39, 0.02] | 1.92 |
| Sub-Saharan Africa | Tanzania | 4212 | 0.01 | [ -0.07, 0.08] | 5.31 |
| | **Overall** | | 0.05 | [ 0.02, 0.08] | |

$\tau^2 = 0.00$, $I^2 = 72.0\%$, $H^2 = 3.57$

p) Feeling very capable in most things

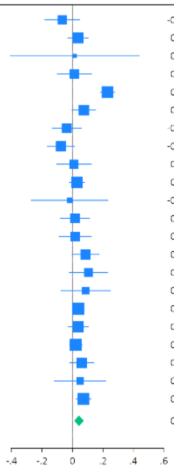

| Region | Country | N | Coef | 95% CI | Weight (%) |
|---|---|---|---|---|---|
| East Asia & Pacific | Australia | 1994 | -0.07 | [ -0.18, 0.05] | 3.84 |
| East Asia & Pacific | China | 3667 | 0.04 | [ -0.03, 0.10] | 5.74 |
| East Asia & Pacific | Hong Kong | 309 | 0.02 | [ -0.41, 0.44] | 0.52 |
| East Asia & Pacific | Indonesia | 1481 | 0.01 | [ -0.10, 0.12] | 3.89 |
| East Asia & Pacific | Japan | 8227 | 0.23 | [ 0.18, 0.28] | 6.59 |
| East Asia & Pacific | Philippines | 2063 | 0.07 | [ -0.00, 0.15] | 5.25 |
| Europe & Central Asia | Germany | 2924 | -0.04 | [ -0.13, 0.06] | 4.49 |
| Europe & Central Asia | Poland | 3151 | -0.08 | [ -0.16, 0.01] | 4.78 |
| Europe & Central Asia | Spain | 1530 | 0.01 | [ -0.10, 0.12] | 3.82 |
| Europe & Central Asia | Sweden | 8791 | 0.03 | [ -0.02, 0.08] | 6.52 |
| Europe & Central Asia | Türkiye | 252 | -0.02 | [ -0.27, 0.23] | 1.30 |
| Europe & Central Asia | United Kingdom | 2466 | 0.02 | [ -0.08, 0.11] | 4.49 |
| Latin America & Caribbean | Argentina | 1543 | 0.02 | [ -0.09, 0.12] | 4.12 |
| Latin America & Caribbean | Brazil | 2208 | 0.09 | [ -0.00, 0.17] | 4.81 |
| Latin America & Caribbean | Mexico | 1094 | 0.10 | [ -0.02, 0.23] | 3.48 |
| Middle East, North Africa, Afghanistan & Pakistan | Egypt | 2209 | 0.09 | [ -0.08, 0.25] | 2.54 |
| North America | United States | 22625 | 0.04 | [ 0.00, 0.07] | 7.00 |
| South Asia | India | 3858 | 0.04 | [ -0.03, 0.10] | 5.78 |
| Sub-Saharan Africa | Kenya | 6735 | 0.02 | [ -0.02, 0.06] | 6.86 |
| Sub-Saharan Africa | Nigeria | 2497 | 0.06 | [ -0.02, 0.14] | 5.20 |
| Sub-Saharan Africa | South Africa | 430 | 0.05 | [ -0.12, 0.22] | 2.38 |
| Sub-Saharan Africa | Tanzania | 4212 | 0.07 | [ 0.02, 0.12] | 6.57 |
| | **Overall** | | 0.04 | [ 0.01, 0.07] | |

$\tau^2 = 0.00$, $I^2 = 70.4\%$, $H^2 = 3.38$

q) Expect more good things to happen to you

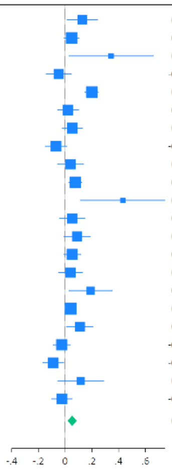

| Region | Country | N | Coef | 95% CI | Weight (%) |
|---|---|---|---|---|---|
| East Asia & Pacific | Australia | 1988 | 0.13 | [ 0.01, 0.24] | 3.92 |
| East Asia & Pacific | China | 3673 | 0.05 | [ -0.01, 0.10] | 5.91 |
| East Asia & Pacific | Hong Kong | 308 | 0.34 | [ 0.03, 0.66] | 1.01 |
| East Asia & Pacific | Indonesia | 1487 | -0.05 | [ -0.14, 0.05] | 4.56 |
| East Asia & Pacific | Japan | 8308 | 0.20 | [ 0.15, 0.25] | 6.05 |
| East Asia & Pacific | Philippines | 2061 | 0.02 | [ -0.06, 0.10] | 5.11 |
| Europe & Central Asia | Germany | 2929 | 0.05 | [ -0.02, 0.13] | 5.25 |
| Europe & Central Asia | Poland | 3191 | -0.07 | [ -0.15, 0.01] | 5.06 |
| Europe & Central Asia | Spain | 1548 | 0.04 | [ -0.06, 0.14] | 4.47 |
| Europe & Central Asia | Sweden | 8827 | 0.08 | [ 0.03, 0.12] | 6.12 |
| Europe & Central Asia | Türkiye | 253 | 0.43 | [ 0.12, 0.75] | 1.01 |
| Europe & Central Asia | United Kingdom | 2470 | 0.05 | [ -0.04, 0.15] | 4.58 |
| Latin America & Caribbean | Argentina | 1545 | 0.09 | [ -0.01, 0.19] | 4.44 |
| Latin America & Caribbean | Brazil | 2221 | 0.05 | [ -0.01, 0.12] | 5.64 |
| Latin America & Caribbean | Mexico | 1103 | 0.04 | [ -0.05, 0.13] | 4.76 |
| Middle East, North Africa, Afghanistan & Pakistan | Egypt | 2161 | 0.19 | [ 0.03, 0.35] | 2.73 |
| North America | United States | 22673 | 0.04 | [ 0.00, 0.08] | 6.42 |
| South Asia | India | 3854 | 0.11 | [ 0.01, 0.21] | 4.43 |
| Sub-Saharan Africa | Kenya | 6747 | -0.03 | [ -0.09, 0.04] | 5.66 |
| Sub-Saharan Africa | Nigeria | 2494 | -0.09 | [ -0.17, -0.01] | 5.13 |
| Sub-Saharan Africa | South Africa | 427 | 0.12 | [ -0.05, 0.29] | 2.55 |
| Sub-Saharan Africa | Tanzania | 4211 | -0.02 | [ -0.10, 0.05] | 5.21 |
| | **Overall** | | 0.05 | [ 0.02, 0.09] | |

$\tau^2 = 0.00$, $I^2 = 75.0\%$, $H^2 = 4.00$

r) Overall mental health

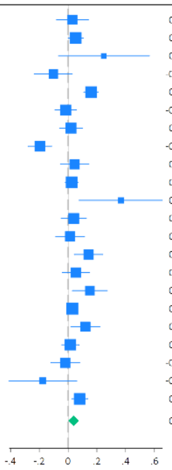

| Region | Country | N | Coef | 95% CI | Weight (%) |
|---|---|---|---|---|---|
| East Asia & Pacific | Australia | 2009 | 0.03 | [ -0.08, 0.14] | 4.23 |
| East Asia & Pacific | China | 3670 | 0.05 | [ 0.00, 0.11] | 5.93 |
| East Asia & Pacific | Hong Kong | 307 | 0.25 | [ -0.07, 0.56] | 1.15 |
| East Asia & Pacific | Indonesia | 1486 | -0.10 | [ -0.23, 0.03] | 3.67 |
| East Asia & Pacific | Japan | 8320 | 0.16 | [ 0.11, 0.21] | 5.99 |
| East Asia & Pacific | Philippines | 2062 | -0.02 | [ -0.09, 0.06] | 5.28 |
| Europe & Central Asia | Germany | 2931 | 0.02 | [ -0.06, 0.10] | 5.15 |
| Europe & Central Asia | Poland | 3196 | -0.20 | [ -0.28, -0.11] | 5.03 |
| Europe & Central Asia | Spain | 1546 | 0.05 | [ -0.05, 0.14] | 4.57 |
| Europe & Central Asia | Sweden | 8855 | 0.02 | [ -0.02, 0.07] | 6.16 |
| Europe & Central Asia | Türkiye | 252 | 0.37 | [ 0.08, 0.66] | 1.33 |
| Europe & Central Asia | United Kingdom | 2465 | 0.04 | [ -0.05, 0.13] | 4.91 |
| Latin America & Caribbean | Argentina | 1546 | 0.01 | [ -0.09, 0.11] | 4.53 |
| Latin America & Caribbean | Brazil | 2225 | 0.14 | [ 0.04, 0.24] | 4.56 |
| Latin America & Caribbean | Mexico | 1100 | 0.05 | [ -0.04, 0.15] | 4.70 |
| Middle East, North Africa, Afghanistan & Pakistan | Egypt | 2199 | 0.15 | [ 0.03, 0.27] | 3.92 |
| North America | United States | 22777 | 0.03 | [ -0.00, 0.06] | 6.39 |
| South Asia | India | 3857 | 0.12 | [ 0.02, 0.22] | 4.53 |
| Sub-Saharan Africa | Kenya | 6738 | 0.02 | [ -0.04, 0.08] | 5.74 |
| Sub-Saharan Africa | Nigeria | 2500 | -0.02 | [ -0.12, 0.08] | 4.54 |
| Sub-Saharan Africa | South Africa | 429 | -0.17 | [ -0.41, 0.06] | 1.83 |
| Sub-Saharan Africa | Tanzania | 4201 | 0.08 | [ 0.02, 0.14] | 5.85 |
| | **Overall** | | 0.04 | [ 0.00, 0.08] | |

$\tau^2 = 0.00$, $I^2 = 78.9\%$, $H^2 = 4.73$

s) Overall physical health

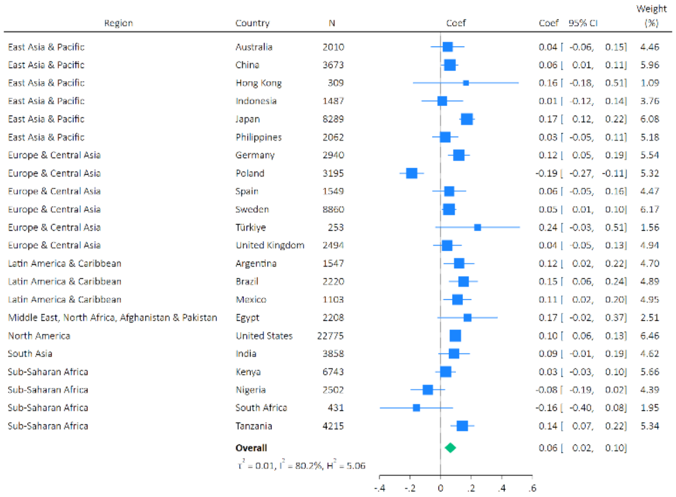

### Supplementary Figure S5: Meta-analysis forest plots for the effects of arts engagement on later health and wellbeing outcomes (nested adjusted only for socio-demographic confounders and baseline health outcome)

a) Feeling nervous, anxious, or on edge

| Region | Country | N | OR | 95% CI | | Weight (%) |
|---|---|---|---|---|---|---|
| East Asia & Pacific | Australia | 2000 | 1.00 | 0.90 | 1.10 | 4.40 |
| East Asia & Pacific | China | 3672 | 1.01 | 0.94 | 1.07 | 5.69 |
| East Asia & Pacific | Hong Kong | 309 | 1.02 | 0.72 | 1.45 | 0.74 |
| East Asia & Pacific | Indonesia | 1487 | 1.05 | 0.94 | 1.18 | 3.71 |
| East Asia & Pacific | Japan | 8240 | 1.02 | 0.97 | 1.07 | 6.32 |
| East Asia & Pacific | Philippines | 2061 | 1.07 | 0.99 | 1.15 | 5.11 |
| Europe & Central Asia | Germany | 2921 | 1.02 | 0.95 | 1.10 | 5.41 |
| Europe & Central Asia | Poland | 3175 | 1.27 | 1.15 | 1.41 | 4.24 |
| Europe & Central Asia | Spain | 1546 | 1.02 | 0.92 | 1.14 | 3.92 |
| Europe & Central Asia | Sweden | 8822 | 1.02 | 0.98 | 1.07 | 6.47 |
| Europe & Central Asia | Türkiye | 251 | 0.87 | 0.68 | 1.12 | 1.31 |
| Europe & Central Asia | United Kingdom | 2476 | 0.99 | 0.91 | 1.09 | 4.65 |
| Latin America & Caribbean | Argentina | 1541 | 0.89 | 0.81 | 0.98 | 4.31 |
| Latin America & Caribbean | Brazil | 2217 | 0.94 | 0.86 | 1.03 | 4.63 |
| Latin America & Caribbean | Mexico | 1099 | 1.01 | 0.88 | 1.16 | 3.06 |
| Middle East, North Africa, Afghanistan & Pakistan | Egypt | 2211 | 0.78 | 0.68 | 0.90 | 3.15 |
| North America | United States | 22708 | 1.09 | 1.06 | 1.13 | 6.93 |
| South Asia | India | 3849 | 1.00 | 0.94 | 1.06 | 5.74 |
| Sub-Saharan Africa | Kenya | 6728 | 1.03 | 0.99 | 1.08 | 6.62 |
| Sub-Saharan Africa | Nigeria | 2480 | 1.03 | 0.95 | 1.11 | 5.27 |
| Sub-Saharan Africa | South Africa | 429 | 1.08 | 0.90 | 1.30 | 2.12 |
| Sub-Saharan Africa | Tanzania | 4208 | 1.01 | 0.96 | 1.06 | 6.20 |
| | **Overall** | | 1.01 | 0.98 | 1.05 | |
| | $\tau^2 = 0.00$, $I^2 = 73.5\%$, $H^2 = 3.77$ | | | | | |

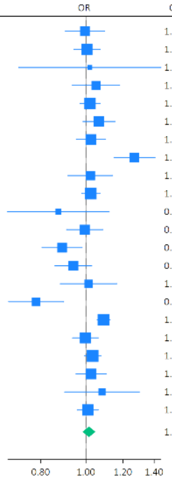

b) Feeling down, depressed or hopeless

| Region | Country | N | OR | 95% CI | | Weight (%) |
|---|---|---|---|---|---|---|
| East Asia & Pacific | Australia | 2009 | 0.98 | 0.89 | 1.09 | 4.06 |
| East Asia & Pacific | China | 3671 | 1.01 | 0.94 | 1.07 | 5.98 |
| East Asia & Pacific | Hong Kong | 306 | 1.02 | 0.73 | 1.42 | 0.62 |
| East Asia & Pacific | Indonesia | 1483 | 1.25 | 1.11 | 1.41 | 3.31 |
| East Asia & Pacific | Japan | 8242 | 0.97 | 0.92 | 1.02 | 6.84 |
| East Asia & Pacific | Philippines | 2060 | 1.09 | 1.01 | 1.18 | 5.05 |
| Europe & Central Asia | Germany | 2923 | 1.05 | 0.97 | 1.13 | 5.24 |
| Europe & Central Asia | Poland | 3180 | 1.16 | 1.03 | 1.30 | 3.39 |
| Europe & Central Asia | Spain | 1547 | 0.94 | 0.84 | 1.06 | 3.51 |
| Europe & Central Asia | Sweden | 8816 | 0.97 | 0.93 | 1.02 | 7.19 |
| Europe & Central Asia | Türkiye | 253 | 0.91 | 0.71 | 1.16 | 1.08 |
| Europe & Central Asia | United Kingdom | 2478 | 0.98 | 0.90 | 1.07 | 4.81 |
| Latin America & Caribbean | Argentina | 1540 | 1.02 | 0.91 | 1.14 | 3.42 |
| Latin America & Caribbean | Brazil | 2225 | 0.96 | 0.87 | 1.05 | 4.48 |
| Latin America & Caribbean | Mexico | 1099 | 0.98 | 0.86 | 1.12 | 2.95 |
| Middle East, North Africa, Afghanistan & Pakistan | Egypt | 2213 | 0.79 | 0.68 | 0.92 | 2.33 |
| North America | United States | 22727 | 1.05 | 1.01 | 1.08 | 8.01 |
| South Asia | India | 3852 | 0.98 | 0.92 | 1.05 | 5.89 |
| Sub-Saharan Africa | Kenya | 6737 | 1.00 | 0.96 | 1.04 | 7.59 |
| Sub-Saharan Africa | Nigeria | 2496 | 1.07 | 0.99 | 1.15 | 5.40 |
| Sub-Saharan Africa | South Africa | 429 | 0.99 | 0.84 | 1.18 | 1.99 |
| Sub-Saharan Africa | Tanzania | 4210 | 1.03 | 0.98 | 1.08 | 6.88 |
| | **Overall** | | 1.01 | 0.98 | 1.04 | |
| | $\tau^2 = 0.00$, $I^2 = 60.9\%$, $H^2 = 2.56$ | | | | | |

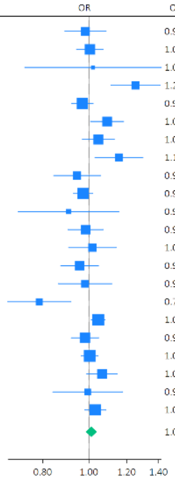

c) Unable to stop or control worrying

| Region | Country | N | OR | 95% CI | | Weight (%) |
|---|---|---|---|---|---|---|
| East Asia & Pacific | Australia | 2007 | 0.96 | 0.87 | 1.06 | 3.99 |
| East Asia & Pacific | China | 3671 | 1.03 | 0.97 | 1.10 | 5.99 |
| East Asia & Pacific | Hong Kong | 309 | 0.99 | 0.68 | 1.44 | 0.47 |
| East Asia & Pacific | Indonesia | 1484 | 1.13 | 1.00 | 1.27 | 3.28 |
| East Asia & Pacific | Japan | 8300 | 1.02 | 0.97 | 1.08 | 6.66 |
| East Asia & Pacific | Philippines | 2058 | 1.05 | 0.98 | 1.14 | 5.17 |
| Europe & Central Asia | Germany | 2933 | 0.97 | 0.90 | 1.05 | 5.29 |
| Europe & Central Asia | Poland | 3182 | 1.31 | 1.18 | 1.46 | 3.59 |
| Europe & Central Asia | Spain | 1549 | 1.07 | 0.94 | 1.20 | 3.14 |
| Europe & Central Asia | Sweden | 8810 | 1.00 | 0.95 | 1.05 | 6.97 |
| Europe & Central Asia | Türkiye | 252 | 0.92 | 0.73 | 1.16 | 1.12 |
| Europe & Central Asia | United Kingdom | 2485 | 0.98 | 0.89 | 1.07 | 4.48 |
| Latin America & Caribbean | Argentina | 1537 | 1.08 | 0.97 | 1.21 | 3.52 |
| Latin America & Caribbean | Brazil | 2222 | 0.97 | 0.90 | 1.05 | 4.90 |
| Latin America & Caribbean | Mexico | 1094 | 0.92 | 0.81 | 1.05 | 2.80 |
| Middle East, North Africa, Afghanistan & Pakistan | Egypt | 2209 | 0.85 | 0.74 | 0.98 | 2.54 |
| North America | United States | 22765 | 1.03 | 0.99 | 1.06 | 8.17 |
| South Asia | India | 3849 | 0.99 | 0.92 | 1.05 | 6.01 |
| Sub-Saharan Africa | Kenya | 6739 | 1.02 | 0.98 | 1.06 | 7.69 |
| Sub-Saharan Africa | Nigeria | 2487 | 1.05 | 0.97 | 1.13 | 5.17 |
| Sub-Saharan Africa | South Africa | 429 | 0.98 | 0.84 | 1.14 | 2.19 |
| Sub-Saharan Africa | Tanzania | 4201 | 1.06 | 1.00 | 1.11 | 6.86 |
| | **Overall** | | 1.02 | 0.99 | 1.05 | |
| | $\tau^2 = 0.00$, $I^2 = 58.9\%$, $H^2 = 2.43$ | | | | | |

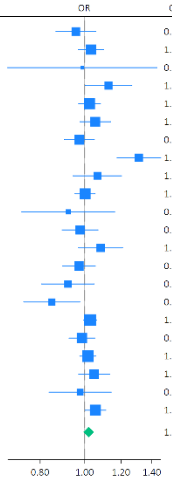

d) Little interest or pleasure in doing things

| Region | Country | N | OR | 95% CI | | Weight (%) |
|---|---|---|---|---|---|---|
| East Asia & Pacific | Australia | 2008 | 0.91 | 0.82 | 1.00 | 3.79 |
| East Asia & Pacific | China | 3670 | 1.02 | 0.96 | 1.09 | 6.11 |
| East Asia & Pacific | Hong Kong | 309 | 0.76 | 0.57 | 1.02 | 0.61 |
| East Asia & Pacific | Indonesia | 1475 | 1.14 | 1.01 | 1.29 | 2.76 |
| East Asia & Pacific | Japan | 8278 | 0.98 | 0.93 | 1.03 | 7.09 |
| East Asia & Pacific | Philippines | 2060 | 1.02 | 0.95 | 1.11 | 5.01 |
| Europe & Central Asia | Germany | 2934 | 1.04 | 0.96 | 1.13 | 4.65 |
| Europe & Central Asia | Poland | 3180 | 1.24 | 1.12 | 1.37 | 3.60 |
| Europe & Central Asia | Spain | 1543 | 1.07 | 0.96 | 1.20 | 3.29 |
| Europe & Central Asia | Sweden | 8763 | 0.98 | 0.93 | 1.02 | 7.54 |
| Europe & Central Asia | Türkiye | 250 | 0.86 | 0.68 | 1.08 | 0.91 |
| Europe & Central Asia | United Kingdom | 2481 | 0.97 | 0.89 | 1.06 | 4.57 |
| Latin America & Caribbean | Argentina | 1538 | 0.99 | 0.90 | 1.09 | 3.76 |
| Latin America & Caribbean | Brazil | 2218 | 1.02 | 0.94 | 1.11 | 4.55 |
| Latin America & Caribbean | Mexico | 1089 | 0.92 | 0.82 | 1.04 | 2.83 |
| Middle East, North Africa, Afghanistan & Pakistan | Egypt | 2203 | 0.97 | 0.85 | 1.11 | 2.39 |
| North America | United States | 22735 | 1.01 | 0.98 | 1.05 | 8.66 |
| South Asia | India | 3848 | 0.96 | 0.91 | 1.03 | 6.01 |
| Sub-Saharan Africa | Kenya | 6740 | 1.04 | 0.99 | 1.08 | 7.94 |
| Sub-Saharan Africa | Nigeria | 2489 | 1.03 | 0.96 | 1.11 | 5.08 |
| Sub-Saharan Africa | South Africa | 430 | 0.90 | 0.76 | 1.07 | 1.57 |
| Sub-Saharan Africa | Tanzania | 4203 | 1.03 | 0.98 | 1.08 | 7.27 |
| | **Overall** | | 1.01 | 0.99 | 1.03 | |
| | $\tau^2 = 0.00$, $I^2 = 51.5\%$, $H^2 = 2.06$ | | | | | |

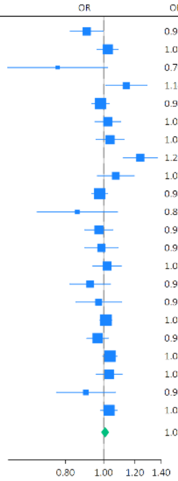

e) Suffering

| Region | Country | N | OR | 95% CI | | Weight (%) |
|---|---|---|---|---|---|---|
| East Asia & Pacific | Australia | 2005 | 1.00 | 0.90 | 1.10 | 2.85 |
| East Asia & Pacific | China | 3672 | 0.98 | 0.92 | 1.04 | 6.11 |
| East Asia & Pacific | Hong Kong | 309 | 0.93 | 0.67 | 1.29 | 0.29 |
| East Asia & Pacific | Indonesia | 1480 | 1.09 | 0.97 | 1.24 | 1.86 |
| East Asia & Pacific | Japan | 8278 | 0.98 | 0.93 | 1.02 | 8.58 |
| East Asia & Pacific | Philippines | 2060 | 1.00 | 0.93 | 1.08 | 4.22 |
| Europe & Central Asia | Germany | 2931 | 1.06 | 0.98 | 1.14 | 4.55 |
| Europe & Central Asia | Poland | 3189 | 1.20 | 1.09 | 1.32 | 3.11 |
| Europe & Central Asia | Spain | 1535 | 0.96 | 0.87 | 1.07 | 2.56 |
| Europe & Central Asia | Sweden | 8811 | 1.01 | 0.97 | 1.05 | 9.50 |
| Europe & Central Asia | Türkiye | 251 | 0.70 | 0.54 | 0.90 | 0.48 |
| Europe & Central Asia | United Kingdom | 2480 | 1.00 | 0.92 | 1.08 | 3.99 |
| Latin America & Caribbean | Argentina | 1544 | 1.02 | 0.92 | 1.13 | 2.80 |
| Latin America & Caribbean | Brazil | 2209 | 1.01 | 0.93 | 1.09 | 3.97 |
| Latin America & Caribbean | Mexico | 1099 | 1.00 | 0.89 | 1.12 | 2.07 |
| Middle East, North Africa, Afghanistan & Pakistan | Egypt | 2213 | 0.89 | 0.77 | 1.04 | 1.34 |
| North America | United States | 22700 | 1.04 | 1.01 | 1.07 | 13.61 |
| South Asia | India | 3850 | 1.02 | 0.95 | 1.09 | 4.97 |
| Sub-Saharan Africa | Kenya | 6743 | 1.01 | 0.97 | 1.05 | 9.87 |
| Sub-Saharan Africa | Nigeria | 2485 | 0.97 | 0.90 | 1.05 | 4.22 |
| Sub-Saharan Africa | South Africa | 428 | 1.03 | 0.87 | 1.23 | 1.01 |
| Sub-Saharan Africa | Tanzania | 4213 | 1.02 | 0.97 | 1.07 | 8.04 |
| | **Overall** | | 1.01 | 0.99 | 1.03 | |
| | $\tau^2 = 0.00$, $I^2 = 24.0\%$, $H^2 = 1.32$ | | | | | |

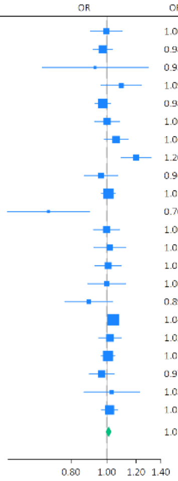

f) Feeling happy

| Region | Country | N | Coef | 95% CI | | Weight (%) |
|---|---|---|---|---|---|---|
| East Asia & Pacific | Australia | 2007 | -0.03 | -0.10 | 0.04 | 5.48 |
| East Asia & Pacific | China | 3671 | 0.05 | 0.00 | 0.10 | 7.12 |
| East Asia & Pacific | Hong Kong | 309 | 0.13 | -0.08 | 0.34 | 0.99 |
| East Asia & Pacific | Indonesia | 1486 | -0.02 | -0.14 | 0.11 | 2.54 |
| East Asia & Pacific | Japan | 8260 | 0.08 | 0.04 | 0.12 | 8.34 |
| East Asia & Pacific | Philippines | 2061 | 0.04 | -0.05 | 0.13 | 4.03 |
| Europe & Central Asia | Germany | 2933 | 0.08 | 0.03 | 0.13 | 7.13 |
| Europe & Central Asia | Poland | 3197 | -0.04 | -0.11 | 0.02 | 6.05 |
| Europe & Central Asia | Spain | 1550 | 0.05 | -0.04 | 0.14 | 3.91 |
| Europe & Central Asia | Sweden | 8834 | 0.04 | 0.01 | 0.08 | 9.14 |
| Europe & Central Asia | Türkiye | 253 | 0.30 | -0.01 | 0.61 | 0.47 |
| Europe & Central Asia | United Kingdom | 2488 | 0.07 | 0.01 | 0.14 | 5.69 |
| Latin America & Caribbean | Argentina | 1545 | 0.09 | 0.01 | 0.18 | 4.16 |
| Latin America & Caribbean | Brazil | 2219 | 0.13 | 0.04 | 0.22 | 4.04 |
| Latin America & Caribbean | Mexico | 1102 | 0.10 | -0.00 | 0.19 | 3.46 |
| Middle East, North Africa, Afghanistan & Pakistan | Egypt | 2211 | 0.21 | -0.01 | 0.44 | 0.90 |
| North America | United States | 22767 | -0.00 | -0.03 | 0.03 | 9.95 |
| South Asia | India | 3863 | 0.07 | -0.04 | 0.18 | 2.92 |
| Sub-Saharan Africa | Kenya | 6741 | 0.03 | -0.04 | 0.10 | 5.52 |
| Sub-Saharan Africa | Nigeria | 2499 | -0.03 | -0.14 | 0.08 | 3.00 |
| Sub-Saharan Africa | South Africa | 431 | -0.12 | -0.42 | 0.18 | 0.51 |
| Sub-Saharan Africa | Tanzania | 4212 | 0.09 | 0.01 | 0.17 | 4.64 |
| | **Overall** | | 0.05 | 0.02 | 0.07 | |
| | $\tau^2 = 0.00$, $I^2 = 50.4\%$, $H^2 = 2.02$ | | | | | |

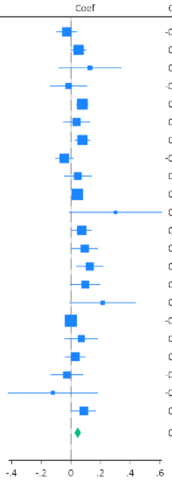

##### g) Understanding purpose in life

| Region | Country | N | Coef | 95% CI | Weight (%) |
|---|---|---|---|---|---|
| East Asia & Pacific | Australia | 2008 | -0.04 | [ -0.14, 0.06] | 4.22 |
| East Asia & Pacific | China | 3670 | 0.07 | [ 0.01, 0.13] | 5.94 |
| East Asia & Pacific | Hong Kong | 309 | 0.14 | [ -0.06, 0.34] | 1.82 |
| East Asia & Pacific | Indonesia | 1487 | 0.04 | [ -0.08, 0.17] | 3.41 |
| East Asia & Pacific | Japan | 8262 | 0.11 | [ 0.07, 0.15] | 6.73 |
| East Asia & Pacific | Philippines | 2060 | -0.00 | [ -0.08, 0.07] | 5.17 |
| Europe & Central Asia | Germany | 2931 | 0.00 | [ -0.06, 0.06] | 5.83 |
| Europe & Central Asia | Poland | 3187 | -0.11 | [ -0.18, -0.03] | 5.22 |
| Europe & Central Asia | Spain | 1543 | 0.05 | [ -0.04, 0.15] | 4.52 |
| Europe & Central Asia | Sweden | 8766 | 0.03 | [ -0.01, 0.07] | 6.58 |
| Europe & Central Asia | Türkiye | 252 | 0.30 | [ 0.02, 0.57] | 1.10 |
| Europe & Central Asia | United Kingdom | 2478 | 0.01 | [ -0.06, 0.09] | 5.27 |
| Latin America & Caribbean | Argentina | 1540 | 0.13 | [ 0.03, 0.23] | 4.19 |
| Latin America & Caribbean | Brazil | 2216 | 0.09 | [ 0.00, 0.17] | 4.77 |
| Latin America & Caribbean | Mexico | 1099 | 0.14 | [ 0.07, 0.22] | 5.25 |
| Middle East, North Africa, Afghanistan & Pakistan | Egypt | 2193 | 0.18 | [ 0.02, 0.34] | 2.58 |
| North America | United States | 22735 | 0.04 | [ 0.01, 0.07] | 6.88 |
| South Asia | India | 3849 | 0.09 | [ -0.01, 0.19] | 4.31 |
| Sub-Saharan Africa | Kenya | 6740 | -0.01 | [ -0.07, 0.06] | 5.49 |
| Sub-Saharan Africa | Nigeria | 2494 | -0.05 | [ -0.15, 0.05] | 4.18 |
| Sub-Saharan Africa | South Africa | 431 | -0.25 | [ -0.49, -0.01] | 1.40 |
| Sub-Saharan Africa | Tanzania | 4176 | 0.12 | [ 0.04, 0.19] | 5.16 |
| | **Overall** | | 0.05 | [ 0.01, 0.08] | |

$\tau^2 = 0.00$, $I^2 = 72.3\%$, $H^2 = 3.61$

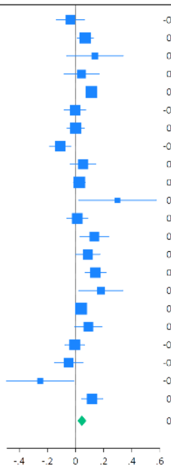

##### h) Life satisfaction

| Region | Country | N | Coef | 95% CI | Weight (%) |
|---|---|---|---|---|---|
| East Asia & Pacific | Australia | 2010 | 0.03 | [ -0.05, 0.10] | 4.91 |
| East Asia & Pacific | China | 3652 | 0.05 | [ -0.01, 0.10] | 6.85 |
| East Asia & Pacific | Hong Kong | 297 | 0.23 | [ 0.04, 0.43] | 1.25 |
| East Asia & Pacific | Indonesia | 1484 | -0.01 | [ -0.12, 0.10] | 3.00 |
| East Asia & Pacific | Japan | 8226 | 0.08 | [ 0.04, 0.11] | 8.51 |
| East Asia & Pacific | Philippines | 2059 | 0.01 | [ -0.08, 0.09] | 4.19 |
| Europe & Central Asia | Germany | 2913 | 0.08 | [ 0.03, 0.14] | 6.81 |
| Europe & Central Asia | Poland | 3180 | -0.07 | [ -0.13, -0.00] | 5.82 |
| Europe & Central Asia | Spain | 1542 | 0.07 | [ -0.01, 0.16] | 4.42 |
| Europe & Central Asia | Sweden | 8822 | 0.02 | [ -0.02, 0.06] | 8.69 |
| Europe & Central Asia | Türkiye | 253 | 0.18 | [ -0.09, 0.46] | 0.66 |
| Europe & Central Asia | United Kingdom | 2459 | 0.09 | [ 0.03, 0.15] | 6.40 |
| Latin America & Caribbean | Argentina | 1544 | 0.11 | [ 0.01, 0.20] | 3.82 |
| Latin America & Caribbean | Brazil | 2219 | 0.07 | [ -0.02, 0.16] | 4.22 |
| Latin America & Caribbean | Mexico | 1102 | 0.06 | [ -0.05, 0.16] | 3.41 |
| Middle East, North Africa, Afghanistan & Pakistan | Egypt | 2199 | 0.16 | [ 0.01, 0.31] | 1.89 |
| North America | United States | 22756 | -0.00 | [ -0.03, 0.02] | 9.43 |
| South Asia | India | 3859 | 0.10 | [ -0.01, 0.22] | 3.00 |
| Sub-Saharan Africa | Kenya | 6744 | 0.03 | [ -0.05, 0.10] | 5.16 |
| Sub-Saharan Africa | Nigeria | 2499 | -0.01 | [ -0.13, 0.10] | 2.81 |
| Sub-Saharan Africa | South Africa | 426 | -0.13 | [ -0.43, 0.17] | 0.56 |
| Sub-Saharan Africa | Tanzania | 4211 | -0.02 | [ -0.11, 0.07] | 4.18 |
| | **Overall** | | 0.04 | [ 0.02, 0.06] | |

$\tau^2 = 0.00$, $I^2 = 52.5\%$, $H^2 = 2.11$

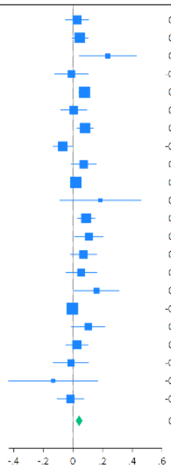

##### i) Life evaluation: Today

| Region | Country | N | Coef | 95% CI | Weight (%) |
|---|---|---|---|---|---|
| East Asia & Pacific | Australia | 2011 | -0.03 | [ -0.10, 0.05] | 5.09 |
| East Asia & Pacific | China | 3671 | 0.02 | [ -0.03, 0.08] | 6.37 |
| East Asia & Pacific | Hong Kong | 309 | 0.17 | [ 0.01, 0.33] | 2.05 |
| East Asia & Pacific | Indonesia | 1485 | 0.08 | [ -0.06, 0.22] | 2.51 |
| East Asia & Pacific | Japan | 8293 | 0.12 | [ 0.08, 0.16] | 7.61 |
| East Asia & Pacific | Philippines | 2056 | 0.12 | [ 0.03, 0.21] | 4.43 |
| Europe & Central Asia | Germany | 2928 | 0.11 | [ 0.05, 0.16] | 6.76 |
| Europe & Central Asia | Poland | 3195 | 0.00 | [ -0.06, 0.07] | 5.75 |
| Europe & Central Asia | Spain | 1550 | 0.04 | [ -0.04, 0.12] | 4.90 |
| Europe & Central Asia | Sweden | 8831 | 0.03 | [ -0.00, 0.06] | 8.02 |
| Europe & Central Asia | Türkiye | 252 | 0.20 | [ -0.10, 0.50] | 0.67 |
| Europe & Central Asia | United Kingdom | 2485 | 0.09 | [ 0.03, 0.15] | 5.95 |
| Latin America & Caribbean | Argentina | 1547 | 0.13 | [ 0.03, 0.24] | 3.74 |
| Latin America & Caribbean | Brazil | 2226 | 0.12 | [ 0.03, 0.20] | 4.43 |
| Latin America & Caribbean | Mexico | 1102 | 0.03 | [ -0.07, 0.13] | 3.92 |
| Middle East, North Africa, Afghanistan & Pakistan | Egypt | 2201 | 0.26 | [ 0.07, 0.44] | 1.58 |
| North America | United States | 22777 | 0.02 | [ -0.01, 0.04] | 8.50 |
| South Asia | India | 3842 | 0.10 | [ -0.01, 0.21] | 3.38 |
| Sub-Saharan Africa | Kenya | 6744 | 0.03 | [ -0.04, 0.10] | 5.58 |
| Sub-Saharan Africa | Nigeria | 2503 | -0.04 | [ -0.16, 0.07] | 3.21 |
| Sub-Saharan Africa | South Africa | 429 | -0.17 | [ -0.46, 0.12] | 0.73 |
| Sub-Saharan Africa | Tanzania | 4215 | 0.14 | [ 0.06, 0.22] | 4.81 |
| | **Overall** | | 0.06 | [ 0.04, 0.09] | |

$\tau^2 = 0.00$, $I^2 = 63.5\%$, $H^2 = 2.74$

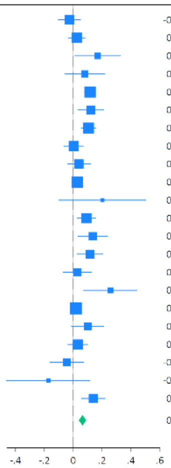

##### j) Life evaluation: Five years from now

| Region | Country | N | Coef | 95% CI | Weight (%) |
|---|---|---|---|---|---|
| East Asia & Pacific | Australia | 2010 | 0.01 | [ -0.08, 0.09] | 4.65 |
| East Asia & Pacific | China | 3653 | 0.04 | [ -0.02, 0.09] | 6.53 |
| East Asia & Pacific | Hong Kong | 300 | 0.21 | [ 0.01, 0.41] | 1.30 |
| East Asia & Pacific | Indonesia | 1467 | 0.09 | [ -0.01, 0.20] | 3.55 |
| East Asia & Pacific | Japan | 8297 | 0.10 | [ 0.07, 0.14] | 7.52 |
| East Asia & Pacific | Philippines | 2057 | 0.04 | [ -0.04, 0.12] | 4.77 |
| Europe & Central Asia | Germany | 2912 | 0.11 | [ 0.06, 0.17] | 6.48 |
| Europe & Central Asia | Poland | 3153 | 0.01 | [ -0.06, 0.09] | 4.93 |
| Europe & Central Asia | Spain | 1540 | 0.05 | [ -0.04, 0.13] | 4.44 |
| Europe & Central Asia | Sweden | 8806 | 0.04 | [ 0.00, 0.07] | 7.92 |
| Europe & Central Asia | Türkiye | 236 | 0.12 | [ -0.19, 0.43] | 0.60 |
| Europe & Central Asia | United Kingdom | 2460 | 0.10 | [ 0.03, 0.17] | 5.39 |
| Latin America & Caribbean | Argentina | 1510 | 0.05 | [ -0.06, 0.15] | 3.64 |
| Latin America & Caribbean | Brazil | 2216 | 0.08 | [ 0.00, 0.15] | 5.13 |
| Latin America & Caribbean | Mexico | 1078 | 0.06 | [ -0.03, 0.16] | 3.98 |
| Middle East, North Africa, Afghanistan & Pakistan | Egypt | 1632 | 0.25 | [ 0.05, 0.44] | 1.37 |
| North America | United States | 22765 | -0.03 | [ -0.06, 0.00] | 8.12 |
| South Asia | India | 3576 | 0.10 | [ 0.00, 0.20] | 3.80 |
| Sub-Saharan Africa | Kenya | 6699 | 0.03 | [ -0.03, 0.09] | 5.73 |
| Sub-Saharan Africa | Nigeria | 2470 | -0.06 | [ -0.15, 0.03] | 4.17 |
| Sub-Saharan Africa | South Africa | 423 | -0.19 | [ -0.43, 0.05] | 0.98 |
| Sub-Saharan Africa | Tanzania | 3979 | 0.11 | [ 0.04, 0.19] | 4.98 |
| | **Overall** | | 0.05 | [ 0.03, 0.08] | |

$\tau^2 = 0.00$, $I^2 = 61.7\%$, $H^2 = 2.61$

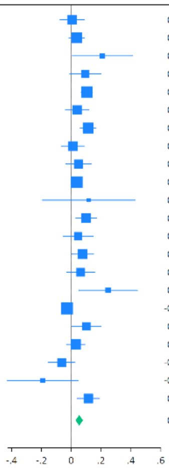

##### k) Feeling lonely

| Region | Country | N | Coef | 95% CI | Weight (%) |
|---|---|---|---|---|---|
| East Asia & Pacific | Australia | 2010 | 0.01 | [ -0.10, 0.12] | 2.82 |
| East Asia & Pacific | China | 3672 | -0.03 | [ -0.10, 0.04] | 6.64 |
| East Asia & Pacific | Hong Kong | 308 | -0.31 | [ -0.58, -0.03] | 0.42 |
| East Asia & Pacific | Indonesia | 1487 | 0.03 | [ -0.16, 0.21] | 0.96 |
| East Asia & Pacific | Japan | 8309 | -0.07 | [ -0.13, -0.02] | 11.30 |
| East Asia & Pacific | Philippines | 2063 | -0.06 | [ -0.18, 0.06] | 2.43 |
| Europe & Central Asia | Germany | 2938 | -0.03 | [ -0.10, 0.04] | 6.02 |
| Europe & Central Asia | Poland | 3190 | 0.02 | [ -0.08, 0.12] | 3.41 |
| Europe & Central Asia | Spain | 1553 | 0.02 | [ -0.10, 0.14] | 2.26 |
| Europe & Central Asia | Sweden | 8848 | -0.04 | [ -0.08, 0.00] | 17.28 |
| Europe & Central Asia | Türkiye | 250 | 0.02 | [ -0.31, 0.34] | 0.32 |
| Europe & Central Asia | United Kingdom | 2476 | -0.02 | [ -0.11, 0.07] | 4.43 |
| Latin America & Caribbean | Argentina | 1549 | -0.12 | [ -0.26, 0.02] | 1.60 |
| Latin America & Caribbean | Brazil | 2221 | -0.06 | [ -0.18, 0.07] | 2.13 |
| Latin America & Caribbean | Mexico | 1099 | -0.09 | [ -0.27, 0.08] | 1.10 |
| Middle East, North Africa, Afghanistan & Pakistan | Egypt | 2210 | -0.12 | [ -0.43, 0.20] | 0.33 |
| North America | United States | 22786 | -0.03 | [ -0.07, 0.00] | 22.77 |
| South Asia | India | 3863 | 0.12 | [ -0.01, 0.25] | 2.04 |
| Sub-Saharan Africa | Kenya | 6739 | -0.03 | [ -0.11, 0.05] | 5.19 |
| Sub-Saharan Africa | Nigeria | 2496 | 0.08 | [ -0.05, 0.22] | 1.84 |
| Sub-Saharan Africa | South Africa | 431 | 0.18 | [ -0.09, 0.46] | 0.44 |
| Sub-Saharan Africa | Tanzania | 4213 | -0.04 | [ -0.13, 0.05] | 4.25 |
| | **Overall** | | -0.03 | [ -0.05, -0.01] | |

$\tau^2 = 0.00$, $I^2 = 0.00\%$, $H^2 = 1.00$

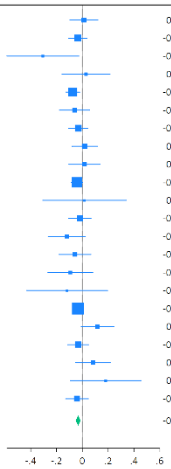

##### l) Life is worthwhile

| Region | Country | N | Coef | 95% CI | Weight (%) |
|---|---|---|---|---|---|
| East Asia & Pacific | Australia | 2008 | 0.00 | [ -0.09, 0.09] | 4.33 |
| East Asia & Pacific | China | 3669 | 0.04 | [ -0.01, 0.09] | 6.92 |
| East Asia & Pacific | Hong Kong | 306 | 0.19 | [ 0.02, 0.35] | 1.89 |
| East Asia & Pacific | Indonesia | 1486 | 0.05 | [ -0.05, 0.15] | 3.79 |
| East Asia & Pacific | Japan | 8290 | 0.09 | [ 0.05, 0.13] | 7.93 |
| East Asia & Pacific | Philippines | 2060 | 0.00 | [ -0.09, 0.09] | 4.43 |
| Europe & Central Asia | Germany | 2932 | 0.07 | [ 0.01, 0.12] | 6.60 |
| Europe & Central Asia | Poland | 3189 | -0.12 | [ -0.19, -0.05] | 5.69 |
| Europe & Central Asia | Spain | 1541 | 0.03 | [ -0.07, 0.13] | 3.87 |
| Europe & Central Asia | Sweden | 8832 | 0.05 | [ 0.01, 0.08] | 7.88 |
| Europe & Central Asia | Türkiye | 253 | 0.17 | [ -0.10, 0.44] | 0.78 |
| Europe & Central Asia | United Kingdom | 2473 | 0.05 | [ -0.01, 0.11] | 6.23 |
| Latin America & Caribbean | Argentina | 1541 | 0.12 | [ 0.02, 0.22] | 3.95 |
| Latin America & Caribbean | Brazil | 2224 | 0.11 | [ 0.02, 0.20] | 4.52 |
| Latin America & Caribbean | Mexico | 1101 | 0.06 | [ -0.04, 0.17] | 3.49 |
| Middle East, North Africa, Afghanistan & Pakistan | Egypt | 2207 | 0.09 | [ -0.16, 0.34] | 0.91 |
| North America | United States | 22770 | 0.03 | [ -0.00, 0.06] | 8.47 |
| South Asia | India | 3844 | 0.01 | [ -0.09, 0.12] | 3.70 |
| Sub-Saharan Africa | Kenya | 6743 | 0.03 | [ -0.04, 0.10] | 5.47 |
| Sub-Saharan Africa | Nigeria | 2498 | 0.04 | [ -0.07, 0.15] | 3.39 |
| Sub-Saharan Africa | South Africa | 428 | 0.05 | [ -0.19, 0.29] | 0.94 |
| Sub-Saharan Africa | Tanzania | 4209 | 0.11 | [ 0.03, 0.19] | 4.81 |
| | **Overall** | | 0.05 | [ 0.02, 0.07] | |

$\tau^2 = 0.00$, $I^2 = 57.9\%$, $H^2 = 2.38$

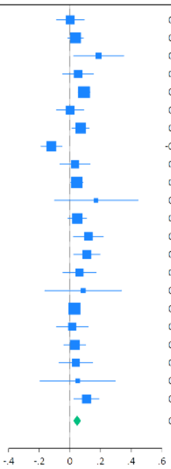

m) Feeling grateful

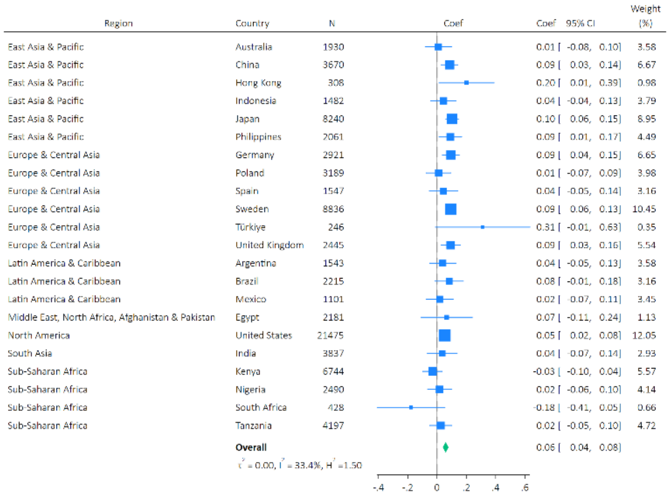

n) Feeling at peace

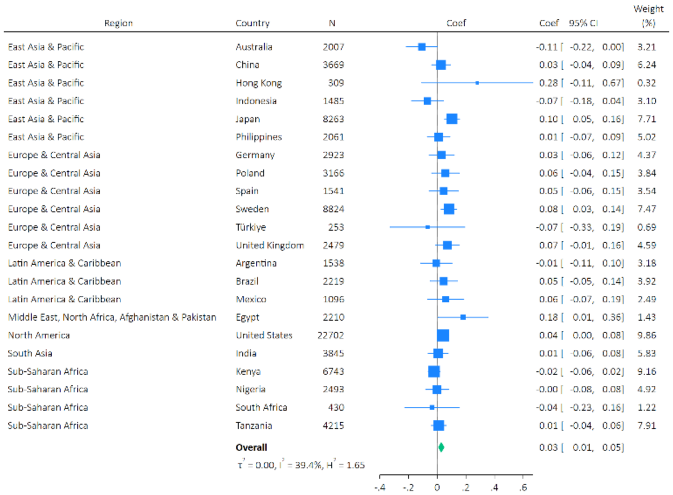

o) Remain hopeful about the future

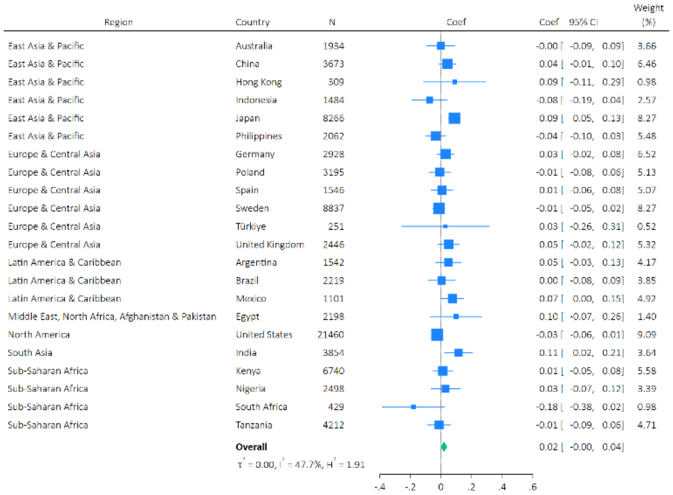

p) Feeling very capable in most things

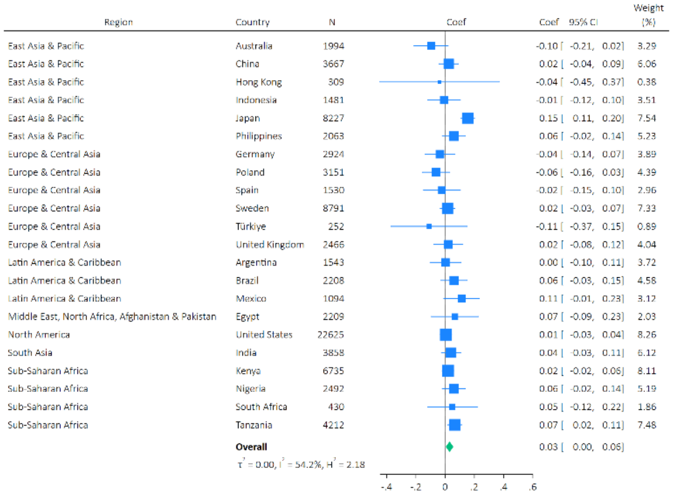

q) Expect more good things to happen to you

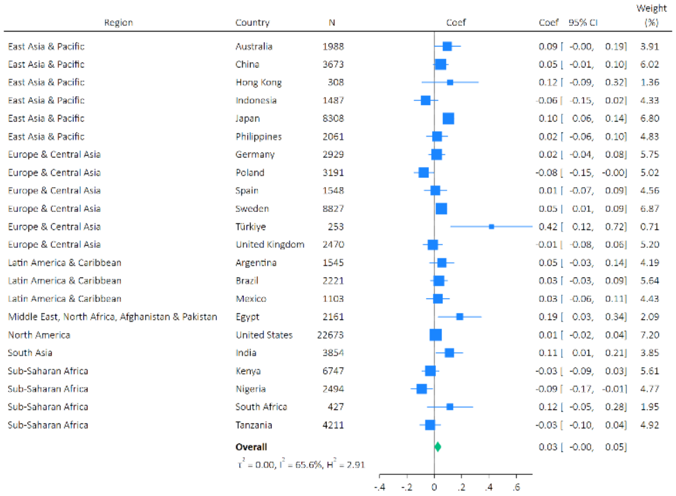

r) Overall mental health

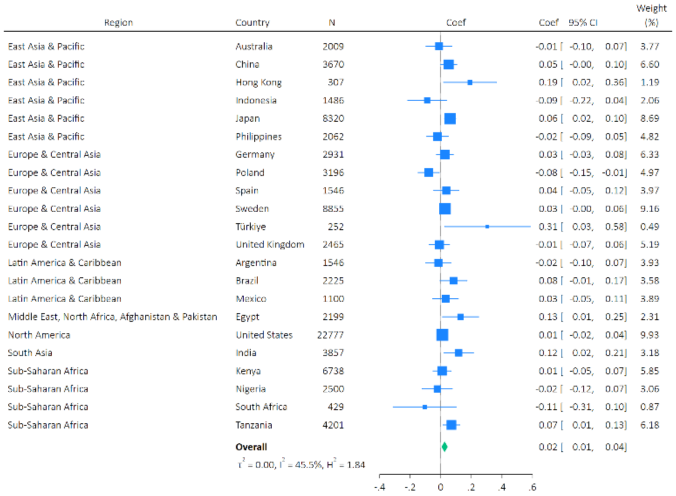

s) Overall physical health

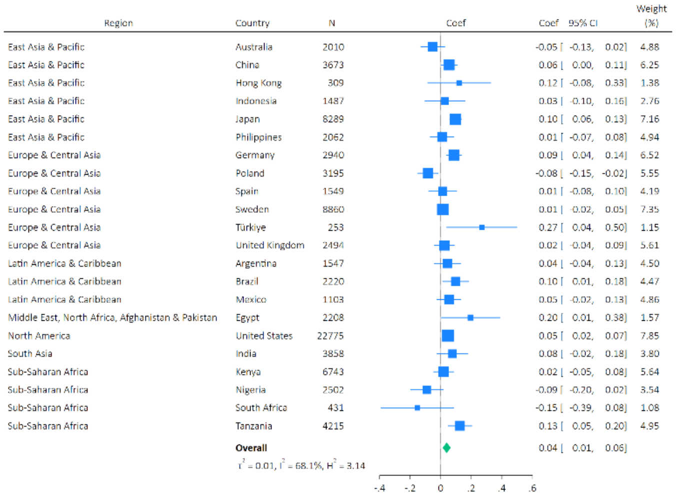

#### Supplementary Figure S6: Meta-analysis forest plots for the effects of arts engagement on later health and wellbeing outcomes (nested adjusted only for socio-demographic confounders, family and social relationships confounders and baseline health outcome)

a) Feeling nervous, anxious, or on edge

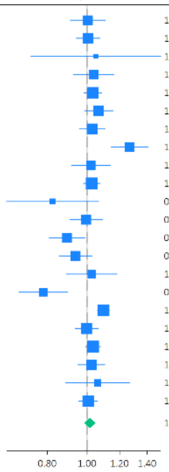

| Region | Country | N | OR | 95% CI | Weight (%) |
|---|---|---|---|---|---|
| East Asia & Pacific | Australia | 2000 | 1.00 | [0.91, 1.11] | 4.39 |
| East Asia & Pacific | China | 3672 | 1.01 | [0.94, 1.07] | 5.69 |
| East Asia & Pacific | Hong Kong | 309 | 1.05 | [0.73, 1.51] | 0.70 |
| East Asia & Pacific | Indonesia | 1487 | 1.04 | [0.93, 1.16] | 3.82 |
| East Asia & Pacific | Japan | 8240 | 1.03 | [0.98, 1.08] | 6.32 |
| East Asia & Pacific | Philippines | 2061 | 1.07 | [0.99, 1.16] | 5.09 |
| Europe & Central Asia | Germany | 2921 | 1.03 | [0.96, 1.10] | 5.42 |
| Europe & Central Asia | Poland | 3175 | 1.27 | [1.15, 1.41] | 4.22 |
| Europe & Central Asia | Spain | 1546 | 1.02 | [0.92, 1.14] | 3.97 |
| Europe & Central Asia | Sweden | 8822 | 1.03 | [0.98, 1.07] | 6.46 |
| Europe & Central Asia | Türkiye | 251 | 0.82 | [0.64, 1.07] | 1.27 |
| Europe & Central Asia | United Kingdom | 2476 | 0.99 | [0.91, 1.09] | 4.66 |
| Latin America & Caribbean | Argentina | 1541 | 0.89 | [0.81, 0.99] | 4.33 |
| Latin America & Caribbean | Brazil | 2217 | 0.94 | [0.85, 1.03] | 4.60 |
| Latin America & Caribbean | Mexico | 1099 | 1.03 | [0.89, 1.18] | 3.02 |
| Middle East, North Africa, Afghanistan & Pakistan | Egypt | 2211 | 0.78 | [0.68, 0.90] | 3.17 |
| North America | United States | 22708 | 1.10 | [1.06, 1.13] | 6.91 |
| South Asia | India | 3849 | 1.00 | [0.94, 1.06] | 5.73 |
| Sub-Saharan Africa | Kenya | 6728 | 1.03 | [0.99, 1.08] | 6.62 |
| Sub-Saharan Africa | Nigeria | 2480 | 1.02 | [0.95, 1.10] | 5.27 |
| Sub-Saharan Africa | South Africa | 429 | 1.06 | [0.89, 1.27] | 2.20 |
| Sub-Saharan Africa | Tanzania | 4208 | 1.01 | [0.95, 1.06] | 6.18 |
| | **Overall** | | 1.02 | [0.98, 1.05] | |

$\tau^2 = 0.00$, $I^2 = 73.5\%$, $H^2 = 3.77$

b) Feeling down, depressed or hopeless

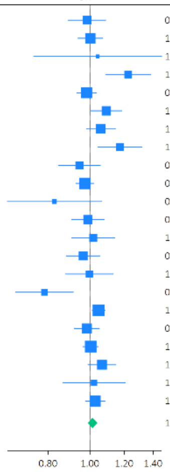

| Region | Country | N | OR | 95% CI | Weight (%) |
|---|---|---|---|---|---|
| East Asia & Pacific | Australia | 2009 | 0.98 | [0.89, 1.09] | 3.92 |
| East Asia & Pacific | China | 3671 | 1.00 | [0.94, 1.07] | 6.04 |
| East Asia & Pacific | Hong Kong | 306 | 1.04 | [0.74, 1.46] | 0.52 |
| East Asia & Pacific | Indonesia | 1483 | 1.23 | [1.09, 1.38] | 3.09 |
| East Asia & Pacific | Japan | 8242 | 0.98 | [0.93, 1.03] | 7.06 |
| East Asia & Pacific | Philippines | 2060 | 1.09 | [1.01, 1.18] | 4.96 |
| Europe & Central Asia | Germany | 2923 | 1.06 | [0.98, 1.14] | 5.20 |
| Europe & Central Asia | Poland | 3180 | 1.17 | [1.04, 1.32] | 3.19 |
| Europe & Central Asia | Spain | 1547 | 0.95 | [0.85, 1.06] | 3.43 |
| Europe & Central Asia | Sweden | 8816 | 0.97 | [0.93, 1.02] | 7.48 |
| Europe & Central Asia | Türkiye | 253 | 0.83 | [0.64, 1.06] | 0.92 |
| Europe & Central Asia | United Kingdom | 2478 | 0.99 | [0.91, 1.08] | 4.65 |
| Latin America & Caribbean | Argentina | 1540 | 1.02 | [0.91, 1.14] | 3.30 |
| Latin America & Caribbean | Brazil | 2225 | 0.96 | [0.88, 1.05] | 4.45 |
| Latin America & Caribbean | Mexico | 1099 | 1.00 | [0.88, 1.13] | 2.85 |
| Middle East, North Africa, Afghanistan & Pakistan | Egypt | 2213 | 0.78 | [0.67, 0.91] | 2.15 |
| North America | United States | 22727 | 1.05 | [1.01, 1.08] | 8.52 |
| South Asia | India | 3852 | 0.98 | [0.92, 1.05] | 5.93 |
| Sub-Saharan Africa | Kenya | 6737 | 1.00 | [0.96, 1.05] | 7.99 |
| Sub-Saharan Africa | Nigeria | 2496 | 1.07 | [0.99, 1.15] | 5.38 |
| Sub-Saharan Africa | South Africa | 429 | 1.02 | [0.87, 1.21] | 1.88 |
| Sub-Saharan Africa | Tanzania | 4210 | 1.03 | [0.98, 1.08] | 7.09 |
| | **Overall** | | 1.01 | [0.99, 1.04] | |

$\tau^2 = 0.00$, $I^2 = 55.4\%$, $H^2 = 2.24$

c) Unable to stop or control worrying

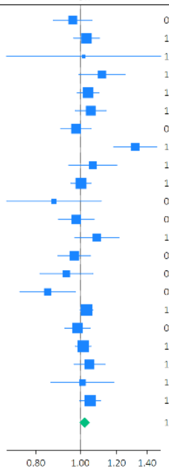

| Region | Country | N | OR | 95% CI | Weight (%) |
|---|---|---|---|---|---|
| East Asia & Pacific | Australia | 2007 | 0.96 | [0.87, 1.06] | 4.04 |
| East Asia & Pacific | China | 3671 | 1.03 | [0.97, 1.10] | 6.00 |
| East Asia & Pacific | Hong Kong | 309 | 1.02 | [0.69, 1.50] | 0.43 |
| East Asia & Pacific | Indonesia | 1484 | 1.12 | [0.99, 1.25] | 3.29 |
| East Asia & Pacific | Japan | 8300 | 1.04 | [0.98, 1.10] | 6.71 |
| East Asia & Pacific | Philippines | 2058 | 1.05 | [0.97, 1.14] | 5.14 |
| Europe & Central Asia | Germany | 2933 | 0.98 | [0.91, 1.06] | 5.24 |
| Europe & Central Asia | Poland | 3182 | 1.32 | [1.18, 1.47] | 3.58 |
| Europe & Central Asia | Spain | 1549 | 1.06 | [0.94, 1.20] | 3.12 |
| Europe & Central Asia | Sweden | 8810 | 1.00 | [0.95, 1.06] | 7.00 |
| Europe & Central Asia | Türkiye | 252 | 0.88 | [0.69, 1.11] | 1.07 |
| Europe & Central Asia | United Kingdom | 2485 | 0.98 | [0.90, 1.07] | 4.45 |
| Latin America & Caribbean | Argentina | 1537 | 1.09 | [0.97, 1.21] | 3.51 |
| Latin America & Caribbean | Brazil | 2222 | 0.97 | [0.89, 1.05] | 4.92 |
| Latin America & Caribbean | Mexico | 1094 | 0.93 | [0.82, 1.07] | 2.73 |
| Middle East, North Africa, Afghanistan & Pakistan | Egypt | 2209 | 0.85 | [0.74, 0.98] | 2.55 |
| North America | United States | 22765 | 1.03 | [1.00, 1.07] | 8.27 |
| South Asia | India | 3849 | 0.99 | [0.92, 1.05] | 6.03 |
| Sub-Saharan Africa | Kenya | 6739 | 1.01 | [0.97, 1.06] | 7.77 |
| Sub-Saharan Africa | Nigeria | 2487 | 1.05 | [0.97, 1.13] | 5.16 |
| Sub-Saharan Africa | South Africa | 429 | 1.01 | [0.86, 1.19] | 2.09 |
| Sub-Saharan Africa | Tanzania | 4201 | 1.05 | [1.00, 1.11] | 6.89 |
| | **Overall** | | 1.02 | [1.00, 1.05] | |

$\tau^2 = 0.00$, $I^2 = 58.0\%$, $H^2 = 2.38$

d) Little interest or pleasure in doing things

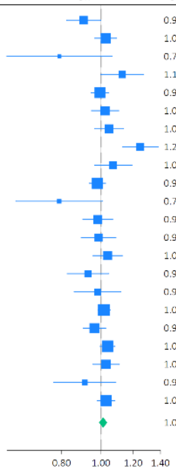

| Region | Country | N | OR | 95% CI | Weight (%) |
|---|---|---|---|---|---|
| East Asia & Pacific | Australia | 2008 | 0.91 | [0.82, 1.00] | 3.73 |
| East Asia & Pacific | China | 3670 | 1.03 | [0.97, 1.09] | 6.13 |
| East Asia & Pacific | Hong Kong | 309 | 0.79 | [0.59, 1.06] | 0.57 |
| East Asia & Pacific | Indonesia | 1475 | 1.13 | [1.00, 1.27] | 2.73 |
| East Asia & Pacific | Japan | 8278 | 0.99 | [0.94, 1.04] | 7.19 |
| East Asia & Pacific | Philippines | 2060 | 1.02 | [0.95, 1.10] | 4.94 |
| Europe & Central Asia | Germany | 2934 | 1.05 | [0.96, 1.14] | 4.64 |
| Europe & Central Asia | Poland | 3180 | 1.25 | [1.13, 1.38] | 3.55 |
| Europe & Central Asia | Spain | 1543 | 1.07 | [0.96, 1.19] | 3.32 |
| Europe & Central Asia | Sweden | 8763 | 0.98 | [0.94, 1.03] | 7.65 |
| Europe & Central Asia | Türkiye | 250 | 0.79 | [0.62, 1.01] | 0.82 |
| Europe & Central Asia | United Kingdom | 2481 | 0.98 | [0.90, 1.07] | 4.45 |
| Latin America & Caribbean | Argentina | 1538 | 0.99 | [0.89, 1.09] | 3.66 |
| Latin America & Caribbean | Brazil | 2218 | 1.04 | [0.96, 1.13] | 4.56 |
| Latin America & Caribbean | Mexico | 1089 | 0.93 | [0.82, 1.04] | 2.83 |
| Middle East, North Africa, Afghanistan & Pakistan | Egypt | 2203 | 0.98 | [0.86, 1.12] | 2.38 |
| North America | United States | 22735 | 1.02 | [0.98, 1.05] | 8.84 |
| South Asia | India | 3848 | 0.96 | [0.90, 1.03] | 6.02 |
| Sub-Saharan Africa | Kenya | 6740 | 1.04 | [0.99, 1.08] | 8.08 |
| Sub-Saharan Africa | Nigeria | 2489 | 1.03 | [0.95, 1.11] | 5.08 |
| Sub-Saharan Africa | South Africa | 430 | 0.91 | [0.76, 1.09] | 1.49 |
| Sub-Saharan Africa | Tanzania | 4203 | 1.03 | [0.98, 1.08] | 7.35 |
| | **Overall** | | 1.01 | [0.99, 1.03] | |

$\tau^2 = 0.00$, $I^2 = 49.5\%$, $H^2 = 1.98$

e) Suffering

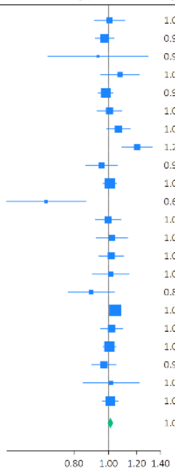

| Region | Country | N | OR | 95% CI | Weight (%) |
|---|---|---|---|---|---|
| East Asia & Pacific | Australia | 2005 | 1.01 | [0.91, 1.11] | 2.86 |
| East Asia & Pacific | China | 3672 | 0.97 | [0.92, 1.04] | 6.12 |
| East Asia & Pacific | Hong Kong | 309 | 0.93 | [0.68, 1.29] | 0.30 |
| East Asia & Pacific | Indonesia | 1480 | 1.08 | [0.95, 1.22] | 1.86 |
| East Asia & Pacific | Japan | 8278 | 0.98 | [0.94, 1.03] | 8.61 |
| East Asia & Pacific | Philippines | 2060 | 1.01 | [0.93, 1.09] | 4.17 |
| Europe & Central Asia | Germany | 2931 | 1.07 | [0.99, 1.15] | 4.50 |
| Europe & Central Asia | Poland | 3189 | 1.20 | [1.09, 1.32] | 2.92 |
| Europe & Central Asia | Spain | 1535 | 0.96 | [0.86, 1.06] | 2.63 |
| Europe & Central Asia | Sweden | 8811 | 1.01 | [0.97, 1.05] | 9.59 |
| Europe & Central Asia | Türkiye | 251 | 0.67 | [0.52, 0.86] | 0.47 |
| Europe & Central Asia | United Kingdom | 2480 | 1.00 | [0.92, 1.08] | 3.89 |
| Latin America & Caribbean | Argentina | 1544 | 1.02 | [0.92, 1.13] | 2.76 |
| Latin America & Caribbean | Brazil | 2209 | 1.02 | [0.94, 1.10] | 3.97 |
| Latin America & Caribbean | Mexico | 1099 | 1.01 | [0.90, 1.14] | 2.03 |
| Middle East, North Africa, Afghanistan & Pakistan | Egypt | 2213 | 0.89 | [0.77, 1.04] | 1.34 |
| North America | United States | 22700 | 1.04 | [1.01, 1.08] | 13.81 |
| South Asia | India | 3850 | 1.02 | [0.95, 1.09] | 4.95 |
| Sub-Saharan Africa | Kenya | 6743 | 1.01 | [0.96, 1.05] | 9.96 |
| Sub-Saharan Africa | Nigeria | 2485 | 0.97 | [0.90, 1.05] | 4.24 |
| Sub-Saharan Africa | South Africa | 428 | 1.02 | [0.85, 1.22] | 0.91 |
| Sub-Saharan Africa | Tanzania | 4213 | 1.01 | [0.96, 1.06] | 8.11 |
| | **Overall** | | 1.01 | [0.99, 1.03] | |

$\tau^2 = 0.00$, $I^2 = 22.9\%$, $H^2 = 1.30$

f) Feeling happy

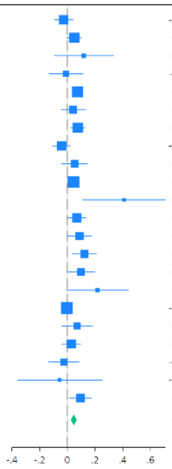

| Region | Country | N | Coef | 95% CI | Weight (%) |
|---|---|---|---|---|---|
| East Asia & Pacific | Australia | 2007 | -0.03 | [-0.09, 0.04] | 5.55 |
| East Asia & Pacific | China | 3671 | 0.05 | [-0.00, 0.10] | 7.09 |
| East Asia & Pacific | Hong Kong | 309 | 0.12 | [-0.09, 0.33] | 0.95 |
| East Asia & Pacific | Indonesia | 1486 | -0.01 | [-0.13, 0.11] | 2.51 |
| East Asia & Pacific | Japan | 8260 | 0.07 | [0.03, 0.11] | 8.44 |
| East Asia & Pacific | Philippines | 2061 | 0.04 | [-0.04, 0.13] | 4.06 |
| Europe & Central Asia | Germany | 2933 | 0.08 | [0.03, 0.13] | 7.17 |
| Europe & Central Asia | Poland | 3197 | -0.04 | [-0.10, 0.02] | 5.88 |
| Europe & Central Asia | Spain | 1550 | 0.05 | [-0.04, 0.14] | 3.81 |
| Europe & Central Asia | Sweden | 8834 | 0.04 | [0.01, 0.08] | 9.18 |
| Europe & Central Asia | Türkiye | 253 | 0.41 | [0.11, 0.71] | 0.50 |
| Europe & Central Asia | United Kingdom | 2488 | 0.07 | [0.00, 0.13] | 5.67 |
| Latin America & Caribbean | Argentina | 1545 | 0.09 | [0.00, 0.17] | 4.23 |
| Latin America & Caribbean | Brazil | 2219 | 0.12 | [0.04, 0.21] | 4.08 |
| Latin America & Caribbean | Mexico | 1102 | 0.10 | [0.00, 0.20] | 3.47 |
| Middle East, North Africa, Afghanistan & Pakistan | Egypt | 2211 | 0.22 | [-0.01, 0.44] | 0.88 |
| North America | United States | 22767 | -0.00 | [-0.03, 0.02] | 9.99 |
| South Asia | India | 3863 | 0.07 | [-0.04, 0.18] | 2.90 |
| Sub-Saharan Africa | Kenya | 6741 | 0.03 | [-0.04, 0.10] | 5.49 |
| Sub-Saharan Africa | Nigeria | 2499 | -0.03 | [-0.13, 0.08] | 2.98 |
| Sub-Saharan Africa | South Africa | 431 | -0.05 | [-0.36, 0.25] | 0.49 |
| Sub-Saharan Africa | Tanzania | 4212 | 0.09 | [0.02, 0.17] | 4.67 |
| | **Overall** | | 0.05 | [0.02, 0.07] | |

$\tau^2 = 0.00$, $I^2 = 49.8\%$, $H^2 = 1.99$

#### g) Understanding purpose in life

| Region | Country | N | Coef | 95% CI | Weight (%) |
|---|---|---|---|---|---|
| East Asia & Pacific | Australia | 2008 | -0.04 | [ -0.14, 0.05] | 4.20 |
| East Asia & Pacific | China | 3670 | 0.07 | [ 0.01, 0.12] | 5.98 |
| East Asia & Pacific | Hong Kong | 309 | 0.12 | [ -0.08, 0.32] | 1.73 |
| East Asia & Pacific | Indonesia | 1487 | 0.05 | [ -0.06, 0.17] | 3.56 |
| East Asia & Pacific | Japan | 8262 | 0.11 | [ 0.07, 0.15] | 6.86 |
| East Asia & Pacific | Philippines | 2060 | -0.00 | [ -0.08, 0.07] | 5.18 |
| Europe & Central Asia | Germany | 2931 | 0.00 | [ -0.06, 0.06] | 5.89 |
| Europe & Central Asia | Poland | 3187 | -0.09 | [ -0.17, -0.02] | 5.22 |
| Europe & Central Asia | Spain | 1543 | 0.06 | [ -0.04, 0.15] | 4.43 |
| Europe & Central Asia | Sweden | 8766 | 0.03 | [ -0.01, 0.07] | 6.67 |
| Europe & Central Asia | Türkiye | 252 | 0.33 | [ 0.05, 0.61] | 1.00 |
| Europe & Central Asia | United Kingdom | 2478 | 0.01 | [ -0.07, 0.08] | 5.24 |
| Latin America & Caribbean | Argentina | 1540 | 0.13 | [ 0.03, 0.23] | 4.06 |
| Latin America & Caribbean | Brazil | 2216 | 0.07 | [ -0.01, 0.15] | 4.95 |
| Latin America & Caribbean | Mexico | 1099 | 0.14 | [ 0.06, 0.21] | 5.24 |
| Middle East, North Africa, Afghanistan & Pakistan | Egypt | 2193 | 0.18 | [ 0.02, 0.34] | 2.42 |
| North America | United States | 22735 | 0.04 | [ 0.00, 0.07] | 7.03 |
| South Asia | India | 3849 | 0.09 | [ -0.01, 0.19] | 4.24 |
| Sub-Saharan Africa | Kenya | 6740 | -0.00 | [ -0.07, 0.06] | 5.49 |
| Sub-Saharan Africa | Nigeria | 2494 | -0.05 | [ -0.15, 0.05] | 4.13 |
| Sub-Saharan Africa | South Africa | 431 | -0.27 | [ -0.51, -0.03] | 1.31 |
| Sub-Saharan Africa | Tanzania | 4176 | 0.12 | [ 0.04, 0.20] | 5.16 |
| | **Overall** | | 0.04 | [ 0.01, 0.07] | |

$\tau^2 = 0.00$, $I^2 = 70.3\%$, $H^2 = 3.37$

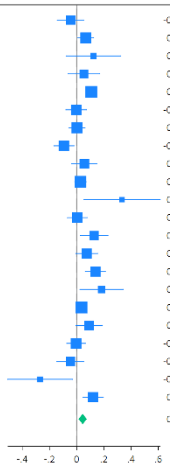

#### h) Life satisfaction

| Region | Country | N | Coef | 95% CI | Weight (%) |
|---|---|---|---|---|---|
| East Asia & Pacific | Australia | 2010 | 0.02 | [ -0.05, 0.10] | 4.96 |
| East Asia & Pacific | China | 3652 | 0.05 | [ -0.01, 0.10] | 6.86 |
| East Asia & Pacific | Hong Kong | 297 | 0.22 | [ 0.02, 0.42] | 1.13 |
| East Asia & Pacific | Indonesia | 1484 | -0.01 | [ -0.12, 0.11] | 2.94 |
| East Asia & Pacific | Japan | 8226 | 0.07 | [ 0.04, 0.11] | 8.67 |
| East Asia & Pacific | Philippines | 2059 | 0.01 | [ -0.08, 0.10] | 4.20 |
| Europe & Central Asia | Germany | 2913 | 0.08 | [ 0.03, 0.13] | 6.84 |
| Europe & Central Asia | Poland | 3180 | -0.07 | [ -0.13, 0.00] | 5.65 |
| Europe & Central Asia | Spain | 1542 | 0.08 | [ -0.01, 0.16] | 4.33 |
| Europe & Central Asia | Sweden | 8822 | 0.02 | [ -0.01, 0.06] | 8.88 |
| Europe & Central Asia | Türkiye | 253 | 0.27 | [ 0.01, 0.53] | 0.69 |
| Europe & Central Asia | United Kingdom | 2459 | 0.08 | [ 0.02, 0.14] | 6.29 |
| Latin America & Caribbean | Argentina | 1544 | 0.11 | [ 0.01, 0.20] | 3.81 |
| Latin America & Caribbean | Brazil | 2219 | 0.07 | [ -0.02, 0.15] | 4.49 |
| Latin America & Caribbean | Mexico | 1102 | 0.06 | [ -0.04, 0.16] | 3.36 |
| Middle East, North Africa, Afghanistan & Pakistan | Egypt | 2199 | 0.16 | [ 0.00, 0.31] | 1.79 |
| North America | United States | 22756 | -0.01 | [ -0.04, 0.02] | 9.69 |
| South Asia | India | 3859 | 0.10 | [ -0.01, 0.22] | 2.91 |
| Sub-Saharan Africa | Kenya | 6744 | 0.02 | [ -0.05, 0.10] | 5.10 |
| Sub-Saharan Africa | Nigeria | 2499 | -0.02 | [ -0.14, 0.10] | 2.73 |
| Sub-Saharan Africa | South Africa | 426 | -0.09 | [ -0.39, 0.20] | 0.54 |
| Sub-Saharan Africa | Tanzania | 4211 | -0.02 | [ -0.10, 0.07] | 4.13 |
| | **Overall** | | 0.04 | [ 0.02, 0.06] | |

$\tau^2 = 0.00$, $I^2 = 50.2\%$, $H^2 = 2.01$

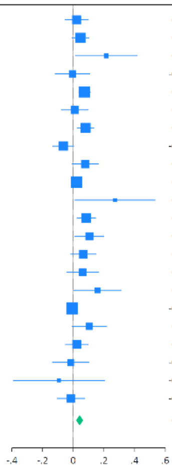

#### i) Life evaluation: Today

| Region | Country | N | Coef | 95% CI | Weight (%) |
|---|---|---|---|---|---|
| East Asia & Pacific | Australia | 2011 | -0.03 | [ -0.10, 0.04] | 5.17 |
| East Asia & Pacific | China | 3671 | 0.03 | [ -0.03, 0.08] | 6.39 |
| East Asia & Pacific | Hong Kong | 309 | 0.13 | [ -0.02, 0.28] | 2.12 |
| East Asia & Pacific | Indonesia | 1485 | 0.09 | [ -0.05, 0.22] | 2.48 |
| East Asia & Pacific | Japan | 8293 | 0.11 | [ 0.07, 0.15] | 7.84 |
| East Asia & Pacific | Philippines | 2056 | 0.12 | [ 0.04, 0.21] | 4.36 |
| Europe & Central Asia | Germany | 2928 | 0.11 | [ 0.05, 0.16] | 6.82 |
| Europe & Central Asia | Poland | 3195 | 0.01 | [ -0.06, 0.08] | 5.73 |
| Europe & Central Asia | Spain | 1550 | 0.05 | [ -0.03, 0.13] | 4.72 |
| Europe & Central Asia | Sweden | 8831 | 0.03 | [ -0.00, 0.06] | 8.23 |
| Europe & Central Asia | Türkiye | 252 | 0.26 | [ -0.04, 0.55] | 0.66 |
| Europe & Central Asia | United Kingdom | 2485 | 0.09 | [ 0.02, 0.15] | 5.96 |
| Latin America & Caribbean | Argentina | 1547 | 0.12 | [ 0.02, 0.23] | 3.68 |
| Latin America & Caribbean | Brazil | 2226 | 0.12 | [ 0.03, 0.20] | 4.45 |
| Latin America & Caribbean | Mexico | 1102 | 0.03 | [ -0.07, 0.13] | 3.85 |
| Middle East, North Africa, Afghanistan & Pakistan | Egypt | 2201 | 0.26 | [ 0.07, 0.45] | 1.45 |
| North America | United States | 22777 | 0.02 | [ -0.01, 0.04] | 8.77 |
| South Asia | India | 3842 | 0.10 | [ -0.01, 0.21] | 3.27 |
| Sub-Saharan Africa | Kenya | 6744 | 0.03 | [ -0.04, 0.10] | 5.56 |
| Sub-Saharan Africa | Nigeria | 2503 | -0.04 | [ -0.16, 0.07] | 3.07 |
| Sub-Saharan Africa | South Africa | 429 | -0.10 | [ -0.38, 0.18] | 0.71 |
| Sub-Saharan Africa | Tanzania | 4215 | 0.14 | [ 0.06, 0.22] | 4.72 |
| | **Overall** | | 0.06 | [ 0.04, 0.09] | |

$\tau^2 = 0.00$, $I^2 = 61.0\%$, $H^2 = 2.56$

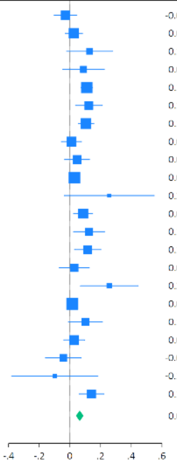

#### j) Life evaluation: Five years from now

| Region | Country | N | Coef | 95% CI | Weight (%) |
|---|---|---|---|---|---|
| East Asia & Pacific | Australia | 2010 | 0.01 | [ -0.07, 0.09] | 4.70 |
| East Asia & Pacific | China | 3653 | 0.04 | [ -0.02, 0.09] | 6.56 |
| East Asia & Pacific | Hong Kong | 300 | 0.18 | [ -0.02, 0.39] | 1.24 |
| East Asia & Pacific | Indonesia | 1467 | 0.09 | [ -0.01, 0.20] | 3.54 |
| East Asia & Pacific | Japan | 8297 | 0.10 | [ 0.06, 0.14] | 7.60 |
| East Asia & Pacific | Philippines | 2057 | 0.04 | [ -0.04, 0.12] | 4.74 |
| Europe & Central Asia | Germany | 2912 | 0.11 | [ 0.06, 0.17] | 6.51 |
| Europe & Central Asia | Poland | 3153 | 0.02 | [ -0.06, 0.10] | 4.83 |
| Europe & Central Asia | Spain | 1540 | 0.05 | [ -0.03, 0.14] | 4.29 |
| Europe & Central Asia | Sweden | 8806 | 0.04 | [ 0.00, 0.07] | 8.00 |
| Europe & Central Asia | Türkiye | 236 | 0.16 | [ -0.12, 0.44] | 0.72 |
| Europe & Central Asia | United Kingdom | 2460 | 0.09 | [ 0.03, 0.16] | 5.36 |
| Latin America & Caribbean | Argentina | 1510 | 0.04 | [ -0.06, 0.14] | 3.64 |
| Latin America & Caribbean | Brazil | 2216 | 0.08 | [ 0.01, 0.16] | 5.11 |
| Latin America & Caribbean | Mexico | 1078 | 0.07 | [ -0.03, 0.16] | 3.93 |
| Middle East, North Africa, Afghanistan & Pakistan | Egypt | 1632 | 0.25 | [ 0.05, 0.44] | 1.35 |
| North America | United States | 22765 | -0.03 | [ -0.06, -0.00] | 8.21 |
| South Asia | India | 3576 | 0.10 | [ 0.01, 0.20] | 3.80 |
| Sub-Saharan Africa | Kenya | 6699 | 0.03 | [ -0.03, 0.10] | 5.73 |
| Sub-Saharan Africa | Nigeria | 2470 | -0.06 | [ -0.15, 0.03] | 4.13 |
| Sub-Saharan Africa | South Africa | 423 | -0.12 | [ -0.35, 0.11] | 1.02 |
| Sub-Saharan Africa | Tanzania | 3979 | 0.11 | [ 0.04, 0.19] | 5.00 |
| | **Overall** | | 0.05 | [ 0.03, 0.08] | |

$\tau^2 = 0.00$, $I^2 = 60.8\%$, $H^2 = 2.55$

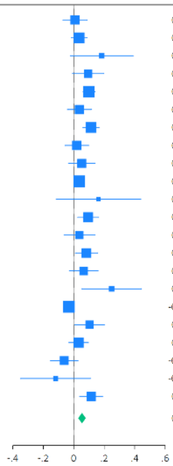

#### k) Feeling lonely

| Region | Country | N | Coef | 95% CI | Weight (%) |
|---|---|---|---|---|---|
| East Asia & Pacific | Australia | 2010 | 0.02 | [ -0.09, 0.12] | 2.78 |
| East Asia & Pacific | China | 3672 | -0.03 | [ -0.10, 0.04] | 6.48 |
| East Asia & Pacific | Hong Kong | 308 | -0.28 | [ -0.55, 0.00] | 0.42 |
| East Asia & Pacific | Indonesia | 1487 | -0.00 | [ -0.19, 0.19] | 0.93 |
| East Asia & Pacific | Japan | 8309 | -0.06 | [ -0.11, -0.01] | 11.61 |
| East Asia & Pacific | Philippines | 2063 | -0.06 | [ -0.17, 0.06] | 2.44 |
| Europe & Central Asia | Germany | 2938 | -0.03 | [ -0.10, 0.05] | 5.96 |
| Europe & Central Asia | Poland | 3190 | 0.03 | [ -0.07, 0.13] | 3.33 |
| Europe & Central Asia | Spain | 1553 | 0.01 | [ -0.12, 0.13] | 2.16 |
| Europe & Central Asia | Sweden | 8848 | -0.04 | [ -0.08, 0.00] | 17.62 |
| Europe & Central Asia | Türkiye | 250 | -0.08 | [ -0.37, 0.21] | 0.37 |
| Europe & Central Asia | United Kingdom | 2476 | -0.02 | [ -0.11, 0.07] | 4.36 |
| Latin America & Caribbean | Argentina | 1549 | -0.11 | [ -0.25, 0.03] | 1.67 |
| Latin America & Caribbean | Brazil | 2221 | -0.03 | [ -0.15, 0.09] | 2.17 |
| Latin America & Caribbean | Mexico | 1099 | -0.09 | [ -0.26, 0.09] | 1.06 |
| Middle East, North Africa, Afghanistan & Pakistan | Egypt | 2210 | -0.12 | [ -0.43, 0.20] | 0.32 |
| North America | United States | 22786 | -0.02 | [ -0.06, 0.01] | 22.84 |
| South Asia | India | 3863 | 0.12 | [ -0.01, 0.25] | 2.00 |
| Sub-Saharan Africa | Kenya | 6739 | -0.03 | [ -0.11, 0.05] | 5.09 |
| Sub-Saharan Africa | Nigeria | 2496 | 0.08 | [ -0.06, 0.21] | 1.79 |
| Sub-Saharan Africa | South Africa | 431 | 0.18 | [ -0.10, 0.46] | 0.42 |
| Sub-Saharan Africa | Tanzania | 4213 | -0.05 | [ -0.14, 0.04] | 4.15 |
| | **Overall** | | -0.03 | [ -0.05, -0.01] | |

$\tau^2 = 0.00$, $I^2 = 0.00\%$, $H^2 = 1.00$

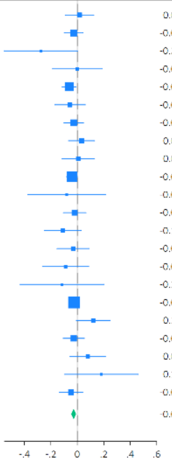

#### l) Life is worthwhile

| Region | Country | N | Coef | 95% CI | Weight (%) |
|---|---|---|---|---|---|
| East Asia & Pacific | Australia | 2008 | 0.01 | [ -0.08, 0.10] | 4.39 |
| East Asia & Pacific | China | 3669 | 0.03 | [ -0.02, 0.08] | 7.08 |
| East Asia & Pacific | Hong Kong | 306 | 0.16 | [ -0.01, 0.34] | 1.58 |
| East Asia & Pacific | Indonesia | 1486 | 0.05 | [ -0.05, 0.15] | 3.62 |
| East Asia & Pacific | Japan | 8290 | 0.09 | [ 0.05, 0.13] | 8.29 |
| East Asia & Pacific | Philippines | 2060 | 0.00 | [ -0.08, 0.09] | 4.39 |
| Europe & Central Asia | Germany | 2932 | 0.07 | [ 0.01, 0.12] | 6.76 |
| Europe & Central Asia | Poland | 3189 | -0.12 | [ -0.19, -0.05] | 5.49 |
| Europe & Central Asia | Spain | 1541 | 0.04 | [ -0.06, 0.14] | 3.69 |
| Europe & Central Asia | Sweden | 8832 | 0.05 | [ 0.01, 0.09] | 8.23 |
| Europe & Central Asia | Türkiye | 253 | 0.26 | [ -0.01, 0.52] | 0.73 |
| Europe & Central Asia | United Kingdom | 2473 | 0.05 | [ -0.01, 0.11] | 6.23 |
| Latin America & Caribbean | Argentina | 1541 | 0.11 | [ 0.02, 0.21] | 3.94 |
| Latin America & Caribbean | Brazil | 2224 | 0.10 | [ 0.02, 0.19] | 4.50 |
| Latin America & Caribbean | Mexico | 1101 | 0.06 | [ -0.04, 0.17] | 3.43 |
| Middle East, North Africa, Afghanistan & Pakistan | Egypt | 2207 | 0.09 | [ -0.16, 0.33] | 0.87 |
| North America | United States | 22770 | 0.02 | [ -0.01, 0.06] | 8.96 |
| South Asia | India | 3844 | 0.02 | [ -0.08, 0.12] | 3.60 |
| Sub-Saharan Africa | Kenya | 6743 | 0.03 | [ -0.04, 0.10] | 5.45 |
| Sub-Saharan Africa | Nigeria | 2498 | 0.04 | [ -0.07, 0.15] | 3.23 |
| Sub-Saharan Africa | South Africa | 428 | 0.02 | [ -0.24, 0.29] | 0.76 |
| Sub-Saharan Africa | Tanzania | 4209 | 0.11 | [ 0.03, 0.19] | 4.78 |
| | **Overall** | | 0.05 | [ 0.02, 0.07] | |

$\tau^2 = 0.00$, $I^2 = 53.5\%$, $H^2 = 2.15$

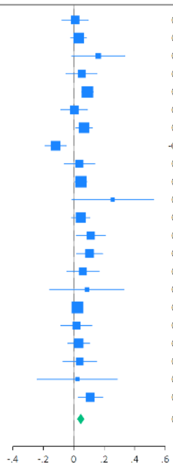

m) Feeling grateful

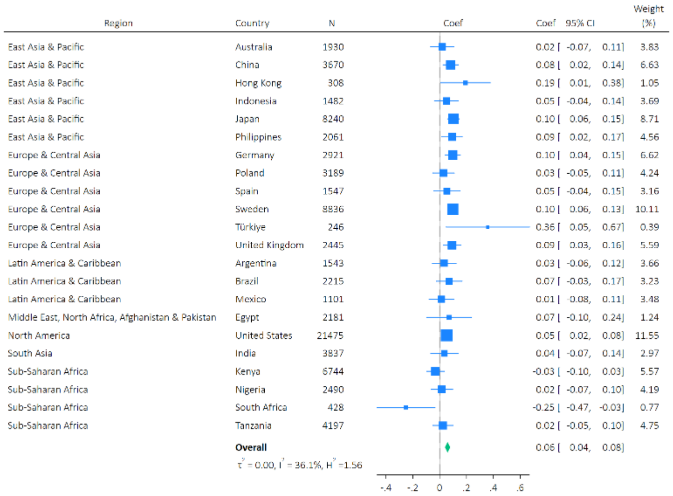

n) Feeling at peace

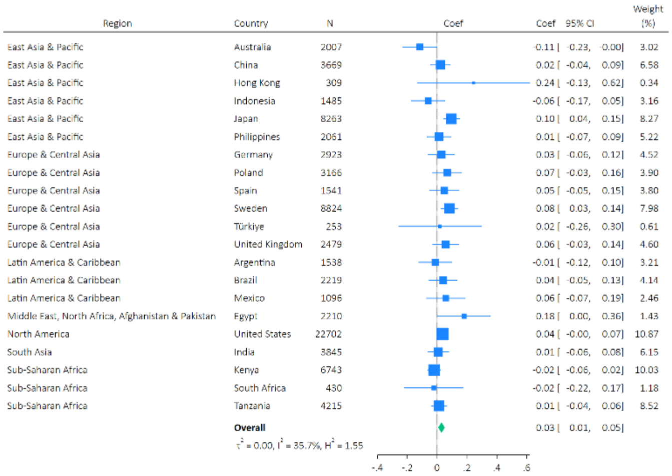

o) Remain hopeful about the future

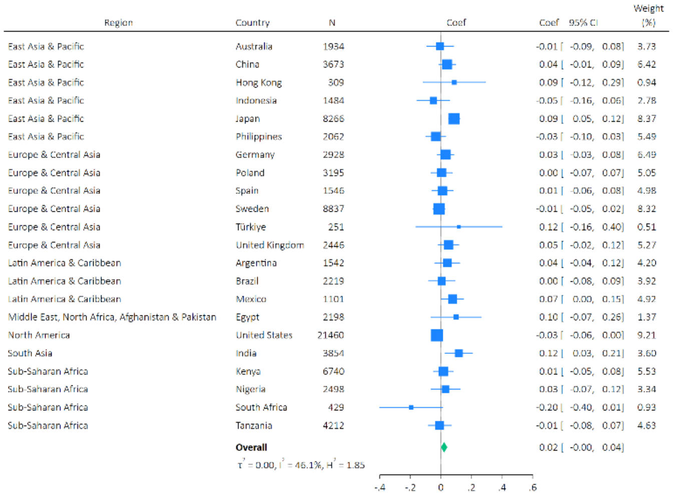

p) Feeling very capable in most things

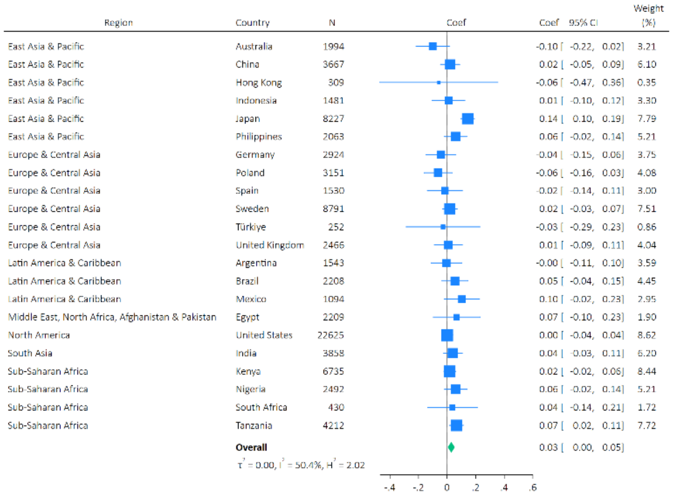

q) Expect more good things to happen to you

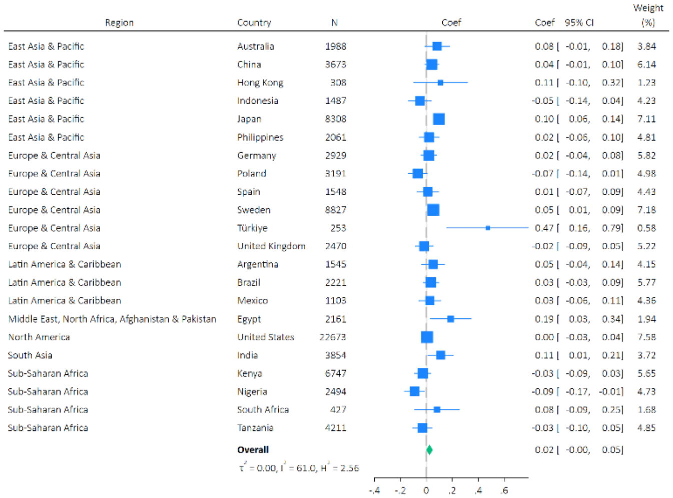

r) Overall mental health

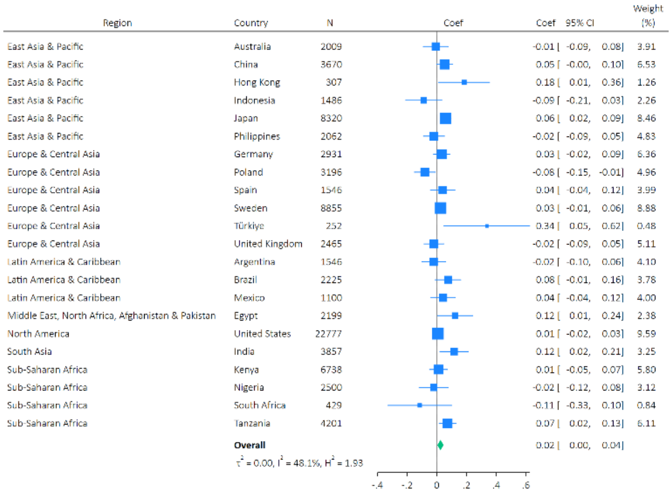

s) Overall physical health

| Region | Country | N | Coef | Coef | 95% CI | Weight (%) |
|---|---|---|---|---|---|---|
| East Asia & Pacific | Australia | 2010 | | -0.05 | [ -0.13, 0.02] | 4.90 |
| East Asia & Pacific | China | 3673 | | 0.06 | [ 0.00, 0.11] | 6.19 |
| East Asia & Pacific | Hong Kong | 309 | | 0.11 | [ -0.10, 0.32] | 1.37 |
| East Asia & Pacific | Indonesia | 1487 | | 0.05 | [ -0.07, 0.18] | 2.94 |
| East Asia & Pacific | Japan | 8289 | | 0.10 | [ 0.06, 0.13] | 7.05 |
| East Asia & Pacific | Philippines | 2062 | | 0.01 | [ -0.07, 0.09] | 4.94 |
| Europe & Central Asia | Germany | 2940 | | 0.09 | [ 0.04, 0.13] | 6.48 |
| Europe & Central Asia | Poland | 3195 | | -0.09 | [ -0.15, -0.02] | 5.54 |
| Europe & Central Asia | Spain | 1549 | | 0.01 | [ -0.07, 0.10] | 4.30 |
| Europe & Central Asia | Sweden | 8860 | | 0.02 | [ -0.02, 0.05] | 7.21 |
| Europe & Central Asia | Türkiye | 253 | | 0.30 | [ 0.07, 0.53] | 1.17 |
| Europe & Central Asia | United Kingdom | 2494 | | 0.02 | [ -0.04, 0.09] | 5.55 |
| Latin America & Caribbean | Argentina | 1547 | | 0.04 | [ -0.04, 0.13] | 4.60 |
| Latin America & Caribbean | Brazil | 2220 | | 0.10 | [ 0.01, 0.18] | 4.51 |
| Latin America & Caribbean | Mexico | 1103 | | 0.06 | [ -0.02, 0.13] | 4.80 |
| Middle East, North Africa, Afghanistan & Pakistan | Egypt | 2208 | | 0.19 | [ 0.00, 0.38] | 1.64 |
| North America | United States | 22775 | | 0.04 | [ 0.02, 0.07] | 7.68 |
| South Asia | India | 3858 | | 0.08 | [ -0.02, 0.18] | 3.86 |
| Sub-Saharan Africa | Kenya | 6743 | | 0.02 | [ -0.04, 0.08] | 5.62 |
| Sub-Saharan Africa | Nigeria | 2502 | | -0.09 | [ -0.20, 0.01] | 3.58 |
| Sub-Saharan Africa | South Africa | 431 | | -0.20 | [ -0.44, 0.04] | 1.08 |
| Sub-Saharan Africa | Tanzania | 4215 | | 0.13 | [ 0.05, 0.20] | 4.99 |
| | **Overall** | | | 0.04 | [ 0.01, 0.07] | |

$\tau^2 = 0.01$, $I^2 = 70.0\%$, $H^2 = 3.30$

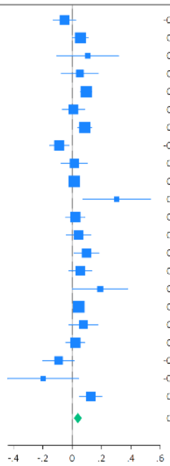

Supplementary Figure S7: Meta-analysis forest plots for the effects of arts engagement on later health and wellbeing outcomes (the main model; fully adjusted)

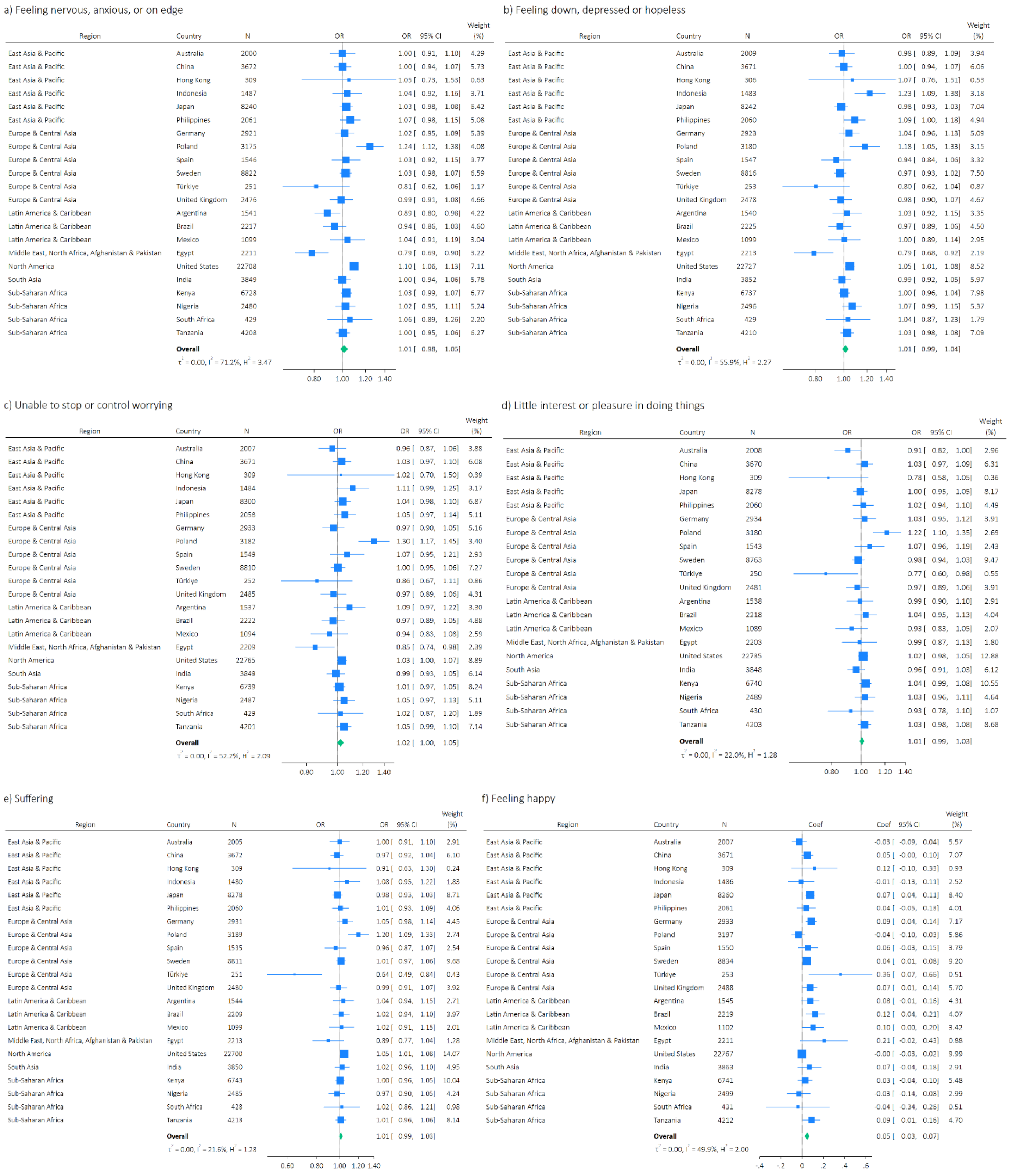

g) Understanding purpose in life

| Region | Country | N | Coef | 95% CI | Weight (%) |
|---|---|---|---|---|---|
| East Asia & Pacific | Australia | 2008 | -0.05 | [-0.15, 0.05] | 4.13 |
| East Asia & Pacific | China | 3670 | 0.07 | [0.01, 0.13] | 6.14 |
| East Asia & Pacific | Hong Kong | 309 | 0.14 | [-0.07, 0.34] | 1.55 |
| East Asia & Pacific | Indonesia | 1487 | 0.06 | [-0.06, 0.17] | 3.46 |
| East Asia & Pacific | Japan | 8262 | 0.11 | [0.07, 0.14] | 7.27 |
| East Asia & Pacific | Philippines | 2060 | -0.00 | [-0.08, 0.07] | 5.16 |
| Europe & Central Asia | Germany | 2931 | 0.01 | [-0.05, 0.07] | 6.04 |
| Europe & Central Asia | Poland | 3187 | -0.07 | [-0.15, 0.01] | 5.01 |
| Europe & Central Asia | Spain | 1543 | 0.07 | [-0.03, 0.16] | 4.26 |
| Europe & Central Asia | Sweden | 8766 | 0.03 | [-0.01, 0.07] | 7.03 |
| Europe & Central Asia | Türkiye | 252 | 0.30 | [0.02, 0.57] | 0.90 |
| Europe & Central Asia | United Kingdom | 2478 | 0.01 | [-0.06, 0.09] | 5.25 |
| Latin America & Caribbean | Argentina | 1540 | 0.12 | [0.02, 0.22] | 3.93 |
| Latin America & Caribbean | Brazil | 2216 | 0.07 | [-0.01, 0.16] | 4.89 |
| Latin America & Caribbean | Mexico | 1099 | 0.14 | [0.07, 0.22] | 5.16 |
| Middle East, North Africa, Afghanistan & Pakistan | Egypt | 2193 | 0.18 | [0.02, 0.33] | 2.24 |
| North America | United States | 22735 | 0.03 | [0.00, 0.07] | 7.50 |
| South Asia | India | 3849 | 0.09 | [-0.01, 0.19] | 4.13 |
| Sub-Saharan Africa | Kenya | 6740 | 0.00 | [-0.07, 0.07] | 5.56 |
| Sub-Saharan Africa | Nigeria | 2494 | -0.05 | [-0.15, 0.05] | 4.01 |
| Sub-Saharan Africa | South Africa | 431 | -0.26 | [-0.49, -0.03] | 1.21 |
| Sub-Saharan Africa | Tanzania | 4176 | 0.11 | [0.04, 0.19] | 5.17 |
| | **Overall** | | 0.05 | [0.02, 0.07] | |
| | $\tau^2 = 0.00$, $I^2 = 65.0\%$, $H^2 = 2.85$ | | | | |

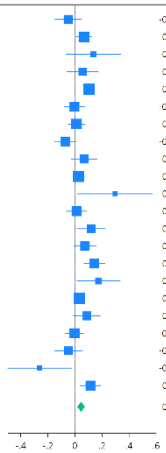

h) Life satisfaction

| Region | Country | N | Coef | 95% CI | Weight (%) |
|---|---|---|---|---|---|
| East Asia & Pacific | Australia | 2010 | 0.02 | [-0.05, 0.09] | 5.09 |
| East Asia & Pacific | China | 3652 | 0.05 | [-0.01, 0.10] | 6.82 |
| East Asia & Pacific | Hong Kong | 297 | 0.21 | [0.01, 0.41] | 1.10 |
| East Asia & Pacific | Indonesia | 1484 | -0.01 | [-0.12, 0.11] | 2.93 |
| East Asia & Pacific | Japan | 8226 | 0.07 | [0.04, 0.11] | 8.64 |
| East Asia & Pacific | Philippines | 2059 | 0.01 | [-0.08, 0.09] | 4.17 |
| Europe & Central Asia | Germany | 2913 | 0.09 | [0.04, 0.14] | 6.80 |
| Europe & Central Asia | Poland | 3180 | -0.06 | [-0.13, 0.01] | 5.63 |
| Europe & Central Asia | Spain | 1542 | 0.08 | [-0.00, 0.17] | 4.19 |
| Europe & Central Asia | Sweden | 8822 | 0.02 | [-0.01, 0.06] | 8.89 |
| Europe & Central Asia | Türkiye | 253 | 0.22 | [-0.02, 0.47] | 0.76 |
| Europe & Central Asia | United Kingdom | 2459 | 0.09 | [0.03, 0.15] | 6.38 |
| Latin America & Caribbean | Argentina | 1544 | 0.09 | [-0.00, 0.18] | 3.90 |
| Latin America & Caribbean | Brazil | 2219 | 0.07 | [-0.02, 0.15] | 4.49 |
| Latin America & Caribbean | Mexico | 1102 | 0.07 | [-0.04, 0.17] | 3.31 |
| Middle East, North Africa, Afghanistan & Pakistan | Egypt | 2199 | 0.15 | [-0.00, 0.31] | 1.75 |
| North America | United States | 22756 | -0.01 | [-0.04, 0.02] | 9.68 |
| South Asia | India | 3859 | 0.10 | [-0.02, 0.21] | 2.94 |
| Sub-Saharan Africa | Kenya | 6744 | 0.02 | [-0.05, 0.10] | 5.08 |
| Sub-Saharan Africa | Nigeria | 2499 | -0.02 | [-0.14, 0.10] | 2.75 |
| Sub-Saharan Africa | South Africa | 426 | -0.10 | [-0.39, 0.19] | 0.55 |
| Sub-Saharan Africa | Tanzania | 4211 | -0.02 | [-0.10, 0.07] | 4.15 |
| | **Overall** | | 0.04 | [0.02, 0.06] | |
| | $\tau^2 = 0.00$, $I^2 = 50.4\%$, $H^2 = 2.02$ | | | | |

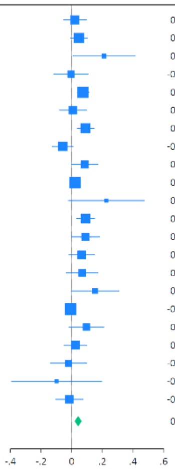

i) Life evaluation: Today

| Region | Country | N | Coef | 95% CI | Weight (%) |
|---|---|---|---|---|---|
| East Asia & Pacific | Australia | 2011 | -0.03 | [-0.10, 0.04] | 5.18 |
| East Asia & Pacific | China | 3671 | 0.03 | [-0.03, 0.09] | 6.39 |
| East Asia & Pacific | Hong Kong | 309 | 0.12 | [-0.03, 0.27] | 2.03 |
| East Asia & Pacific | Indonesia | 1485 | 0.09 | [-0.05, 0.22] | 2.44 |
| East Asia & Pacific | Japan | 8293 | 0.11 | [0.07, 0.14] | 7.89 |
| East Asia & Pacific | Philippines | 2056 | 0.12 | [0.03, 0.21] | 4.34 |
| Europe & Central Asia | Germany | 2928 | 0.12 | [0.07, 0.17] | 6.88 |
| Europe & Central Asia | Poland | 3195 | 0.02 | [-0.05, 0.08] | 5.59 |
| Europe & Central Asia | Spain | 1550 | 0.06 | [-0.02, 0.14] | 4.81 |
| Europe & Central Asia | Sweden | 8831 | 0.03 | [-0.00, 0.06] | 8.33 |
| Europe & Central Asia | Türkiye | 252 | 0.25 | [-0.03, 0.52] | 0.71 |
| Europe & Central Asia | United Kingdom | 2485 | 0.09 | [0.03, 0.15] | 6.01 |
| Latin America & Caribbean | Argentina | 1547 | 0.11 | [0.01, 0.21] | 3.65 |
| Latin America & Caribbean | Brazil | 2226 | 0.12 | [0.03, 0.20] | 4.40 |
| Latin America & Caribbean | Mexico | 1102 | 0.03 | [-0.06, 0.13] | 3.73 |
| Middle East, North Africa, Afghanistan & Pakistan | Egypt | 2201 | 0.25 | [0.06, 0.44] | 1.43 |
| North America | United States | 22777 | 0.02 | [-0.01, 0.04] | 8.88 |
| South Asia | India | 3842 | 0.10 | [-0.01, 0.21] | 3.25 |
| Sub-Saharan Africa | Kenya | 6744 | 0.03 | [-0.04, 0.10] | 5.56 |
| Sub-Saharan Africa | Nigeria | 2503 | -0.05 | [-0.16, 0.07] | 3.03 |
| Sub-Saharan Africa | South Africa | 429 | -0.09 | [-0.35, 0.18] | 0.77 |
| Sub-Saharan Africa | Tanzania | 4215 | 0.13 | [0.05, 0.21] | 4.70 |
| | **Overall** | | 0.06 | [0.04, 0.09] | |
| | $\tau^2 = 0.00$, $I^2 = 60.4\%$, $H^2 = 2.52$ | | | | |

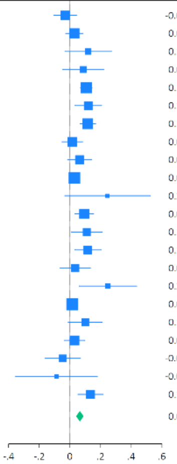

j) Life evaluation: Five years from now

| Region | Country | N | Coef | 95% CI | Weight (%) |
|---|---|---|---|---|---|
| East Asia & Pacific | Australia | 2010 | 0.01 | [-0.07, 0.09] | 4.71 |
| East Asia & Pacific | China | 3653 | 0.04 | [-0.02, 0.09] | 6.48 |
| East Asia & Pacific | Hong Kong | 300 | 0.19 | [-0.01, 0.39] | 1.33 |
| East Asia & Pacific | Indonesia | 1467 | 0.10 | [-0.00, 0.20] | 3.63 |
| East Asia & Pacific | Japan | 8297 | 0.10 | [0.06, 0.14] | 7.50 |
| East Asia & Pacific | Philippines | 2057 | 0.04 | [-0.04, 0.12] | 4.73 |
| Europe & Central Asia | Germany | 2912 | 0.12 | [0.07, 0.17] | 6.40 |
| Europe & Central Asia | Poland | 3153 | 0.02 | [-0.06, 0.10] | 4.67 |
| Europe & Central Asia | Spain | 1540 | 0.08 | [-0.01, 0.16] | 4.53 |
| Europe & Central Asia | Sweden | 8806 | 0.04 | [0.00, 0.07] | 7.90 |
| Europe & Central Asia | Türkiye | 236 | 0.13 | [-0.16, 0.42] | 0.69 |
| Europe & Central Asia | United Kingdom | 2460 | 0.10 | [0.03, 0.17] | 5.40 |
| Latin America & Caribbean | Argentina | 1510 | 0.03 | [-0.07, 0.13] | 3.70 |
| Latin America & Caribbean | Brazil | 2216 | 0.08 | [0.01, 0.16] | 5.11 |
| Latin America & Caribbean | Mexico | 1078 | 0.08 | [-0.02, 0.17] | 3.96 |
| Middle East, North Africa, Afghanistan & Pakistan | Egypt | 1632 | 0.24 | [0.04, 0.44] | 1.34 |
| North America | United States | 22765 | -0.03 | [-0.06, -0.00] | 8.10 |
| South Asia | India | 3576 | 0.10 | [0.00, 0.19] | 3.84 |
| Sub-Saharan Africa | Kenya | 6699 | 0.03 | [-0.03, 0.10] | 5.70 |
| Sub-Saharan Africa | Nigeria | 2470 | -0.06 | [-0.15, 0.03] | 4.13 |
| Sub-Saharan Africa | South Africa | 423 | -0.11 | [-0.33, 0.11] | 1.12 |
| Sub-Saharan Africa | Tanzania | 3979 | 0.11 | [0.03, 0.18] | 5.02 |
| | **Overall** | | 0.06 | [0.03, 0.08] | |
| | $\tau^2 = 0.00$, $I^2 = 62.0\%$, $H^2 = 2.63$ | | | | |

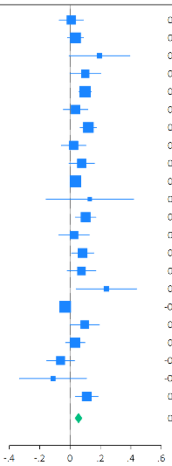

k) Feeling lonely

| Region | Country | N | Coef | 95% CI | Weight (%) |
|---|---|---|---|---|---|
| East Asia & Pacific | Australia | 2010 | 0.01 | [-0.10, 0.12] | 2.78 |
| East Asia & Pacific | China | 3672 | -0.03 | [-0.10, 0.04] | 6.39 |
| East Asia & Pacific | Hong Kong | 308 | -0.30 | [-0.58, -0.01] | 0.39 |
| East Asia & Pacific | Indonesia | 1487 | -0.00 | [-0.19, 0.19] | 0.92 |
| East Asia & Pacific | Japan | 8309 | -0.06 | [-0.11, -0.01] | 11.34 |
| East Asia & Pacific | Philippines | 2063 | -0.06 | [-0.17, 0.06] | 2.45 |
| Europe & Central Asia | Germany | 2938 | -0.05 | [-0.12, 0.03] | 6.02 |
| Europe & Central Asia | Poland | 3190 | 0.04 | [-0.05, 0.14] | 3.57 |
| Europe & Central Asia | Spain | 1553 | 0.02 | [-0.10, 0.14] | 2.13 |
| Europe & Central Asia | Sweden | 8848 | -0.04 | [-0.08, 0.00] | 17.67 |
| Europe & Central Asia | Türkiye | 250 | -0.08 | [-0.37, 0.22] | 0.37 |
| Europe & Central Asia | United Kingdom | 2476 | -0.02 | [-0.11, 0.06] | 4.34 |
| Latin America & Caribbean | Argentina | 1549 | -0.09 | [-0.22, 0.05] | 1.70 |
| Latin America & Caribbean | Brazil | 2221 | -0.03 | [-0.16, 0.09] | 2.17 |
| Latin America & Caribbean | Mexico | 1099 | -0.11 | [-0.29, 0.07] | 1.04 |
| Middle East, North Africa, Afghanistan & Pakistan | Egypt | 2210 | -0.10 | [-0.43, 0.22] | 0.31 |
| North America | United States | 22786 | -0.02 | [-0.06, 0.01] | 22.99 |
| South Asia | India | 3863 | 0.12 | [-0.00, 0.25] | 1.99 |
| Sub-Saharan Africa | Kenya | 6739 | -0.03 | [-0.11, 0.05] | 5.07 |
| Sub-Saharan Africa | Nigeria | 2496 | 0.08 | [-0.05, 0.21] | 1.81 |
| Sub-Saharan Africa | South Africa | 431 | 0.19 | [-0.08, 0.47] | 0.43 |
| Sub-Saharan Africa | Tanzania | 4213 | -0.05 | [-0.13, 0.04] | 4.10 |
| | **Overall** | | -0.03 | [-0.05, -0.01] | |
| | $\tau^2 = 0.00$, $I^2 = 0.02\%$, $H^2 = 1.00$ | | | | |

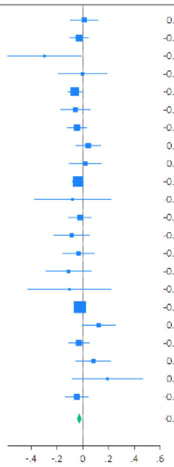

l) Life is worthwhile

| Region | Country | N | Coef | 95% CI | Weight (%) |
|---|---|---|---|---|---|
| East Asia & Pacific | Australia | 2008 | 0.01 | [-0.07, 0.10] | 4.28 |
| East Asia & Pacific | China | 3669 | 0.03 | [-0.02, 0.09] | 7.34 |
| East Asia & Pacific | Hong Kong | 306 | 0.17 | [-0.01, 0.34] | 1.33 |
| East Asia & Pacific | Indonesia | 1486 | 0.06 | [-0.04, 0.16] | 3.37 |
| East Asia & Pacific | Japan | 8290 | 0.09 | [0.05, 0.13] | 8.98 |
| East Asia & Pacific | Philippines | 2060 | -0.00 | [-0.09, 0.09] | 4.17 |
| Europe & Central Asia | Germany | 2932 | 0.07 | [0.02, 0.13] | 6.94 |
| Europe & Central Asia | Poland | 3189 | -0.10 | [-0.17, -0.03] | 5.24 |
| Europe & Central Asia | Spain | 1541 | 0.04 | [-0.06, 0.14] | 3.44 |
| Europe & Central Asia | Sweden | 8832 | 0.05 | [0.01, 0.09] | 8.95 |
| Europe & Central Asia | Türkiye | 253 | 0.19 | [-0.07, 0.46] | 0.63 |
| Europe & Central Asia | United Kingdom | 2473 | 0.05 | [-0.01, 0.11] | 6.38 |
| Latin America & Caribbean | Argentina | 1541 | 0.10 | [0.01, 0.20] | 3.77 |
| Latin America & Caribbean | Brazil | 2224 | 0.10 | [0.02, 0.19] | 4.23 |
| Latin America & Caribbean | Mexico | 1101 | 0.06 | [-0.05, 0.16] | 3.19 |
| Middle East, North Africa, Afghanistan & Pakistan | Egypt | 2207 | 0.08 | [-0.16, 0.32] | 0.75 |
| North America | United States | 22770 | 0.02 | [-0.01, 0.05] | 10.02 |
| South Asia | India | 3844 | 0.02 | [-0.08, 0.12] | 3.36 |
| Sub-Saharan Africa | Kenya | 6743 | 0.03 | [-0.04, 0.10] | 5.38 |
| Sub-Saharan Africa | Nigeria | 2498 | 0.04 | [-0.07, 0.14] | 2.98 |
| Sub-Saharan Africa | South Africa | 428 | 0.01 | [-0.26, 0.27] | 0.62 |
| Sub-Saharan Africa | Tanzania | 4209 | 0.11 | [0.03, 0.18] | 4.64 |
| | **Overall** | | 0.05 | [0.02, 0.07] | |
| | $\tau^2 = 0.00$, $I^2 = 44.9\%$, $H^2 = 1.81$ | | | | |

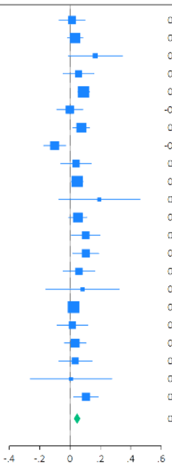

m) Feeling grateful

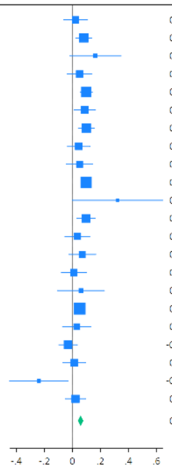

| Region | Country | N | Coef | 95% CI | Weight (%) |
|---|---|---|---|---|---|
| East Asia & Pacific | Australia | 1930 | 0.02 | [ -0.06, 0.11] | 3.86 |
| East Asia & Pacific | China | 3670 | 0.08 | [ 0.02, 0.14] | 6.59 |
| East Asia & Pacific | Hong Kong | 308 | 0.16 | [ -0.02, 0.35] | 1.04 |
| East Asia & Pacific | Indonesia | 1482 | 0.05 | [ -0.04, 0.14] | 3.70 |
| East Asia & Pacific | Japan | 8240 | 0.10 | [ 0.06, 0.14] | 8.72 |
| East Asia & Pacific | Philippines | 2061 | 0.09 | [ 0.01, 0.17] | 4.48 |
| Europe & Central Asia | Germany | 2921 | 0.10 | [ 0.04, 0.16] | 6.59 |
| Europe & Central Asia | Poland | 3189 | 0.05 | [ -0.04, 0.13] | 4.18 |
| Europe & Central Asia | Spain | 1547 | 0.05 | [ -0.04, 0.15] | 3.31 |
| Europe & Central Asia | Sweden | 8836 | 0.10 | [ 0.06, 0.13] | 10.20 |
| Europe & Central Asia | Türkiye | 246 | 0.32 | [ -0.00, 0.65] | 0.35 |
| Europe & Central Asia | United Kingdom | 2445 | 0.10 | [ 0.03, 0.16] | 5.53 |
| Latin America & Caribbean | Argentina | 1543 | 0.04 | [ -0.05, 0.12] | 3.64 |
| Latin America & Caribbean | Brazil | 2215 | 0.07 | [ -0.03, 0.17] | 3.16 |
| Latin America & Caribbean | Mexico | 1101 | 0.01 | [ -0.08, 0.10] | 3.59 |
| Middle East, North Africa, Afghanistan & Pakistan | Egypt | 2181 | 0.06 | [ -0.11, 0.23] | 1.24 |
| North America | United States | 21475 | 0.05 | [ 0.02, 0.08] | 11.67 |
| South Asia | India | 3837 | 0.03 | [ -0.07, 0.13] | 2.94 |
| Sub-Saharan Africa | Kenya | 6744 | -0.03 | [ -0.10, 0.03] | 5.53 |
| Sub-Saharan Africa | Nigeria | 2490 | 0.01 | [ -0.07, 0.10] | 4.15 |
| Sub-Saharan Africa | South Africa | 428 | -0.24 | [ -0.45, -0.03] | 0.82 |
| Sub-Saharan Africa | Tanzania | 4197 | 0.02 | [ -0.05, 0.10] | 4.74 |
| | **Overall** | | 0.06 | [ 0.04, 0.08] | |
| | $\tau^2 = 0.00$, $I^2 = 35.2\%$, $H^2 = 1.54$ | | | | |

n) Feeling at peace

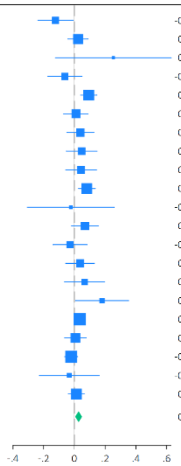

| Region | Country | N | Coef | 95% CI | Weight (%) |
|---|---|---|---|---|---|
| East Asia & Pacific | Australia | 2007 | -0.12 | [ -0.24, -0.01] | 3.04 |
| East Asia & Pacific | China | 3669 | 0.02 | [ -0.04, 0.09] | 6.62 |
| East Asia & Pacific | Hong Kong | 309 | 0.25 | [ -0.12, 0.63] | 0.34 |
| East Asia & Pacific | Indonesia | 1485 | -0.06 | [ -0.17, 0.05] | 3.16 |
| East Asia & Pacific | Japan | 8263 | 0.09 | [ 0.04, 0.15] | 8.24 |
| East Asia & Pacific | Philippines | 2061 | 0.01 | [ -0.07, 0.09] | 5.20 |
| Europe & Central Asia | Germany | 2923 | 0.04 | [ -0.05, 0.13] | 4.55 |
| Europe & Central Asia | Poland | 3166 | 0.05 | [ -0.05, 0.15] | 3.70 |
| Europe & Central Asia | Spain | 1541 | 0.04 | [ -0.06, 0.14] | 3.79 |
| Europe & Central Asia | Sweden | 8824 | 0.08 | [ 0.03, 0.14] | 8.04 |
| Europe & Central Asia | Türkiye | 253 | -0.02 | [ -0.31, 0.26] | 0.59 |
| Europe & Central Asia | United Kingdom | 2479 | 0.07 | [ -0.02, 0.16] | 4.63 |
| Latin America & Caribbean | Argentina | 1538 | -0.03 | [ -0.14, 0.08] | 3.21 |
| Latin America & Caribbean | Brazil | 2219 | 0.04 | [ -0.06, 0.13] | 4.15 |
| Latin America & Caribbean | Mexico | 1096 | 0.07 | [ -0.06, 0.20] | 2.47 |
| Middle East, North Africa, Afghanistan & Pakistan | Egypt | 2210 | 0.18 | [ 0.01, 0.35] | 1.50 |
| North America | United States | 22702 | 0.04 | [ -0.00, 0.07] | 10.87 |
| South Asia | India | 3845 | 0.01 | [ -0.06, 0.08] | 6.19 |
| Sub-Saharan Africa | Kenya | 6743 | -0.02 | [ -0.06, 0.02] | 10.03 |
| Sub-Saharan Africa | South Africa | 430 | -0.03 | [ -0.23, 0.16] | 1.21 |
| Sub-Saharan Africa | Tanzania | 4215 | 0.01 | [ -0.04, 0.06] | 8.47 |
| | **Overall** | | 0.03 | [ 0.01, 0.05] | |
| | $\tau^2 = 0.00$, $I^2 = 36.0\%$, $H^2 = 1.56$ | | | | |

o) Remain hopeful about the future

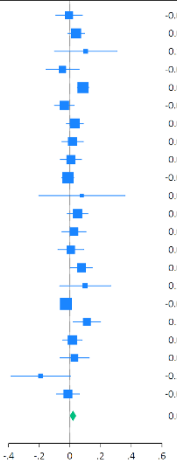

| Region | Country | N | Coef | 95% CI | Weight (%) |
|---|---|---|---|---|---|
| East Asia & Pacific | Australia | 1934 | -0.01 | [ -0.09, 0.08] | 3.80 |
| East Asia & Pacific | China | 3673 | 0.04 | [ -0.01, 0.09] | 6.40 |
| East Asia & Pacific | Hong Kong | 309 | 0.10 | [ -0.10, 0.31] | 0.93 |
| East Asia & Pacific | Indonesia | 1484 | -0.05 | [ -0.16, 0.06] | 2.77 |
| East Asia & Pacific | Japan | 8266 | 0.09 | [ 0.05, 0.12] | 8.35 |
| East Asia & Pacific | Philippines | 2062 | -0.04 | [ -0.10, 0.03] | 5.43 |
| Europe & Central Asia | Germany | 2928 | 0.03 | [ -0.02, 0.09] | 6.39 |
| Europe & Central Asia | Poland | 3195 | 0.02 | [ -0.05, 0.09] | 4.92 |
| Europe & Central Asia | Spain | 1546 | 0.01 | [ -0.06, 0.08] | 5.03 |
| Europe & Central Asia | Sweden | 8837 | -0.01 | [ -0.05, 0.02] | 8.35 |
| Europe & Central Asia | Türkiye | 251 | 0.08 | [ -0.20, 0.36] | 0.51 |
| Europe & Central Asia | United Kingdom | 2446 | 0.05 | [ -0.02, 0.12] | 5.25 |
| Latin America & Caribbean | Argentina | 1542 | 0.03 | [ -0.05, 0.10] | 4.33 |
| Latin America & Caribbean | Brazil | 2219 | 0.01 | [ -0.08, 0.09] | 3.93 |
| Latin America & Caribbean | Mexico | 1101 | 0.07 | [ 0.00, 0.15] | 4.92 |
| Middle East, North Africa, Afghanistan & Pakistan | Egypt | 2198 | 0.10 | [ -0.07, 0.27] | 1.33 |
| North America | United States | 21460 | -0.03 | [ -0.06, 0.00] | 9.25 |
| South Asia | India | 3854 | 0.11 | [ 0.02, 0.20] | 3.62 |
| Sub-Saharan Africa | Kenya | 6740 | 0.02 | [ -0.05, 0.08] | 5.56 |
| Sub-Saharan Africa | Nigeria | 2498 | 0.03 | [ -0.07, 0.13] | 3.28 |
| Sub-Saharan Africa | South Africa | 429 | -0.19 | [ -0.39, 0.00] | 1.02 |
| Sub-Saharan Africa | Tanzania | 4212 | -0.01 | [ -0.09, 0.06] | 4.64 |
| | **Overall** | | 0.02 | [ -0.00, 0.04] | |
| | $\tau^2 = 0.00$, $I^2 = 45.9\%$, $H^2 = 1.85$ | | | | |

p) Feeling very capable in most things

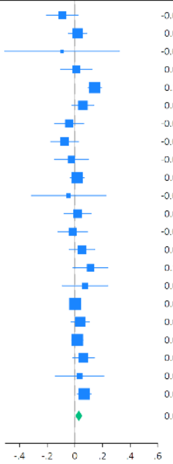

| Region | Country | N | Coef | 95% CI | Weight (%) |
|---|---|---|---|---|---|
| East Asia & Pacific | Australia | 1994 | -0.09 | [ -0.21, 0.02] | 3.28 |
| East Asia & Pacific | China | 3667 | 0.02 | [ -0.05, 0.09] | 6.13 |
| East Asia & Pacific | Hong Kong | 309 | -0.09 | [ -0.51, 0.32] | 0.35 |
| East Asia & Pacific | Indonesia | 1481 | 0.01 | [ -0.10, 0.12] | 3.34 |
| East Asia & Pacific | Japan | 8227 | 0.14 | [ 0.10, 0.19] | 7.74 |
| East Asia & Pacific | Philippines | 2063 | 0.06 | [ -0.02, 0.14] | 5.24 |
| Europe & Central Asia | Germany | 2924 | -0.04 | [ -0.15, 0.06] | 3.76 |
| Europe & Central Asia | Poland | 3151 | -0.07 | [ -0.18, 0.03] | 3.93 |
| Europe & Central Asia | Spain | 1530 | -0.03 | [ -0.15, 0.10] | 3.01 |
| Europe & Central Asia | Sweden | 8791 | 0.02 | [ -0.03, 0.07] | 7.49 |
| Europe & Central Asia | Türkiye | 252 | -0.05 | [ -0.31, 0.22] | 0.81 |
| Europe & Central Asia | United Kingdom | 2466 | 0.02 | [ -0.08, 0.12] | 4.12 |
| Latin America & Caribbean | Argentina | 1543 | -0.02 | [ -0.12, 0.09] | 3.68 |
| Latin America & Caribbean | Brazil | 2208 | 0.05 | [ -0.04, 0.14] | 4.44 |
| Latin America & Caribbean | Mexico | 1094 | 0.11 | [ -0.01, 0.24] | 2.91 |
| Middle East, North Africa, Afghanistan & Pakistan | Egypt | 2209 | 0.07 | [ -0.09, 0.24] | 1.91 |
| North America | United States | 22625 | 0.00 | [ -0.04, 0.04] | 8.59 |
| South Asia | India | 3858 | 0.04 | [ -0.03, 0.10] | 6.19 |
| Sub-Saharan Africa | Kenya | 6735 | 0.02 | [ -0.02, 0.06] | 8.41 |
| Sub-Saharan Africa | Nigeria | 2492 | 0.06 | [ -0.02, 0.14] | 5.26 |
| Sub-Saharan Africa | South Africa | 430 | 0.03 | [ -0.14, 0.21] | 1.71 |
| Sub-Saharan Africa | Tanzania | 4212 | 0.07 | [ 0.02, 0.12] | 7.69 |
| | **Overall** | | 0.03 | [ 0.00, 0.05] | |
| | $\tau^2 = 0.00$, $I^2 = 51.0\%$, $H^2 = 2.04$ | | | | |

q) Expect more good things to happen to you

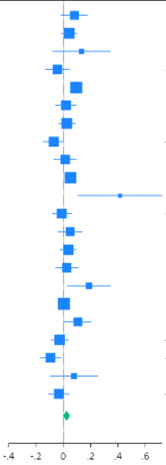

| Region | Country | N | Coef | 95% CI | Weight (%) |
|---|---|---|---|---|---|
| East Asia & Pacific | Australia | 1988 | 0.08 | [ -0.01, 0.18] | 3.79 |
| East Asia & Pacific | China | 3673 | 0.04 | [ -0.01, 0.10] | 6.13 |
| East Asia & Pacific | Hong Kong | 308 | 0.13 | [ -0.08, 0.34] | 1.18 |
| East Asia & Pacific | Indonesia | 1487 | -0.04 | [ -0.13, 0.04] | 4.18 |
| East Asia & Pacific | Japan | 8308 | 0.09 | [ 0.05, 0.14] | 7.11 |
| East Asia & Pacific | Philippines | 2061 | 0.02 | [ -0.06, 0.09] | 4.80 |
| Europe & Central Asia | Germany | 2929 | 0.03 | [ -0.03, 0.09] | 5.80 |
| Europe & Central Asia | Poland | 3191 | -0.07 | [ -0.14, 0.00] | 4.95 |
| Europe & Central Asia | Spain | 1548 | 0.01 | [ -0.07, 0.10] | 4.44 |
| Europe & Central Asia | Sweden | 8827 | 0.05 | [ 0.01, 0.09] | 7.20 |
| Europe & Central Asia | Türkiye | 253 | 0.42 | [ 0.11, 0.72] | 0.60 |
| Europe & Central Asia | United Kingdom | 2470 | -0.01 | [ -0.08, 0.06] | 5.19 |
| Latin America & Caribbean | Argentina | 1545 | 0.05 | [ -0.04, 0.14] | 4.12 |
| Latin America & Caribbean | Brazil | 2221 | 0.04 | [ -0.02, 0.10] | 5.81 |
| Latin America & Caribbean | Mexico | 1103 | 0.03 | [ -0.06, 0.11] | 4.46 |
| Middle East, North Africa, Afghanistan & Pakistan | Egypt | 2161 | 0.19 | [ 0.03, 0.35] | 1.91 |
| North America | United States | 22673 | 0.00 | [ -0.03, 0.04] | 7.62 |
| South Asia | India | 3854 | 0.11 | [ 0.01, 0.20] | 3.71 |
| Sub-Saharan Africa | Kenya | 6747 | -0.03 | [ -0.09, 0.03] | 5.68 |
| Sub-Saharan Africa | Nigeria | 2494 | -0.09 | [ -0.17, -0.02] | 4.83 |
| Sub-Saharan Africa | South Africa | 427 | 0.08 | [ -0.10, 0.25] | 1.63 |
| Sub-Saharan Africa | Tanzania | 4211 | -0.03 | [ -0.11, 0.04] | 4.87 |
| | **Overall** | | 0.02 | [ -0.00, 0.05] | |
| | $\tau^2 = 0.00$, $I^2 = 60.8\%$, $H^2 = 2.55$ | | | | |

r) Overall mental health

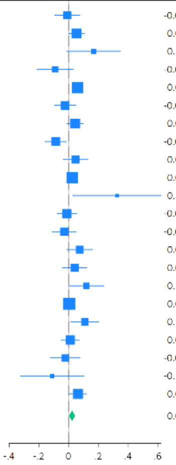

| Region | Country | N | Coef | 95% CI | Weight (%) |
|---|---|---|---|---|---|
| East Asia & Pacific | Australia | 2009 | -0.01 | [ -0.09, 0.08] | 3.92 |
| East Asia & Pacific | China | 3670 | 0.05 | [ 0.00, 0.11] | 6.50 |
| East Asia & Pacific | Hong Kong | 307 | 0.17 | [ -0.01, 0.35] | 1.14 |
| East Asia & Pacific | Indonesia | 1486 | -0.09 | [ -0.21, 0.03] | 2.25 |
| East Asia & Pacific | Japan | 8320 | 0.06 | [ 0.02, 0.10] | 8.43 |
| East Asia & Pacific | Philippines | 2062 | -0.02 | [ -0.09, 0.05] | 4.78 |
| Europe & Central Asia | Germany | 2931 | 0.05 | [ -0.01, 0.10] | 6.32 |
| Europe & Central Asia | Poland | 3196 | -0.09 | [ -0.16, -0.02] | 4.86 |
| Europe & Central Asia | Spain | 1546 | 0.05 | [ -0.04, 0.13] | 3.97 |
| Europe & Central Asia | Sweden | 8855 | 0.02 | [ -0.01, 0.06] | 8.91 |
| Europe & Central Asia | Türkiye | 252 | 0.33 | [ 0.03, 0.62] | 0.45 |
| Europe & Central Asia | United Kingdom | 2465 | -0.01 | [ -0.08, 0.05] | 5.30 |
| Latin America & Caribbean | Argentina | 1546 | -0.03 | [ -0.11, 0.05] | 4.11 |
| Latin America & Caribbean | Brazil | 2225 | 0.08 | [ -0.01, 0.16] | 3.79 |
| Latin America & Caribbean | Mexico | 1100 | 0.04 | [ -0.04, 0.12] | 4.08 |
| Middle East, North Africa, Afghanistan & Pakistan | Egypt | 2199 | 0.12 | [ 0.00, 0.24] | 2.33 |
| North America | United States | 22777 | 0.01 | [ -0.02, 0.03] | 9.62 |
| South Asia | India | 3857 | 0.11 | [ 0.02, 0.20] | 3.33 |
| Sub-Saharan Africa | Kenya | 6738 | 0.01 | [ -0.05, 0.07] | 5.78 |
| Sub-Saharan Africa | Nigeria | 2500 | -0.02 | [ -0.12, 0.08] | 3.13 |
| Sub-Saharan Africa | South Africa | 429 | -0.11 | [ -0.32, 0.10] | 0.84 |
| Sub-Saharan Africa | Tanzania | 4201 | 0.06 | [ 0.01, 0.12] | 6.18 |
| | **Overall** | | 0.02 | [ 0.00, 0.04] | |
| | $\tau^2 = 0.00$, $I^2 = 48.1\%$, $H^2 = 1.93$ | | | | |

s) Overall physical health

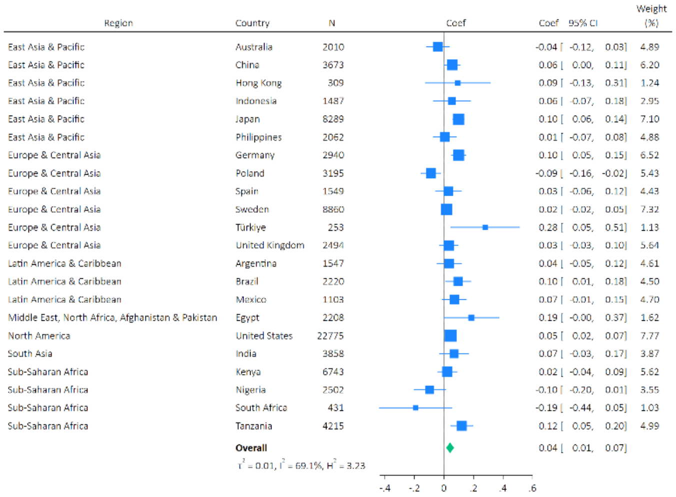

Supplementary Figure S8: Sensitivity analysis exploring the association between arts engagement and the composite measure of psychological distress (PHQ-4).

#### PHQ-4 (a composite measure of psychological distress)

| Region | Country | N | Coef | 95% CI | Weight (%) |
|---|---|---|---|---|---|
| East Asia & Pacific | Australia | 1989 | 0.97 | [ 0.86, 1.09] | 4.42 |
| East Asia & Pacific | China | 3665 | 1.01 | [ 0.94, 1.09] | 6.29 |
| East Asia & Pacific | Hong Kong | 306 | 1.00 | [ 0.76, 1.33] | 1.35 |
| East Asia & Pacific | Indonesia | 1469 | 1.23 | [ 1.04, 1.46] | 2.97 |
| East Asia & Pacific | Japan | 8055 | 1.00 | [ 0.95, 1.06] | 7.17 |
| East Asia & Pacific | Philippines | 2054 | 1.12 | [ 0.99, 1.26] | 4.28 |
| Europe & Central Asia | Germany | 2872 | 1.03 | [ 0.96, 1.11] | 6.26 |
| Europe & Central Asia | Poland | 3119 | 1.22 | [ 1.11, 1.34] | 5.44 |
| Europe & Central Asia | Spain | 1527 | 1.05 | [ 0.91, 1.21] | 3.58 |
| Europe & Central Asia | Sweden | 8564 | 1.01 | [ 0.96, 1.06] | 7.42 |
| Europe & Central Asia | Türkiye | 248 | 0.65 | [ 0.44, 0.96] | 0.75 |
| Europe & Central Asia | United Kingdom | 2431 | 0.95 | [ 0.86, 1.04] | 5.20 |
| Latin America & Caribbean | Argentina | 1514 | 1.02 | [ 0.88, 1.18] | 3.46 |
| Latin America & Caribbean | Brazil | 2198 | 1.00 | [ 0.89, 1.12] | 4.65 |
| Latin America & Caribbean | Mexico | 1077 | 0.94 | [ 0.80, 1.11] | 3.04 |
| Middle East, North Africa, Afghanistan & Pakistan | Egypt | 2196 | 0.73 | [ 0.61, 0.87] | 2.75 |
| North America | United States | 22527 | 1.07 | [ 1.03, 1.12] | 7.71 |
| South Asia | India | 3810 | 0.94 | [ 0.85, 1.05] | 4.82 |
| Sub-Saharan Africa | Kenya | 6692 | 1.04 | [ 0.98, 1.10] | 6.84 |
| Sub-Saharan Africa | Nigeria | 2441 | 1.10 | [ 0.96, 1.26] | 3.80 |
| Sub-Saharan Africa | South Africa | 425 | 1.01 | [ 0.81, 1.26] | 2.01 |
| Sub-Saharan Africa | Tanzania | 4170 | 1.06 | [ 0.97, 1.15] | 5.78 |
| | **Overall** | | 1.02 | [ 0.99, 1.06] | |

$\tau^2 = 0.00$, $I^2 = 64.6\%$, $H^2 = 2.83$

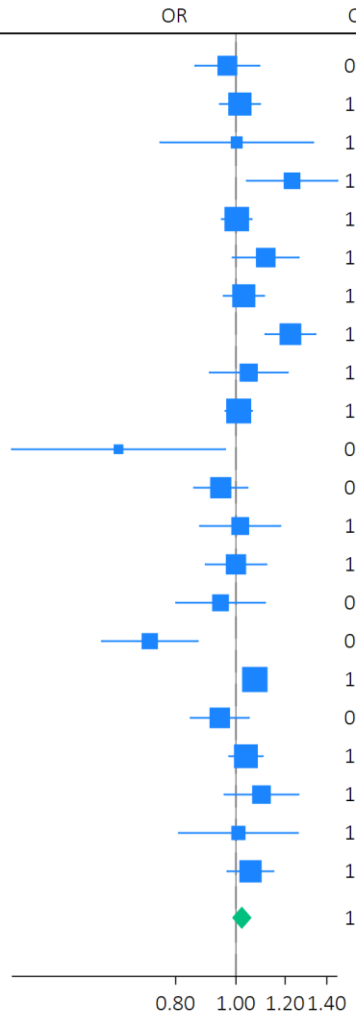

### Supplementary Figure S9: Sensitivity analysis excluding adjustment for baseline health (nested)

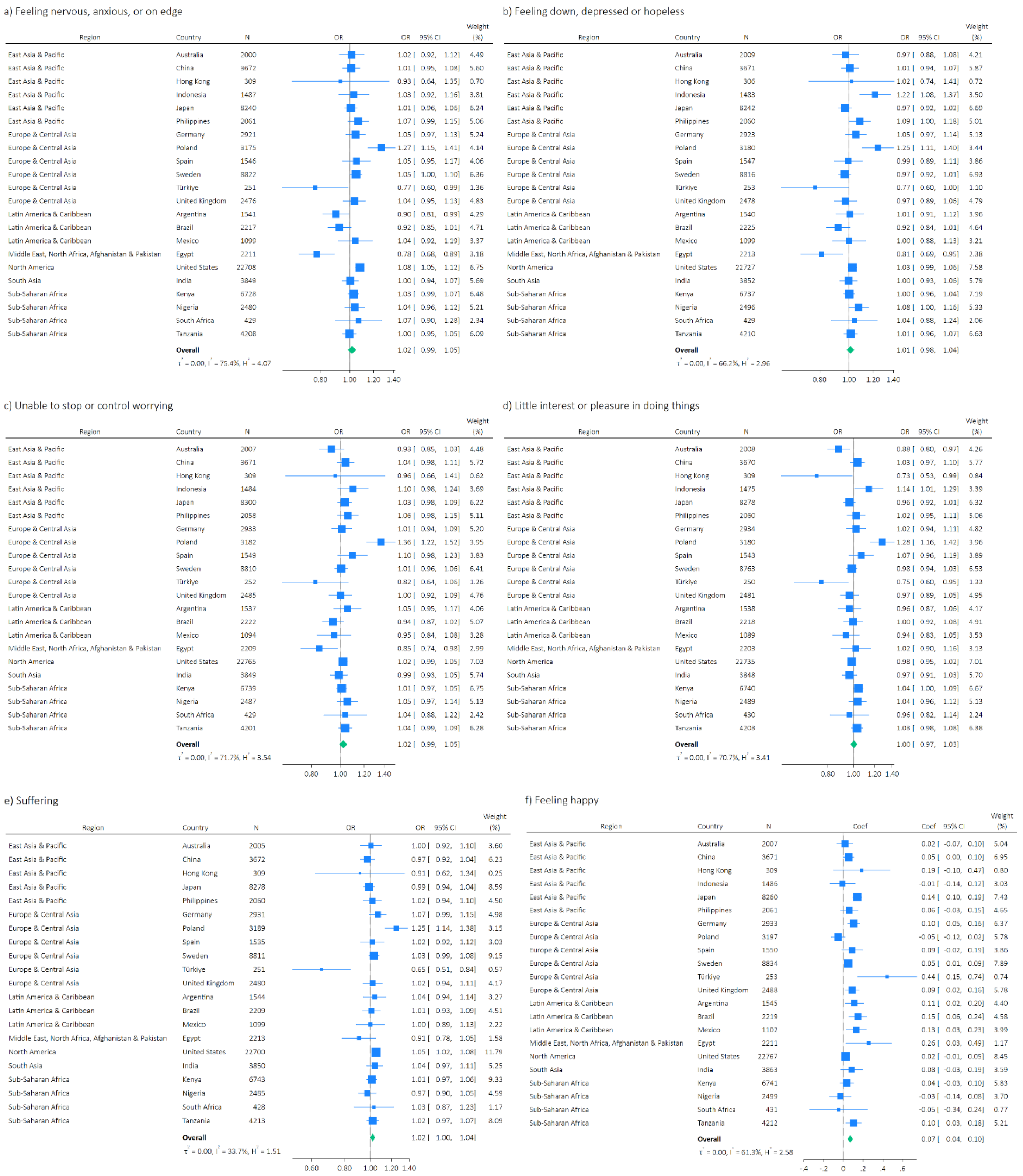

g) Understanding purpose in life

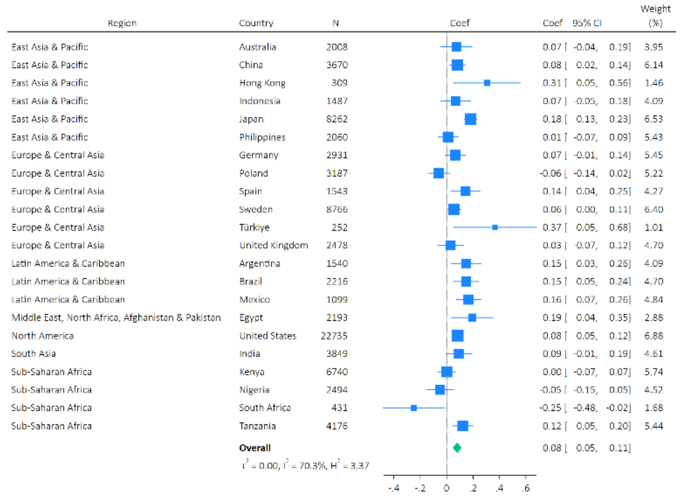

h) Life satisfaction

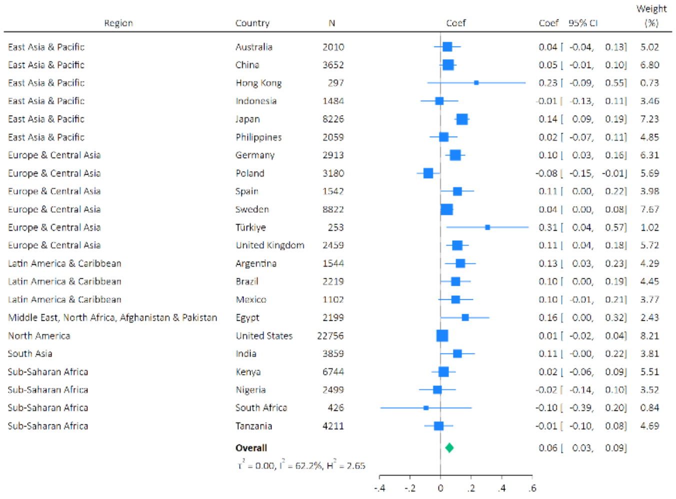

i) Life evaluation: Today

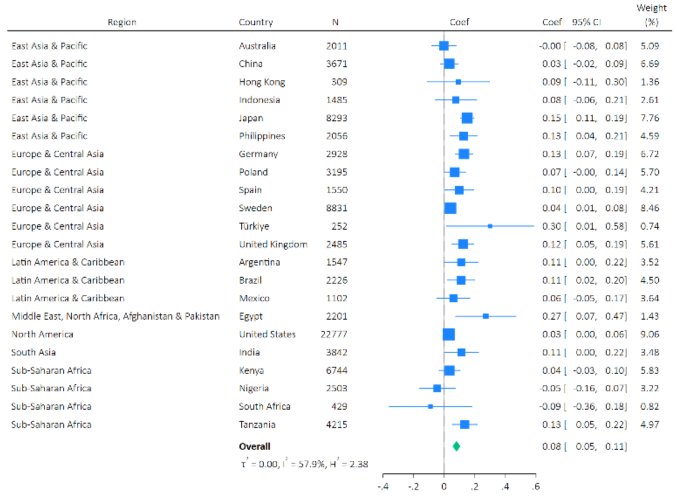

j) Life evaluation: Five years from now

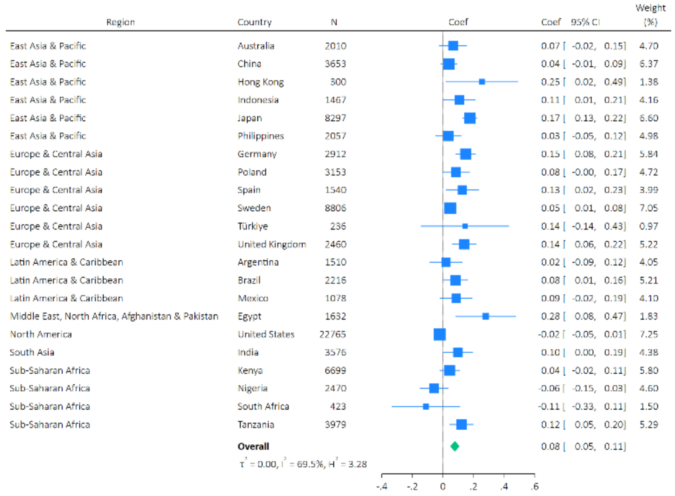

k) Feeling lonely

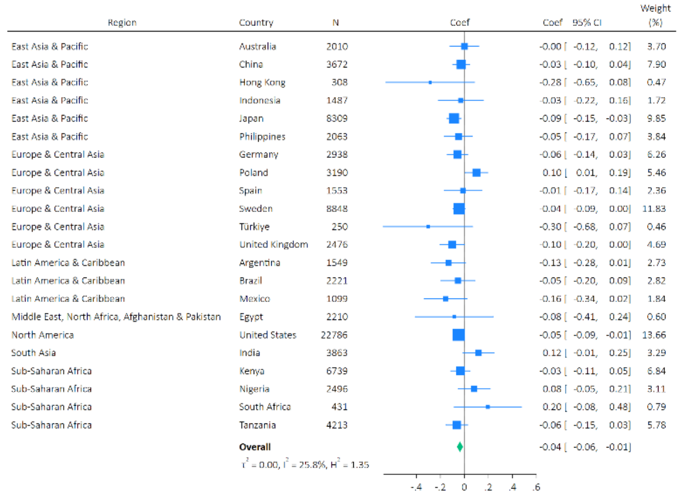

l) Life is worthwhile

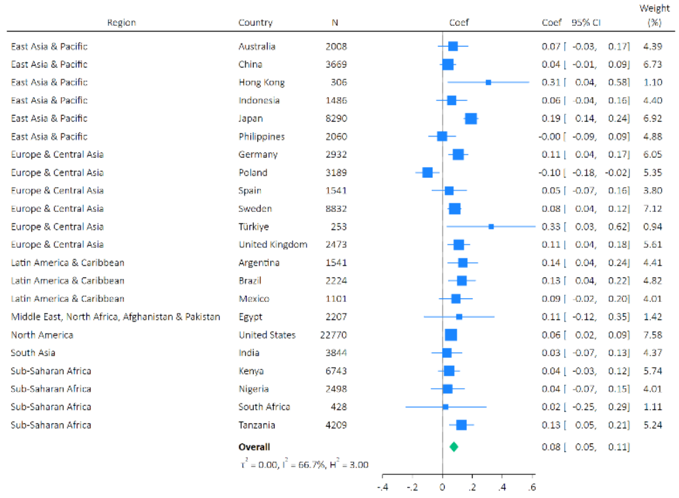

m) Feeling grateful

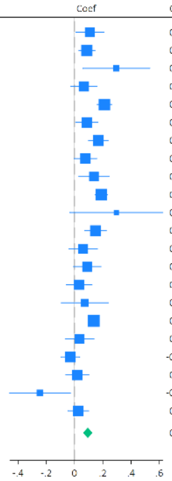

| Region | Country | N | Coef | 95% CI | Weight (%) |
|---|---|---|---|---|---|
| East Asia & Pacific | Australia | 1930 | 0.11 | [ 0.01, 0.21] | 4.42 |
| East Asia & Pacific | China | 3670 | 0.09 | [ 0.03, 0.14] | 5.98 |
| East Asia & Pacific | Hong Kong | 308 | 0.30 | [ 0.06, 0.53] | 1.53 |
| East Asia & Pacific | Indonesia | 1482 | 0.07 | [ -0.03, 0.16] | 4.66 |
| East Asia & Pacific | Japan | 8240 | 0.21 | [ 0.16, 0.26] | 6.19 |
| East Asia & Pacific | Philippines | 2061 | 0.09 | [ 0.01, 0.17] | 5.12 |
| Europe & Central Asia | Germany | 2921 | 0.17 | [ 0.10, 0.24] | 5.52 |
| Europe & Central Asia | Poland | 3189 | 0.08 | [ -0.00, 0.16] | 5.03 |
| Europe & Central Asia | Spain | 1547 | 0.14 | [ 0.03, 0.24] | 4.13 |
| Europe & Central Asia | Sweden | 8836 | 0.19 | [ 0.15, 0.23] | 6.46 |
| Europe & Central Asia | Türkiye | 246 | 0.30 | [ -0.03, 0.63] | 0.88 |
| Europe & Central Asia | United Kingdom | 2445 | 0.15 | [ 0.07, 0.22] | 5.19 |
| Latin America & Caribbean | Argentina | 1543 | 0.06 | [ -0.04, 0.16] | 4.34 |
| Latin America & Caribbean | Brazil | 2215 | 0.09 | [ -0.01, 0.19] | 4.46 |
| Latin America & Caribbean | Mexico | 1101 | 0.03 | [ -0.06, 0.12] | 4.68 |
| Middle East, North Africa, Afghanistan & Pakistan | Egypt | 2181 | 0.07 | [ -0.09, 0.24] | 2.57 |
| North America | United States | 21475 | 0.14 | [ 0.10, 0.17] | 6.74 |
| South Asia | India | 3837 | 0.04 | [ -0.07, 0.14] | 4.31 |
| Sub-Saharan Africa | Kenya | 6744 | -0.03 | [ -0.10, 0.04] | 5.62 |
| Sub-Saharan Africa | Nigeria | 2490 | 0.02 | [ -0.06, 0.10] | 5.04 |
| Sub-Saharan Africa | South Africa | 428 | -0.24 | [ -0.46, -0.03] | 1.79 |
| Sub-Saharan Africa | Tanzania | 4197 | 0.03 | [ -0.05, 0.10] | 5.34 |
| | **Overall** | | 0.09 | [ 0.06, 0.13] | |

$\tau^2 = 0.00$, $I^2 = 73.1\%$, $H^2 = 3.72$

n) Feeling at peace

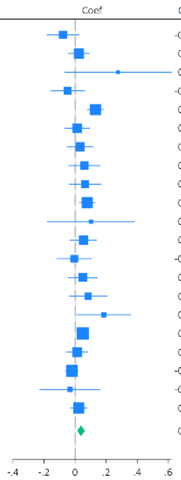

| Region | Country | N | Coef | 95% CI | Weight (%) |
|---|---|---|---|---|---|
| East Asia & Pacific | Australia | 2007 | -0.08 | [ -0.18, 0.02] | 3.85 |
| East Asia & Pacific | China | 3669 | 0.02 | [ -0.04, 0.09] | 6.39 |
| East Asia & Pacific | Hong Kong | 309 | 0.28 | [ -0.07, 0.62] | 0.48 |
| East Asia & Pacific | Indonesia | 1485 | -0.05 | [ -0.16, 0.06] | 3.51 |
| East Asia & Pacific | Japan | 8263 | 0.13 | [ 0.08, 0.18] | 7.84 |
| East Asia & Pacific | Philippines | 2061 | 0.01 | [ -0.06, 0.09] | 5.30 |
| Europe & Central Asia | Germany | 2923 | 0.03 | [ -0.05, 0.12] | 5.00 |
| Europe & Central Asia | Poland | 3166 | 0.06 | [ -0.04, 0.16] | 3.93 |
| Europe & Central Asia | Spain | 1541 | 0.06 | [ -0.04, 0.16] | 3.90 |
| Europe & Central Asia | Sweden | 8824 | 0.08 | [ 0.03, 0.13] | 7.91 |
| Europe & Central Asia | Türkiye | 253 | 0.10 | [ -0.18, 0.38] | 0.70 |
| Europe & Central Asia | United Kingdom | 2479 | 0.05 | [ -0.03, 0.14] | 4.88 |
| Latin America & Caribbean | Argentina | 1538 | -0.01 | [ -0.12, 0.11] | 3.40 |
| Latin America & Caribbean | Brazil | 2219 | 0.05 | [ -0.04, 0.14] | 4.40 |
| Latin America & Caribbean | Mexico | 1096 | 0.08 | [ -0.04, 0.21] | 2.96 |
| Middle East, North Africa, Afghanistan & Pakistan | Egypt | 2210 | 0.18 | [ 0.01, 0.36] | 1.69 |
| North America | United States | 22702 | 0.05 | [ 0.01, 0.08] | 9.71 |
| South Asia | India | 3845 | 0.01 | [ -0.06, 0.08] | 6.11 |
| Sub-Saharan Africa | Kenya | 6743 | -0.02 | [ -0.06, 0.02] | 8.91 |
| Sub-Saharan Africa | South Africa | 430 | -0.03 | [ -0.23, 0.16] | 1.37 |
| Sub-Saharan Africa | Tanzania | 4215 | 0.02 | [ -0.03, 0.08] | 7.77 |
| | **Overall** | | 0.04 | [ 0.01, 0.06] | |

$\tau^2 = 0.00$, $I^2 = 47.1\%$, $H^2 = 1.89$

o) Remain hopeful about the future

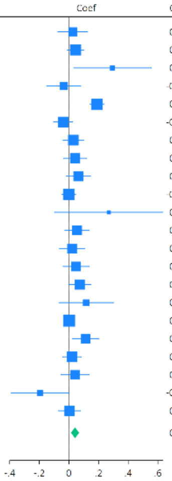

| Region | Country | N | Coef | 95% CI | Weight (%) |
|---|---|---|---|---|---|
| East Asia & Pacific | Australia | 1934 | 0.03 | [ -0.07, 0.13] | 3.99 |
| East Asia & Pacific | China | 3673 | 0.04 | [ -0.01, 0.10] | 6.30 |
| East Asia & Pacific | Hong Kong | 309 | 0.29 | [ 0.03, 0.56] | 0.99 |
| East Asia & Pacific | Indonesia | 1484 | -0.04 | [ -0.15, 0.08] | 3.44 |
| East Asia & Pacific | Japan | 8266 | 0.19 | [ 0.14, 0.24] | 6.69 |
| East Asia & Pacific | Philippines | 2062 | -0.04 | [ -0.10, 0.03] | 5.78 |
| Europe & Central Asia | Germany | 2928 | 0.03 | [ -0.04, 0.10] | 5.53 |
| Europe & Central Asia | Poland | 3195 | 0.04 | [ -0.03, 0.12] | 5.12 |
| Europe & Central Asia | Spain | 1546 | 0.06 | [ -0.02, 0.15] | 4.86 |
| Europe & Central Asia | Sweden | 8837 | -0.00 | [ -0.05, 0.04] | 6.79 |
| Europe & Central Asia | Türkiye | 251 | 0.27 | [ -0.10, 0.63] | 0.54 |
| Europe & Central Asia | United Kingdom | 2446 | 0.05 | [ -0.03, 0.14] | 4.83 |
| Latin America & Caribbean | Argentina | 1542 | 0.02 | [ -0.06, 0.11] | 4.72 |
| Latin America & Caribbean | Brazil | 2219 | 0.05 | [ -0.04, 0.14] | 4.56 |
| Latin America & Caribbean | Mexico | 1101 | 0.07 | [ -0.00, 0.15] | 5.23 |
| Middle East, North Africa, Afghanistan & Pakistan | Egypt | 2198 | 0.12 | [ -0.07, 0.30] | 1.79 |
| North America | United States | 21460 | 0.00 | [ -0.03, 0.04] | 7.39 |
| South Asia | India | 3854 | 0.11 | [ 0.02, 0.20] | 4.47 |
| Sub-Saharan Africa | Kenya | 6740 | 0.02 | [ -0.04, 0.08] | 5.84 |
| Sub-Saharan Africa | Nigeria | 2498 | 0.04 | [ -0.05, 0.14] | 4.21 |
| Sub-Saharan Africa | South Africa | 429 | -0.20 | [ -0.39, -0.00] | 1.65 |
| Sub-Saharan Africa | Tanzania | 4212 | 0.00 | [ -0.07, 0.08] | 5.27 |
| | **Overall** | | 0.04 | [ 0.01, 0.07] | |

$\tau^2 = 0.00$, $I^2 = 63.7\%$, $H^2 = 2.75$

p) Feeling very capable in most things

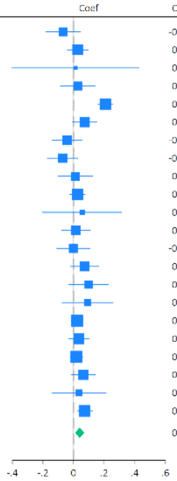

| Region | Country | N | Coef | 95% CI | Weight (%) |
|---|---|---|---|---|---|
| East Asia & Pacific | Australia | 1994 | -0.07 | [ -0.18, 0.04] | 3.77 |
| East Asia & Pacific | China | 3667 | 0.03 | [ -0.04, 0.09] | 5.87 |
| East Asia & Pacific | Hong Kong | 309 | 0.01 | [ -0.40, 0.43] | 0.48 |
| East Asia & Pacific | Indonesia | 1481 | 0.03 | [ -0.09, 0.14] | 3.75 |
| East Asia & Pacific | Japan | 8227 | 0.21 | [ 0.16, 0.26] | 6.87 |
| East Asia & Pacific | Philippines | 2063 | 0.07 | [ -0.01, 0.15] | 5.27 |
| Europe & Central Asia | Germany | 2924 | -0.04 | [ -0.14, 0.06] | 4.40 |
| Europe & Central Asia | Poland | 3151 | -0.07 | [ -0.17, 0.03] | 4.33 |
| Europe & Central Asia | Spain | 1530 | 0.01 | [ -0.10, 0.12] | 3.78 |
| Europe & Central Asia | Sweden | 8791 | 0.03 | [ -0.02, 0.08] | 6.80 |
| Europe & Central Asia | Türkiye | 252 | 0.06 | [ -0.20, 0.31] | 1.13 |
| Europe & Central Asia | United Kingdom | 2466 | 0.02 | [ -0.08, 0.11] | 4.54 |
| Latin America & Caribbean | Argentina | 1543 | -0.00 | [ -0.11, 0.11] | 3.99 |
| Latin America & Caribbean | Brazil | 2208 | 0.07 | [ -0.02, 0.16] | 4.69 |
| Latin America & Caribbean | Mexico | 1094 | 0.10 | [ -0.03, 0.23] | 3.24 |
| Middle East, North Africa, Afghanistan & Pakistan | Egypt | 2209 | 0.09 | [ -0.07, 0.26] | 2.29 |
| North America | United States | 22625 | 0.02 | [ -0.01, 0.06] | 7.41 |
| South Asia | India | 3858 | 0.03 | [ -0.03, 0.10] | 5.91 |
| Sub-Saharan Africa | Kenya | 6735 | 0.02 | [ -0.02, 0.06] | 7.23 |
| Sub-Saharan Africa | Nigeria | 2492 | 0.06 | [ -0.01, 0.14] | 5.28 |
| Sub-Saharan Africa | South Africa | 430 | 0.04 | [ -0.14, 0.21] | 2.12 |
| Sub-Saharan Africa | Tanzania | 4212 | 0.07 | [ 0.02, 0.12] | 6.85 |
| | **Overall** | | 0.04 | [ 0.01, 0.07] | |

$\tau^2 = 0.00$, $I^2 = 65.8\%$, $H^2 = 2.92$

q) Expect more good things to happen to you

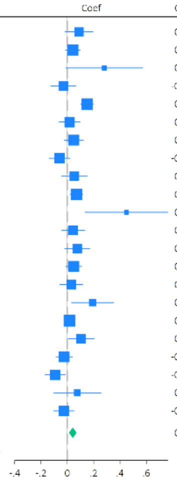

| Region | Country | N | Coef | 95% CI | Weight (%) |
|---|---|---|---|---|---|
| East Asia & Pacific | Australia | 1988 | 0.09 | [ -0.02, 0.19] | 3.89 |
| East Asia & Pacific | China | 3673 | 0.04 | [ -0.01, 0.10] | 6.16 |
| East Asia & Pacific | Hong Kong | 308 | 0.28 | [ -0.01, 0.57] | 0.89 |
| East Asia & Pacific | Indonesia | 1487 | -0.03 | [ -0.12, 0.07] | 4.28 |
| East Asia & Pacific | Japan | 8308 | 0.15 | [ 0.10, 0.20] | 6.54 |
| East Asia & Pacific | Philippines | 2061 | 0.02 | [ -0.06, 0.09] | 5.04 |
| Europe & Central Asia | Germany | 2929 | 0.05 | [ -0.02, 0.12] | 5.37 |
| Europe & Central Asia | Poland | 3191 | -0.06 | [ -0.14, 0.02] | 5.02 |
| Europe & Central Asia | Spain | 1548 | 0.05 | [ -0.04, 0.15] | 4.20 |
| Europe & Central Asia | Sweden | 8827 | 0.07 | [ 0.02, 0.12] | 6.61 |
| Europe & Central Asia | Türkiye | 253 | 0.45 | [ 0.13, 0.76] | 0.77 |
| Europe & Central Asia | United Kingdom | 2470 | 0.04 | [ -0.05, 0.13] | 4.60 |
| Latin America & Caribbean | Argentina | 1545 | 0.07 | [ -0.02, 0.17] | 4.30 |
| Latin America & Caribbean | Brazil | 2221 | 0.05 | [ -0.01, 0.11] | 5.87 |
| Latin America & Caribbean | Mexico | 1103 | 0.03 | [ -0.05, 0.11] | 4.74 |
| Middle East, North Africa, Afghanistan & Pakistan | Egypt | 2161 | 0.19 | [ 0.03, 0.35] | 2.35 |
| North America | United States | 22673 | 0.02 | [ -0.02, 0.05] | 7.01 |
| South Asia | India | 3854 | 0.11 | [ 0.01, 0.20] | 4.18 |
| Sub-Saharan Africa | Kenya | 6747 | -0.02 | [ -0.09, 0.04] | 5.82 |
| Sub-Saharan Africa | Nigeria | 2494 | -0.09 | [ -0.17, -0.02] | 5.17 |
| Sub-Saharan Africa | South Africa | 427 | 0.08 | [ -0.10, 0.25] | 2.00 |
| Sub-Saharan Africa | Tanzania | 4211 | -0.03 | [ -0.10, 0.05] | 5.18 |
| | **Overall** | | 0.04 | [ 0.01, 0.07] | |

$\tau^2 = 0.00$, $I^2 = 67.1\%$, $H^2 = 3.04$

r) Overall mental health

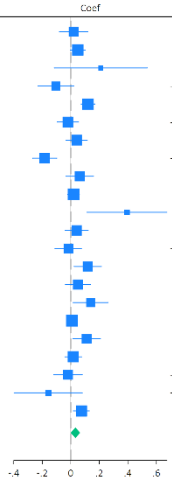

| Region | Country | N | Coef | 95% CI | Weight (%) |
|---|---|---|---|---|---|
| East Asia & Pacific | Australia | 2009 | 0.02 | [ -0.08, 0.12] | 4.35 |
| East Asia & Pacific | China | 3670 | 0.05 | [ -0.00, 0.10] | 6.08 |
| East Asia & Pacific | Hong Kong | 307 | 0.21 | [ -0.12, 0.54] | 0.89 |
| East Asia & Pacific | Indonesia | 1486 | -0.10 | [ -0.23, 0.02] | 3.51 |
| East Asia & Pacific | Japan | 8320 | 0.12 | [ 0.07, 0.17] | 6.27 |
| East Asia & Pacific | Philippines | 2062 | -0.02 | [ -0.10, 0.05] | 5.26 |
| Europe & Central Asia | Germany | 2931 | 0.04 | [ -0.03, 0.12] | 5.27 |
| Europe & Central Asia | Poland | 3196 | -0.18 | [ -0.27, -0.10] | 4.90 |
| Europe & Central Asia | Spain | 1546 | 0.06 | [ -0.03, 0.16] | 4.47 |
| Europe & Central Asia | Sweden | 8855 | 0.02 | [ -0.02, 0.06] | 6.51 |
| Europe & Central Asia | Türkiye | 252 | 0.39 | [ 0.12, 0.67] | 1.16 |
| Europe & Central Asia | United Kingdom | 2465 | 0.04 | [ -0.04, 0.12] | 5.01 |
| Latin America & Caribbean | Argentina | 1546 | -0.02 | [ -0.11, 0.08] | 4.53 |
| Latin America & Caribbean | Brazil | 2225 | 0.12 | [ 0.02, 0.22] | 4.48 |
| Latin America & Caribbean | Mexico | 1100 | 0.05 | [ -0.04, 0.14] | 4.75 |
| Middle East, North Africa, Afghanistan & Pakistan | Egypt | 2199 | 0.14 | [ 0.02, 0.26] | 3.63 |
| North America | United States | 22777 | 0.01 | [ -0.02, 0.04] | 6.79 |
| South Asia | India | 3857 | 0.11 | [ 0.01, 0.21] | 4.44 |
| Sub-Saharan Africa | Kenya | 6738 | 0.02 | [ -0.04, 0.08] | 5.82 |
| Sub-Saharan Africa | Nigeria | 2500 | -0.02 | [ -0.12, 0.08] | 4.36 |
| Sub-Saharan Africa | South Africa | 429 | -0.16 | [ -0.39, 0.08] | 1.52 |
| Sub-Saharan Africa | Tanzania | 4201 | 0.07 | [ 0.02, 0.13] | 6.00 |
| | **Overall** | | 0.03 | [ -0.00, 0.07] | |

$\tau^2 = 0.00$, $I^2 = 74.7\%$, $H^2 = 3.95$

s) Overall physical health

| Region | Country | N | Coef | 95% CI | Weight (%) |
|---|---|---|---|---|---|
| East Asia & Pacific | Australia | 2010 | 0.04 | [ -0.05, 0.14] | 4.59 |
| East Asia & Pacific | China | 3673 | 0.06 | [ 0.00, 0.11] | 5.97 |
| East Asia & Pacific | Hong Kong | 309 | 0.16 | [ -0.17, 0.49] | 1.00 |
| East Asia & Pacific | Indonesia | 1487 | 0.03 | [ -0.10, 0.16] | 3.66 |
| East Asia & Pacific | Japan | 8289 | 0.15 | [ 0.10, 0.19] | 6.21 |
| East Asia & Pacific | Philippines | 2062 | 0.02 | [ -0.06, 0.10] | 5.05 |
| Europe & Central Asia | Germany | 2940 | 0.15 | [ 0.09, 0.21] | 5.70 |
| Europe & Central Asia | Poland | 3195 | -0.17 | [ -0.25, -0.09] | 5.20 |
| Europe & Central Asia | Spain | 1549 | 0.07 | [ -0.02, 0.17] | 4.67 |
| Europe & Central Asia | Sweden | 8860 | 0.05 | [ 0.01, 0.09] | 6.39 |
| Europe & Central Asia | Türkiye | 253 | 0.27 | [ 0.02, 0.52] | 1.59 |
| Europe & Central Asia | United Kingdom | 2494 | 0.07 | [ -0.00, 0.15] | 5.25 |
| Latin America & Caribbean | Argentina | 1547 | 0.08 | [ -0.02, 0.17] | 4.76 |
| Latin America & Caribbean | Brazil | 2220 | 0.13 | [ 0.04, 0.21] | 4.92 |
| Latin America & Caribbean | Mexico | 1103 | 0.12 | [ 0.03, 0.21] | 4.76 |
| Middle East, North Africa, Afghanistan & Pakistan | Egypt | 2208 | 0.16 | [ -0.02, 0.35] | 2.38 |
| North America | United States | 22775 | 0.08 | [ 0.05, 0.11] | 6.63 |
| South Asia | India | 3858 | 0.08 | [ -0.02, 0.18] | 4.52 |
| Sub-Saharan Africa | Kenya | 6743 | 0.03 | [ -0.03, 0.10] | 5.64 |
| Sub-Saharan Africa | Nigeria | 2502 | -0.09 | [ -0.20, 0.02] | 4.21 |
| Sub-Saharan Africa | South Africa | 431 | -0.19 | [ -0.44, 0.05] | 1.64 |
| Sub-Saharan Africa | Tanzania | 4215 | 0.14 | [ 0.06, 0.21] | 5.28 |
| | **Overall** | | 0.06 | [ 0.03, 0.10] | |

$\tau^2 = 0.00$, $I^2 = 79.0\%$, $H^2 = 4.77$

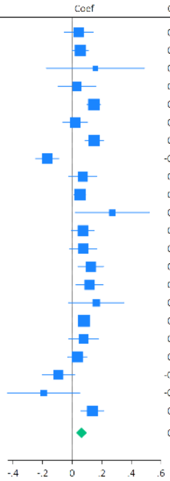

Supplementary Figure S10: Sensitivity analysis restricting the sample to participants below the healthiest baseline category

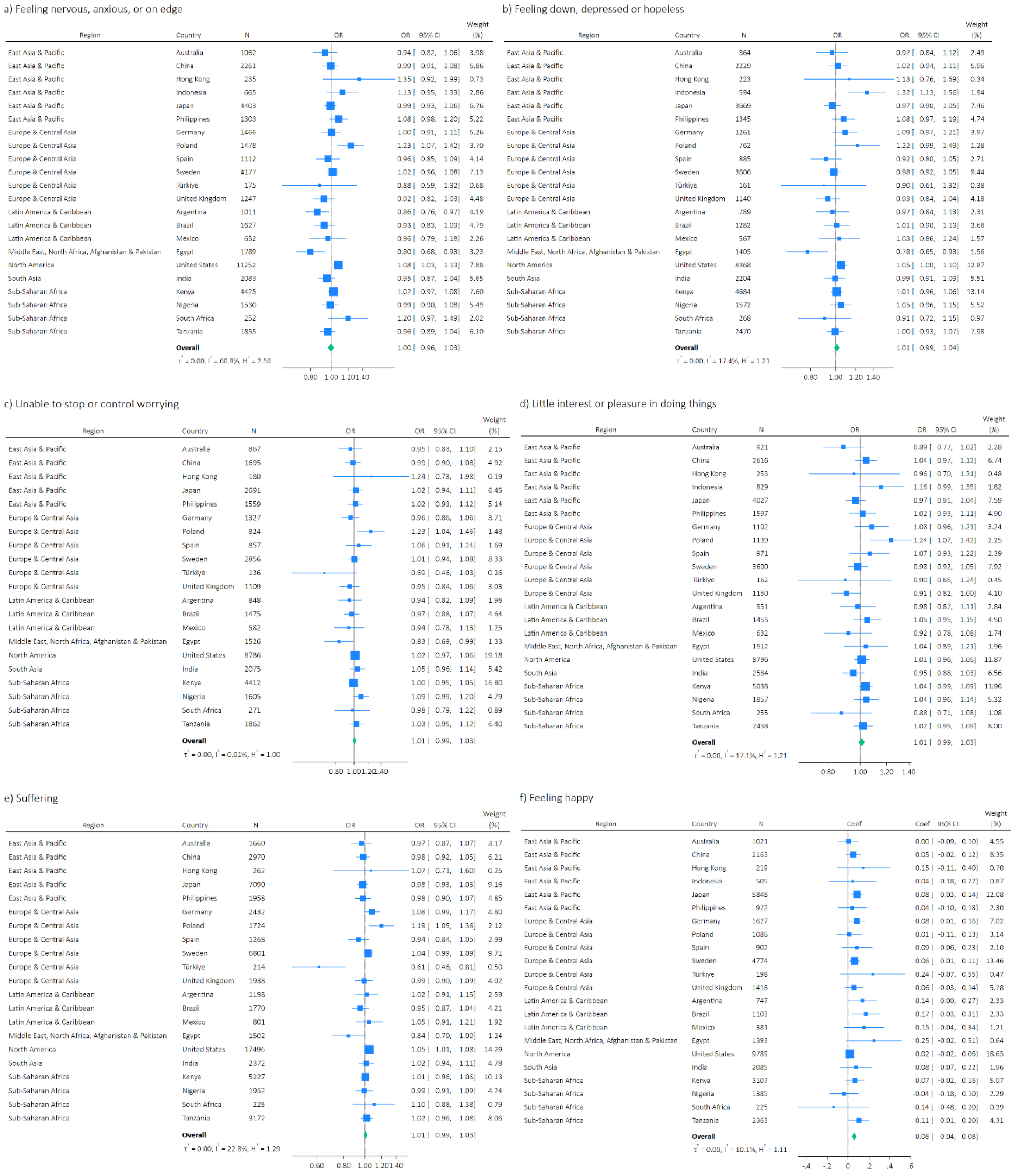

g) Understanding purpose in life

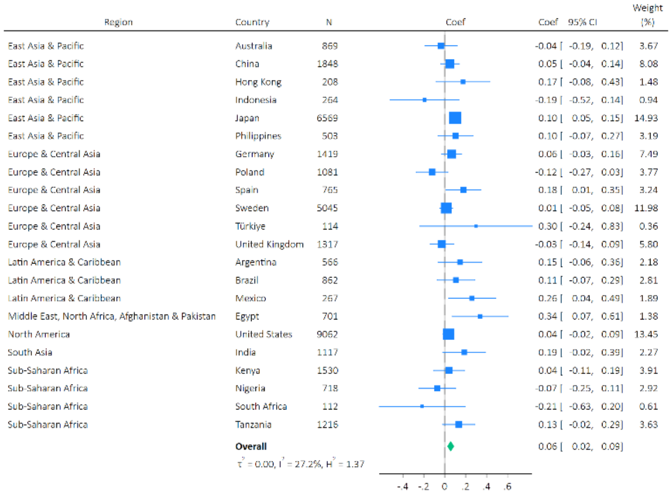

h) Life satisfaction

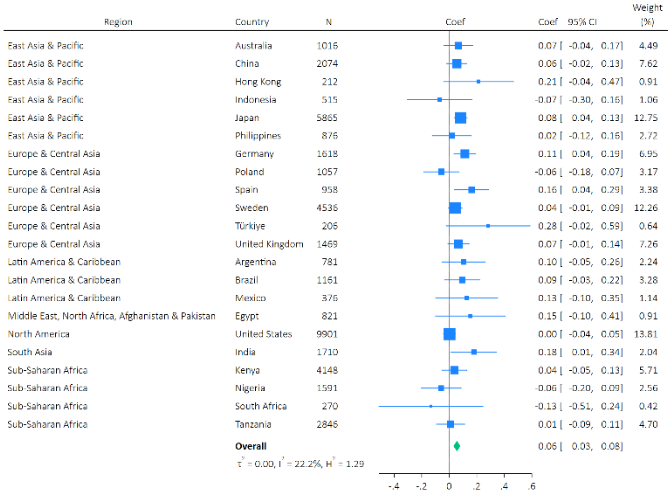

i) Life evaluation: Today

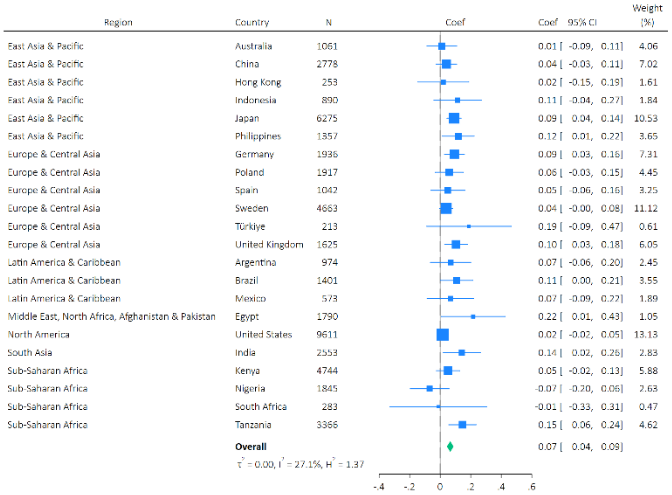

j) Life evaluation: Five years from now

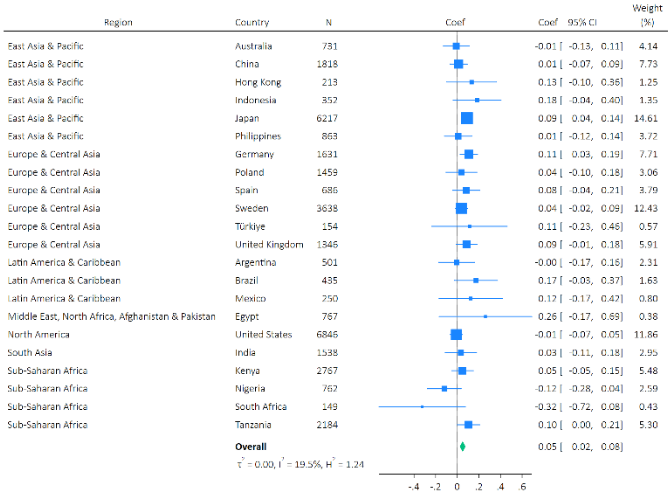

k) Feeling lonely

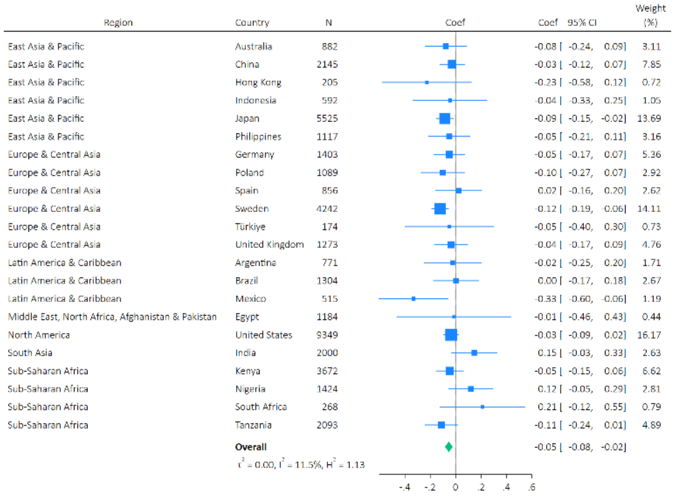

l) Life is worthwhile

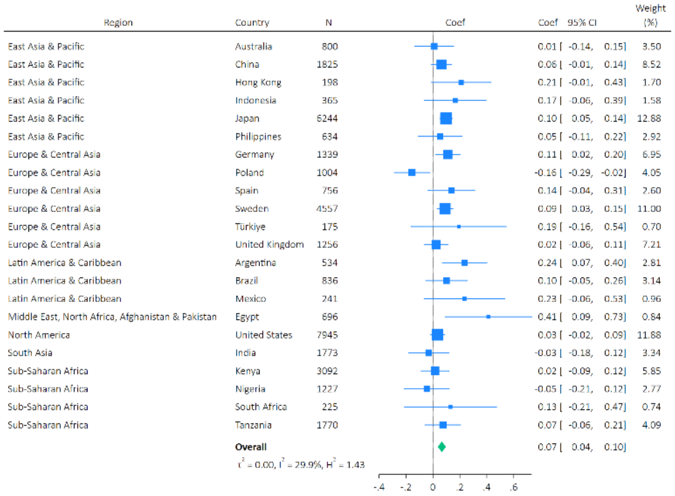

m) Feeling grateful

| Region | Country | N | Coef | 95% CI | Weight (%) |
|---|---|---|---|---|---|
| East Asia & Pacific | Australia | 531 | -0.13 | [ -0.34, 0.08] | 1.49 |
| East Asia & Pacific | China | 1780 | 0.09 | [ 0.01, 0.18] | 8.91 |
| East Asia & Pacific | Hong Kong | 225 | 0.16 | [ -0.11, 0.44] | 0.87 |
| East Asia & Pacific | Indonesia | 196 | 0.07 | [ -0.16, 0.30] | 1.21 |
| East Asia & Pacific | Japan | 6526 | 0.10 | [ 0.05, 0.15] | 22.02 |
| East Asia & Pacific | Philippines | 370 | 0.05 | [ -0.15, 0.24] | 1.76 |
| Europe & Central Asia | Germany | 1330 | 0.06 | [ -0.04, 0.17] | 5.74 |
| Europe & Central Asia | Poland | 1513 | 0.06 | [ -0.07, 0.19] | 3.97 |
| Europe & Central Asia | Spain | 637 | 0.07 | [ -0.08, 0.21] | 2.98 |
| Europe & Central Asia | Sweden | 4151 | 0.12 | [ 0.05, 0.18] | 14.88 |
| Europe & Central Asia | Türkiye | 146 | 0.24 | [ -0.14, 0.61] | 0.48 |
| Europe & Central Asia | United Kingdom | 1112 | 0.06 | [ -0.04, 0.17] | 5.82 |
| Latin America & Caribbean | Argentina | 371 | 0.26 | [ 0.05, 0.47] | 1.51 |
| Latin America & Caribbean | Brazil | 515 | 0.06 | [ -0.16, 0.28] | 1.36 |
| Latin America & Caribbean | Mexico | 177 | -0.08 | [ -0.44, 0.29] | 0.49 |
| Middle East, North Africa, Afghanistan & Pakistan | Egypt | 805 | -0.02 | [ -0.30, 0.26] | 0.83 |
| North America | United States | 4705 | 0.03 | [ -0.04, 0.11] | 11.76 |
| South Asia | India | 1433 | 0.16 | [ -0.01, 0.33] | 2.20 |
| Sub-Saharan Africa | Kenya | 2159 | 0.00 | [ -0.11, 0.12] | 4.94 |
| Sub-Saharan Africa | Nigeria | 752 | 0.08 | [ -0.08, 0.24] | 2.56 |
| Sub-Saharan Africa | South Africa | 140 | -0.45 | [ -0.79, -0.12] | 0.59 |
| Sub-Saharan Africa | Tanzania | 1331 | 0.01 | [ -0.12, 0.14] | 3.63 |
| | **Overall** | | 0.07 | [ 0.05, 0.10] | |

$\tau^2 = 0.00$, $I^2 = 3.06\%$, $H^2 = 1.03$

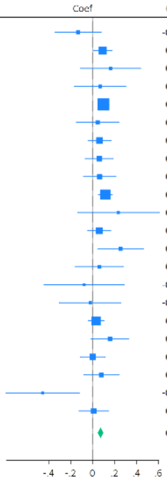

n) Feeling at peace

| Region | Country | N | Coef | 95% CI | Weight (%) |
|---|---|---|---|---|---|
| East Asia & Pacific | Australia | 1732 | -0.08 | [ -0.21, 0.04] | 3.11 |
| East Asia & Pacific | China | 2927 | -0.01 | [ -0.08, 0.07] | 6.67 |
| East Asia & Pacific | Hong Kong | 247 | 0.14 | [ -0.29, 0.56] | 0.32 |
| East Asia & Pacific | Indonesia | 989 | -0.07 | [ -0.22, 0.08] | 2.32 |
| East Asia & Pacific | Japan | 7283 | 0.08 | [ 0.02, 0.14] | 8.87 |
| East Asia & Pacific | Philippines | 1609 | 0.02 | [ -0.07, 0.11] | 5.10 |
| Europe & Central Asia | Germany | 2562 | 0.01 | [ -0.09, 0.11] | 4.61 |
| Europe & Central Asia | Poland | 2609 | 0.01 | [ -0.10, 0.11] | 3.88 |
| Europe & Central Asia | Spain | 1359 | 0.04 | [ -0.06, 0.15] | 4.17 |
| Europe & Central Asia | Sweden | 7838 | 0.07 | [ 0.01, 0.13] | 8.80 |
| Europe & Central Asia | Türkiye | 213 | 0.01 | [ -0.31, 0.34] | 0.53 |
| Europe & Central Asia | United Kingdom | 2119 | 0.06 | [ -0.04, 0.15] | 5.02 |
| Latin America & Caribbean | Argentina | 1123 | -0.04 | [ -0.18, 0.10] | 2.59 |
| Latin America & Caribbean | Brazil | 1754 | 0.07 | [ -0.04, 0.18] | 4.01 |
| Latin America & Caribbean | Mexico | 817 | 0.13 | [ -0.03, 0.28] | 2.19 |
| Middle East, North Africa, Afghanistan & Pakistan | Egypt | 1408 | 0.27 | [ 0.04, 0.50] | 1.02 |
| North America | United States | 18281 | 0.05 | [ 0.01, 0.10] | 11.81 |
| South Asia | India | 2294 | 0.01 | [ -0.08, 0.10] | 5.31 |
| Sub-Saharan Africa | Kenya | 4994 | -0.04 | [ -0.09, 0.01] | 10.58 |
| Sub-Saharan Africa | South Africa | 250 | 0.04 | [ -0.21, 0.28] | 0.92 |
| Sub-Saharan Africa | Tanzania | 2919 | 0.02 | [ -0.05, 0.08] | 8.15 |
| | **Overall** | | 0.03 | [ 0.00, 0.05] | |

$\tau^2 = 0.00$, $I^2 = 29.0\%$, $H^2 = 1.41$

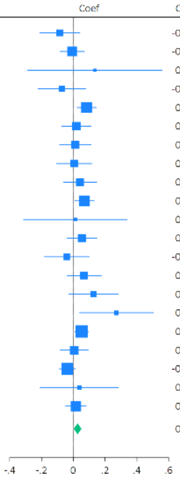

o) Remain hopeful about the future

| Region | Country | N | Coef | 95% CI | Weight (%) |
|---|---|---|---|---|---|
| East Asia & Pacific | Australia | 637 | 0.05 | [ -0.10, 0.21] | 3.64 |
| East Asia & Pacific | China | 1634 | 0.04 | [ -0.04, 0.13] | 9.36 |
| East Asia & Pacific | Hong Kong | 207 | 0.05 | [ -0.19, 0.29] | 1.66 |
| East Asia & Pacific | Indonesia | 145 | -0.14 | [ -0.50, 0.21] | 0.80 |
| East Asia & Pacific | Japan | 6376 | 0.08 | [ 0.04, 0.13] | 15.47 |
| East Asia & Pacific | Philippines | 301 | 0.04 | [ -0.14, 0.22] | 2.74 |
| Europe & Central Asia | Germany | 1209 | 0.04 | [ -0.06, 0.14] | 6.95 |
| Europe & Central Asia | Poland | 1004 | 0.05 | [ -0.08, 0.18] | 4.70 |
| Europe & Central Asia | Spain | 509 | 0.11 | [ -0.01, 0.23] | 5.34 |
| Europe & Central Asia | Sweden | 4425 | -0.02 | [ -0.08, 0.04] | 12.71 |
| Europe & Central Asia | Türkiye | 108 | 0.22 | [ -0.42, 0.85] | 0.26 |
| Europe & Central Asia | United Kingdom | 1081 | -0.00 | [ -0.11, 0.11] | 6.24 |
| Latin America & Caribbean | Argentina | 280 | 0.06 | [ -0.18, 0.30] | 1.71 |
| Latin America & Caribbean | Brazil | 415 | 0.19 | [ -0.02, 0.41] | 2.09 |
| Latin America & Caribbean | Mexico | 108 | 0.35 | [ 0.01, 0.68] | 0.92 |
| Middle East, North Africa, Afghanistan & Pakistan | Egypt | 601 | 0.03 | [ -0.32, 0.37] | 0.85 |
| North America | United States | 6902 | -0.04 | [ -0.10, 0.02] | 12.92 |
| South Asia | India | 979 | 0.07 | [ -0.15, 0.29] | 1.99 |
| Sub-Saharan Africa | Kenya | 1314 | 0.02 | [ -0.13, 0.17] | 3.95 |
| Sub-Saharan Africa | Nigeria | 523 | -0.04 | [ -0.25, 0.17] | 2.15 |
| Sub-Saharan Africa | South Africa | 64 | 0.25 | [ -0.32, 0.82] | 0.32 |
| Sub-Saharan Africa | Tanzania | 1014 | 0.02 | [ -0.14, 0.19] | 3.23 |
| | **Overall** | | 0.03 | [ 0.00, 0.07] | |

$\tau^2 = 0.00$, $I^2 = 25.2\%$, $H^2 = 1.34$

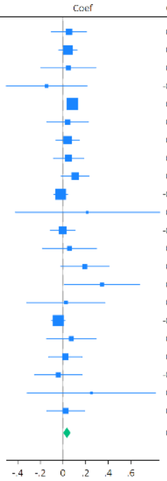

p) Feeling very capable in most things

| Region | Country | N | Coef | 95% CI | Weight (%) |
|---|---|---|---|---|---|
| East Asia & Pacific | Australia | 1688 | -0.07 | [ -0.21, 0.06] | 3.62 |
| East Asia & Pacific | China | 2929 | 0.02 | [ -0.06, 0.09] | 6.51 |
| East Asia & Pacific | Hong Kong | 252 | -0.17 | [ -0.69, 0.35] | 0.34 |
| East Asia & Pacific | Indonesia | 980 | -0.03 | [ -0.17, 0.11] | 3.32 |
| East Asia & Pacific | Japan | 8021 | 0.15 | [ 0.10, 0.20] | 8.46 |
| East Asia & Pacific | Philippines | 1453 | 0.03 | [ -0.07, 0.13] | 5.31 |
| Europe & Central Asia | Germany | 2691 | -0.03 | [ -0.14, 0.08] | 4.52 |
| Europe & Central Asia | Poland | 2120 | -0.09 | [ -0.22, 0.04] | 3.81 |
| Europe & Central Asia | Spain | 1186 | -0.09 | [ -0.23, 0.05] | 3.43 |
| Europe & Central Asia | Sweden | 7526 | -0.00 | [ -0.06, 0.06] | 7.89 |
| Europe & Central Asia | Türkiye | 172 | 0.01 | [ -0.37, 0.39] | 0.62 |
| Europe & Central Asia | United Kingdom | 2136 | 0.01 | [ -0.10, 0.12] | 4.62 |
| Latin America & Caribbean | Argentina | 993 | 0.02 | [ -0.13, 0.17] | 3.12 |
| Latin America & Caribbean | Brazil | 1620 | 0.06 | [ -0.06, 0.17] | 4.30 |
| Latin America & Caribbean | Mexico | 646 | 0.22 | [ 0.05, 0.38] | 2.54 |
| Middle East, North Africa, Afghanistan & Pakistan | Egypt | 1137 | -0.06 | [ -0.32, 0.19] | 1.31 |
| North America | United States | 18250 | 0.00 | [ -0.04, 0.05] | 8.69 |
| South Asia | India | 1768 | 0.02 | [ -0.08, 0.11] | 5.31 |
| Sub-Saharan Africa | Kenya | 4270 | 0.01 | [ -0.04, 0.06] | 8.31 |
| Sub-Saharan Africa | Nigeria | 1473 | 0.03 | [ -0.07, 0.13] | 4.98 |
| Sub-Saharan Africa | South Africa | 215 | -0.09 | [ -0.33, 0.14] | 1.47 |
| Sub-Saharan Africa | Tanzania | 2698 | 0.09 | [ 0.02, 0.15] | 7.51 |
| | **Overall** | | 0.02 | [ -0.01, 0.05] | |

$\tau^2 = 0.00$, $I^2 = 52.0\%$, $H^2 = 2.08$

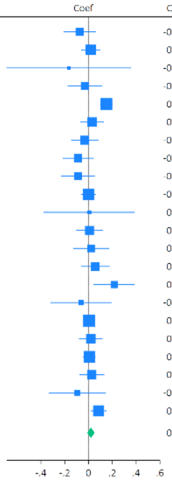

q) Expect more good things to happen to you

| Region | Country | N | Coef | 95% CI | Weight (%) |
|---|---|---|---|---|---|
| East Asia & Pacific | Australia | 708 | 0.18 | [ 0.02, 0.34] | 3.45 |
| East Asia & Pacific | China | 1476 | 0.04 | [ -0.05, 0.13] | 8.79 |
| East Asia & Pacific | Hong Kong | 195 | 0.07 | [ -0.19, 0.33] | 1.33 |
| East Asia & Pacific | Indonesia | 165 | -0.38 | [ -0.83, 0.08] | 0.45 |
| East Asia & Pacific | Japan | 5061 | 0.06 | [ -0.00, 0.12] | 15.85 |
| East Asia & Pacific | Philippines | 330 | 0.10 | [ -0.13, 0.32] | 1.76 |
| Europe & Central Asia | Germany | 1291 | 0.02 | [ -0.08, 0.12] | 7.93 |
| Europe & Central Asia | Poland | 1159 | -0.04 | [ -0.19, 0.11] | 3.62 |
| Europe & Central Asia | Spain | 466 | 0.11 | [ -0.03, 0.25] | 4.12 |
| Europe & Central Asia | Sweden | 4209 | 0.06 | [ -0.00, 0.13] | 14.23 |
| Europe & Central Asia | Türkiye | 106 | 0.28 | [ -0.19, 0.75] | 0.41 |
| Europe & Central Asia | United Kingdom | 1264 | -0.04 | [ -0.15, 0.06] | 6.70 |
| Latin America & Caribbean | Argentina | 284 | 0.10 | [ -0.17, 0.37] | 1.25 |
| Latin America & Caribbean | Brazil | 288 | 0.08 | [ -0.16, 0.32] | 1.54 |
| Latin America & Caribbean | Mexico | 115 | 0.06 | [ -0.33, 0.45] | 0.61 |
| Middle East, North Africa, Afghanistan & Pakistan | Egypt | 826 | 0.23 | [ -0.01, 0.46] | 1.61 |
| North America | United States | 6722 | -0.02 | [ -0.08, 0.04] | 14.76 |
| South Asia | India | 1093 | 0.15 | [ -0.05, 0.35] | 2.21 |
| Sub-Saharan Africa | Kenya | 1134 | -0.09 | [ -0.24, 0.07] | 3.43 |
| Sub-Saharan Africa | Nigeria | 424 | -0.03 | [ -0.19, 0.14] | 3.03 |
| Sub-Saharan Africa | South Africa | 108 | 0.26 | [ -0.10, 0.62] | 0.72 |
| Sub-Saharan Africa | Tanzania | 745 | -0.05 | [ -0.25, 0.15] | 2.18 |
| | **Overall** | | 0.04 | [ 0.00, 0.07] | |

$\tau^2 = 0.00$, $I^2 = 13.7\%$, $H^2 = 1.16$

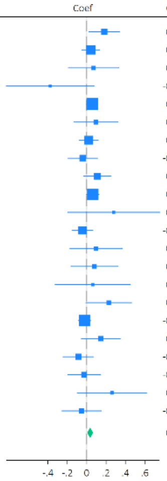

r) Overall mental health

| Region | Country | N | Coef | 95% CI | Weight (%) |
|---|---|---|---|---|---|
| East Asia & Pacific | Australia | 904 | 0.01 | [ -0.12, 0.14] | 2.90 |
| East Asia & Pacific | China | 1544 | 0.09 | [ 0.00, 0.18] | 5.99 |
| East Asia & Pacific | Hong Kong | 195 | -0.02 | [ -0.23, 0.19] | 1.06 |
| East Asia & Pacific | Indonesia | 334 | -0.21 | [ -0.50, 0.07] | 0.58 |
| East Asia & Pacific | Japan | 5653 | 0.06 | [ 0.02, 0.11] | 19.62 |
| East Asia & Pacific | Philippines | 641 | 0.02 | [ -0.11, 0.15] | 2.72 |
| Europe & Central Asia | Germany | 1440 | 0.04 | [ -0.04, 0.13] | 6.43 |
| Europe & Central Asia | Poland | 816 | 0.05 | [ -0.12, 0.21] | 1.75 |
| Europe & Central Asia | Spain | 689 | 0.10 | [ -0.04, 0.23] | 2.58 |
| Europe & Central Asia | Sweden | 4156 | 0.05 | [ -0.01, 0.10] | 17.10 |
| Europe & Central Asia | Türkiye | 169 | 0.29 | [ -0.03, 0.61] | 0.45 |
| Europe & Central Asia | United Kingdom | 1175 | -0.02 | [ -0.12, 0.08] | 4.53 |
| Latin America & Caribbean | Argentina | 527 | -0.07 | [ -0.23, 0.10] | 1.74 |
| Latin America & Caribbean | Brazil | 1015 | 0.14 | [ 0.02, 0.26] | 3.23 |
| Latin America & Caribbean | Mexico | 265 | 0.06 | [ -0.11, 0.22] | 1.74 |
| Middle East, North Africa, Afghanistan & Pakistan | Egypt | 442 | 0.08 | [ -0.37, 0.54] | 0.22 |
| North America | United States | 7645 | 0.01 | [ -0.03, 0.06] | 21.51 |
| South Asia | India | 1461 | 0.09 | [ -0.08, 0.26] | 1.61 |
| Sub-Saharan Africa | Kenya | 1345 | 0.04 | [ -0.11, 0.20] | 1.90 |
| Sub-Saharan Africa | Nigeria | 588 | -0.16 | [ -0.41, 0.08] | 0.79 |
| Sub-Saharan Africa | South Africa | 114 | -0.27 | [ -0.77, 0.22] | 0.19 |
| Sub-Saharan Africa | Tanzania | 651 | 0.10 | [ -0.09, 0.29] | 1.35 |
| | **Overall** | | 0.04 | [ 0.02, 0.06] | |

$\tau^2 = 0.00$, $I^2 = 0.00\%$, $H^2 = 1.00$

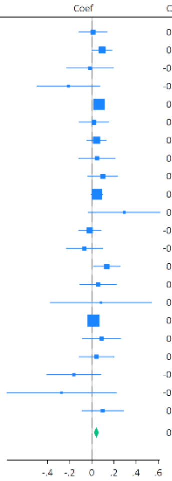

s) Overall physical health

| Region | Country | N | Coef | Coef [95% CI] | Weight (%) |
|---|---|---|---|---|---|
| East Asia & Pacific | Australia | 1201 | | -0.04 [ -0.15, 0.06] | 5.09 |
| East Asia & Pacific | China | 1874 | | 0.04 [ -0.04, 0.12] | 6.77 |
| East Asia & Pacific | Hong Kong | 234 | | -0.13 [ -0.32, 0.06] | 2.45 |
| East Asia & Pacific | Indonesia | 418 | | 0.19 [ -0.02, 0.40] | 2.09 |
| East Asia & Pacific | Japan | 6355 | | 0.12 [ 0.08, 0.17] | 8.86 |
| East Asia & Pacific | Philippines | 819 | | 0.02 [ -0.11, 0.15] | 4.04 |
| Europe & Central Asia | Germany | 1953 | | 0.11 [ 0.04, 0.17] | 7.52 |
| Europe & Central Asia | Poland | 1060 | | -0.02 [ -0.14, 0.11] | 4.23 |
| Europe & Central Asia | Spain | 947 | | 0.03 [ -0.09, 0.15] | 4.63 |
| Europe & Central Asia | Sweden | 6113 | | 0.03 [ -0.01, 0.08] | 9.08 |
| Europe & Central Asia | Türkiye | 131 | | 0.28 [ -0.03, 0.59] | 1.10 |
| Europe & Central Asia | United Kingdom | 1672 | | 0.02 [ -0.06, 0.10] | 6.53 |
| Latin America & Caribbean | Argentina | 814 | | 0.05 [ -0.08, 0.19] | 4.12 |
| Latin America & Caribbean | Brazil | 1131 | | 0.18 [ 0.07, 0.29] | 5.19 |
| Latin America & Caribbean | Mexico | 385 | | 0.11 [ -0.05, 0.27] | 3.31 |
| Middle East, North Africa, Afghanistan & Pakistan | Egypt | 1221 | | 0.21 [ -0.11, 0.52] | 1.05 |
| North America | United States | 11763 | | 0.06 [ 0.02, 0.09] | 9.40 |
| South Asia | India | 1655 | | 0.05 [ -0.10, 0.20] | 3.40 |
| Sub-Saharan Africa | Kenya | 2081 | | 0.07 [ -0.05, 0.19] | 4.54 |
| Sub-Saharan Africa | Nigeria | 735 | | -0.32 [ -0.52, -0.13] | 2.41 |
| Sub-Saharan Africa | South Africa | 153 | | -0.10 [ -0.61, 0.42] | 0.43 |
| Sub-Saharan Africa | Tanzania | 1492 | | 0.18 [ 0.03, 0.32] | 3.74 |
| | **Overall** | | | 0.05 [ 0.02, 0.09] | |

$\tau^2 = 0.00$, $I^2 = 60.0\%$, $H^2 = 2.50$

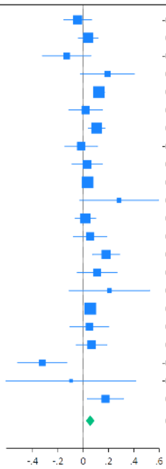

### Supplementary Figure S11: Sensitivity analysis modelling arts engagement as a binary variable

#### a) Feeling nervous, anxious, or on edge

| Region | Country | N | OR | 95% CI | Weight (%) |
|---|---|---|---|---|---|
| East Asia & Pacific | Australia | 2000 | 1.32 | [0.87, 2.00] | 2.87 |
| East Asia & Pacific | China | 3672 | 1.12 | [0.93, 1.34] | 5.88 |
| East Asia & Pacific | Hong Kong | 309 | 0.96 | [0.52, 1.77] | 1.65 |
| East Asia & Pacific | Indonesia | 1487 | 1.32 | [0.97, 1.79] | 4.03 |
| East Asia & Pacific | Japan | 8240 | 1.08 | [0.97, 1.21] | 6.87 |
| East Asia & Pacific | Philippines | 2061 | 1.29 | [1.05, 1.60] | 5.35 |
| Europe & Central Asia | Germany | 2921 | 1.06 | [0.85, 1.32] | 5.22 |
| Europe & Central Asia | Poland | 3175 | 1.52 | [1.24, 1.86] | 5.54 |
| Europe & Central Asia | Spain | 1546 | 1.07 | [0.74, 1.56] | 3.30 |
| Europe & Central Asia | Sweden | 8822 | 0.92 | [0.80, 1.07] | 6.41 |
| Europe & Central Asia | Türkiye | 251 | 0.75 | [0.28, 2.00] | 0.75 |
| Europe & Central Asia | United Kingdom | 2476 | 0.94 | [0.71, 1.23] | 4.45 |
| Latin America & Caribbean | Argentina | 1541 | 0.71 | [0.52, 0.97] | 3.97 |
| Latin America & Caribbean | Brazil | 2217 | 0.84 | [0.65, 1.09] | 4.63 |
| Latin America & Caribbean | Mexico | 1099 | 1.07 | [0.75, 1.53] | 3.45 |
| Middle East, North Africa, Afghanistan & Pakistan | Egypt | 2211 | 0.66 | [0.50, 0.88] | 4.32 |
| North America | United States | 22708 | 1.35 | [1.19, 1.54] | 6.62 |
| South Asia | India | 3849 | 0.96 | [0.82, 1.13] | 6.20 |
| Sub-Saharan Africa | Kenya | 6728 | 1.16 | [1.01, 1.34] | 6.47 |
| Sub-Saharan Africa | Nigeria | 2480 | 0.88 | [0.64, 1.20] | 3.88 |
| Sub-Saharan Africa | South Africa | 429 | 1.01 | [0.57, 1.76] | 1.92 |
| Sub-Saharan Africa | Tanzania | 4208 | 1.02 | [0.87, 1.19] | 6.22 |
| | **Overall** | | 1.05 | [0.96, 1.15] | |

$\tau^2 = 0.03$, $I^2 = 70.0\%$, $H^2 = 3.31$

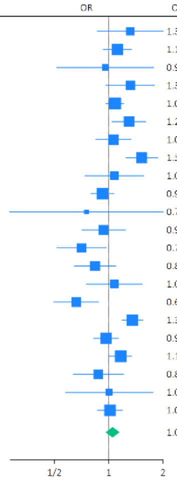

#### b) Feeling down, depressed or hopeless

| Region | Country | N | OR | 95% CI | Weight (%) |
|---|---|---|---|---|---|
| East Asia & Pacific | Australia | 2009 | 1.13 | [0.75, 1.72] | 2.42 |
| East Asia & Pacific | China | 3671 | 1.07 | [0.90, 1.27] | 6.46 |
| East Asia & Pacific | Hong Kong | 306 | 1.36 | [0.72, 2.56] | 1.23 |
| East Asia & Pacific | Indonesia | 1483 | 1.64 | [1.19, 2.28] | 3.45 |
| East Asia & Pacific | Japan | 8242 | 0.94 | [0.83, 1.06] | 7.97 |
| East Asia & Pacific | Philippines | 2060 | 1.33 | [1.07, 1.65] | 5.41 |
| Europe & Central Asia | Germany | 2923 | 1.15 | [0.89, 1.49] | 4.59 |
| Europe & Central Asia | Poland | 3180 | 1.47 | [1.15, 1.88] | 4.81 |
| Europe & Central Asia | Spain | 1547 | 0.79 | [0.53, 1.15] | 2.75 |
| Europe & Central Asia | Sweden | 8816 | 0.84 | [0.72, 0.98] | 7.09 |
| Europe & Central Asia | Türkiye | 253 | 0.87 | [0.32, 2.38] | 0.52 |
| Europe & Central Asia | United Kingdom | 2478 | 0.94 | [0.71, 1.24] | 4.19 |
| Latin America & Caribbean | Argentina | 1540 | 1.09 | [0.79, 1.49] | 3.57 |
| Latin America & Caribbean | Brazil | 2225 | 1.00 | [0.75, 1.32] | 4.17 |
| Latin America & Caribbean | Mexico | 1099 | 0.94 | [0.68, 1.28] | 3.61 |
| Middle East, North Africa, Afghanistan & Pakistan | Egypt | 2213 | 0.72 | [0.52, 0.99] | 3.56 |
| North America | United States | 22727 | 1.05 | [0.92, 1.20] | 7.63 |
| South Asia | India | 3852 | 0.98 | [0.83, 1.15] | 6.85 |
| Sub-Saharan Africa | Kenya | 6737 | 1.06 | [0.92, 1.22] | 7.34 |
| Sub-Saharan Africa | Nigeria | 2496 | 1.13 | [0.83, 1.54] | 3.72 |
| Sub-Saharan Africa | South Africa | 429 | 1.31 | [0.76, 2.24] | 1.62 |
| Sub-Saharan Africa | Tanzania | 4210 | 1.12 | [0.96, 1.30] | 7.03 |
| | **Overall** | | 1.05 | [0.98, 1.14] | |

$\tau^2 = 0.01$, $I^2 = 54.4\%$, $H^2 = 2.19$

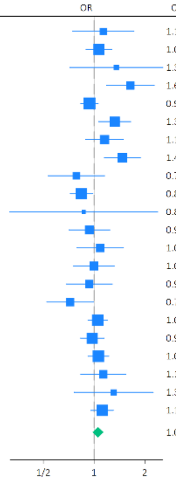

#### c) Unable to stop or control worrying

| Region | Country | N | OR | 95% CI | Weight (%) |
|---|---|---|---|---|---|
| East Asia & Pacific | Australia | 2007 | 1.06 | [0.72, 1.57] | 2.66 |
| East Asia & Pacific | China | 3671 | 1.05 | [0.88, 1.25] | 6.35 |
| East Asia & Pacific | Hong Kong | 309 | 0.80 | [0.41, 1.58] | 1.08 |
| East Asia & Pacific | Indonesia | 1484 | 1.31 | [0.95, 1.81] | 3.47 |
| East Asia & Pacific | Japan | 8300 | 1.04 | [0.91, 1.19] | 7.65 |
| East Asia & Pacific | Philippines | 2058 | 1.37 | [1.11, 1.68] | 5.63 |
| Europe & Central Asia | Germany | 2933 | 0.92 | [0.73, 1.17] | 4.98 |
| Europe & Central Asia | Poland | 3182 | 1.67 | [1.33, 2.10] | 5.18 |
| Europe & Central Asia | Spain | 1549 | 1.12 | [0.75, 1.65] | 2.66 |
| Europe & Central Asia | Sweden | 8810 | 0.87 | [0.74, 1.02] | 6.90 |
| Europe & Central Asia | Türkiye | 252 | 1.58 | [0.58, 4.25] | 0.54 |
| Europe & Central Asia | United Kingdom | 2485 | 0.90 | [0.67, 1.20] | 3.99 |
| Latin America & Caribbean | Argentina | 1537 | 1.06 | [0.77, 1.48] | 3.41 |
| Latin America & Caribbean | Brazil | 2222 | 0.97 | [0.75, 1.25] | 4.65 |
| Latin America & Caribbean | Mexico | 1094 | 0.85 | [0.59, 1.21] | 3.04 |
| Middle East, North Africa, Afghanistan & Pakistan | Egypt | 2209 | 0.79 | [0.59, 1.05] | 3.99 |
| North America | United States | 22765 | 1.07 | [0.94, 1.22] | 7.62 |
| South Asia | India | 3849 | 1.00 | [0.85, 1.17] | 6.86 |
| Sub-Saharan Africa | Kenya | 6739 | 1.08 | [0.94, 1.25] | 7.33 |
| Sub-Saharan Africa | Nigeria | 2487 | 0.96 | [0.69, 1.34] | 3.37 |
| Sub-Saharan Africa | South Africa | 429 | 0.92 | [0.55, 1.53] | 1.74 |
| Sub-Saharan Africa | Tanzania | 4201 | 1.11 | [0.95, 1.30] | 6.90 |
| | **Overall** | | 1.05 | [0.97, 1.13] | |

$\tau^2 = 0.01$, $I^2 = 53.8\%$, $H^2 = 2.16$

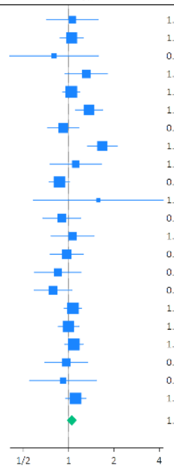

#### d) Little interest or pleasure in doing things

| Region | Country | N | OR | 95% CI | Weight (%) |
|---|---|---|---|---|---|
| East Asia & Pacific | Australia | 2008 | 0.70 | [0.46, 1.07] | 1.69 |
| East Asia & Pacific | China | 3670 | 0.94 | [0.80, 1.11] | 7.17 |
| East Asia & Pacific | Hong Kong | 309 | 0.69 | [0.41, 1.17] | 1.11 |
| East Asia & Pacific | Japan | 8278 | 0.95 | [0.85, 1.06] | 10.79 |
| East Asia & Pacific | Philippines | 2060 | 1.16 | [0.94, 1.42] | 5.39 |
| Europe & Central Asia | Germany | 2934 | 0.98 | [0.75, 1.28] | 3.74 |
| Europe & Central Asia | Poland | 3180 | 1.30 | [1.06, 1.59] | 5.47 |
| Europe & Central Asia | Spain | 1543 | 1.06 | [0.75, 1.50] | 2.41 |
| Europe & Central Asia | Sweden | 8763 | 0.95 | [0.82, 1.09] | 8.41 |
| Europe & Central Asia | Türkiye | 250 | 0.87 | [0.37, 2.08] | 0.42 |
| Europe & Central Asia | United Kingdom | 2481 | 0.93 | [0.70, 1.22] | 3.40 |
| Latin America & Caribbean | Argentina | 1538 | 1.14 | [0.83, 1.56] | 2.84 |
| Latin America & Caribbean | Brazil | 2218 | 1.10 | [0.85, 1.43] | 3.83 |
| Latin America & Caribbean | Mexico | 1089 | 0.80 | [0.57, 1.12] | 2.48 |
| Middle East, North Africa, Afghanistan & Pakistan | Egypt | 2203 | 0.87 | [0.65, 1.17] | 3.18 |
| North America | United States | 22735 | 0.96 | [0.85, 1.10] | 9.37 |
| South Asia | India | 3848 | 0.90 | [0.77, 1.05] | 7.86 |
| Sub-Saharan Africa | Kenya | 6740 | 1.16 | [1.01, 1.34] | 8.73 |
| Sub-Saharan Africa | Nigeria | 2489 | 1.09 | [0.78, 1.51] | 2.62 |
| Sub-Saharan Africa | South Africa | 430 | 0.91 | [0.51, 1.63] | 0.92 |
| Sub-Saharan Africa | Tanzania | 4203 | 1.10 | [0.95, 1.28] | 8.17 |
| | **Overall** | | 1.00 | [0.95, 1.06] | |

$\tau^2 = 0.00$, $I^2 = 29.6\%$, $H^2 = 1.42$

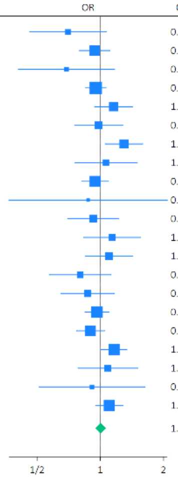

#### e) Suffering

| Region | Country | N | OR | 95% CI | Weight (%) |
|---|---|---|---|---|---|
| East Asia & Pacific | Australia | 2005 | 0.92 | [0.63, 1.34] | 1.88 |
| East Asia & Pacific | China | 3672 | 1.06 | [0.89, 1.26] | 6.65 |
| East Asia & Pacific | Hong Kong | 309 | 1.09 | [0.56, 2.14] | 0.65 |
| East Asia & Pacific | Indonesia | 1480 | 1.29 | [0.97, 1.71] | 3.16 |
| East Asia & Pacific | Japan | 8278 | 0.97 | [0.87, 1.07] | 11.11 |
| East Asia & Pacific | Philippines | 2060 | 1.05 | [0.84, 1.31] | 4.71 |
| Europe & Central Asia | Germany | 2931 | 1.03 | [0.81, 1.32] | 3.96 |
| Europe & Central Asia | Poland | 3189 | 1.41 | [1.15, 1.72] | 5.49 |
| Europe & Central Asia | Spain | 1535 | 0.88 | [0.61, 1.27] | 2.07 |
| Europe & Central Asia | Sweden | 8811 | 0.92 | [0.79, 1.05] | 8.47 |
| Europe & Central Asia | Türkiye | 251 | 0.43 | [0.16, 1.14] | 0.31 |
| Europe & Central Asia | United Kingdom | 2480 | 0.98 | [0.75, 1.28] | 3.53 |
| Latin America & Caribbean | Argentina | 1544 | 1.04 | [0.77, 1.41] | 2.84 |
| Latin America & Caribbean | Brazil | 2209 | 1.07 | [0.82, 1.40] | 3.45 |
| Latin America & Caribbean | Mexico | 1099 | 1.19 | [0.84, 1.68] | 2.28 |
| Middle East, North Africa, Afghanistan & Pakistan | Egypt | 2213 | 0.83 | [0.62, 1.11] | 2.95 |
| North America | United States | 22700 | 1.15 | [1.02, 1.30] | 9.98 |
| South Asia | India | 3850 | 1.02 | [0.87, 1.19] | 7.41 |
| Sub-Saharan Africa | Kenya | 6743 | 1.11 | [0.97, 1.28] | 8.46 |
| Sub-Saharan Africa | Nigeria | 2485 | 1.02 | [0.74, 1.41] | 2.50 |
| Sub-Saharan Africa | South Africa | 428 | 0.94 | [0.54, 1.63] | 0.96 |
| Sub-Saharan Africa | Tanzania | 4213 | 1.01 | [0.86, 1.19] | 7.19 |
| | **Overall** | | 1.04 | [0.99, 1.10] | |

$\tau^2 = 0.00$, $I^2 = 26.8\%$, $H^2 = 1.37$

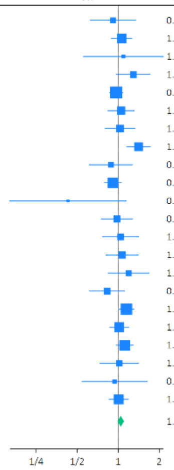

#### f) Feeling happy

| Region | Country | N | Coef | 95% CI | Weight (%) |
|---|---|---|---|---|---|
| East Asia & Pacific | Australia | 2007 | -0.05 | [-0.30, 0.20] | 3.63 |
| East Asia & Pacific | China | 3671 | 0.01 | [-0.14, 0.13] | 7.88 |
| East Asia & Pacific | Hong Kong | 309 | 0.22 | [-0.11, 0.55] | 2.33 |
| East Asia & Pacific | Indonesia | 1486 | -0.04 | [-0.36, 0.28] | 2.43 |
| East Asia & Pacific | Japan | 8260 | 0.18 | [0.10, 0.27] | 11.71 |
| East Asia & Pacific | Philippines | 2061 | -0.02 | [-0.25, 0.22] | 4.04 |
| Europe & Central Asia | Germany | 2933 | 0.24 | [0.07, 0.40] | 6.51 |
| Europe & Central Asia | Poland | 3197 | -0.04 | [-0.16, 0.09] | 8.65 |
| Europe & Central Asia | Spain | 1550 | 0.27 | [0.01, 0.54] | 3.29 |
| Europe & Central Asia | Sweden | 8834 | 0.13 | [0.03, 0.23] | 10.38 |
| Europe & Central Asia | Türkiye | 253 | 0.05 | [-1.04, 1.14] | 0.24 |
| Europe & Central Asia | United Kingdom | 2488 | 0.19 | [-0.01, 0.39] | 5.00 |
| Latin America & Caribbean | Argentina | 1545 | 0.11 | [-0.16, 0.38] | 3.15 |
| Latin America & Caribbean | Brazil | 2219 | 0.13 | [-0.13, 0.40] | 3.32 |
| Latin America & Caribbean | Mexico | 1102 | 0.28 | [0.01, 0.56] | 3.11 |
| Middle East, North Africa, Afghanistan & Pakistan | Egypt | 2211 | 0.36 | [-0.10, 0.81] | 1.31 |
| North America | United States | 22767 | 0.03 | [-0.08, 0.13] | 10.32 |
| South Asia | India | 3863 | 0.22 | [-0.05, 0.48] | 3.37 |
| Sub-Saharan Africa | Kenya | 6741 | 0.04 | [-0.19, 0.27] | 4.07 |
| Sub Saharan Africa | Nigeria | 2499 | 0.05 | [-0.52, 0.42] | 1.24 |
| Sub-Saharan Africa | South Africa | 431 | 0.10 | [-0.82, 1.01] | 0.34 |
| Sub-Saharan Africa | Tanzania | 4212 | 0.31 | [0.06, 0.56] | 3.68 |
| | **Overall** | | 0.11 | [0.06, 0.17] | |

$\tau^2 = 0.00$, $I^2 = 34.4$, $H^2 = 1.52$

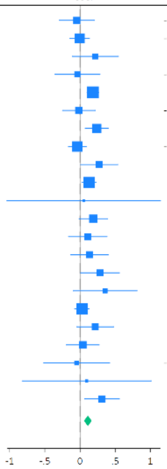

g) Understanding purpose in life

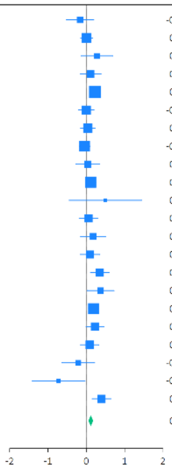

| Region | Country | N | Coef | 95% CI | Weight (%) |
|---|---|---|---|---|---|
| East Asia & Pacific | Australia | 2008 | -0.16 | [ -0.52, 0.19] | 2.47 |
| East Asia & Pacific | China | 3670 | 0.01 | [ -0.15, 0.17] | 7.05 |
| East Asia & Pacific | Hong Kong | 309 | 0.28 | [ -0.13, 0.69] | 1.94 |
| East Asia & Pacific | Indonesia | 1487 | 0.11 | [ -0.16, 0.38] | 3.69 |
| East Asia & Pacific | Japan | 8262 | 0.23 | [ 0.14, 0.32] | 10.42 |
| East Asia & Pacific | Philippines | 2060 | -0.00 | [ -0.20, 0.20] | 5.59 |
| Europe & Central Asia | Germany | 2931 | 0.04 | [ -0.15, 0.23] | 5.99 |
| Europe & Central Asia | Poland | 3187 | -0.04 | [ -0.18, 0.10] | 7.98 |
| Europe & Central Asia | Spain | 1543 | 0.04 | [ -0.27, 0.34] | 3.10 |
| Europe & Central Asia | Sweden | 8766 | 0.12 | [ -0.01, 0.24] | 8.57 |
| Europe & Central Asia | Türkiye | 252 | 0.50 | [ -0.45, 1.44] | 0.42 |
| Europe & Central Asia | United Kingdom | 2478 | 0.06 | [ -0.18, 0.30] | 4.39 |
| Latin America & Caribbean | Argentina | 1540 | 0.18 | [ -0.15, 0.51] | 2.78 |
| Latin America & Caribbean | Brazil | 2216 | 0.09 | [ -0.16, 0.35] | 4.04 |
| Latin America & Caribbean | Mexico | 1099 | 0.35 | [ 0.11, 0.59] | 4.42 |
| Middle East, North Africa, Afghanistan & Pakistan | Egypt | 2193 | 0.37 | [ 0.02, 0.72] | 2.54 |
| North America | United States | 22735 | 0.19 | [ 0.06, 0.32] | 8.52 |
| South Asia | India | 3849 | 0.23 | [ -0.00, 0.46] | 4.66 |
| Sub-Saharan Africa | Kenya | 6740 | 0.09 | [ -0.15, 0.32] | 4.51 |
| Sub-Saharan Africa | Nigeria | 2494 | -0.21 | [ -0.64, 0.22] | 1.80 |
| Sub-Saharan Africa | South Africa | 431 | -0.73 | [ -1.42, -0.04] | 0.76 |
| Sub-Saharan Africa | Tanzania | 4176 | 0.39 | [ 0.15, 0.64] | 4.38 |
| | **Overall** | | 0.12 | [ 0.05, 0.18] | |

$\tau^2 = 0.01$, $I^2 = 40.5\%$, $H^2 = 1.68$

h) Life satisfaction

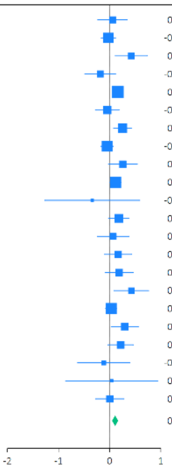

| Region | Country | N | Coef | 95% CI | Weight (%) |
|---|---|---|---|---|---|
| East Asia & Pacific | Australia | 2010 | 0.06 | [ -0.23, 0.35] | 3.22 |
| East Asia & Pacific | China | 3652 | -0.03 | [ -0.16, 0.11] | 7.76 |
| East Asia & Pacific | Hong Kong | 297 | 0.42 | [ 0.11, 0.74] | 2.82 |
| East Asia & Pacific | Indonesia | 1484 | -0.19 | [ -0.49, 0.12] | 3.04 |
| East Asia & Pacific | Japan | 8226 | 0.16 | [ 0.08, 0.24] | 10.68 |
| East Asia & Pacific | Philippines | 2059 | -0.05 | [ -0.28, 0.19] | 4.40 |
| Europe & Central Asia | Germany | 2913 | 0.25 | [ 0.08, 0.42] | 6.30 |
| Europe & Central Asia | Poland | 3180 | -0.05 | [ -0.17, 0.07] | 8.62 |
| Europe & Central Asia | Spain | 1542 | 0.26 | [ -0.03, 0.54] | 3.34 |
| Europe & Central Asia | Sweden | 8822 | 0.11 | [ 0.01, 0.22] | 9.45 |
| Europe & Central Asia | Türkiye | 253 | -0.34 | [ -1.27, 0.59] | 0.40 |
| Europe & Central Asia | United Kingdom | 2459 | 0.18 | [ -0.02, 0.38] | 5.37 |
| Latin America & Caribbean | Argentina | 1544 | 0.07 | [ -0.24, 0.38] | 2.92 |
| Latin America & Caribbean | Brazil | 2219 | 0.17 | [ -0.10, 0.43] | 3.68 |
| Latin America & Caribbean | Mexico | 1102 | 0.19 | [ -0.09, 0.46] | 3.47 |
| Middle East, North Africa, Afghanistan & Pakistan | Egypt | 2199 | 0.43 | [ 0.09, 0.76] | 2.53 |
| North America | United States | 22756 | 0.03 | [ -0.08, 0.14] | 9.26 |
| South Asia | India | 3859 | 0.29 | [ 0.03, 0.56] | 3.66 |
| Sub-Saharan Africa | Kenya | 6744 | 0.21 | [ -0.04, 0.46] | 3.96 |
| Sub-Saharan Africa | Nigeria | 2499 | -0.11 | [ -0.63, 0.40] | 1.22 |
| Sub-Saharan Africa | South Africa | 426 | 0.04 | [ -0.86, 0.94] | 0.42 |
| Sub-Saharan Africa | Tanzania | 4211 | 0.00 | [ -0.27, 0.28] | 3.45 |
| | **Overall** | | 0.11 | [ 0.05, 0.17] | |

$\tau^2 = 0.01$, $I^2 = 42.1\%$, $H^2 = 1.73$

i) Life evaluation: Today

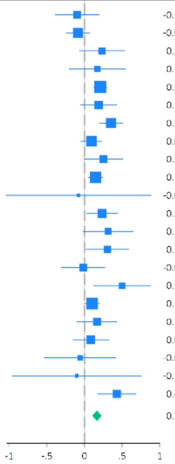

| Region | Country | N | Coef | 95% CI | Weight (%) |
|---|---|---|---|---|---|
| East Asia & Pacific | Australia | 2011 | -0.10 | [ -0.39, 0.19] | 3.41 |
| East Asia & Pacific | China | 3671 | -0.09 | [ -0.24, 0.07] | 6.98 |
| East Asia & Pacific | Hong Kong | 309 | 0.23 | [ -0.07, 0.53] | 3.23 |
| East Asia & Pacific | Indonesia | 1485 | 0.17 | [ -0.20, 0.55] | 2.28 |
| East Asia & Pacific | Japan | 8293 | 0.21 | [ 0.12, 0.29] | 9.94 |
| East Asia & Pacific | Philippines | 2056 | 0.19 | [ -0.05, 0.43] | 4.41 |
| Europe & Central Asia | Germany | 2928 | 0.35 | [ 0.20, 0.51] | 7.03 |
| Europe & Central Asia | Poland | 3195 | 0.09 | [ -0.05, 0.23] | 7.56 |
| Europe & Central Asia | Spain | 1550 | 0.25 | [ -0.00, 0.50] | 4.09 |
| Europe & Central Asia | Sweden | 8831 | 0.15 | [ 0.05, 0.24] | 9.41 |
| Europe & Central Asia | Türkiye | 252 | -0.08 | [ -1.04, 0.88] | 0.41 |
| Europe & Central Asia | United Kingdom | 2485 | 0.23 | [ 0.03, 0.43] | 5.40 |
| Latin America & Caribbean | Argentina | 1547 | 0.31 | [ -0.02, 0.64] | 2.77 |
| Latin America & Caribbean | Brazil | 2226 | 0.30 | [ 0.02, 0.58] | 3.54 |
| Latin America & Caribbean | Mexico | 1102 | -0.02 | [ -0.30, 0.27] | 3.45 |
| Middle East, North Africa, Afghanistan & Pakistan | Egypt | 2201 | 0.50 | [ 0.12, 0.88] | 2.27 |
| North America | United States | 22777 | 0.10 | [ 0.00, 0.20] | 9.36 |
| South Asia | India | 3842 | 0.17 | [ -0.10, 0.43] | 3.89 |
| Sub-Saharan Africa | Kenya | 6744 | 0.08 | [ -0.15, 0.32] | 4.47 |
| Sub-Saharan Africa | Nigeria | 2503 | -0.06 | [ -0.53, 0.42] | 1.52 |
| Sub-Saharan Africa | South Africa | 429 | -0.11 | [ -0.96, 0.75] | 0.52 |
| Sub-Saharan Africa | Tanzania | 4215 | 0.43 | [ 0.18, 0.68] | 4.06 |
| | **Overall** | | 0.16 | [ 0.10, 0.23] | |

$\tau^2 = 0.01$, $I^2 = 48.2\%$, $H^2 = 1.93$

j) Life evaluation: Five years from now

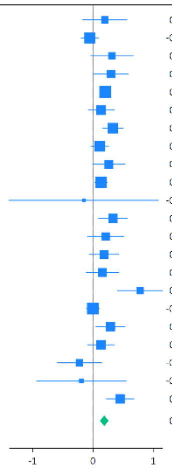

| Region | Country | N | Coef | 95% CI | Weight (%) |
|---|---|---|---|---|---|
| East Asia & Pacific | Australia | 2010 | 0.20 | [ -0.17, 0.56] | 2.76 |
| East Asia & Pacific | China | 3653 | -0.05 | [ -0.19, 0.09] | 6.84 |
| East Asia & Pacific | Hong Kong | 300 | 0.32 | [ -0.03, 0.67] | 2.92 |
| East Asia & Pacific | Indonesia | 1467 | 0.30 | [ 0.01, 0.58] | 3.77 |
| East Asia & Pacific | Japan | 8297 | 0.20 | [ 0.12, 0.29] | 8.16 |
| East Asia & Pacific | Philippines | 2057 | 0.14 | [ -0.07, 0.35] | 5.16 |
| Europe & Central Asia | Germany | 2912 | 0.33 | [ 0.17, 0.50] | 6.23 |
| Europe & Central Asia | Poland | 3153 | 0.11 | [ -0.03, 0.26] | 6.73 |
| Europe & Central Asia | Spain | 1540 | 0.26 | [ -0.00, 0.53] | 4.08 |
| Europe & Central Asia | Sweden | 8806 | 0.14 | [ 0.03, 0.24] | 7.86 |
| Europe & Central Asia | Türkiye | 236 | -0.15 | [ -1.39, 1.08] | 0.33 |
| Europe & Central Asia | United Kingdom | 2460 | 0.33 | [ 0.09, 0.57] | 4.64 |
| Latin America & Caribbean | Argentina | 1510 | 0.21 | [ -0.09, 0.51] | 3.55 |
| Latin America & Caribbean | Brazil | 2216 | 0.18 | [ -0.06, 0.43] | 4.53 |
| Latin America & Caribbean | Mexico | 1078 | 0.16 | [ -0.11, 0.43] | 4.09 |
| Middle East, North Africa, Afghanistan & Pakistan | Egypt | 1632 | 0.78 | [ 0.41, 1.16] | 2.67 |
| North America | United States | 22765 | -0.00 | [ -0.11, 0.11] | 7.71 |
| South Asia | India | 3576 | 0.29 | [ 0.05, 0.53] | 4.61 |
| Sub-Saharan Africa | Kenya | 6699 | 0.13 | [ -0.09, 0.36] | 4.94 |
| Sub-Saharan Africa | Nigeria | 2470 | -0.22 | [ -0.59, 0.14] | 2.76 |
| Sub-Saharan Africa | South Africa | 423 | -0.19 | [ -0.93, 0.55] | 0.86 |
| Sub-Saharan Africa | Tanzania | 3979 | 0.45 | [ 0.22, 0.68] | 4.80 |
| | **Overall** | | 0.19 | [ 0.12, 0.26] | |

$\tau^2 = 0.01$, $I^2 = 60.7\%$, $H^2 = 2.54$

k) Feeling lonely

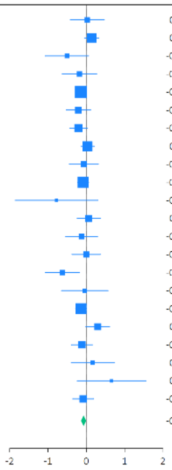

| Region | Country | N | Coef | 95% CI | Weight (%) |
|---|---|---|---|---|---|
| East Asia & Pacific | Australia | 2010 | 0.03 | [ -0.42, 0.47] | 2.08 |
| East Asia & Pacific | China | 3672 | 0.14 | [ -0.05, 0.33] | 8.10 |
| East Asia & Pacific | Hong Kong | 308 | -0.50 | [ -1.07, 0.06] | 1.33 |
| East Asia & Pacific | Indonesia | 1487 | -0.18 | [ -0.63, 0.27] | 2.04 |
| East Asia & Pacific | Japan | 8309 | -0.14 | [ -0.26, -0.02] | 13.83 |
| East Asia & Pacific | Philippines | 2063 | -0.21 | [ -0.52, 0.11] | 3.77 |
| Europe & Central Asia | Germany | 2938 | -0.20 | [ -0.44, 0.03] | 6.08 |
| Europe & Central Asia | Poland | 3190 | 0.03 | [ -0.14, 0.21] | 9.11 |
| Europe & Central Asia | Spain | 1553 | -0.07 | [ -0.44, 0.31] | 2.79 |
| Europe & Central Asia | Sweden | 8848 | -0.09 | [ -0.23, 0.06] | 11.34 |
| Europe & Central Asia | Türkiye | 250 | -0.78 | [ -1.86, 0.30] | 0.38 |
| Europe & Central Asia | United Kingdom | 2476 | 0.06 | [ -0.24, 0.36] | 4.08 |
| Latin America & Caribbean | Argentina | 1549 | -0.12 | [ -0.54, 0.29] | 2.33 |
| Latin America & Caribbean | Brazil | 2221 | -0.00 | [ -0.38, 0.37] | 2.82 |
| Latin America & Caribbean | Mexico | 1099 | -0.63 | [ -1.07, -0.19] | 2.12 |
| Middle East, North Africa, Afghanistan & Pakistan | Egypt | 2210 | -0.04 | [ -0.65, 0.57] | 1.16 |
| North America | United States | 22786 | -0.14 | [ -0.28, 0.01] | 11.26 |
| South Asia | India | 3863 | 0.30 | [ -0.01, 0.61] | 3.90 |
| Sub-Saharan Africa | Kenya | 6739 | -0.12 | [ -0.39, 0.16] | 4.76 |
| Sub-Saharan Africa | Nigeria | 2496 | 0.17 | [ -0.39, 0.73] | 1.34 |
| Sub-Saharan Africa | South Africa | 431 | 0.66 | [ -0.24, 1.56] | 0.55 |
| Sub-Saharan Africa | Tanzania | 4213 | -0.08 | [ -0.35, 0.19] | 4.84 |
| | **Overall** | | -0.07 | [ -0.14, -0.00] | |

$\tau^2 = 0.01$, $I^2 = 22.4\%$, $H^2 = 1.29$

l) Life is worthwhile

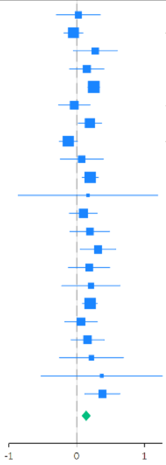

| Region | Country | N | Coef | 95% CI | Weight (%) |
|---|---|---|---|---|---|
| East Asia & Pacific | Australia | 2008 | 0.02 | [ -0.30, 0.35] | 3.04 |
| East Asia & Pacific | China | 3669 | -0.05 | [ -0.18, 0.09] | 7.56 |
| East Asia & Pacific | Hong Kong | 306 | 0.28 | [ -0.05, 0.60] | 3.04 |
| East Asia & Pacific | Indonesia | 1486 | 0.15 | [ -0.10, 0.40] | 4.25 |
| East Asia & Pacific | Japan | 8290 | 0.25 | [ 0.17, 0.34] | 9.50 |
| East Asia & Pacific | Philippines | 2060 | -0.04 | [ -0.27, 0.20] | 4.73 |
| Europe & Central Asia | Germany | 2932 | 0.20 | [ 0.03, 0.37] | 6.45 |
| Europe & Central Asia | Poland | 3189 | -0.12 | [ -0.26, 0.01] | 7.68 |
| Europe & Central Asia | Spain | 1541 | 0.07 | [ -0.24, 0.39] | 3.11 |
| Europe & Central Asia | Sweden | 8832 | 0.20 | [ 0.08, 0.32] | 8.29 |
| Europe & Central Asia | Türkiye | 253 | 0.17 | [ -0.86, 1.19] | 0.40 |
| Europe & Central Asia | United Kingdom | 2473 | 0.10 | [ -0.11, 0.30] | 5.40 |
| Latin America & Caribbean | Argentina | 1541 | 0.19 | [ -0.10, 0.49] | 3.51 |
| Latin America & Caribbean | Brazil | 2224 | 0.31 | [ 0.05, 0.57] | 4.07 |
| Latin America & Caribbean | Mexico | 1101 | 0.18 | [ -0.12, 0.49] | 3.32 |
| Middle East, North Africa, Afghanistan & Pakistan | Egypt | 2207 | 0.21 | [ -0.22, 0.64] | 1.94 |
| North America | United States | 22770 | 0.20 | [ 0.09, 0.31] | 8.60 |
| South Asia | India | 3844 | 0.06 | [ -0.18, 0.31] | 4.46 |
| Sub-Saharan Africa | Kenya | 6743 | 0.16 | [ -0.09, 0.41] | 4.37 |
| Sub-Saharan Africa | Nigeria | 2498 | 0.22 | [ -0.25, 0.69] | 1.68 |
| Sub-Saharan Africa | South Africa | 428 | 0.37 | [ -0.53, 1.26] | 0.52 |
| Sub-Saharan Africa | Tanzania | 4209 | 0.38 | [ 0.12, 0.64] | 4.09 |
| | **Overall** | | 0.14 | [ 0.07, 0.21] | |

$\tau^2 = 0.01$, $I^2 = 50.0\%$, $H^2 = 2.00$

m) Feeling grateful

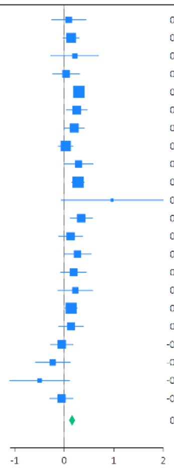

| Region | Country | N | Coef | 95% CI | Weight (%) |
|---|---|---|---|---|---|
| East Asia & Pacific | Australia | 1930 | 0.09 | [ -0.25, 0.43] | 2.54 |
| East Asia & Pacific | China | 3670 | 0.14 | [ -0.02, 0.30] | 6.98 |
| East Asia & Pacific | Hong Kong | 308 | 0.21 | [ -0.27, 0.69] | 1.43 |
| East Asia & Pacific | Indonesia | 1482 | 0.04 | [ -0.23, 0.31] | 3.72 |
| East Asia & Pacific | Japan | 8240 | 0.30 | [ 0.20, 0.39] | 10.15 |
| East Asia & Pacific | Philippines | 2061 | 0.25 | [ 0.04, 0.46] | 5.15 |
| Europe & Central Asia | Germany | 2921 | 0.20 | [ -0.00, 0.41] | 5.33 |
| Europe & Central Asia | Poland | 3189 | 0.03 | [ -0.12, 0.18] | 7.33 |
| Europe & Central Asia | Spain | 1547 | 0.29 | [ -0.00, 0.58] | 3.32 |
| Europe & Central Asia | Sweden | 8836 | 0.28 | [ 0.16, 0.40] | 8.81 |
| Europe & Central Asia | Türkiye | 246 | 0.97 | [ -0.06, 1.99] | 0.34 |
| Europe & Central Asia | United Kingdom | 2445 | 0.35 | [ 0.12, 0.57] | 4.80 |
| Latin America & Caribbean | Argentina | 1543 | 0.13 | [ -0.11, 0.37] | 4.37 |
| Latin America & Caribbean | Brazil | 2215 | 0.27 | [ -0.00, 0.54] | 3.68 |
| Latin America & Caribbean | Mexico | 1101 | 0.19 | [ -0.07, 0.45] | 3.95 |
| Middle East, North Africa, Afghanistan & Pakistan | Egypt | 2181 | 0.22 | [ -0.13, 0.57] | 2.48 |
| North America | United States | 21475 | 0.14 | [ 0.02, 0.26] | 8.76 |
| South Asia | India | 3837 | 0.14 | [ -0.11, 0.38] | 4.21 |
| Sub-Saharan Africa | Kenya | 6744 | -0.05 | [ -0.28, 0.17] | 4.69 |
| Sub-Saharan Africa | Nigeria | 2490 | -0.23 | [ -0.58, 0.12] | 2.49 |
| Sub-Saharan Africa | South Africa | 428 | -0.49 | [ -1.10, 0.11] | 0.95 |
| Sub-Saharan Africa | Tanzania | 4197 | -0.05 | [ -0.29, 0.18] | 4.52 |
|  | **Overall** |  | 0.16 | [ 0.10, 0.22] |  |

$\tau^2 = 0.01$, $I^2 = 39.7\%$, $H^2 = 1.66$

n) Feeling at peace

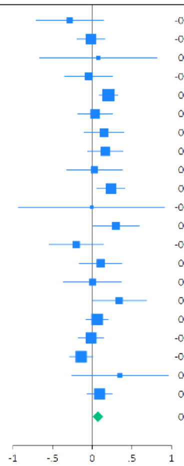

| Region | Country | N | Coef | 95% CI | Weight (%) |
|---|---|---|---|---|---|
| East Asia & Pacific | Australia | 2007 | -0.28 | [ -0.70, 0.14] | 2.06 |
| East Asia & Pacific | China | 3669 | -0.02 | [ -0.19, 0.16] | 7.09 |
| East Asia & Pacific | Hong Kong | 309 | 0.07 | [ -0.66, 0.81] | 0.75 |
| East Asia & Pacific | Indonesia | 1485 | -0.05 | [ -0.35, 0.25] | 3.59 |
| East Asia & Pacific | Japan | 8263 | 0.20 | [ 0.09, 0.32] | 9.98 |
| East Asia & Pacific | Philippines | 2061 | 0.04 | [ -0.18, 0.26] | 5.52 |
| Europe & Central Asia | Germany | 2923 | 0.15 | [ -0.10, 0.40] | 4.62 |
| Europe & Central Asia | Poland | 3166 | 0.16 | [ -0.06, 0.38] | 5.48 |
| Europe & Central Asia | Spain | 1541 | 0.03 | [ -0.32, 0.38] | 2.80 |
| Europe & Central Asia | Sweden | 8824 | 0.24 | [ 0.06, 0.41] | 7.15 |
| Europe & Central Asia | Türkiye | 253 | -0.01 | [ -0.93, 0.91] | 0.49 |
| Europe & Central Asia | United Kingdom | 2479 | 0.30 | [ 0.01, 0.59] | 3.74 |
| Latin America & Caribbean | Argentina | 1538 | -0.20 | [ -0.54, 0.14] | 2.94 |
| Latin America & Caribbean | Brazil | 2219 | 0.11 | [ -0.16, 0.37] | 4.21 |
| Latin America & Caribbean | Mexico | 1096 | 0.00 | [ -0.36, 0.37] | 2.62 |
| Middle East, North Africa, Afghanistan & Pakistan | Egypt | 2210 | 0.34 | [ -0.00, 0.68] | 2.93 |
| North America | United States | 22702 | 0.06 | [ -0.08, 0.20] | 8.71 |
| South Asia | India | 3845 | -0.01 | [ -0.17, 0.14] | 7.83 |
| Sub-Saharan Africa | Kenya | 6743 | -0.14 | [ -0.28, 0.01] | 8.46 |
| Sub-Saharan Africa | South Africa | 430 | 0.35 | [ -0.26, 0.96] | 1.08 |
| Sub-Saharan Africa | Tanzania | 4215 | 0.09 | [ -0.06, 0.25] | 7.96 |
|  | **Overall** |  | 0.07 | [ 0.01, 0.14] |  |

$\tau^2 = 0.01$, $I^2 = 38.2\%$, $H^2 = 1.62$

o) Remain hopeful about the future

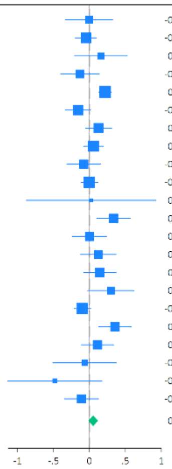

| Region | Country | N | Coef | 95% CI | Weight (%) |
|---|---|---|---|---|---|
| East Asia & Pacific | Australia | 1934 | -0.01 | [ -0.33, 0.32] | 3.01 |
| East Asia & Pacific | China | 3673 | -0.05 | [ -0.19, 0.10] | 6.55 |
| East Asia & Pacific | Hong Kong | 309 | 0.16 | [ -0.21, 0.52] | 2.57 |
| East Asia & Pacific | Indonesia | 1484 | -0.13 | [ -0.40, 0.14] | 3.89 |
| East Asia & Pacific | Japan | 8266 | 0.22 | [ 0.13, 0.31] | 8.06 |
| East Asia & Pacific | Philippines | 2062 | -0.16 | [ -0.33, 0.01] | 5.85 |
| Europe & Central Asia | Germany | 2928 | 0.13 | [ -0.05, 0.31] | 5.67 |
| Europe & Central Asia | Poland | 3195 | 0.06 | [ -0.08, 0.19] | 6.83 |
| Europe & Central Asia | Spain | 1546 | -0.08 | [ -0.31, 0.15] | 4.48 |
| Europe & Central Asia | Sweden | 8837 | -0.00 | [ -0.12, 0.11] | 7.36 |
| Europe & Central Asia | Türkiye | 251 | 0.02 | [ -0.88, 0.92] | 0.55 |
| Europe & Central Asia | United Kingdom | 2446 | 0.34 | [ 0.10, 0.57] | 4.50 |
| Latin America & Caribbean | Argentina | 1542 | 0.00 | [ -0.24, 0.24] | 4.45 |
| Latin America & Caribbean | Brazil | 2219 | 0.12 | [ -0.12, 0.37] | 4.27 |
| Latin America & Caribbean | Mexico | 1101 | 0.14 | [ -0.08, 0.37] | 4.67 |
| Middle East, North Africa, Afghanistan & Pakistan | Egypt | 2198 | 0.30 | [ -0.02, 0.62] | 3.07 |
| North America | United States | 21460 | -0.10 | [ -0.21, 0.02] | 7.41 |
| South Asia | India | 3854 | 0.36 | [ 0.13, 0.58] | 4.67 |
| Sub-Saharan Africa | Kenya | 6740 | 0.11 | [ -0.11, 0.33] | 4.74 |
| Sub-Saharan Africa | Nigeria | 2498 | -0.07 | [ -0.51, 0.38] | 1.93 |
| Sub-Saharan Africa | South Africa | 429 | -0.48 | [ -1.13, 0.17] | 0.99 |
| Sub-Saharan Africa | Tanzania | 4212 | -0.11 | [ -0.35, 0.12] | 4.46 |
|  | **Overall** |  | 0.05 | [ -0.02, 0.12] |  |

$\tau^2 = 0.01$, $I^2 = 58.6\%$, $H^2 = 2.41$

p) Feeling very capable in most things

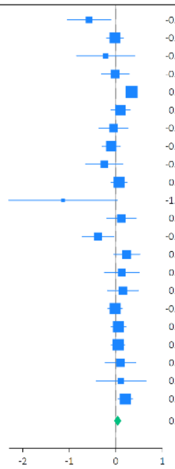

| Region | Country | N | Coef | 95% CI | Weight (%) |
|---|---|---|---|---|---|
| East Asia & Pacific | Australia | 1994 | -0.58 | [ -1.04, -0.11] | 2.17 |
| East Asia & Pacific | China | 3667 | -0.02 | [ -0.20, 0.16] | 6.47 |
| East Asia & Pacific | Hong Kong | 309 | -0.22 | [ -0.84, 0.40] | 1.34 |
| East Asia & Pacific | Indonesia | 1481 | -0.01 | [ -0.31, 0.28] | 4.06 |
| East Asia & Pacific | Japan | 8227 | 0.34 | [ 0.23, 0.45] | 8.24 |
| East Asia & Pacific | Philippines | 2063 | 0.10 | [ -0.11, 0.31] | 5.85 |
| Europe & Central Asia | Germany | 2924 | -0.05 | [ -0.36, 0.26] | 3.84 |
| Europe & Central Asia | Poland | 3151 | -0.10 | [ -0.30, 0.09] | 6.12 |
| Europe & Central Asia | Spain | 1530 | -0.25 | [ -0.65, 0.15] | 2.72 |
| Europe & Central Asia | Sweden | 8791 | 0.07 | [ -0.10, 0.24] | 6.67 |
| Europe & Central Asia | Türkiye | 252 | -1.14 | [ -2.30, 0.03] | 0.43 |
| Europe & Central Asia | United Kingdom | 2466 | 0.12 | [ -0.20, 0.44] | 3.75 |
| Latin America & Caribbean | Argentina | 1543 | -0.38 | [ -0.73, -0.04] | 3.40 |
| Latin America & Caribbean | Brazil | 2208 | 0.24 | [ -0.05, 0.52] | 4.31 |
| Latin America & Caribbean | Mexico | 1094 | 0.13 | [ -0.24, 0.50] | 3.03 |
| Middle East, North Africa, Afghanistan & Pakistan | Egypt | 2209 | 0.15 | [ -0.18, 0.48] | 3.55 |
| North America | United States | 22625 | -0.02 | [ -0.17, 0.14] | 7.16 |
| South Asia | India | 3858 | 0.05 | [ -0.11, 0.21] | 6.98 |
| Sub-Saharan Africa | Kenya | 6735 | 0.05 | [ -0.10, 0.19] | 7.43 |
| Sub-Saharan Africa | Nigeria | 2492 | 0.10 | [ -0.23, 0.43] | 3.61 |
| Sub-Saharan Africa | South Africa | 430 | 0.11 | [ -0.43, 0.65] | 1.74 |
| Sub-Saharan Africa | Tanzania | 4212 | 0.21 | [ 0.05, 0.36] | 7.14 |
|  | **Overall** |  | 0.04 | [ -0.04, 0.12] |  |

$\tau^2 = 0.02$, $I^2 = 55.2\%$, $H^2 = 2.23$

q) Expect more good things to happen to you

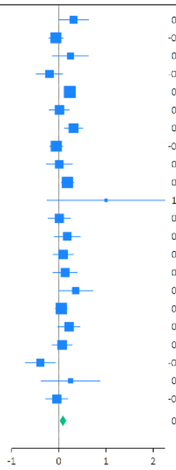

| Region | Country | N | Coef | 95% CI | Weight (%) |
|---|---|---|---|---|---|
| East Asia & Pacific | Australia | 1988 | 0.31 | [ -0.01, 0.63] | 3.17 |
| East Asia & Pacific | China | 3673 | -0.06 | [ -0.21, 0.09] | 6.69 |
| East Asia & Pacific | Hong Kong | 308 | 0.24 | [ -0.13, 0.62] | 2.48 |
| East Asia & Pacific | Indonesia | 1487 | -0.20 | [ -0.48, 0.08] | 3.75 |
| East Asia & Pacific | Japan | 8308 | 0.23 | [ 0.13, 0.33] | 8.19 |
| East Asia & Pacific | Philippines | 2061 | 0.01 | [ -0.20, 0.22] | 5.10 |
| Europe & Central Asia | Germany | 2929 | 0.31 | [ 0.12, 0.50] | 5.54 |
| Europe & Central Asia | Poland | 3191 | -0.05 | [ -0.19, 0.09] | 7.04 |
| Europe & Central Asia | Spain | 1548 | 0.01 | [ -0.26, 0.28] | 3.83 |
| Europe & Central Asia | Sweden | 8827 | 0.18 | [ 0.05, 0.31] | 7.35 |
| Europe & Central Asia | Türkiye | 253 | 1.00 | [ -0.25, 2.25] | 0.29 |
| Europe & Central Asia | United Kingdom | 2470 | 0.01 | [ -0.23, 0.24] | 4.57 |
| Latin America & Caribbean | Argentina | 1545 | 0.18 | [ -0.09, 0.45] | 3.85 |
| Latin America & Caribbean | Brazil | 2221 | 0.09 | [ -0.12, 0.31] | 5.00 |
| Latin America & Caribbean | Mexico | 1103 | 0.13 | [ -0.12, 0.38] | 4.21 |
| Middle East, North Africa, Afghanistan & Pakistan | Egypt | 2161 | 0.36 | [ -0.00, 0.72] | 2.65 |
| North America | United States | 22673 | 0.05 | [ -0.07, 0.17] | 7.49 |
| South Asia | India | 3854 | 0.22 | [ -0.02, 0.45] | 4.63 |
| Sub-Saharan Africa | Kenya | 6747 | 0.07 | [ -0.14, 0.27] | 5.14 |
| Sub-Saharan Africa | Nigeria | 2494 | -0.39 | [ -0.70, -0.08] | 3.25 |
| Sub-Saharan Africa | South Africa | 427 | 0.25 | [ -0.37, 0.87] | 1.09 |
| Sub-Saharan Africa | Tanzania | 4211 | -0.05 | [ -0.27, 0.18] | 4.71 |
|  | **Overall** |  | 0.09 | [ 0.02, 0.15] |  |

$\tau^2 = 0.01$, $I^2 = 54.5\%$, $H^2 = 2.20$

r) Overall mental health

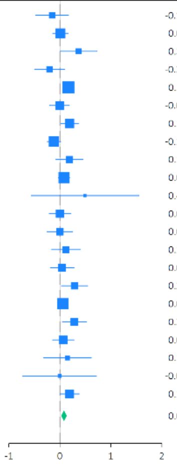

| Region | Country | N | Coef | 95% CI | Weight (%) |
|---|---|---|---|---|---|
| East Asia & Pacific | Australia | 2009 | -0.15 | [ -0.47, 0.17] | 2.41 |
| East Asia & Pacific | China | 3670 | 0.01 | [ -0.14, 0.16] | 6.90 |
| East Asia & Pacific | Hong Kong | 307 | 0.37 | [ 0.01, 0.73] | 2.00 |
| East Asia & Pacific | Indonesia | 1486 | -0.20 | [ -0.50, 0.09] | 2.75 |
| East Asia & Pacific | Japan | 8320 | 0.17 | [ 0.08, 0.25] | 10.48 |
| East Asia & Pacific | Philippines | 2062 | -0.01 | [ -0.20, 0.18] | 5.18 |
| Europe & Central Asia | Germany | 2931 | 0.19 | [ 0.02, 0.36] | 5.87 |
| Europe & Central Asia | Poland | 3196 | -0.11 | [ -0.24, 0.01] | 7.89 |
| Europe & Central Asia | Spain | 1546 | 0.19 | [ -0.08, 0.45] | 3.29 |
| Europe & Central Asia | Sweden | 8855 | 0.08 | [ -0.03, 0.19] | 9.11 |
| Europe & Central Asia | Türkiye | 252 | 0.49 | [ -0.56, 1.55] | 0.26 |
| Europe & Central Asia | United Kingdom | 2465 | 0.00 | [ -0.21, 0.21] | 4.58 |
| Latin America & Caribbean | Argentina | 1546 | 0.00 | [ -0.25, 0.25] | 3.60 |
| Latin America & Caribbean | Brazil | 2225 | 0.12 | [ -0.16, 0.40] | 3.04 |
| Latin America & Caribbean | Mexico | 1100 | 0.05 | [ -0.18, 0.28] | 4.01 |
| Middle East, North Africa, Afghanistan & Pakistan | Egypt | 2199 | 0.29 | [ 0.03, 0.55] | 3.42 |
| North America | United States | 22777 | 0.06 | [ -0.04, 0.16] | 9.44 |
| South Asia | India | 3857 | 0.29 | [ 0.05, 0.52] | 3.92 |
| Sub-Saharan Africa | Kenya | 6738 | 0.07 | [ -0.14, 0.28] | 4.62 |
| Sub-Saharan Africa | Nigeria | 2500 | 0.15 | [ -0.32, 0.61] | 1.25 |
| Sub-Saharan Africa | South Africa | 429 | -0.01 | [ -0.73, 0.72] | 0.55 |
| Sub-Saharan Africa | Tanzania | 4201 | 0.19 | [ 0.01, 0.38] | 5.39 |
|  | **Overall** |  | 0.08 | [ 0.03, 0.14] |  |

$\tau^2 = 0.01$, $I^2 = 38.7\%$, $H^2 = 1.63$

s) Overall physical health

| Region | Country | N | Coef | 95% CI | Weight (%) |
|---|---|---|---|---|---|
| East Asia & Pacific | Australia | 2010 | -0.10 | [ -0.41, 0.22] | 3.11 |
| East Asia & Pacific | China | 3673 | 0.07 | [ -0.07, 0.21] | 6.82 |
| East Asia & Pacific | Hong Kong | 309 | 0.10 | [ -0.24, 0.45] | 2.71 |
| East Asia & Pacific | Indonesia | 1487 | 0.19 | [ -0.11, 0.49] | 3.34 |
| East Asia & Pacific | Japan | 8289 | 0.20 | [ 0.11, 0.28] | 8.51 |
| East Asia & Pacific | Philippines | 2062 | -0.01 | [ -0.20, 0.18] | 5.38 |
| Europe & Central Asia | Germany | 2940 | 0.33 | [ 0.18, 0.48] | 6.59 |
| Europe & Central Asia | Poland | 3195 | -0.13 | [ -0.26, -0.01] | 7.32 |
| Europe & Central Asia | Spain | 1549 | 0.12 | [ -0.16, 0.40] | 3.58 |
| Europe & Central Asia | Sweden | 8860 | 0.07 | [ -0.03, 0.18] | 7.94 |
| Europe & Central Asia | Türkiye | 253 | 0.94 | [ -0.08, 1.97] | 0.41 |
| Europe & Central Asia | United Kingdom | 2494 | 0.06 | [ -0.13, 0.25] | 5.42 |
| Latin America & Caribbean | Argentina | 1547 | 0.19 | [ -0.06, 0.43] | 4.29 |
| Latin America & Caribbean | Brazil | 2220 | 0.32 | [ 0.06, 0.59] | 3.89 |
| Latin America & Caribbean | Mexico | 1103 | 0.21 | [ -0.04, 0.47] | 4.06 |
| Middle East, North Africa, Afghanistan & Pakistan | Egypt | 2208 | 0.43 | [ 0.06, 0.79] | 2.50 |
| North America | United States | 22775 | 0.09 | [ -0.00, 0.18] | 8.30 |
| South Asia | India | 3858 | 0.22 | [ -0.02, 0.45] | 4.45 |
| Sub-Saharan Africa | Kenya | 6743 | 0.08 | [ -0.14, 0.30] | 4.79 |
| Sub-Saharan Africa | Nigeria | 2502 | -0.29 | [ -0.76, 0.19] | 1.64 |
| Sub-Saharan Africa | South Africa | 431 | 0.10 | [ -0.67, 0.88] | 0.69 |
| Sub-Saharan Africa | Tanzania | 4215 | 0.42 | [ 0.17, 0.66] | 4.25 |
| | **Overall** | | 0.13 | [ 0.06, 0.20] | |

$\tau^2 = 0.01$, $I^2 = 58.7\%$, $H^2 = 2.42$

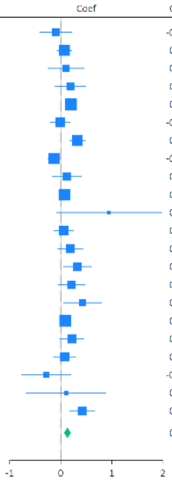

### Supplementary Figure S12: Sensitivity analysis modelling the frequency of arts engagement amongst participants who engaged in the arts

#### a) Feeling nervous, anxious, or on edge

| Region | Country | N | OR | 95% CI | Weight (%) |
|---|---|---|---|---|---|
| East Asia & Pacific | Australia | 1793 | 0.95 | [0.85, 1.08] | 3.84 |
| East Asia & Pacific | China | 2950 | 0.96 | [0.88, 1.05] | 6.35 |
| East Asia & Pacific | Hong Kong | 207 | 1.39 | [0.82, 2.37] | 0.23 |
| East Asia & Pacific | Indonesia | 732 | 0.90 | [0.75, 1.08] | 1.74 |
| East Asia & Pacific | Japan | 3432 | 1.01 | [0.93, 1.10] | 6.94 |
| East Asia & Pacific | Philippines | 1286 | 1.00 | [0.88, 1.13] | 3.64 |
| Europe & Central Asia | Germany | 2098 | 1.01 | [0.92, 1.11] | 5.38 |
| Europe & Central Asia | Poland | 1643 | 1.11 | [0.94, 1.31] | 2.13 |
| Europe & Central Asia | Spain | 1236 | 1.02 | [0.89, 1.17] | 3.02 |
| Europe & Central Asia | Sweden | 7210 | 1.06 | [1.00, 1.12] | 10.93 |
| Europe & Central Asia | Türkiye | 207 | 0.71 | [0.50, 1.00] | 0.54 |
| Europe & Central Asia | United Kingdom | 1798 | 1.03 | [0.92, 1.15] | 4.08 |
| Latin America & Caribbean | Argentina | 1072 | 0.91 | [0.79, 1.04] | 3.15 |
| Latin America & Caribbean | Brazil | 1683 | 0.96 | [0.85, 1.07] | 3.96 |
| Latin America & Caribbean | Mexico | 750 | 1.02 | [0.84, 1.22] | 1.74 |
| Middle East, North Africa, Afghanistan & Pakistan | Egypt | 263 | 0.69 | [0.52, 0.92] | 0.76 |
| North America | United States | 19540 | 1.07 | [1.03, 1.11] | 14.74 |
| South Asia | India | 1868 | 1.03 | [0.93, 1.15] | 4.50 |
| Sub-Saharan Africa | Kenya | 5343 | 1.00 | [0.94, 1.06] | 9.98 |
| Sub-Saharan Africa | Nigeria | 2186 | 1.07 | [0.97, 1.18] | 5.36 |
| Sub-Saharan Africa | South Africa | 299 | 1.12 | [0.87, 1.44] | 1.01 |
| Sub-Saharan Africa | Tanzania | 2108 | 0.99 | [0.91, 1.09] | 5.99 |
| | **Overall** | | 1.01 | [0.99, 1.04] | |

$\tau^2 = 0.00$, $I^2 = 25.0\%$, $H^2 = 1.33$

#### b) Feeling down, depressed or hopeless

| Region | Country | N | OR | 95% CI | Weight (%) |
|---|---|---|---|---|---|
| East Asia & Pacific | Australia | 1803 | 0.95 | [0.85, 1.08] | 3.58 |
| East Asia & Pacific | China | 2949 | 0.98 | [0.90, 1.06] | 6.27 |
| East Asia & Pacific | Hong Kong | 204 | 0.97 | [0.61, 1.55] | 0.27 |
| East Asia & Pacific | Indonesia | 731 | 1.20 | [1.01, 1.43] | 1.83 |
| East Asia & Pacific | Japan | 3435 | 1.01 | [0.93, 1.10] | 6.72 |
| East Asia & Pacific | Philippines | 1284 | 1.03 | [0.91, 1.17] | 3.50 |
| Europe & Central Asia | Germany | 2101 | 1.03 | [0.93, 1.14] | 4.59 |
| Europe & Central Asia | Poland | 1643 | 1.03 | [0.85, 1.24] | 1.59 |
| Europe & Central Asia | Spain | 1238 | 0.97 | [0.85, 1.10] | 3.07 |
| Europe & Central Asia | Sweden | 7200 | 1.00 | [0.95, 1.06] | 11.49 |
| Europe & Central Asia | Türkiye | 208 | 0.73 | [0.55, 0.97] | 0.71 |
| Europe & Central Asia | United Kingdom | 1802 | 0.99 | [0.89, 1.11] | 4.03 |
| Latin America & Caribbean | Argentina | 1070 | 0.98 | [0.84, 1.15] | 2.29 |
| Latin America & Caribbean | Brazil | 1688 | 0.93 | [0.83, 1.05] | 3.94 |
| Latin America & Caribbean | Mexico | 749 | 1.04 | [0.87, 1.23] | 1.83 |
| Middle East, North Africa, Afghanistan & Pakistan | Egypt | 263 | 0.71 | [0.51, 0.97] | 0.58 |
| North America | United States | 19549 | 1.06 | [1.02, 1.10] | 16.20 |
| South Asia | India | 1875 | 0.97 | [0.87, 1.09] | 4.11 |
| Sub-Saharan Africa | Kenya | 5350 | 0.98 | [0.93, 1.04] | 10.97 |
| Sub-Saharan Africa | Nigeria | 2196 | 1.08 | [0.98, 1.18] | 5.43 |
| Sub-Saharan Africa | South Africa | 298 | 0.94 | [0.74, 1.20] | 1.02 |
| Sub-Saharan Africa | Tanzania | 2111 | 0.98 | [0.89, 1.07] | 5.96 |
| | **Overall** | | 1.00 | [0.98, 1.03] | |

$\tau^2 = 0.00$, $I^2 = 18.9\%$, $H^2 = 1.23$

#### c) Unable to stop or control worrying

| Region | Country | N | OR | 95% CI | Weight (%) |
|---|---|---|---|---|---|
| East Asia & Pacific | Australia | 1801 | 0.94 | [0.83, 1.06] | 2.91 |
| East Asia & Pacific | China | 2950 | 1.03 | [0.95, 1.12] | 5.61 |
| East Asia & Pacific | Hong Kong | 207 | 1.46 | [0.87, 2.44] | 0.16 |
| East Asia & Pacific | Indonesia | 733 | 1.09 | [0.90, 1.31] | 1.23 |
| East Asia & Pacific | Japan | 3472 | 1.07 | [0.98, 1.17] | 5.61 |
| East Asia & Pacific | Philippines | 1284 | 0.91 | [0.81, 1.03] | 2.77 |
| Europe & Central Asia | Germany | 2108 | 0.99 | [0.89, 1.10] | 3.95 |
| Europe & Central Asia | Poland | 1648 | 1.15 | [0.96, 1.37] | 1.31 |
| Europe & Central Asia | Spain | 1239 | 1.08 | [0.93, 1.25] | 1.95 |
| Europe & Central Asia | Sweden | 7205 | 1.04 | [0.98, 1.10] | 11.15 |
| Europe & Central Asia | Türkiye | 207 | 0.63 | [0.45, 0.89] | 0.36 |
| Europe & Central Asia | United Kingdom | 1806 | 0.99 | [0.89, 1.11] | 3.17 |
| Latin America & Caribbean | Argentina | 1068 | 1.12 | [0.96, 1.30] | 1.83 |
| Latin America & Caribbean | Brazil | 1684 | 0.94 | [0.85, 1.05] | 3.72 |
| Latin America & Caribbean | Mexico | 744 | 0.96 | [0.80, 1.15] | 1.28 |
| Middle East, North Africa, Afghanistan & Pakistan | Egypt | 263 | 0.73 | [0.55, 0.96] | 0.52 |
| North America | United States | 19576 | 1.03 | [1.00, 1.07] | 26.18 |
| South Asia | India | 1872 | 0.96 | [0.86, 1.07] | 3.71 |
| Sub-Saharan Africa | Kenya | 5353 | 0.99 | [0.94, 1.05] | 12.49 |
| Sub-Saharan Africa | Nigeria | 2190 | 1.08 | [0.98, 1.19] | 4.28 |
| Sub-Saharan Africa | South Africa | 299 | 1.14 | [0.89, 1.46] | 0.67 |
| Sub-Saharan Africa | Tanzania | 2108 | 1.06 | [0.97, 1.15] | 5.16 |
| | **Overall** | | 1.02 | [1.00, 1.04] | |

$\tau^2 = 0.00$, $I^2 = 1.82\%$, $H^2 = 1.02$

#### d) Little interest or pleasure in doing things

| Region | Country | N | OR | 95% CI | Weight (%) |
|---|---|---|---|---|---|
| East Asia & Pacific | Australia | 1800 | 0.93 | [0.83, 1.04] | 2.94 |
| East Asia & Pacific | China | 2948 | 1.08 | [0.99, 1.17] | 5.88 |
| East Asia & Pacific | Hong Kong | 207 | 0.88 | [0.55, 1.42] | 0.17 |
| East Asia & Pacific | Indonesia | 728 | 1.09 | [0.92, 1.30] | 1.29 |
| East Asia & Pacific | Japan | 3471 | 1.03 | [0.95, 1.12] | 5.97 |
| East Asia & Pacific | Philippines | 1284 | 0.96 | [0.85, 1.08] | 2.91 |
| Europe & Central Asia | Germany | 2112 | 1.05 | [0.94, 1.17] | 3.34 |
| Europe & Central Asia | Poland | 1646 | 1.25 | [1.06, 1.48] | 1.43 |
| Europe & Central Asia | Spain | 1235 | 1.08 | [0.95, 1.24] | 2.20 |
| Europe & Central Asia | Sweden | 7156 | 0.99 | [0.93, 1.04] | 13.12 |
| Europe & Central Asia | Türkiye | 205 | 0.63 | [0.44, 0.88] | 0.33 |
| Europe & Central Asia | United Kingdom | 1804 | 0.99 | [0.89, 1.10] | 3.43 |
| Latin America & Caribbean | Argentina | 1070 | 0.91 | [0.80, 1.05] | 2.06 |
| Latin America & Caribbean | Brazil | 1683 | 1.01 | [0.91, 1.12] | 3.59 |
| Latin America & Caribbean | Mexico | 748 | 0.96 | [0.82, 1.13] | 1.51 |
| Middle East, North Africa, Afghanistan & Pakistan | Egypt | 263 | 1.21 | [0.90, 1.61] | 0.48 |
| North America | United States | 19554 | 1.04 | [1.00, 1.08] | 24.41 |
| South Asia | India | 1873 | 1.00 | [0.90, 1.11] | 3.49 |
| Sub-Saharan Africa | Kenya | 5355 | 1.01 | [0.95, 1.07] | 11.25 |
| Sub-Saharan Africa | Nigeria | 2193 | 1.03 | [0.93, 1.13] | 4.49 |
| Sub-Saharan Africa | South Africa | 300 | 0.87 | [0.69, 1.09] | 0.77 |
| Sub-Saharan Africa | Tanzania | 2109 | 1.00 | [0.91, 1.09] | 4.93 |
| | **Overall** | | 1.01 | [0.99, 1.03] | |

$\tau^2 = 0.00$, $I^2 = 0.01\%$, $H^2 = 1.00$

#### e) Suffering

| Region | Country | N | OR | 95% CI | Weight (%) |
|---|---|---|---|---|---|
| East Asia & Pacific | Australia | 1799 | 1.03 | [0.91, 1.15] | 3.61 |
| East Asia & Pacific | China | 2950 | 0.93 | [0.86, 1.01] | 6.39 |
| East Asia & Pacific | Hong Kong | 207 | 0.77 | [0.45, 1.32] | 0.18 |
| East Asia & Pacific | Indonesia | 729 | 0.99 | [0.82, 1.19] | 1.50 |
| East Asia & Pacific | Japan | 3467 | 0.97 | [0.90, 1.05] | 6.88 |
| East Asia & Pacific | Philippines | 1285 | 0.99 | [0.87, 1.11] | 3.22 |
| Europe & Central Asia | Germany | 2104 | 1.08 | [0.98, 1.20] | 4.66 |
| Europe & Central Asia | Poland | 1651 | 1.10 | [0.95, 1.28] | 2.28 |
| Europe & Central Asia | Spain | 1230 | 0.96 | [0.85, 1.09] | 3.04 |
| Europe & Central Asia | Sweden | 7205 | 1.04 | [0.99, 1.09] | 11.81 |
| Europe & Central Asia | Türkiye | 206 | 0.62 | [0.43, 0.87] | 0.43 |
| Europe & Central Asia | United Kingdom | 1803 | 0.99 | [0.90, 1.10] | 4.48 |
| Latin America & Caribbean | Argentina | 1072 | 1.02 | [0.89, 1.16] | 2.78 |
| Latin America & Caribbean | Brazil | 1676 | 1.00 | [0.90, 1.11] | 4.19 |
| Latin America & Caribbean | Mexico | 747 | 0.96 | [0.81, 1.12] | 1.98 |
| Middle East, North Africa, Afghanistan & Pakistan | Egypt | 263 | 0.90 | [0.63, 1.27] | 0.44 |
| North America | United States | 19525 | 1.03 | [1.00, 1.07] | 17.55 |
| South Asia | India | 1873 | 1.06 | [0.94, 1.19] | 3.39 |
| Sub-Saharan Africa | Kenya | 5357 | 0.96 | [0.91, 1.02] | 10.03 |
| Sub-Saharan Africa | Nigeria | 2188 | 0.95 | [0.86, 1.05] | 4.77 |
| Sub-Saharan Africa | South Africa | 298 | 1.10 | [0.88, 1.38] | 1.04 |
| Sub-Saharan Africa | Tanzania | 2112 | 1.04 | [0.95, 1.14] | 5.35 |
| | **Overall** | | 1.00 | [0.98, 1.03] | |

$\tau^2 = 0.00$, $I^2 = 17.8\%$, $H^2 = 1.22$

#### f) Feeling happy

| Region | Country | N | Coef | 95% CI | Weight (%) |
|---|---|---|---|---|---|
| East Asia & Pacific | Australia | 1800 | -0.03 | [-0.11, 0.05] | 6.17 |
| East Asia & Pacific | China | 2949 | 0.09 | [0.02, 0.15] | 7.53 |
| East Asia & Pacific | Hong Kong | 207 | 0.15 | [-0.11, 0.42] | 0.85 |
| East Asia & Pacific | Indonesia | 733 | 0.01 | [-0.16, 0.18] | 1.95 |
| East Asia & Pacific | Japan | 3453 | 0.03 | [-0.03, 0.09] | 8.20 |
| East Asia & Pacific | Philippines | 1287 | 0.11 | [-0.02, 0.24] | 3.12 |
| Europe & Central Asia | Germany | 2108 | 0.06 | [-0.01, 0.12] | 7.62 |
| Europe & Central Asia | Poland | 1659 | -0.04 | [-0.13, 0.06] | 4.77 |
| Europe & Central Asia | Spain | 1240 | 0.02 | [-0.09, 0.13] | 3.88 |
| Europe & Central Asia | Sweden | 7219 | 0.03 | [-0.01, 0.07] | 11.36 |
| Europe & Central Asia | Türkiye | 208 | 0.50 | [0.17, 0.82] | 0.59 |
| Europe & Central Asia | United Kingdom | 1807 | 0.05 | [-0.03, 0.13] | 5.94 |
| Latin America & Caribbean | Argentina | 1072 | 0.10 | [-0.02, 0.22] | 3.64 |
| Latin America & Caribbean | Brazil | 1683 | 0.19 | [0.08, 0.30] | 4.05 |
| Latin America & Caribbean | Mexico | 751 | 0.05 | [-0.07, 0.17] | 3.43 |
| Middle East, North Africa, Afghanistan & Pakistan | Egypt | 262 | 0.22 | [-0.18, 0.61] | 0.40 |
| North America | United States | 19579 | -0.01 | [-0.04, 0.02] | 12.89 |
| South Asia | India | 1879 | -0.02 | [-0.20, 0.16] | 1.80 |
| Sub-Saharan Africa | Kenya | 5355 | 0.04 | [-0.05, 0.13] | 5.23 |
| Sub-Saharan Africa | Nigeria | 2199 | -0.03 | [-0.16, 0.10] | 3.17 |
| Sub-Saharan Africa | South Africa | 300 | -0.17 | [-0.53, 0.20] | 0.47 |
| Sub-Saharan Africa | Tanzania | 2113 | 0.01 | [-0.12, 0.15] | 2.93 |
| | **Overall** | | 0.04 | [0.01, 0.06] | |

$\tau^2 = 0.00$, $I^2 = 39.3\%$, $H^2 = 1.65$

#### g) Understanding purpose in life

| Region | Country | N | Coef | 95% CI | Weight (%) |
|---|---|---|---|---|---|
| East Asia & Pacific | Australia | 1800 | -0.03 | [ -0.15, 0.09] | 3.77 |
| East Asia & Pacific | China | 2949 | 0.11 | [ 0.04, 0.18] | 7.14 |
| East Asia & Pacific | Hong Kong | 207 | 0.01 | [ -0.27, 0.28] | 0.94 |
| East Asia & Pacific | Indonesia | 732 | 0.04 | [ -0.13, 0.20] | 2.42 |
| East Asia & Pacific | Japan | 3459 | 0.07 | [ 0.01, 0.13] | 8.85 |
| East Asia & Pacific | Philippines | 1286 | -0.00 | [ -0.12, 0.11] | 4.09 |
| Europe & Central Asia | Germany | 2106 | -0.00 | [ -0.08, 0.07] | 7.00 |
| Europe & Central Asia | Poland | 1655 | -0.11 | [ -0.25, 0.02] | 3.29 |
| Europe & Central Asia | Spain | 1235 | 0.11 | [ 0.01, 0.22] | 4.55 |
| Europe & Central Asia | Sweden | 7162 | 0.01 | [ -0.04, 0.06] | 9.94 |
| Europe & Central Asia | Türkiye | 207 | 0.22 | [ -0.09, 0.54] | 0.72 |
| Europe & Central Asia | United Kingdom | 1800 | -0.01 | [ -0.10, 0.08] | 5.39 |
| Latin America & Caribbean | Argentina | 1072 | 0.15 | [ 0.02, 0.28] | 3.43 |
| Latin America & Caribbean | Brazil | 1685 | 0.09 | [ -0.01, 0.19] | 4.99 |
| Latin America & Caribbean | Mexico | 751 | 0.11 | [ 0.01, 0.22] | 4.67 |
| Middle East, North Africa, Afghanistan & Pakistan | Egypt | 261 | 0.14 | [ -0.17, 0.46] | 0.73 |
| North America | United States | 19552 | 0.01 | [ -0.03, 0.04] | 11.71 |
| South Asia | India | 1874 | 0.04 | [ -0.12, 0.19] | 2.56 |
| Sub-Saharan Africa | Kenya | 5355 | -0.05 | [ -0.14, 0.04] | 5.85 |
| Sub-Saharan Africa | Nigeria | 2196 | -0.03 | [ -0.15, 0.10] | 3.68 |
| Sub-Saharan Africa | South Africa | 300 | -0.15 | [ -0.44, 0.15] | 0.82 |
| Sub-Saharan Africa | Tanzania | 2094 | 0.04 | [ -0.09, 0.17] | 3.45 |
| | **Overall** | | 0.03 | [ 0.00, 0.06] | |

$\tau^2 = 0.00$, $I^2 = 38.5\%$, $H^2 = 1.63$

#### h) Life satisfaction

| Region | Country | N | Coef | 95% CI | Weight (%) |
|---|---|---|---|---|---|
| East Asia & Pacific | Australia | 1802 | 0.02 | [ -0.07, 0.11] | 5.68 |
| East Asia & Pacific | China | 2934 | 0.09 | [ 0.02, 0.16] | 7.18 |
| East Asia & Pacific | Hong Kong | 200 | 0.21 | [ -0.06, 0.47] | 0.97 |
| East Asia & Pacific | Indonesia | 729 | 0.12 | [ -0.06, 0.31] | 1.81 |
| East Asia & Pacific | Japan | 3439 | 0.05 | [ -0.01, 0.11] | 8.61 |
| East Asia & Pacific | Philippines | 1284 | 0.06 | [ -0.07, 0.19] | 3.29 |
| Europe & Central Asia | Germany | 2089 | 0.06 | [ -0.02, 0.13] | 7.14 |
| Europe & Central Asia | Poland | 1647 | -0.08 | [ -0.19, 0.02] | 4.46 |
| Europe & Central Asia | Spain | 1234 | 0.08 | [ -0.02, 0.18] | 4.87 |
| Europe & Central Asia | Sweden | 7207 | 0.00 | [ -0.04, 0.04] | 10.84 |
| Europe & Central Asia | Türkiye | 208 | 0.42 | [ 0.09, 0.74] | 0.66 |
| Europe & Central Asia | United Kingdom | 1784 | 0.09 | [ 0.02, 0.16] | 7.09 |
| Latin America & Caribbean | Argentina | 1074 | 0.13 | [ 0.01, 0.26] | 3.60 |
| Latin America & Caribbean | Brazil | 1685 | 0.07 | [ -0.03, 0.17] | 4.83 |
| Latin America & Caribbean | Mexico | 751 | 0.03 | [ -0.10, 0.16] | 3.26 |
| Middle East, North Africa, Afghanistan & Pakistan | Egypt | 261 | -0.01 | [ -0.26, 0.25] | 1.02 |
| North America | United States | 19576 | -0.02 | [ -0.05, 0.01] | 12.12 |
| South Asia | India | 1878 | 0.00 | [ -0.19, 0.19] | 1.80 |
| Sub-Saharan Africa | Kenya | 5357 | -0.05 | [ -0.15, 0.05] | 4.86 |
| Sub-Saharan Africa | Nigeria | 2199 | -0.01 | [ -0.15, 0.13] | 3.05 |
| Sub-Saharan Africa | South Africa | 295 | -0.36 | [ -0.73, 0.01] | 0.51 |
| Sub-Saharan Africa | Tanzania | 2112 | -0.05 | [ -0.21, 0.11] | 2.37 |
| | **Overall** | | 0.03 | [ 0.01, 0.06] | |

$\tau^2 = 0.00$, $I^2 = 40.7\%$, $H^2 = 1.69$

#### i) Life evaluation: Today

| Region | Country | N | Coef | 95% CI | Weight (%) |
|---|---|---|---|---|---|
| East Asia & Pacific | Australia | 1803 | -0.03 | [ -0.11, 0.06] | 5.81 |
| East Asia & Pacific | China | 2950 | 0.08 | [ 0.01, 0.16] | 6.72 |
| East Asia & Pacific | Hong Kong | 207 | 0.06 | [ -0.16, 0.27] | 1.42 |
| East Asia & Pacific | Indonesia | 731 | 0.11 | [ -0.08, 0.30] | 1.80 |
| East Asia & Pacific | Japan | 3460 | 0.09 | [ 0.03, 0.14] | 8.13 |
| East Asia & Pacific | Philippines | 1283 | 0.17 | [ 0.04, 0.29] | 3.46 |
| Europe & Central Asia | Germany | 2103 | 0.07 | [ 0.00, 0.13] | 7.37 |
| Europe & Central Asia | Poland | 1656 | -0.07 | [ -0.17, 0.02] | 4.97 |
| Europe & Central Asia | Spain | 1241 | 0.03 | [ -0.06, 0.13] | 4.97 |
| Europe & Central Asia | Sweden | 7217 | 0.00 | [ -0.03, 0.04] | 10.07 |
| Europe & Central Asia | Türkiye | 207 | 0.43 | [ 0.12, 0.74] | 0.74 |
| Europe & Central Asia | United Kingdom | 1805 | 0.08 | [ 0.01, 0.16] | 6.48 |
| Latin America & Caribbean | Argentina | 1074 | 0.08 | [ -0.03, 0.19] | 4.28 |
| Latin America & Caribbean | Brazil | 1689 | 0.12 | [ 0.02, 0.21] | 4.91 |
| Latin America & Caribbean | Mexico | 752 | 0.07 | [ -0.06, 0.20] | 3.28 |
| Middle East, North Africa, Afghanistan & Pakistan | Egypt | 262 | 0.19 | [ -0.12, 0.49] | 0.75 |
| North America | United States | 19592 | -0.00 | [ -0.03, 0.02] | 11.18 |
| South Asia | India | 1871 | 0.13 | [ -0.06, 0.32] | 1.81 |
| Sub-Saharan Africa | Kenya | 5358 | 0.02 | [ -0.06, 0.11] | 5.44 |
| Sub-Saharan Africa | Nigeria | 2203 | -0.06 | [ -0.20, 0.08] | 2.94 |
| Sub-Saharan Africa | South Africa | 299 | -0.14 | [ -0.49, 0.22] | 0.56 |
| Sub-Saharan Africa | Tanzania | 2113 | 0.05 | [ -0.09, 0.19] | 2.92 |
| | **Overall** | | 0.05 | [ 0.02, 0.07] | |

$\tau^2 = 0.00$, $I^2 = 48.9\%$, $H^2 = 1.96$

#### j) Life evaluation: Five years from now

| Region | Country | N | Coef | 95% CI | Weight (%) |
|---|---|---|---|---|---|
| East Asia & Pacific | Australia | 1802 | -0.02 | [ -0.11, 0.06] | 5.42 |
| East Asia & Pacific | China | 2935 | 0.08 | [ 0.01, 0.15] | 6.94 |
| East Asia & Pacific | Hong Kong | 201 | 0.18 | [ -0.10, 0.45] | 0.83 |
| East Asia & Pacific | Indonesia | 723 | 0.04 | [ -0.10, 0.17] | 2.94 |
| East Asia & Pacific | Japan | 3467 | 0.07 | [ 0.01, 0.13] | 8.05 |
| East Asia & Pacific | Philippines | 1282 | -0.01 | [ -0.12, 0.10] | 3.79 |
| Europe & Central Asia | Germany | 2093 | 0.08 | [ 0.01, 0.15] | 6.87 |
| Europe & Central Asia | Poland | 1628 | -0.07 | [ -0.20, 0.05] | 3.22 |
| Europe & Central Asia | Spain | 1231 | 0.05 | [ -0.05, 0.14] | 4.74 |
| Europe & Central Asia | Sweden | 7196 | 0.02 | [ -0.02, 0.06] | 10.19 |
| Europe & Central Asia | Türkiye | 201 | 0.19 | [ -0.10, 0.49] | 0.72 |
| Europe & Central Asia | United Kingdom | 1785 | 0.05 | [ -0.02, 0.13] | 6.13 |
| Latin America & Caribbean | Argentina | 1058 | -0.04 | [ -0.16, 0.07] | 3.76 |
| Latin America & Caribbean | Brazil | 1684 | 0.08 | [ -0.00, 0.16] | 5.62 |
| Latin America & Caribbean | Mexico | 745 | 0.07 | [ -0.05, 0.19] | 3.53 |
| Middle East, North Africa, Afghanistan & Pakistan | Egypt | 232 | -0.21 | [ -0.57, 0.15] | 0.51 |
| North America | United States | 19578 | -0.05 | [ -0.08, -0.01] | 10.95 |
| South Asia | India | 1774 | 0.01 | [ -0.14, 0.16] | 2.52 |
| Sub-Saharan Africa | Kenya | 5331 | 0.01 | [ -0.08, 0.10] | 5.27 |
| Sub-Saharan Africa | Nigeria | 2176 | -0.05 | [ -0.16, 0.06] | 3.86 |
| Sub-Saharan Africa | South Africa | 295 | -0.15 | [ -0.43, 0.12] | 0.85 |
| Sub-Saharan Africa | Tanzania | 2042 | -0.05 | [ -0.17, 0.08] | 3.31 |
| | **Overall** | | 0.02 | [ -0.01, 0.05] | |

$\tau^2 = 0.00$, $I^2 = 44.2\%$, $H^2 = 1.79$

#### k) Feeling lonely

| Region | Country | N | Coef | 95% CI | Weight (%) |
|---|---|---|---|---|---|
| East Asia & Pacific | Australia | 1803 | 0.01 | [ -0.11, 0.13] | 3.33 |
| East Asia & Pacific | China | 2950 | -0.10 | [ -0.19, -0.00] | 5.84 |
| East Asia & Pacific | Hong Kong | 206 | -0.17 | [ -0.59, 0.25] | 0.29 |
| East Asia & Pacific | Indonesia | 732 | 0.18 | [ -0.08, 0.44] | 0.75 |
| East Asia & Pacific | Japan | 3479 | -0.03 | [ -0.11, 0.05] | 7.51 |
| East Asia & Pacific | Philippines | 1287 | 0.01 | [ -0.16, 0.18] | 1.73 |
| Europe & Central Asia | Germany | 2112 | 0.00 | [ -0.09, 0.09] | 6.23 |
| Europe & Central Asia | Poland | 1652 | 0.06 | [ -0.10, 0.22] | 1.96 |
| Europe & Central Asia | Spain | 1243 | 0.03 | [ -0.12, 0.19] | 2.13 |
| Europe & Central Asia | Sweden | 7229 | -0.03 | [ -0.08, 0.01] | 21.39 |
| Europe & Central Asia | Türkiye | 207 | -0.00 | [ -0.35, 0.35] | 0.42 |
| Europe & Central Asia | United Kingdom | 1799 | -0.05 | [ -0.16, 0.05] | 4.43 |
| Latin America & Caribbean | Argentina | 1077 | -0.12 | [ -0.31, 0.08] | 1.35 |
| Latin America & Caribbean | Brazil | 1684 | -0.06 | [ -0.21, 0.10] | 2.02 |
| Latin America & Caribbean | Mexico | 749 | 0.11 | [ -0.10, 0.33] | 1.12 |
| Middle East, North Africa, Afghanistan & Pakistan | Egypt | 263 | -0.22 | [ -0.75, 0.31] | 0.18 |
| North America | United States | 19598 | -0.00 | [ -0.05, 0.04] | 29.20 |
| South Asia | India | 1880 | 0.08 | [ -0.13, 0.28] | 1.22 |
| Sub-Saharan Africa | Kenya | 5352 | -0.01 | [ -0.12, 0.10] | 4.34 |
| Sub-Saharan Africa | Nigeria | 2196 | 0.10 | [ -0.07, 0.26] | 1.88 |
| Sub-Saharan Africa | South Africa | 300 | -0.04 | [ -0.45, 0.36] | 0.31 |
| Sub-Saharan Africa | Tanzania | 2111 | -0.07 | [ -0.22, 0.08] | 2.37 |
| | **Overall** | | -0.02 | [ -0.04, 0.01] | |

$\tau^2 = 0.00$, $I^2 = 0.00\%$, $H^2 = 1.00$

#### l) Life is worthwhile

| Region | Country | N | Coef | 95% CI | Weight (%) |
|---|---|---|---|---|---|
| East Asia & Pacific | Australia | 1800 | 0.02 | [ -0.08, 0.12] | 3.91 |
| East Asia & Pacific | China | 2948 | 0.08 | [ 0.01, 0.14] | 7.81 |
| East Asia & Pacific | Hong Kong | 204 | 0.24 | [ 0.00, 0.48] | 0.75 |
| East Asia & Pacific | Indonesia | 732 | 0.03 | [ -0.13, 0.20] | 1.58 |
| East Asia & Pacific | Japan | 3469 | 0.02 | [ -0.04, 0.08] | 9.33 |
| East Asia & Pacific | Philippines | 1284 | 0.04 | [ -0.08, 0.17] | 2.57 |
| Europe & Central Asia | Germany | 2110 | 0.05 | [ -0.02, 0.12] | 7.80 |
| Europe & Central Asia | Poland | 1650 | -0.10 | [ -0.21, 0.01] | 3.16 |
| Europe & Central Asia | Spain | 1232 | 0.05 | [ -0.07, 0.16] | 3.08 |
| Europe & Central Asia | Sweden | 7215 | 0.01 | [ -0.03, 0.06] | 13.90 |
| Europe & Central Asia | Türkiye | 208 | 0.21 | [ -0.08, 0.51] | 0.49 |
| Europe & Central Asia | United Kingdom | 1798 | 0.04 | [ -0.04, 0.12] | 5.63 |
| Latin America & Caribbean | Argentina | 1070 | 0.11 | [ -0.01, 0.22] | 3.17 |
| Latin America & Caribbean | Brazil | 1687 | 0.07 | [ -0.03, 0.17] | 3.90 |
| Latin America & Caribbean | Mexico | 749 | 0.02 | [ -0.11, 0.15] | 2.58 |
| Middle East, North Africa, Afghanistan & Pakistan | Egypt | 262 | -0.03 | [ -0.39, 0.32] | 0.34 |
| North America | United States | 19584 | -0.01 | [ -0.05, 0.02] | 18.81 |
| South Asia | India | 1871 | -0.02 | [ -0.18, 0.14] | 1.65 |
| Sub-Saharan Africa | Kenya | 5358 | 0.00 | [ -0.09, 0.09] | 4.55 |
| Sub-Saharan Africa | Nigeria | 2199 | 0.01 | [ -0.12, 0.14] | 2.35 |
| Sub-Saharan Africa | South Africa | 297 | -0.20 | [ -0.53, 0.13] | 0.40 |
| Sub-Saharan Africa | Tanzania | 2112 | 0.02 | [ -0.12, 0.15] | 2.26 |
| | **Overall** | | 0.02 | [ 0.00, 0.04] | |

$\tau^2 = 0.00$, $I^2 = 13.4\%$, $H^2 = 1.16$

m) Feeling grateful

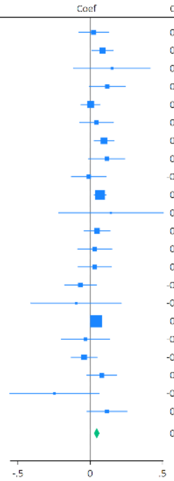

| Region | Country | N | Coef | 95% CI | Weight (%) |
|---|---|---|---|---|---|
| East Asia & Pacific | Australia | 1731 | 0.02 | [ -0.08, 0.13] | 3.17 |
| East Asia & Pacific | China | 2950 | 0.09 | [ 0.01, 0.16] | 6.09 |
| East Asia & Pacific | Hong Kong | 206 | 0.15 | [ -0.11, 0.42] | 0.50 |
| East Asia & Pacific | Indonesia | 729 | 0.12 | [ -0.01, 0.24] | 2.23 |
| East Asia & Pacific | Japan | 3451 | 0.00 | [ -0.06, 0.07] | 7.45 |
| East Asia & Pacific | Philippines | 1285 | 0.04 | [ -0.07, 0.16] | 2.52 |
| Europe & Central Asia | Germany | 2095 | 0.10 | [ 0.03, 0.16] | 7.01 |
| Europe & Central Asia | Poland | 1648 | 0.11 | [ -0.01, 0.24] | 2.20 |
| Europe & Central Asia | Spain | 1238 | -0.01 | [ -0.13, 0.11] | 2.37 |
| Europe & Central Asia | Sweden | 7221 | 0.07 | [ 0.03, 0.11] | 16.87 |
| Europe & Central Asia | Türkiye | 203 | 0.14 | [ -0.22, 0.51] | 0.27 |
| Europe & Central Asia | United Kingdom | 1771 | 0.05 | [ -0.04, 0.14] | 4.25 |
| Latin America & Caribbean | Argentina | 1072 | 0.03 | [ -0.09, 0.15] | 2.47 |
| Latin America & Caribbean | Brazil | 1682 | 0.03 | [ -0.08, 0.14] | 2.66 |
| Latin America & Caribbean | Mexico | 750 | -0.07 | [ -0.18, 0.04] | 2.85 |
| Middle East, North Africa, Afghanistan & Pakistan | Egypt | 262 | -0.10 | [ -0.41, 0.22] | 0.36 |
| North America | United States | 18490 | 0.04 | [ 0.01, 0.07] | 25.84 |
| South Asia | India | 1872 | -0.03 | [ -0.20, 0.14] | 1.24 |
| Sub-Saharan Africa | Kenya | 5358 | -0.04 | [ -0.13, 0.05] | 4.19 |
| Sub-Saharan Africa | Nigeria | 2193 | 0.08 | [ -0.02, 0.18] | 3.24 |
| Sub-Saharan Africa | South Africa | 297 | -0.25 | [ -0.56, 0.06] | 0.37 |
| Sub-Saharan Africa | Tanzania | 2103 | 0.12 | [ -0.02, 0.25] | 1.84 |
| | **Overall** | | 0.05 | [ 0.03, 0.06] | |
| | $\tau^2 = 0.00$, $I^2 = 6.03\%$, $H^2 = 1.06$ | | | | |

n) Feeling at peace

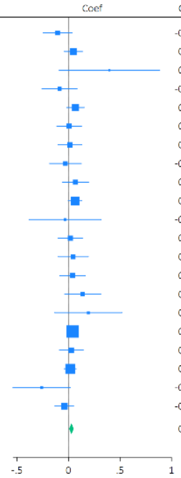

| Region | Country | N | Coef | 95% CI | Weight (%) |
|---|---|---|---|---|---|
| East Asia & Pacific | Australia | 1801 | -0.11 | [ -0.25, 0.03] | 2.45 |
| East Asia & Pacific | China | 2949 | 0.05 | [ -0.04, 0.13] | 6.20 |
| East Asia & Pacific | Hong Kong | 207 | 0.40 | [ -0.09, 0.88] | 0.20 |
| East Asia & Pacific | Indonesia | 731 | -0.09 | [ -0.26, 0.08] | 1.66 |
| East Asia & Pacific | Japan | 3451 | 0.07 | [ -0.02, 0.15] | 6.38 |
| East Asia & Pacific | Philippines | 1285 | 0.01 | [ -0.11, 0.13] | 3.33 |
| Europe & Central Asia | Germany | 2101 | 0.01 | [ -0.10, 0.13] | 3.53 |
| Europe & Central Asia | Poland | 1641 | -0.03 | [ -0.18, 0.12] | 1.99 |
| Europe & Central Asia | Spain | 1232 | 0.07 | [ -0.06, 0.19] | 3.00 |
| Europe & Central Asia | Sweden | 7212 | 0.06 | [ -0.00, 0.13] | 11.02 |
| Europe & Central Asia | Türkiye | 208 | -0.03 | [ -0.38, 0.32] | 0.39 |
| Europe & Central Asia | United Kingdom | 1801 | 0.02 | [ -0.10, 0.14] | 3.24 |
| Latin America & Caribbean | Argentina | 1071 | 0.05 | [ -0.10, 0.19] | 2.23 |
| Latin America & Caribbean | Brazil | 1684 | 0.04 | [ -0.08, 0.16] | 3.12 |
| Latin America & Caribbean | Mexico | 748 | 0.14 | [ -0.04, 0.31] | 1.57 |
| Middle East, North Africa, Afghanistan & Pakistan | Egypt | 263 | 0.19 | [ -0.13, 0.52] | 0.45 |
| North America | United States | 19523 | 0.04 | [ -0.00, 0.08] | 24.45 |
| South Asia | India | 1874 | 0.03 | [ -0.09, 0.14] | 3.57 |
| Sub-Saharan Africa | Kenya | 5357 | 0.02 | [ -0.04, 0.07] | 14.76 |
| Sub-Saharan Africa | South Africa | 300 | -0.26 | [ -0.54, 0.02] | 0.61 |
| Sub-Saharan Africa | Tanzania | 2112 | -0.04 | [ -0.13, 0.05] | 5.84 |
| | **Overall** | | 0.03 | [ 0.01, 0.05] | |
| | $\tau^2 = 0.00$, $I^2 = 0.02\%$, $H^2 = 1.00$ | | | | |

o) Remain hopeful about the future

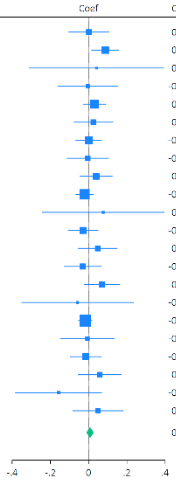

| Region | Country | N | Coef | 95% CI | Weight (%) |
|---|---|---|---|---|---|
| East Asia & Pacific | Australia | 1735 | 0.00 | [ -0.10, 0.11] | 3.19 |
| East Asia & Pacific | China | 2951 | 0.09 | [ 0.02, 0.16] | 6.39 |
| East Asia & Pacific | Hong Kong | 207 | 0.04 | [ -0.31, 0.39] | 0.30 |
| East Asia & Pacific | Indonesia | 731 | -0.00 | [ -0.16, 0.15] | 1.48 |
| East Asia & Pacific | Japan | 3457 | 0.03 | [ -0.03, 0.09] | 9.16 |
| East Asia & Pacific | Philippines | 1286 | 0.03 | [ -0.08, 0.13] | 3.36 |
| Europe & Central Asia | Germany | 2102 | -0.00 | [ -0.07, 0.07] | 7.11 |
| Europe & Central Asia | Poland | 1655 | -0.00 | [ -0.11, 0.10] | 2.97 |
| Europe & Central Asia | Spain | 1238 | 0.04 | [ -0.04, 0.12] | 4.81 |
| Europe & Central Asia | Sweden | 7230 | -0.02 | [ -0.07, 0.02] | 13.24 |
| Europe & Central Asia | Türkiye | 207 | 0.08 | [ -0.24, 0.39] | 0.36 |
| Europe & Central Asia | United Kingdom | 1773 | -0.03 | [ -0.11, 0.05] | 5.41 |
| Latin America & Caribbean | Argentina | 1072 | 0.05 | [ -0.05, 0.15] | 3.37 |
| Latin America & Caribbean | Brazil | 1683 | -0.03 | [ -0.13, 0.07] | 3.71 |
| Latin America & Caribbean | Mexico | 749 | 0.07 | [ -0.02, 0.16] | 4.03 |
| Middle East, North Africa, Afghanistan & Pakistan | Egypt | 263 | -0.06 | [ -0.35, 0.23] | 0.44 |
| North America | United States | 18477 | -0.02 | [ -0.05, 0.02] | 18.16 |
| South Asia | India | 1879 | -0.01 | [ -0.15, 0.13] | 1.85 |
| Sub-Saharan Africa | Kenya | 5355 | -0.02 | [ -0.10, 0.06] | 5.11 |
| Sub-Saharan Africa | Nigeria | 2199 | 0.06 | [ -0.05, 0.17] | 2.77 |
| Sub-Saharan Africa | South Africa | 298 | -0.16 | [ -0.38, 0.07] | 0.73 |
| Sub-Saharan Africa | Tanzania | 2112 | 0.05 | [ -0.08, 0.18] | 2.06 |
| | **Overall** | | 0.01 | [ -0.01, 0.03] | |
| | $\tau^2 = 0.00$, $I^2 = 11.1\%$, $H^2 = 1.12$ | | | | |

p) Feeling very capable in most things

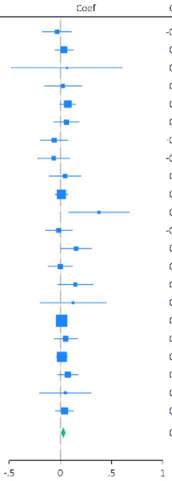

| Region | Country | N | Coef | 95% CI | Weight (%) |
|---|---|---|---|---|---|
| East Asia & Pacific | Australia | 1789 | -0.03 | [ -0.17, 0.11] | 2.27 |
| East Asia & Pacific | China | 2948 | 0.04 | [ -0.05, 0.12] | 5.79 |
| East Asia & Pacific | Hong Kong | 207 | 0.06 | [ -0.48, 0.61] | 0.15 |
| East Asia & Pacific | Indonesia | 730 | 0.03 | [ -0.15, 0.21] | 1.40 |
| East Asia & Pacific | Japan | 3440 | 0.07 | [ -0.00, 0.15] | 7.57 |
| East Asia & Pacific | Philippines | 1287 | 0.06 | [ -0.06, 0.18] | 3.07 |
| Europe & Central Asia | Germany | 2101 | -0.06 | [ -0.19, 0.07] | 2.58 |
| Europe & Central Asia | Poland | 1630 | -0.07 | [ -0.22, 0.09] | 1.89 |
| Europe & Central Asia | Spain | 1223 | 0.04 | [ -0.11, 0.20] | 1.91 |
| Europe & Central Asia | Sweden | 7177 | 0.01 | [ -0.05, 0.07] | 12.25 |
| Europe & Central Asia | Türkiye | 208 | 0.38 | [ 0.08, 0.67] | 0.52 |
| Europe & Central Asia | United Kingdom | 1792 | -0.02 | [ -0.14, 0.11] | 2.71 |
| Latin America & Caribbean | Argentina | 1073 | 0.15 | [ 0.00, 0.30] | 1.99 |
| Latin America & Caribbean | Brazil | 1675 | 0.00 | [ -0.12, 0.12] | 3.31 |
| Latin America & Caribbean | Mexico | 745 | 0.15 | [ -0.03, 0.32] | 1.54 |
| Middle East, North Africa, Afghanistan & Pakistan | Egypt | 263 | 0.12 | [ -0.20, 0.45] | 0.43 |
| North America | United States | 19465 | 0.01 | [ -0.03, 0.06] | 21.34 |
| South Asia | India | 1876 | 0.05 | [ -0.06, 0.16] | 3.61 |
| Sub-Saharan Africa | Kenya | 5348 | 0.02 | [ -0.04, 0.07] | 14.08 |
| Sub-Saharan Africa | Nigeria | 2193 | 0.07 | [ -0.02, 0.17] | 4.73 |
| Sub-Saharan Africa | South Africa | 299 | 0.05 | [ -0.20, 0.30] | 0.71 |
| Sub-Saharan Africa | Tanzania | 2112 | 0.04 | [ -0.04, 0.13] | 6.15 |
| | **Overall** | | 0.03 | [ 0.01, 0.05] | |
| | $\tau^2 = 0.00$, $I^2 = 0.01\%$, $H^2 = 1.00$ | | | | |

q) Expect more good things to happen to you

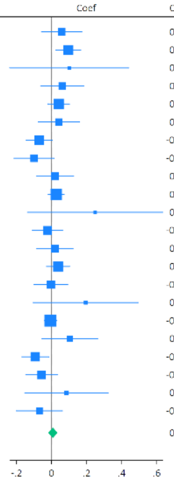

| Region | Country | N | Coef | 95% CI | Weight (%) |
|---|---|---|---|---|---|
| East Asia & Pacific | Australia | 1785 | 0.06 | [ -0.06, 0.17] | 3.70 |
| East Asia & Pacific | China | 2951 | 0.10 | [ 0.02, 0.17] | 6.66 |
| East Asia & Pacific | Hong Kong | 206 | 0.10 | [ -0.24, 0.44] | 0.56 |
| East Asia & Pacific | Indonesia | 732 | 0.06 | [ -0.06, 0.18] | 3.37 |
| East Asia & Pacific | Japan | 3470 | 0.04 | [ -0.02, 0.10] | 7.59 |
| East Asia & Pacific | Philippines | 1286 | 0.04 | [ -0.08, 0.16] | 3.58 |
| Europe & Central Asia | Germany | 2106 | -0.07 | [ -0.15, 0.00] | 6.34 |
| Europe & Central Asia | Poland | 1652 | -0.10 | [ -0.21, 0.01] | 3.73 |
| Europe & Central Asia | Spain | 1239 | 0.02 | [ -0.09, 0.13] | 4.17 |
| Europe & Central Asia | Sweden | 7217 | 0.03 | [ -0.02, 0.07] | 9.45 |
| Europe & Central Asia | Türkiye | 208 | 0.25 | [ -0.14, 0.64] | 0.44 |
| Europe & Central Asia | United Kingdom | 1794 | -0.02 | [ -0.11, 0.06] | 5.40 |
| Latin America & Caribbean | Argentina | 1072 | 0.02 | [ -0.09, 0.12] | 4.20 |
| Latin America & Caribbean | Brazil | 1684 | 0.04 | [ -0.03, 0.10] | 7.13 |
| Latin America & Caribbean | Mexico | 752 | -0.00 | [ -0.10, 0.09] | 4.75 |
| Middle East, North Africa, Afghanistan & Pakistan | Egypt | 258 | 0.20 | [ -0.10, 0.50] | 0.71 |
| North America | United States | 19500 | -0.01 | [ -0.04, 0.03] | 10.58 |
| South Asia | India | 1877 | 0.10 | [ -0.05, 0.26] | 2.23 |
| Sub-Saharan Africa | Kenya | 5359 | -0.09 | [ -0.17, -0.02] | 6.15 |
| Sub-Saharan Africa | Nigeria | 2196 | -0.06 | [ -0.15, 0.03] | 5.11 |
| Sub-Saharan Africa | South Africa | 298 | 0.08 | [ -0.15, 0.32] | 1.10 |
| Sub-Saharan Africa | Tanzania | 2112 | -0.07 | [ -0.20, 0.06] | 3.04 |
| | **Overall** | | 0.01 | [ -0.02, 0.03] | |
| | $\tau^2 = 0.00$, $I^2 = 41.7\%$, $H^2 = 1.72$ | | | | |

r) Overall mental health

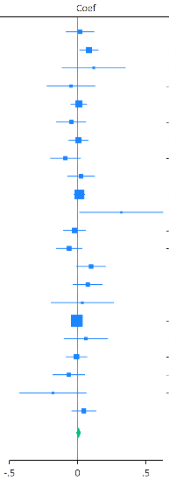

| Region | Country | N | Coef | 95% CI | Weight (%) |
|---|---|---|---|---|---|
| East Asia & Pacific | Australia | 1802 | 0.02 | [ -0.08, 0.12] | 2.57 |
| East Asia & Pacific | China | 2949 | 0.08 | [ 0.02, 0.15] | 5.84 |
| East Asia & Pacific | Hong Kong | 205 | 0.12 | [ -0.11, 0.35] | 0.50 |
| East Asia & Pacific | Indonesia | 732 | -0.05 | [ -0.22, 0.13] | 0.88 |
| East Asia & Pacific | Japan | 3474 | 0.01 | [ -0.05, 0.07] | 8.06 |
| East Asia & Pacific | Philippines | 1286 | -0.05 | [ -0.15, 0.06] | 2.33 |
| Europe & Central Asia | Germany | 2107 | 0.01 | [ -0.06, 0.08] | 5.39 |
| Europe & Central Asia | Poland | 1657 | -0.09 | [ -0.20, 0.02] | 2.16 |
| Europe & Central Asia | Spain | 1236 | 0.03 | [ -0.07, 0.12] | 2.74 |
| Europe & Central Asia | Sweden | 7241 | 0.01 | [ -0.02, 0.05] | 18.11 |
| Europe & Central Asia | Türkiye | 207 | 0.32 | [ 0.02, 0.62] | 0.29 |
| Europe & Central Asia | United Kingdom | 1793 | -0.02 | [ -0.10, 0.06] | 4.12 |
| Latin America & Caribbean | Argentina | 1074 | -0.06 | [ -0.16, 0.03] | 3.08 |
| Latin America & Caribbean | Brazil | 1688 | 0.10 | [ -0.01, 0.20] | 2.44 |
| Latin America & Caribbean | Mexico | 751 | 0.07 | [ -0.03, 0.18] | 2.32 |
| Middle East, North Africa, Afghanistan & Pakistan | Egypt | 262 | 0.03 | [ -0.19, 0.26] | 0.51 |
| North America | United States | 19592 | -0.01 | [ -0.04, 0.02] | 27.31 |
| South Asia | India | 1876 | 0.06 | [ -0.10, 0.22] | 1.04 |
| Sub-Saharan Africa | Kenya | 5352 | -0.01 | [ -0.09, 0.07] | 4.47 |
| Sub-Saharan Africa | Nigeria | 2200 | -0.06 | [ -0.18, 0.05] | 1.97 |
| Sub-Saharan Africa | South Africa | 298 | -0.18 | [ -0.43, 0.06] | 0.45 |
| Sub-Saharan Africa | Tanzania | 2108 | 0.04 | [ -0.04, 0.13] | 3.45 |
| | **Overall** | | 0.01 | [ -0.01, 0.02] | |
| | $\tau^2 = 0.00$, $I^2 = 0.24\%$, $H^2 = 1.00$ | | | | |

s) Overall physical health

| Region | Country | N | Coef | 95% CI | Weight (%) |
|---|---|---|---|---|---|
| East Asia & Pacific | Australia | 1803 | -0.04 | [ -0.13, 0.05] | 3.91 |
| East Asia & Pacific | China | 2951 | 0.07 | [ -0.00, 0.14] | 6.20 |
| East Asia & Pacific | Hong Kong | 207 | 0.20 | [ -0.05, 0.45] | 0.56 |
| East Asia & Pacific | Indonesia | 732 | 0.02 | [ -0.14, 0.18] | 1.31 |
| East Asia & Pacific | Japan | 3469 | 0.07 | [ 0.02, 0.13] | 8.44 |
| East Asia & Pacific | Philippines | 1286 | 0.03 | [ -0.09, 0.15] | 2.37 |
| Europe & Central Asia | Germany | 2111 | 0.05 | [ -0.01, 0.11] | 7.33 |
| Europe & Central Asia | Poland | 1657 | -0.06 | [ -0.17, 0.05] | 2.69 |
| Europe & Central Asia | Spain | 1239 | 0.02 | [ -0.08, 0.12] | 3.23 |
| Europe & Central Asia | Sweden | 7244 | 0.01 | [ -0.03, 0.05] | 14.77 |
| Europe & Central Asia | Türkiye | 208 | 0.12 | [ -0.15, 0.38] | 0.49 |
| Europe & Central Asia | United Kingdom | 1813 | 0.02 | [ -0.06, 0.10] | 4.84 |
| Latin America & Caribbean | Argentina | 1074 | -0.00 | [ -0.11, 0.10] | 2.92 |
| Latin America & Caribbean | Brazil | 1683 | 0.06 | [ -0.04, 0.16] | 3.28 |
| Latin America & Caribbean | Mexico | 751 | 0.03 | [ -0.08, 0.13] | 3.07 |
| Middle East, North Africa, Afghanistan & Pakistan | Egypt | 263 | 0.01 | [ -0.31, 0.34] | 0.33 |
| North America | United States | 19589 | 0.05 | [ 0.02, 0.07] | 23.95 |
| South Asia | India | 1877 | 0.01 | [ -0.16, 0.17] | 1.20 |
| Sub-Saharan Africa | Kenya | 5355 | 0.01 | [ -0.07, 0.10] | 4.46 |
| Sub-Saharan Africa | Nigeria | 2202 | -0.07 | [ -0.20, 0.06] | 2.02 |
| Sub-Saharan Africa | South Africa | 300 | -0.50 | [ -0.79, -0.21] | 0.42 |
| Sub-Saharan Africa | Tanzania | 2111 | 0.03 | [ -0.09, 0.16] | 2.23 |
| | **Overall** | | 0.03 | [ 0.01, 0.05] | |

$\tau^2 = 0.00$, $I^2 = 11.3\%$, $H^2 = 1.13$

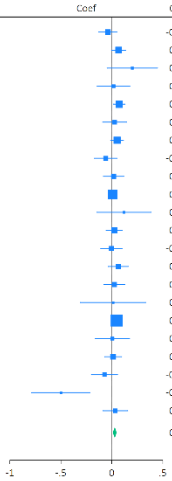

Supplementary Figure S13: Meta-regression analyses examining the moderating effects of country-level moderators

#### 1) Feeling nervous, anxious, or on edge

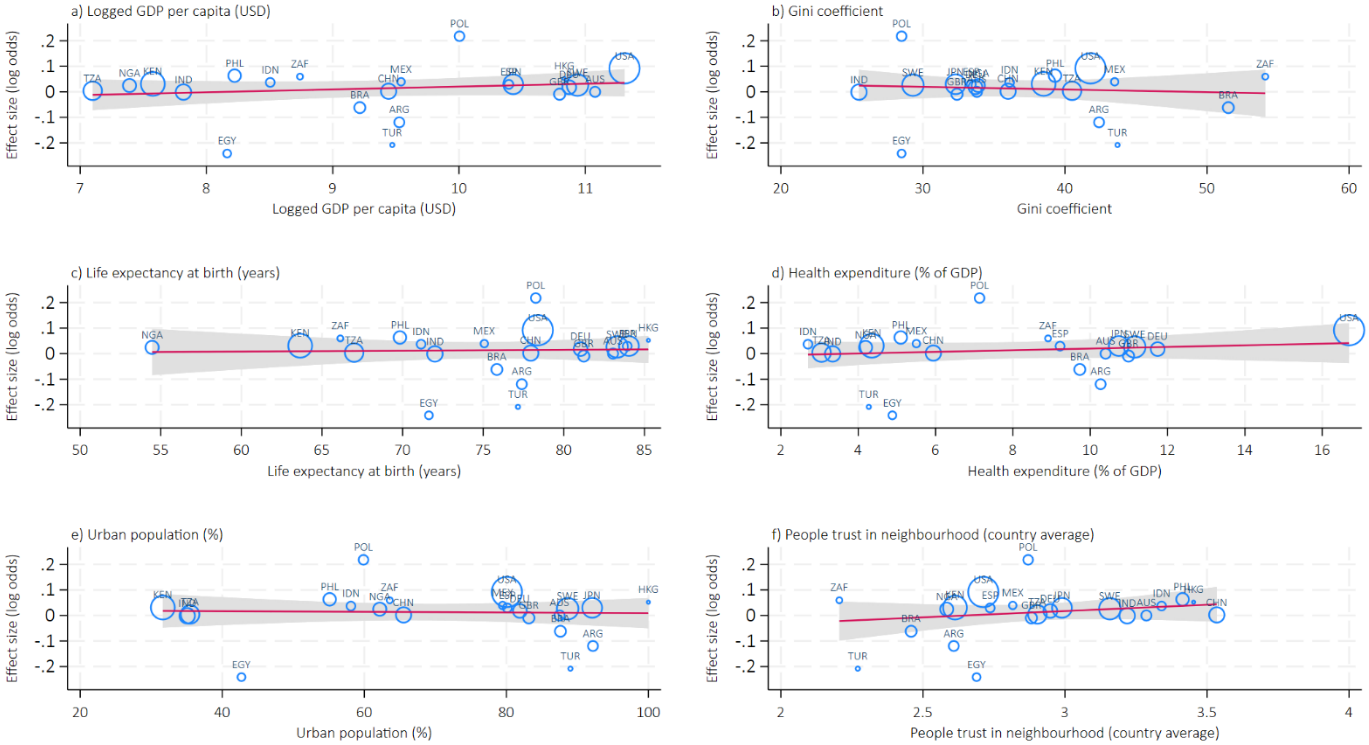

#### 2) Feeling down, depressed or hopeless

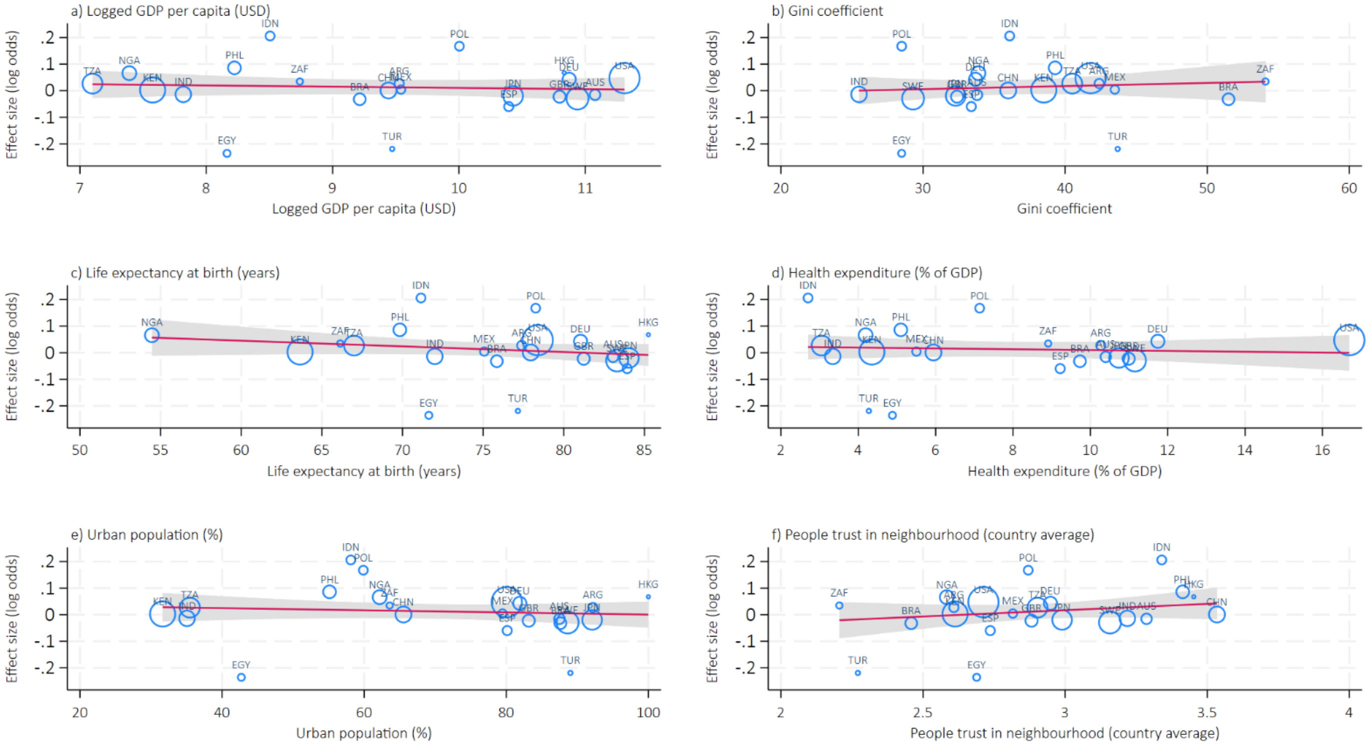

#### 3) Unable to stop or control worrying

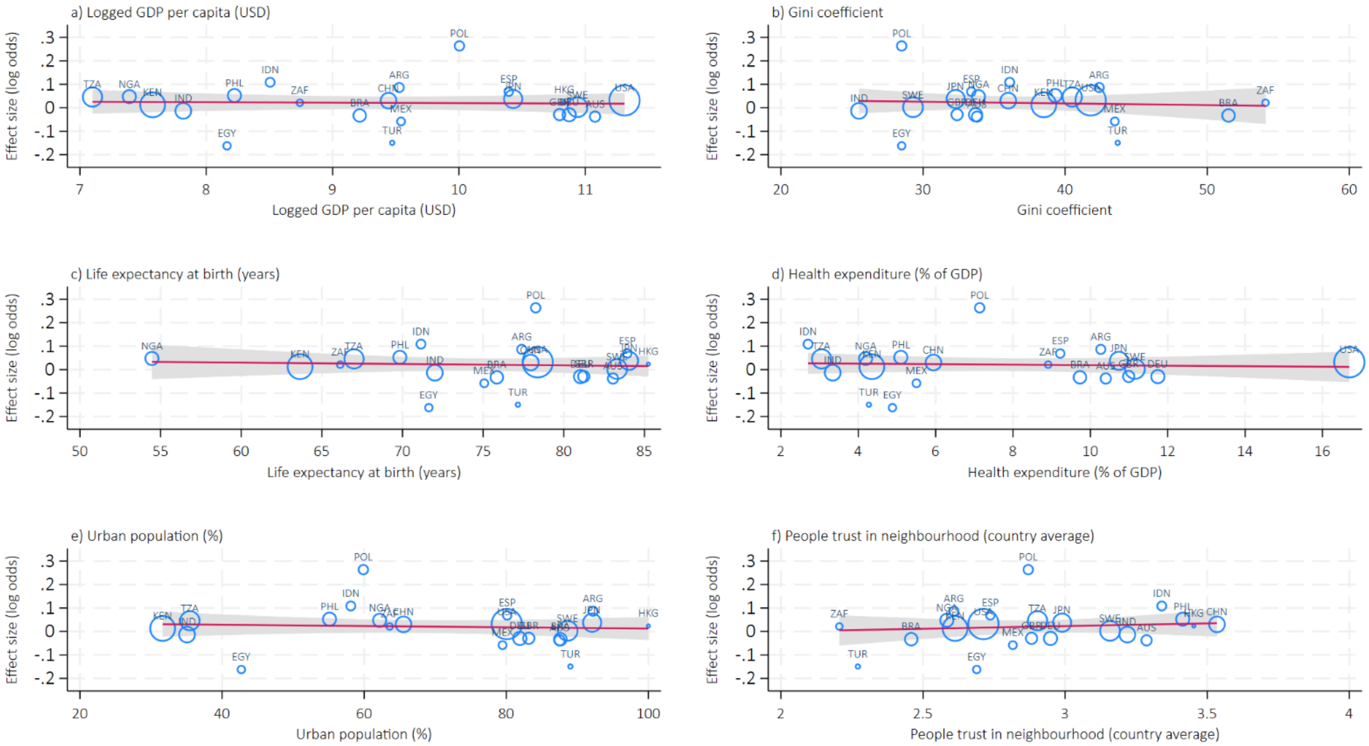

#### 4) Little interest or pleasure in doing things

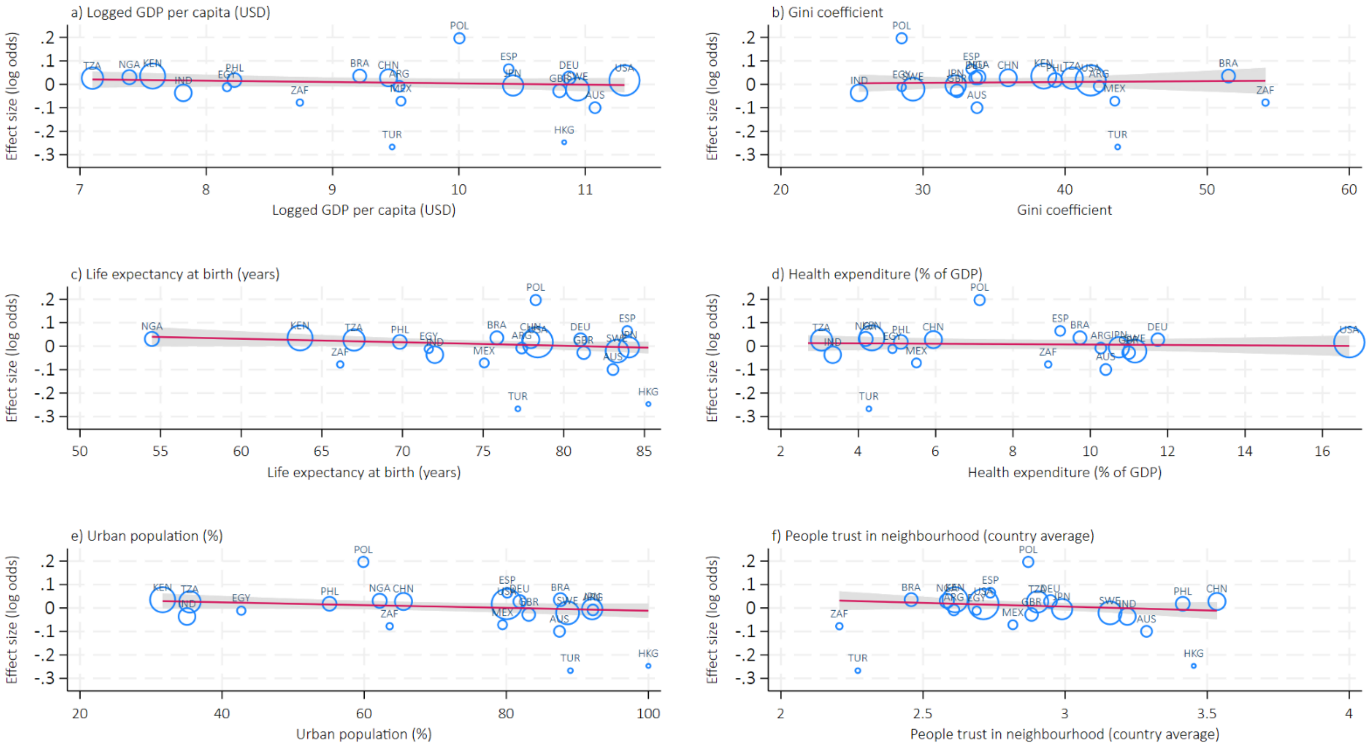

#### 5) Suffering

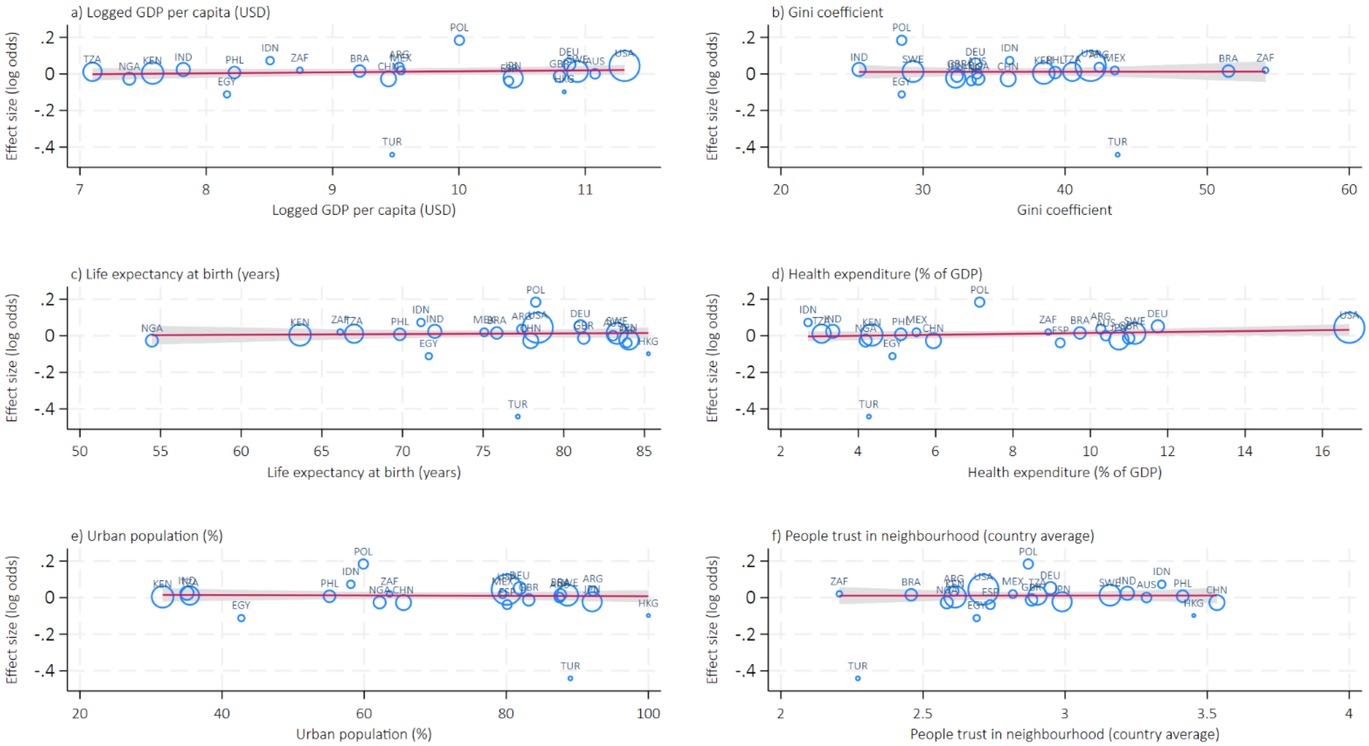

#### 6) Feeling happy

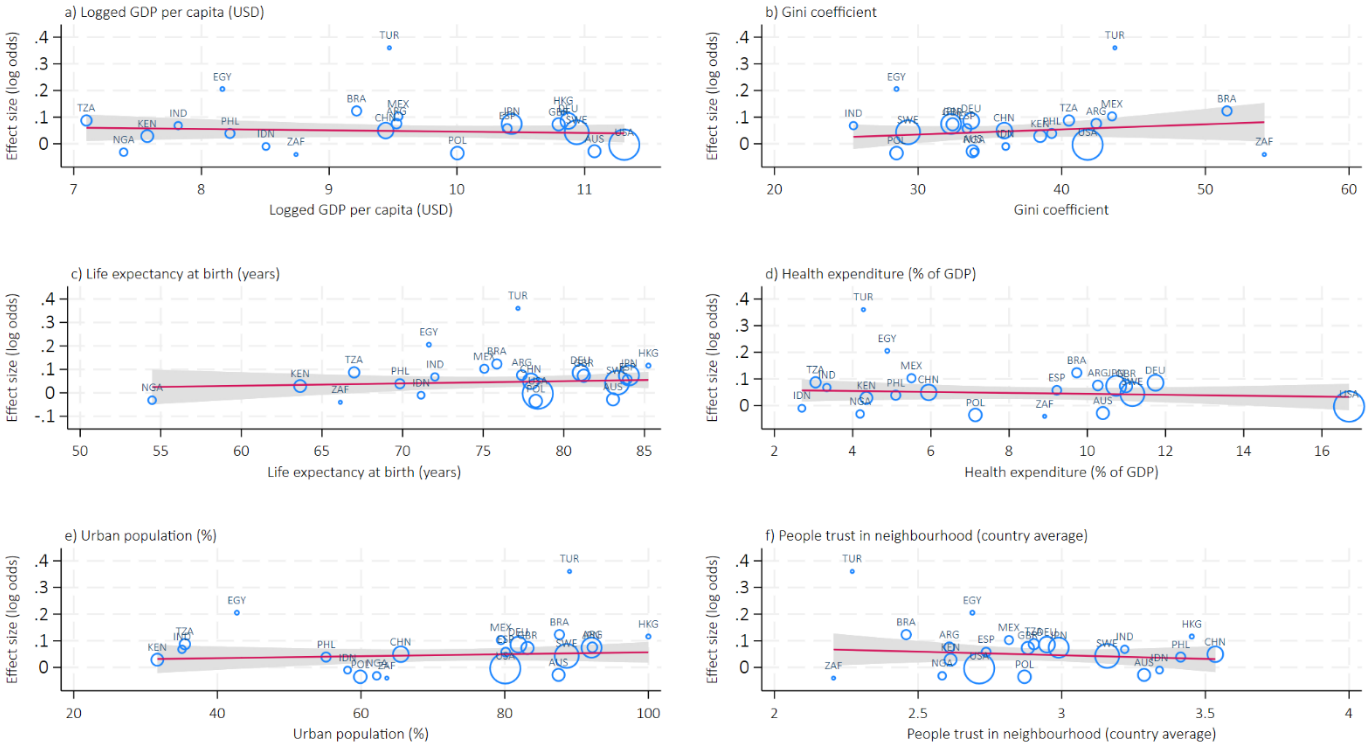

#### 7) Understanding purpose in life

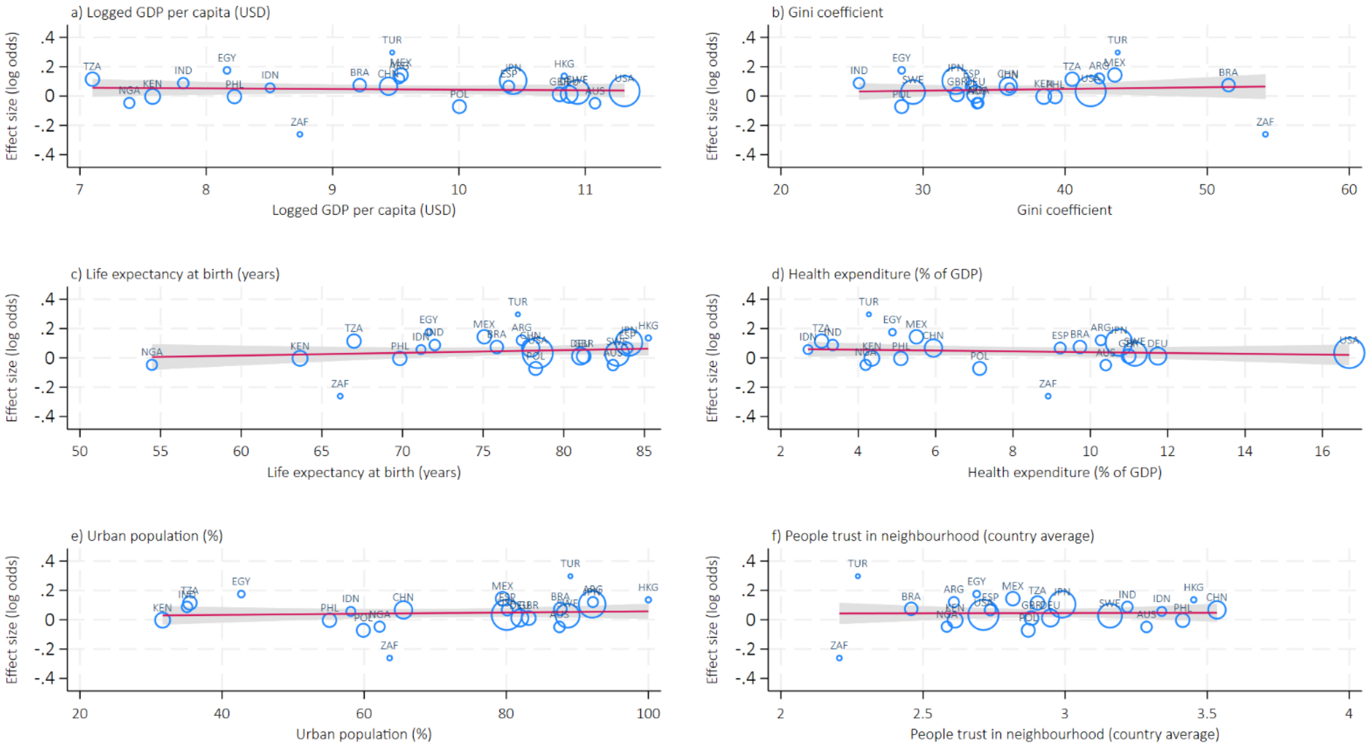

#### 8) Life satisfaction

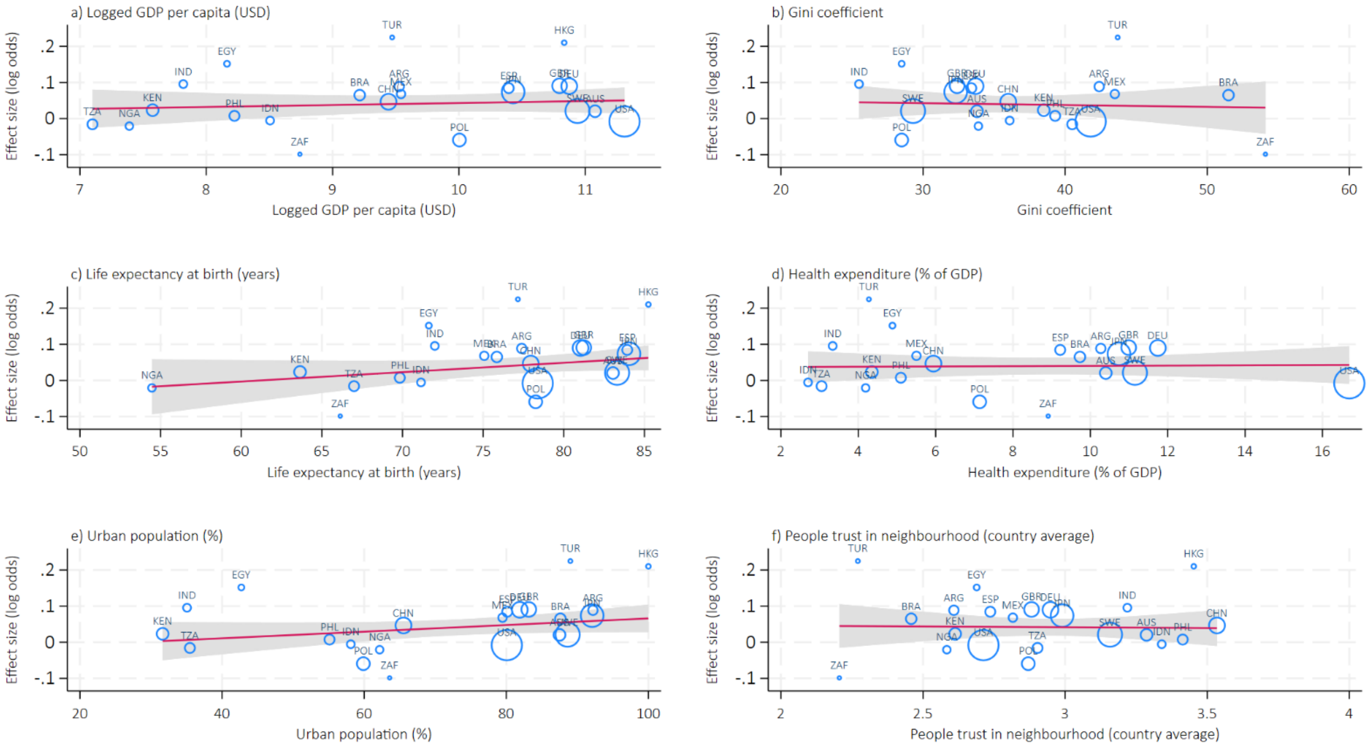

#### 9) Life evaluation: Today

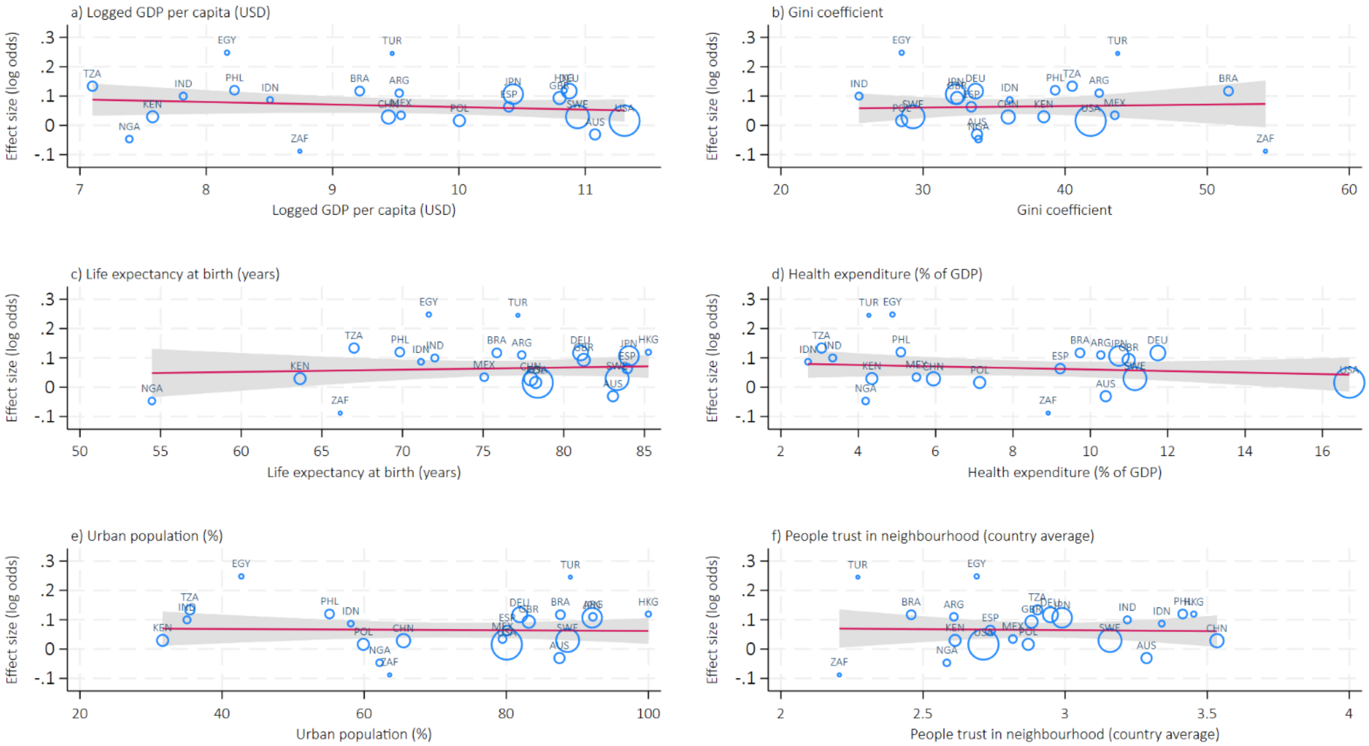

#### 10) Life evaluation: Five years from now

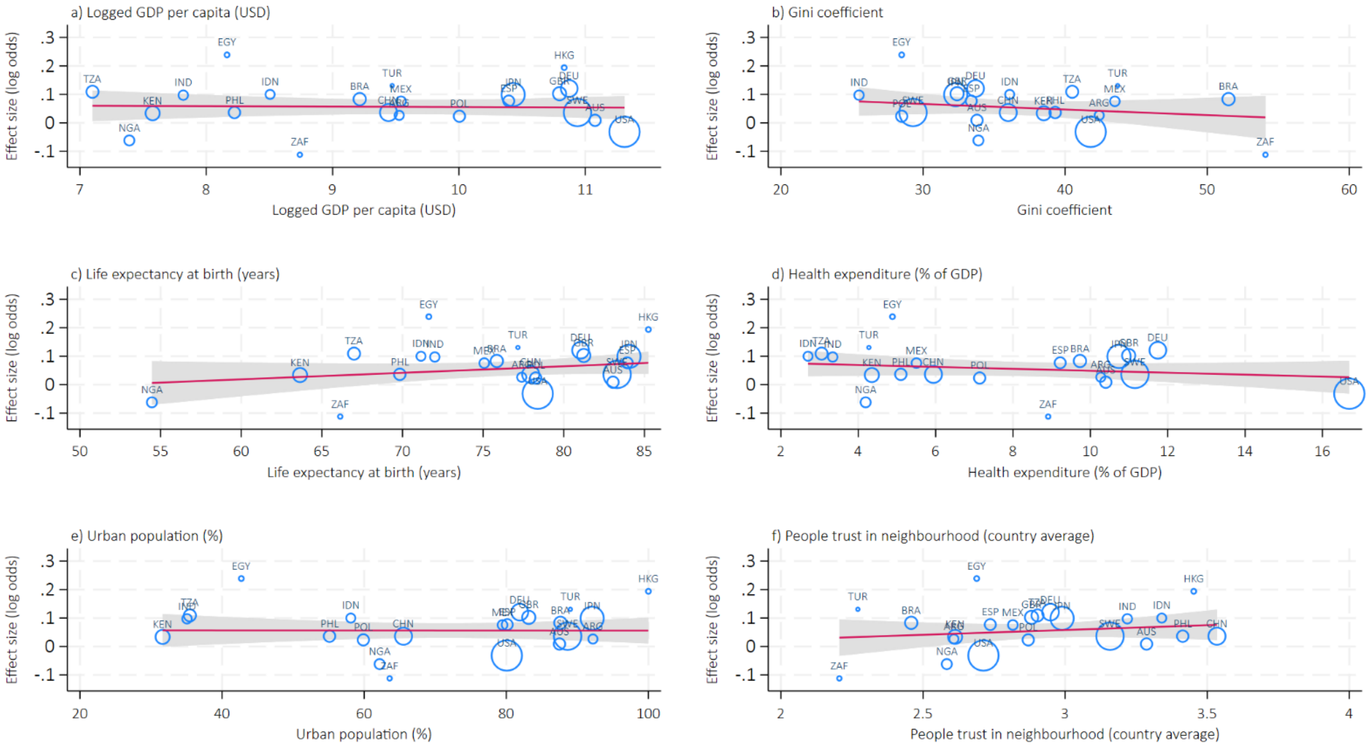

#### 11) Feeling lonely

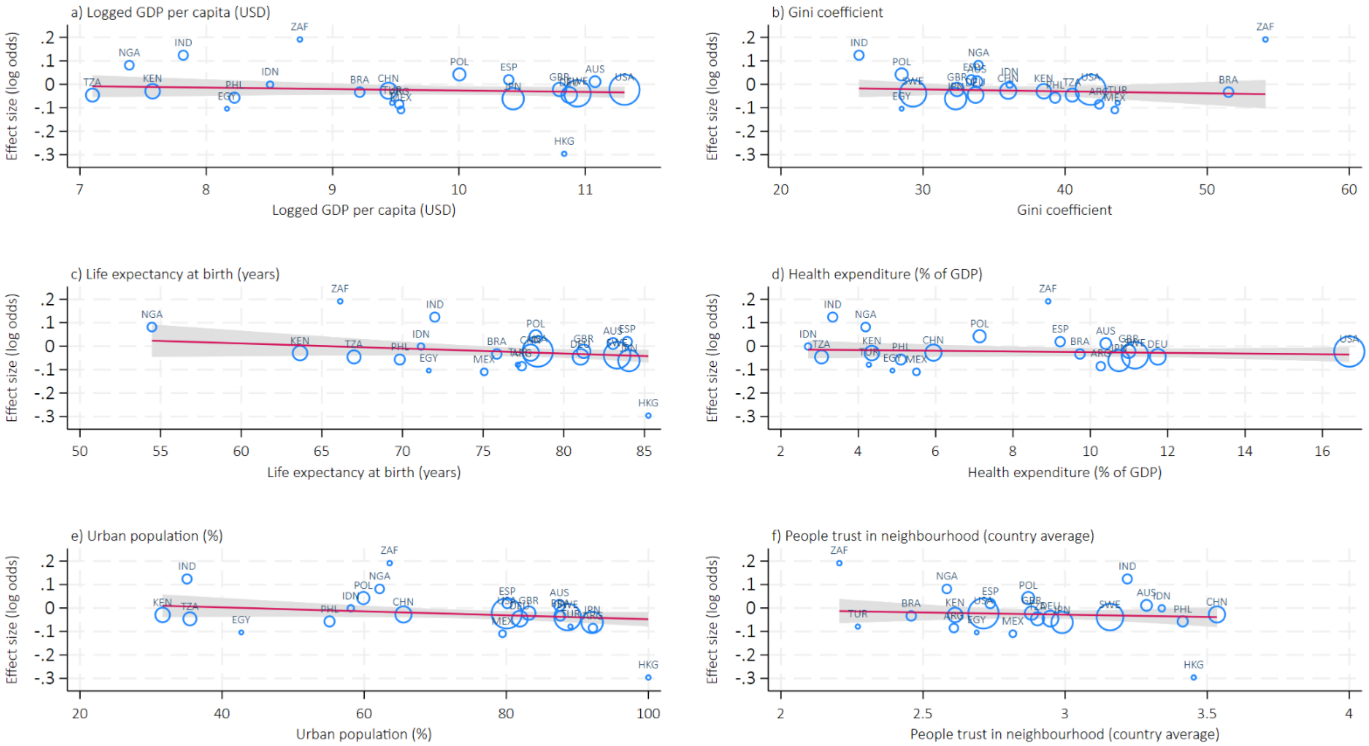

#### 12) Life is worthwhile

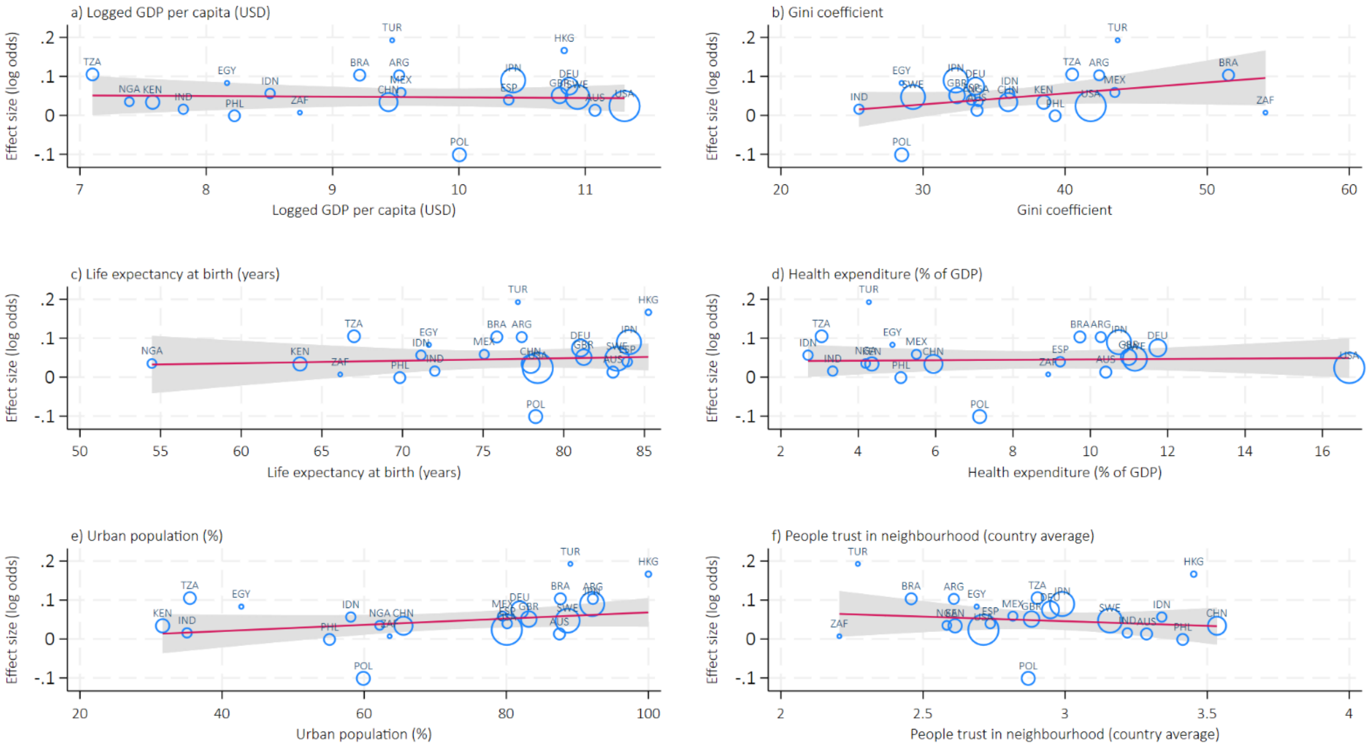

#### 13) Feeling grateful

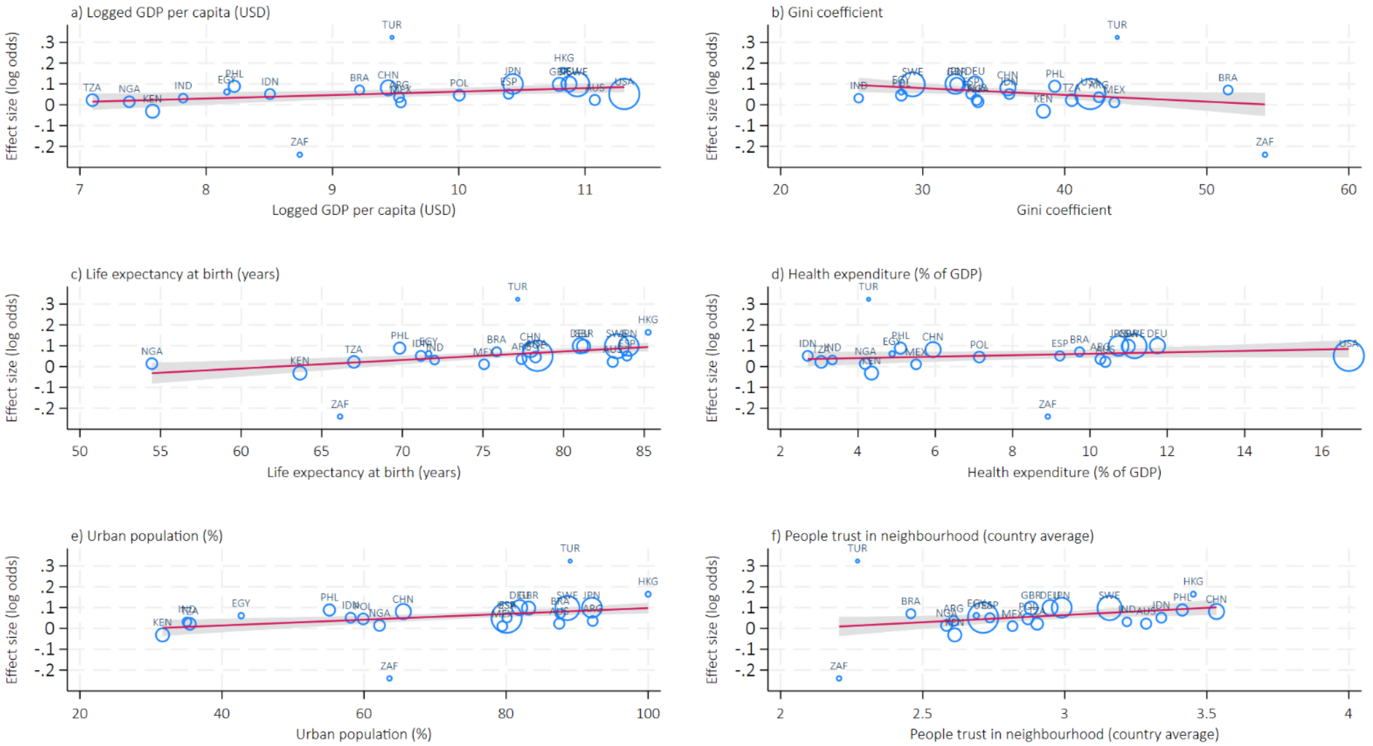

#### 14) Feeling at peace

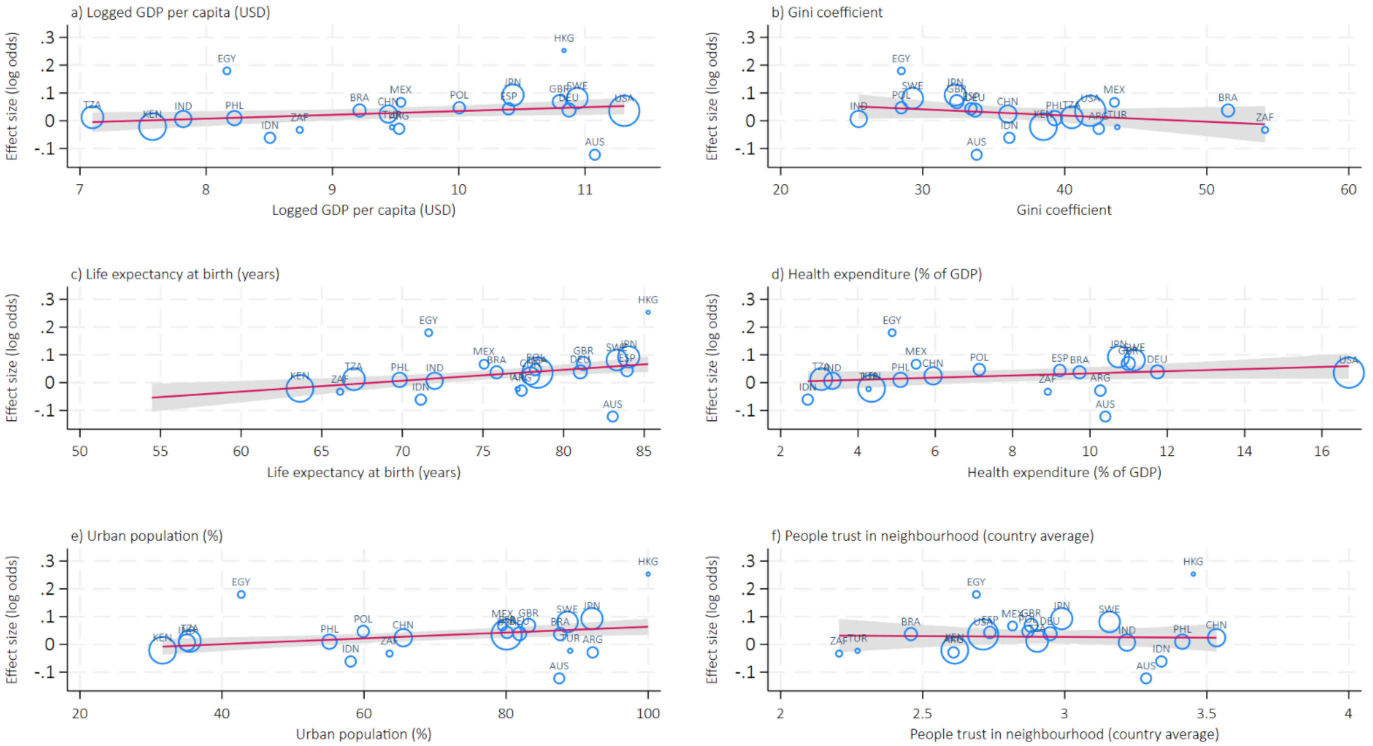

#### 15) Remain hopeful about the future

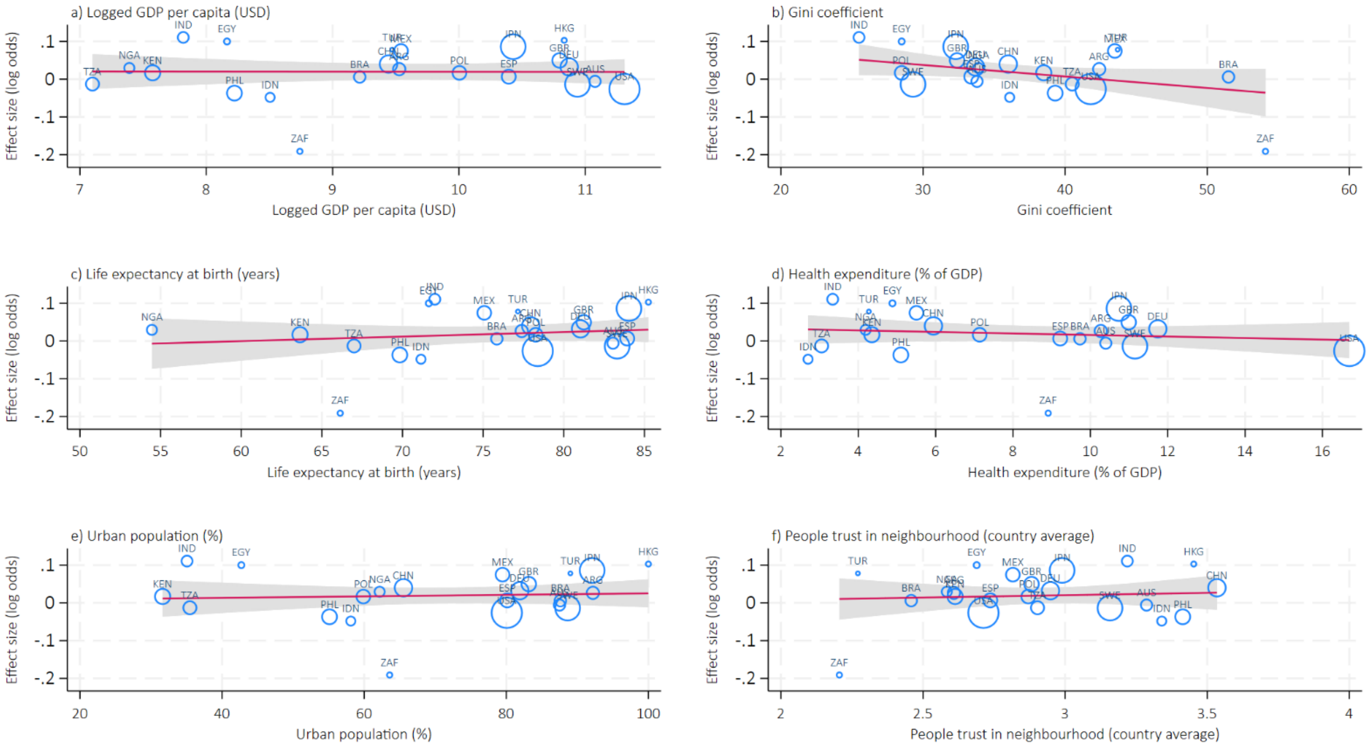

#### 16) Feeling very capable in most things

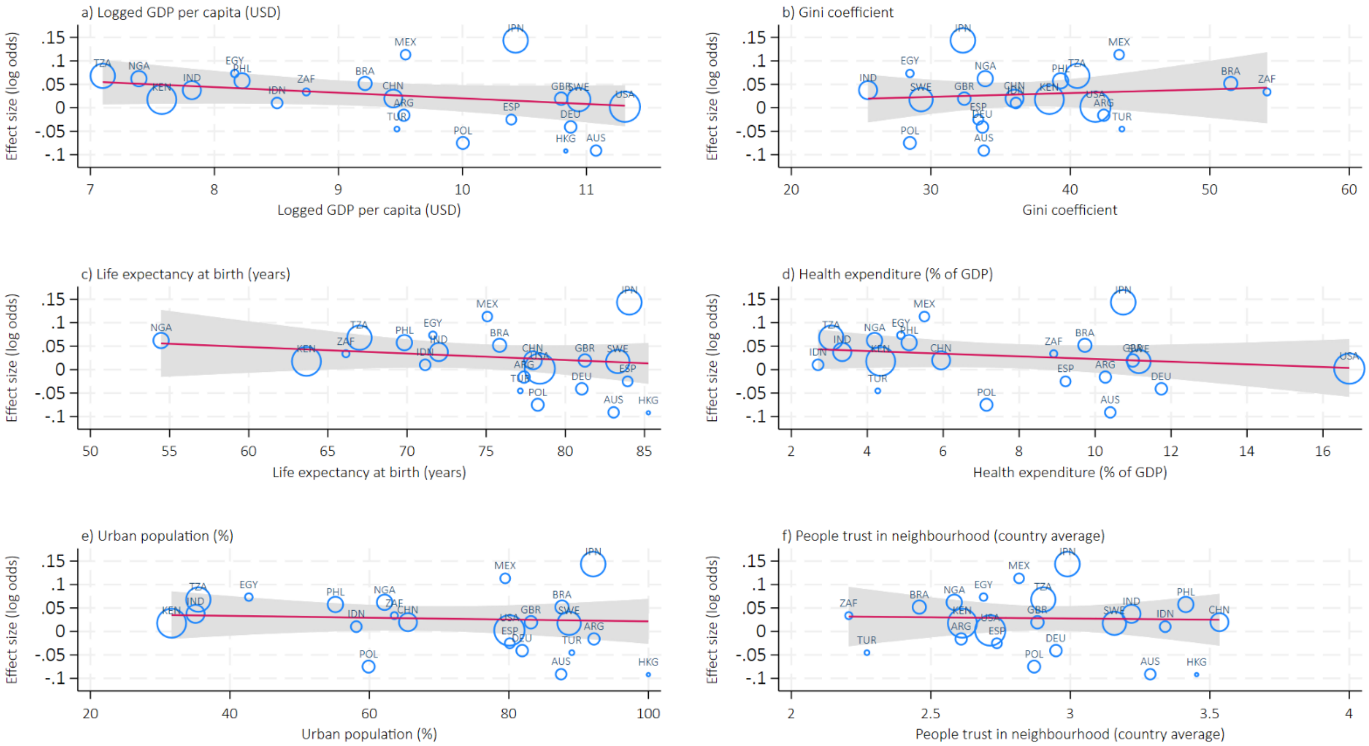

#### 17) Expect more good things to happen to you

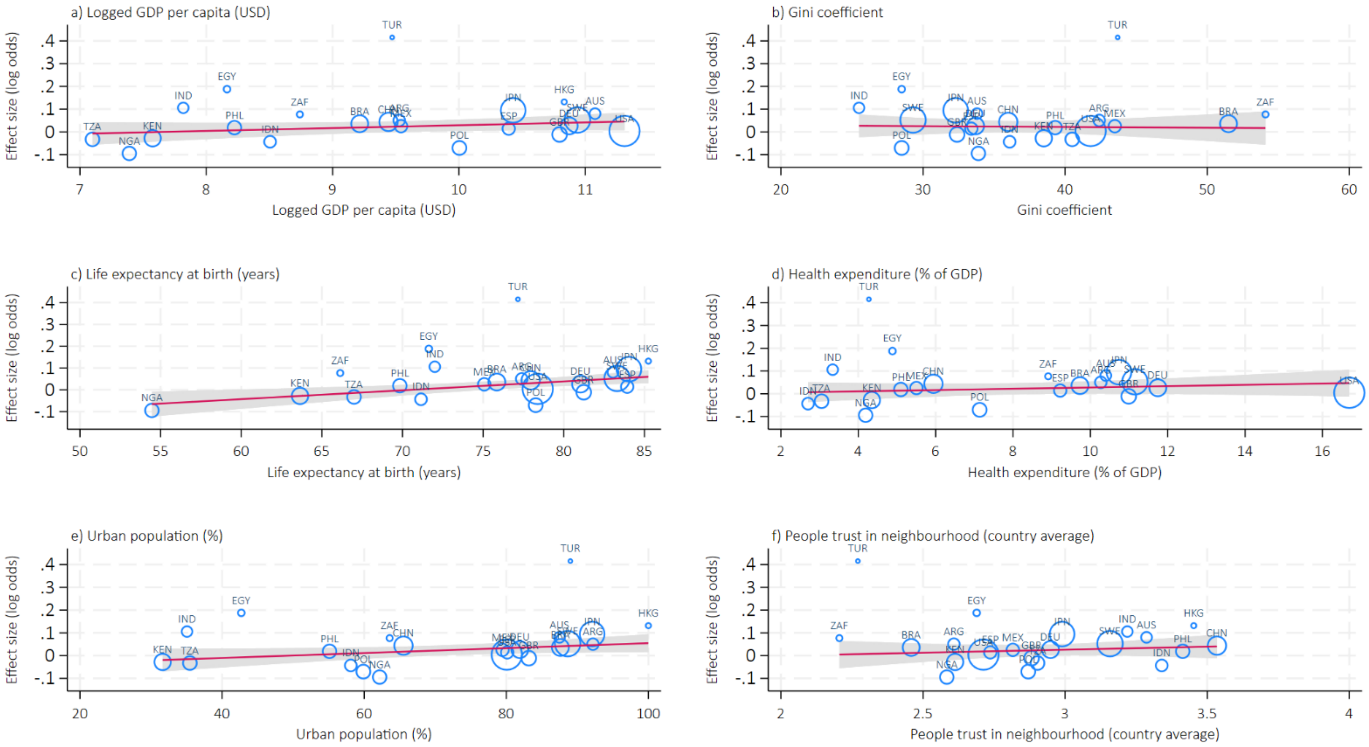

#### 18) Overall mental health

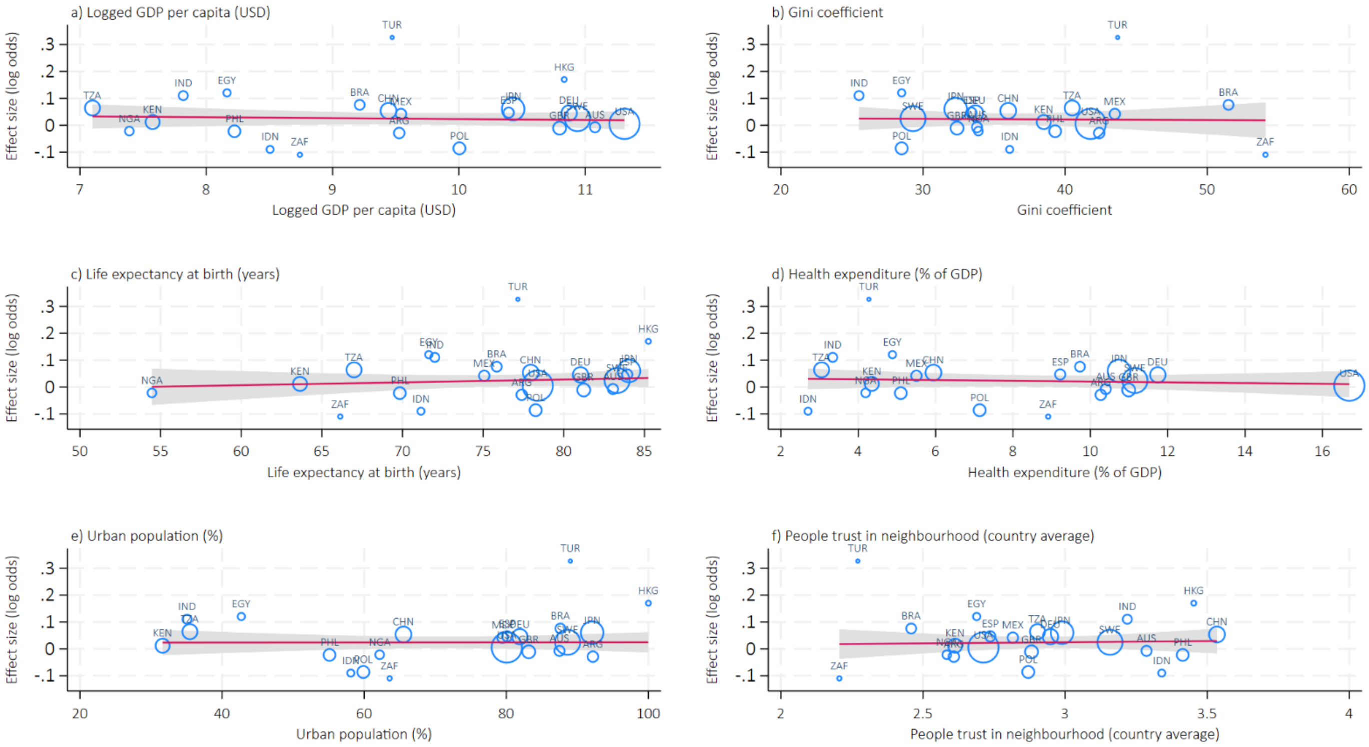

#### 19) Overall physical health

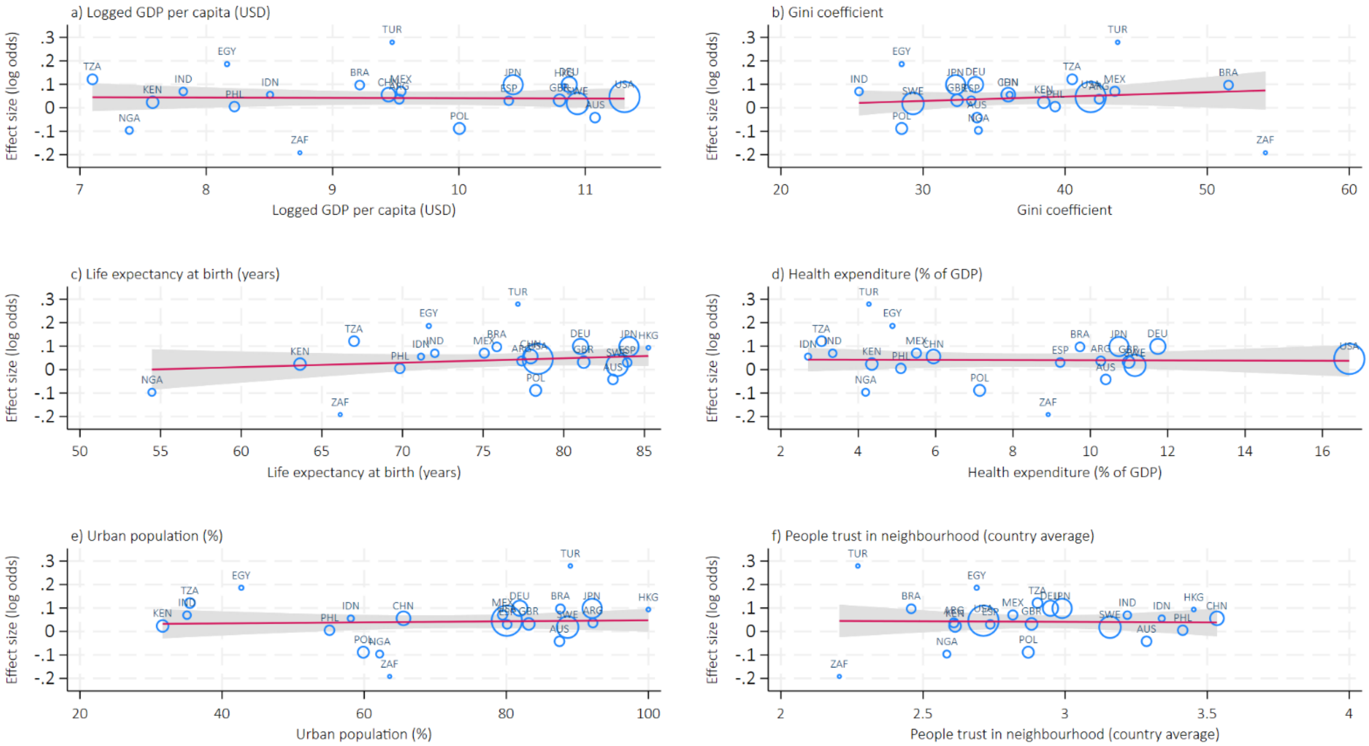
